# The global prevalence of horizontal strabismus: a systematic review and meta-analysis with a focus on ethnic variation

**DOI:** 10.64898/2025.12.23.25342942

**Authors:** Christopher S. von Bartheld, Molly M. Hagen, Jiayi Jiang, Wei Yang, Andrea B. Agarwal

## Abstract

The prevalence of the two types of horizontal strabismus, esotropia and exotropia, varies considerably between studies. This variability has been attributed to factors such as geography/environment, research methodology, age of study subjects, and/or ethnicity. Comprehensive estimates of regional and global prevalences of esotropia and exotropia are lacking, making it difficult to recognize true patterns, trends, and etiologies. Our systematic review compiles prevalences and ratios of esotropia to exotropia from 315 population-based studies and 374 clinic-based studies. We analyze data to assess effects of ethnicity, geography, age, and we identify generational changes of horizontal strabismus. Major ethnicities differ in patterns and ratios of esotropia and exotropia prevalence, not only Caucasians and East Asians, but also Latinos/Hispanics, South Asians, Africans, and Native Americans. Compared to population-based studies, clinic-based studies underestimate exotropia frequency. By weighing prevalences according to the population size of ethnicities, we estimate the worldwide prevalence of horizontal strabismus in the current generation at 1.81% (138.5 million people), comprising 60.0 million people with esotropia (0.67%) and 87.5 million with exotropia (1.14%). In the previous generation, the worldwide prevalence of horizontal strabismus was 1.64% (86.5 million people), comprising 50.5 million with esotropia (0.96%) and 36.0 million with exotropia (0.68%). Esotropia and exotropia prevalences differ between generations within the same ethnicity, indicating that extrinsic factors can modify the underlying intrinsic (genetic) disposition.

## 1 Introduction

Strabismus is a relatively frequent ophthalmological condition that impacts visual function. Studies and review articles most often quote a prevalence of 2-4%.^32,75,79,113,142,150,179,196,210,251,255,261,268,271,290,293,318,373,399,485,448,455,501,545,569,577,596,614^.^645,665,685,753,776,77 7,807,815,817,845,868,A,B^ However, population-based studies have reported the prevalence of strabismus within a surprisingly wide range, with extremes of 0.13% to 9.9% for normal populations (Fig. 1; Table 1). Previous attempts to estimate the global prevalence of strabismus^B,C,318^ were based on a relatively small number of studies and resulted in estimates of 1.9% to 2.9%. Because of the large variability in primary sources, spanning two orders of magnitude, the true global prevalence of horizontal strabismus is unknown. Multiple reasons –not mutually exclusive – for the wide ranges have been considered. These include the methodology employed for examination, selection of non-representative cohorts, the classification of strabismus used, the age of subjects in the cohort, the geography, as well as the ethnicity of the cohorts examined. While older studies had suspected that geography was a factor, possibly due to differences in ambient light exposure with latitude,^100,109,344,372,436^ some of the most recent and detailed reports concluded that methodological and/or age differences are mostly responsible for the wide variability between studies.^A-C,318,356^ To obtain a more valid and global account of the prevalence of horizontal strabismus, we compiled and analyzed in this systematic review all reports that we could find, regardless of origin, language, and type of publication.

**Fig. 1.**
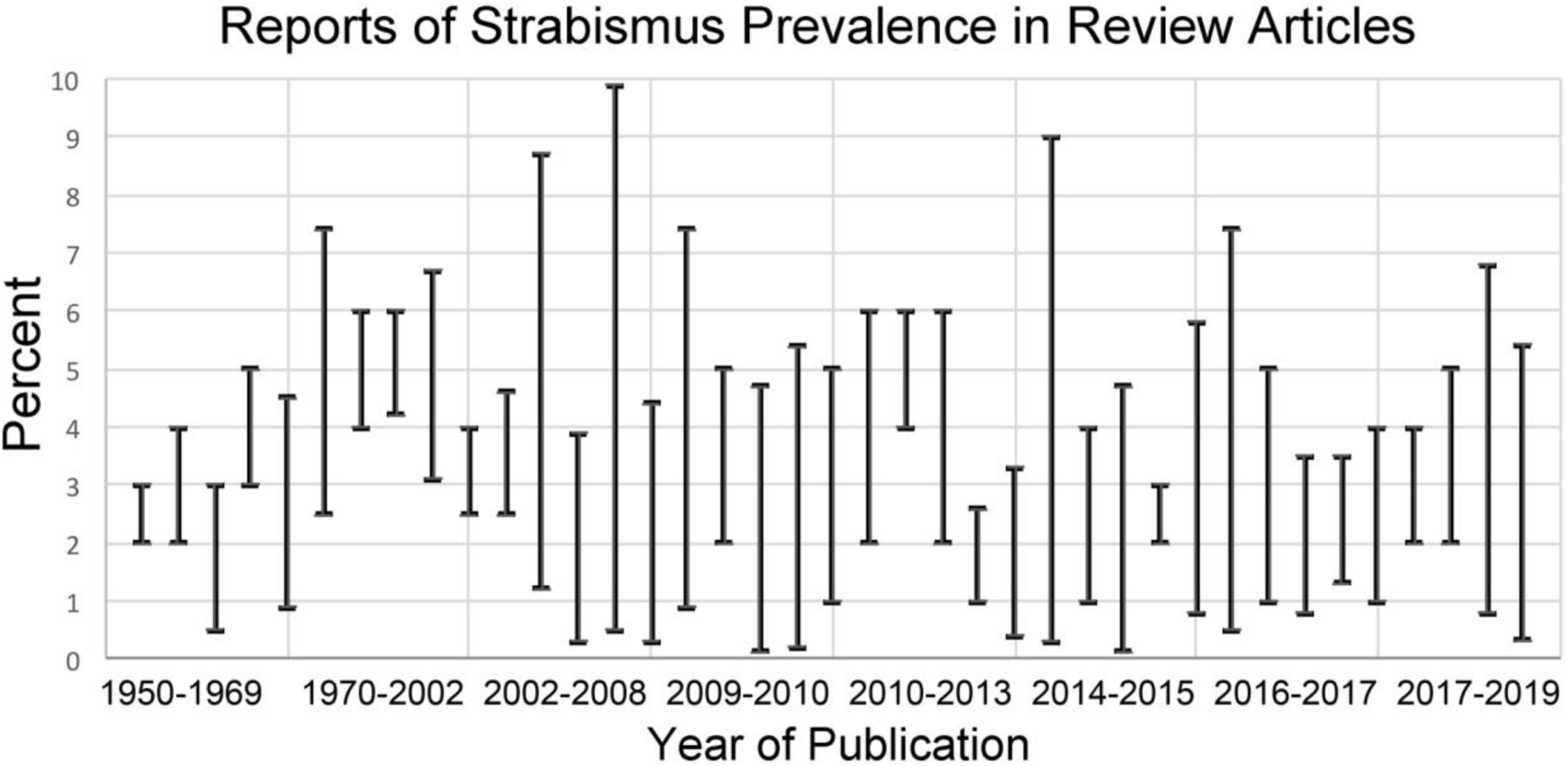
Reports of ranges of strabismus prevalence according to the literature (1951 to 2019), compiled from Table 1, based on reviews and other reports^32,75,79,113,142,150,179,196,210,251,255,261,268,271,290,293,318,373,399,485,448,455,501,545,569,577,596,614.645,665,685,753,776,777,807,815,817,845,86,A,B^

**TABLE 1.** Ranges of strabismus prevalences quoted in reviews.

| Author | Year | Range |
| --- | --- | --- |
| Keiner | 1951 | 2% - 3% |
| Waardenburg | 1954 | 2% - 4% |
| Frandsen | 1960 | 0.5% - 3% |
| Gansner | 1968 | 3% - 5% |
| Rantanen & Tommila | 1971 | 0.9% - 4.5% |
| De Decker & Tessmer | 1973 | 2.5% - 7.4% |
| Aichmair et al. | 1992 | 4% - 6% |
| Stidwill | 1997 | 4.2% - 6% |
| Kvarnström et al. | 2001 | 3.1% - 6.7% |
| Ziakas et al. | 2002 | 2.5% - 4% |
| Donnelly et al. | 2005 | 2.5% - 4.6% |
| Matsuo & Matsuo | 2005 | 1.2% - 8.7% |
| Grönlund et al. | 2006 | 0.3% - 3.9% |
| Robaei et al.(a) | 2006 | 0.5% - 9.9% |
| Multi-ethnic | 2008 | 0.3% - 4.4% |
| Azonobi et al. | 2009 | 0.9% - 7.4% |
| Friedman et al. | 2009 | 2% - 5% |
| Chia et al. | 2010 | 0.13% - 4.7% |
| Green-Simms & Mohny | 2010 | 0.2% - 5.4% |
| Garvey et al. | 2010 | 1% - 5% |
| Torp-Pedersen et al. (a,b) | 2010 | 2% - 6% |
| von Bartheld et al. | 2010 | 4% - 6% |
| Oystreck & Lyons | 2012 | 2% - 5% |
| Sanfilippo et al. | 2012 | 2% - 6% |
| Péchereau et al. | 2013 | 0.99% - 2.6% |
| Fu et al. | 2014 | 0.4% - 3.3% |
| Lin et al. | 2014 | 0.3% - 9% |
| Ye et al. | 2014 | 1% - 4% |
| Jeong & Kim | 2015 | 0.13% - 4.7% |
| Sharbini<sup>A</sup> | 2015 | 2% - 3% |
| Adinanto et al. | 2016 | 0.8% - 5.8% |
| Awoyesuku et al. | 2016 | 0.5% - 7.4% |
| Bruce & Santorelli | 2016 | 1% -5% |
| Chen et al. | 2016 | 0.8% - 3.5% |
| Ehrlich et al. | 2016 | 1.3% - 3.5% |
| Li et al. | 2017 | 1% - 4% |
| Ntizahuyye et al. | 2017 | 2% - 4% |
| Njambi et al. | 2017 | 2% - 5% |
| Wallace et al. | 2018 | 0.8% - 6.8% |
| Hashemi et al. | 2019 | 0.1% - 5.65%* |

\* corrected - one quoted study with a larger percentage (Diniz and Pacha, 2007) was clinic-based, not population-based

Strabismus is known to have a genetic component.^608^ According to Hippocrates (ca. 460-375 BC), strabismus “runs in families.”^339,810,815^ The most frequent form of strabismus is horizontal strabismus, which consists of two types: esotropia (hereafter “ET,” convergent deviation) and exotropia (hereafter “XT,” divergent deviation). Consistent with the view of a genetic component, the pioneering work of Ida Mann^482^ concluded that the distribution of ET “strongly suggests a racial factor,” and that “neither climate nor cultural practices…appear to influence it in any way.” Other authors, however, proposed that horizontal strabismus may also have a considerable environmental component.^100,109,344,352,372,436,625,685,777,845^ Since intensity and duration of sunlight affect eye growth and function,^254,481,497,499,512,566^ differences in geography and climate are thought to be plausible factors that may contribute to the risk and the types of horizontal strabismus.^810^

Other factors that may explain differences in results between studies include the age of the study population, because strabismus prevalence is age-dependent, ^23,90,166,170,173,251,268,287,399,428,443,444,502,503,534,575,697,700,771,748,778,A,D^ and studies vary in the age ranges of the cohorts examined. Additional factors that may be responsible for differences between studies are the definitions and methodologies used for examination,^61,287,293,697,A,B^ and the epoch or decade of investigations because of generational changes.^62,448,498,857,A-C^ Most of the older studies, and especially those that are widely cited, were conducted in countries with predominantly European (Caucasian) populations, and, more recently, with East Asian populations,^174,290,318,501,665,A^ while other populations (Africans and South Asians) have received relatively little attention.^40,861^ If ethnicity is a major determinant of ET and XT prevalence, then this must be considered to avoid Eurocentric bias when attempts are made to estimate the global prevalence of strabismus. Since certain forms of strabismus are associated with other diseases such as mental illness, schizophrenia in particular,^27,28,445,507,535,560,698,701,780^ estimates of the total number of people with ET and XT, and past and current trends of prevalences are relevant for health policy beyond vision care.

Some investigators have compiled and compared the prevalence results and ET/XT ratios from previous reports; however, these studies considered only a small fraction (at most 15%) of the total number of studies on prevalence of ET and XT (Table 2), and rarely included reports written in languages other than English.^174,271,282,290,318,501,665,A,B^ We reason that a systematic review and meta-analysis of all available data on the prevalence of horizontal strabismus in a wide variety of populations and cohorts is more informative about the global prevalence and will help to rectify the relative neglect of ethnicities representing nearly half of the world’s population. On the basis of this approach, we estimate the true prevalence of ET and XT in different countries and worldwide, and we determine patterns and prevalence changes over time that may help to identify risk factors and potential underlying causes for such trends and for the differences between populations.^381^ To include a historical perspective, we review how knowledge about ethnic differences in horizontal strabismus evolved over the last century.

**TABLE 2:** Numbers of studies considered in reviews of the prevalence and relative frequency of horizontal strabismus types.

| Authors and Year | # of Studies Considered | Esotropia (ET) | Exotropia (XT) | Population-based | Clinic-based |
| --- | --- | --- | --- | --- | --- |
| Matsuo and Matsuo 2005 | 12 | 10 | 10 | 12 | 0 |
| Robaei et al 2006a | 20 | 18 | 18 | 20 | 0 |
| Chia et al 2010 | 17 | 14 | 14 | 17 | 0 |
| Curtis et al 2010 | 32* | 31 | 31 | 25 | 7 |
| Garvey et al 2010 | 12 | 12 | 12 | 12 | 0 |
| Green-Simms and Mohny 2010 | 45** | 31 | 34 | 44 | 1 |
| Walline et al. 2012 | 6 | 5 | 5 | 6 | 0 |
| Sharbini 2015 [Thesis]<sup>A</sup> | 70 | 44 | 44 | 62*** | 8 |
| Adinanto et al 2016 [Abstract]<sup>B</sup> | 75 | 30 | 30 | 75 | 0 |
| Bruce and Santorelli 2016 | 7 | 7 | 7 | 7 | 0 |
| Goseki and Ishikawa 2017 | 12 | 11 | 11 | 9 | 3 |
| Wallace et al 2018 | 16 | 15 | 15 | 16 | 0 |
| Hashemi et al 2019 | 56 | 28 | 28 | 55 | 1 |
| Current review (von Bartheld et al.) | 689 | 689 | 689 | 315 | 374 |

\* 1/32 did not report on ET and XT prevalence separately
\*\* 12/45 did not report on ET and XT prevalence separately
\*\*\* 18/62 did not report on ET and XT separately

## 2 Methods

### 2.1 Methods of Literature Search/ PRISMA Guidelines

We followed the PRISMA guidelines for systematic searches and meta-analyses.^528^ Our review protocol was registered in PROSPERO (CRD420251080354).^E^ To identify relevant articles, we searched two databases, PubMed and Google Scholar. PubMed was queried with the keywords “strabismus” and “prevalence” which generated 2,653 records (July 1, 2025) and Google Scholar with the key words “strabismus”, “prevalence”, “esotropia”, and “exotropia,” which generated 7,880 records (July 1, 2025). We accepted all relevant reports that met the inclusion criteria (see below, 2.2), even when they were dissertations, conference proceedings, government reports, or were published in languages other than English (see flow-chart, Fig. 2). When not directly accessible or available through our institution’s library, full-length texts were procured by interlibrary loan or by contacting the author directly. To find additional relevant texts that were missed by the PubMed and Google Scholar searches, we surveyed all references in each of the relevant reports for additional studies that matched our inclusion criteria. We used the link “similar articles” in PubMed to identify additional relevant sources and we examined each paper that cited a relevant report in the Web of Science Core Collection database (“Cited Reference Search”) ^F^ for studies that had been missed. In total, 13,625 titles were screened. We ultimately found 689 reports that met the inclusion criteria. Original sources were read by multiple authors, leveraging the language skills among our team. There were no language restrictions as long as references or English titles or relevant key words appeared in searches; for example, we included 137 studies published in twenty different languages other than English: Chinese, Czech, Danish, Dutch, French, German, Greek, Indonesian, Italian, Japanese, Korean, Macedonian, Norwegian, Persian, Polish, Portuguese, Russian, Spanish, Swedish, and Thai, and we asked colleagues fluent in those languages to translate as needed.

**Fig. 2.**
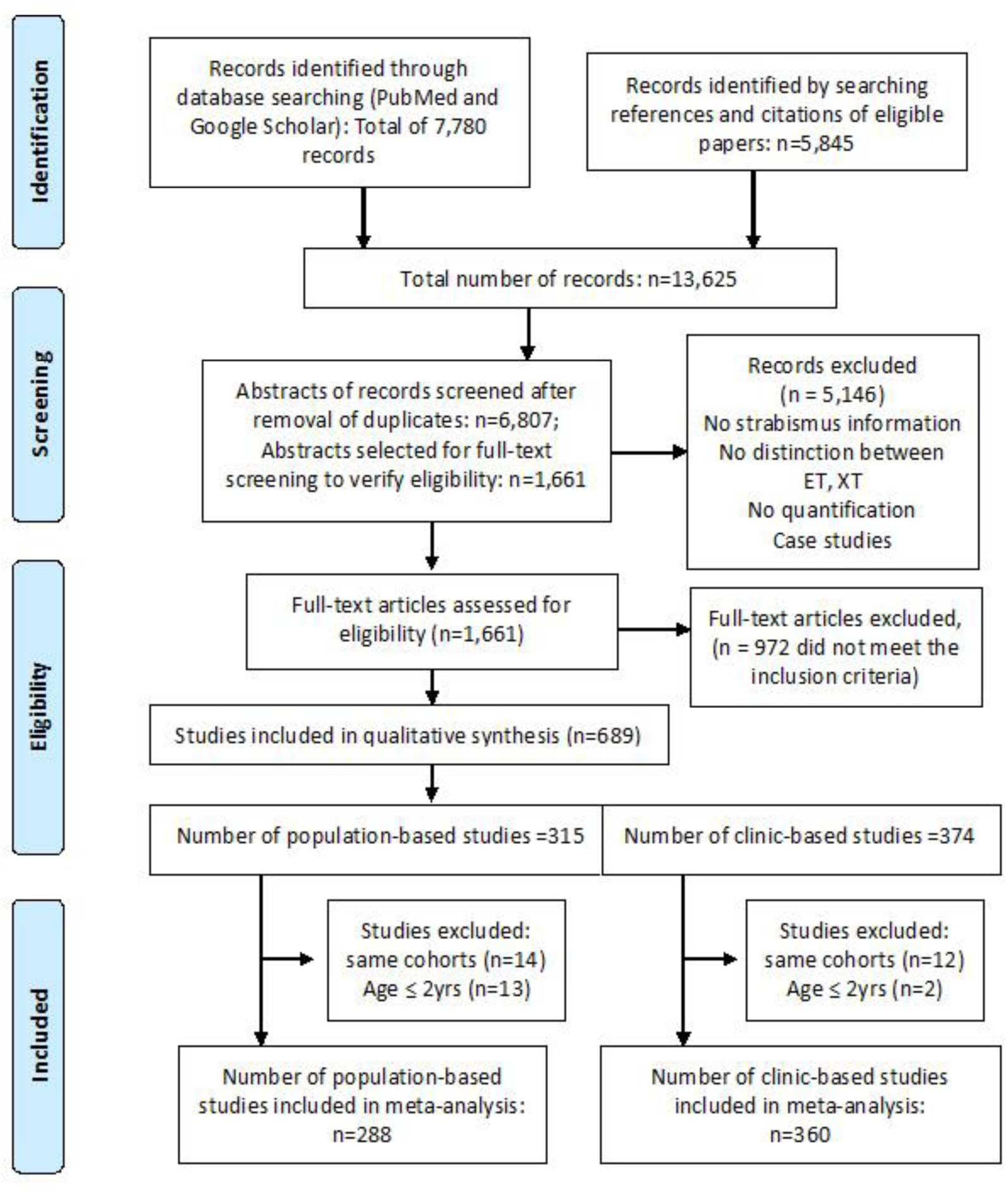
Flow diagram of the search strategy, article selection, application of inclusion and exclusion criteria, and removal of duplicates. The literature search was last updated on July 1, 2025. This review protocol was registered in PROSPERO (CRD420251080354).^E^

### 2.2 Inclusion Criteria for Studies

We accepted all studies that report original and quantitative data on frequencies of esotropia (ET) and exotropia (XT) in human subjects, either obtained from population-based surveys, or from medical records in clinical settings. Reports had to be on humans; we did not consider prevalence studies on non-human primates,^411–412^ and we did not include case reports. Studies were included only when they provided *quantitative* information about both ET and XT separately (not just combined ET and XT prevalence or prevalence of horizontal plus vertical strabismus). We did not include in our quantitative analysis studies that failed to provide exact information^439,508,674^ or studies with only qualitative comments such as “most cases were esotropia”, or “nearly all cases were exotropia.”^244,626,647,683,814,865^ We accepted both, population-based studies (screening of birth cohorts, schools or pre-schools, or military recruits, or inhabitants within a certain region such as an island, town or village), as well as clinic-based studies (surveys of medical records from patients seeking attention at a clinic). We kept these two types of studies separate, in two different tables and spreadsheets, and used them for separate analyses (see below, sections 3.2 and 3.8). In a few studies, the distinction between population- and clinic-based studies was questionable, due to a low response rate to home questionnaires or merely “inspection” by non-expert personnel prior to referral to an ophthalmologist, so that not the entire population was properly screened and would likely have given a significant underestimate of the true prevalence.^373,408,453,503^ Accordingly, these four studies were listed as “clinic-based.”

### 2.3 Data Curation

We compiled data in two main tables (Supplemental Tables S1 and S2): those from population-based studies (n=315) and those from clinic-based studies (n=374). Since many studies reported not only horizontal strabismus, but also the (relatively rare) occurrence of pure vertical strabismus (about 4-5% of all strabismus),^165,290^ the data were verified to report prevalences exclusively for ET and for XT, but not pure vertical strabismus, and quantitative data on prevalence were re-calculated and verified to provide prevalence values to a resolution of two places after the decimal point. We considered all relevant studies that met the inclusion criteria, but we omitted abstracts when a full-length publication of the same cohort had become available. We list all relevant studies by country and region, even when data from the same cohort was published more than once, but we used information from the same cohort only once in the data analyses. Duplicate publications of the same cohort are marked with italic font in Supplemental Tables S1 and S2.

Prevalence of a condition such as horizontal strabismus can refer to lifetime prevalence or to point prevalence.^755^ Here, we report primarily on lifetime prevalence, but acknowledge that this is based at least in part on studies that report point prevalence. Reports of horizontal deviations in newborns are included in the Supplemental Tables S1 and S2 for completeness.^18,59,206,258,330,347,568,772^ However, we did not include in our meta-analyses studies or cohorts that focused entirely on infants two years of age or less, for two reasons. First, it is controversial how many deviations in newborns are “genuine strabismus”^399^ or may be an artifact – human judgement of newborn gaze may not be reliable and should take angle λ into account.^305,351,739,772,831^ Many deviations are thought to be transient and typically resolve within a few days or weeks, although some of them are genuine cases of infantile/congenital esotropia.^59,251,258,347–350,399,568,741^ Second, because only a fraction of horizontal strabismus is known to develop before the age of 2 years, ^23,166,173,251,399,428,503,575,778,D^ we excluded in our meta-analyses such studies (gray-shaded in our Supplemental Tables S1 and S2).^55,59,206,226,227,255,258,330,347,503,568,715,741,747,772^ This does not mean that early types of horizontal strabismus such as the relatively rare infantile/congenital ET^290,810^ were excluded, because they are represented in the many studies (n=56) with larger age ranges beginning at 0-2 years of age. For additional information, we included in our tables the location and number of subjects (cohort size), the age range, and the ethnicity (when such information was available). Data from clinic- (or primarily referral-) based studies are listed in a separate table (Supplemental Table S2), including the country and region/ethnicity, the relative contributions of ET and XT, and, if available, the number of subjects, and mean age or age range of subjects.

### 2.4 Data Analysis

#### 1. Justification for Selection of Ethnicities

We analyzed the main ethnic populations separately, extending the practice of previous studies and reviews.^113,147,149,170,197,255,290,361,380,436,510,545,665,773,793,859,A,D,G^ Based on the compiled cases of ET and XT, we calculated the prevalence values for thirteen major ethnicities, obtained from 76 countries on six continents, according to the population-based studies. Some of these studies evaluated only one ethnicity, some evaluated multiple ethnicities, but did not report them separately, and a third group of studies evaluated multiple ethnicities, and reported them separately. Accordingly, we compiled most data by ethnicity, but in some cases had to present data in aggregate, for “multiple ethnicities,” combined.

In addition to the ethnicities of Caucasian, Native American, Inuit/Eskimo, Latino/Hispanic, African American, African, Middle Eastern, South Asian, East Asian, and Oceanian (Indigenous Australian and Melanesian), there were sufficiently large differences between African populations to warrant recognition of two types: the Niger-Congo-Bantu type in West Africa, and all other populations in Africa, consistent with the conclusions of anthropological, linguistic and genetic studies.^121,167,329,774^ Likewise, there were significant differences between populations in the Northwest and Southeast of the Indian subcontinent that warranted a separation, again confirming genetic data,^91,383,402,475,655^ and Persians have a different genetic history than other populations in the Middle East.^232^ Therefore, a total of thirteen major ethnicities will be considered in this review, including two types in Africa, two types in the Middle East, and two types in South Asia.

Caucasians in Europe, the Americas, South Africa, and Oceania were considered for the total Caucasian count; Europe includes Eastern Europe and Russia or the former Soviet Union. Israelis are listed among the Middle East category, because Jewish people are genetically closest to Middle Eastern populations.^595^ We list Native Americans and Inuit/Eskimos separately, because these populations have distinct ancestries.^246,656,737^ Although Nepal is geographically part of South Asia, we grouped Nepalis among the East Asian ethnicity, because recent genetic evidence indicates that Himalayan valleys were largely populated from Tibet by East Asians, about 6,000 years ago.^374,820^ For this review, we did not further divide the studies of indigenous people in Oceania, although recent work shows that the genetic make-up of the indigenous people in Australia and the Solomon Islands appears to differ from those in Papua-New Guinea, possibly due to divergence some 30,000 years ago.^398,477,738^

#### 2. Meta-Analysis and Comparison of Prevalence between Ethnicities

Prevalence of ET and XT was compiled from population-based studies separately for each major ethnicity. We used R Studio, version 3.6.1, for statistical analyses (R Foundation for Statistical Computing, Vienna, Austria). To calculate estimates of pooled prevalence and 95% confidence intervals for both ET and XT we used the R-meta package, version 4.9-5, and the metaprop function.^87^ The ET/XT ratios for small cohorts were adjusted to avoid having a “0” in the numerator or in the denominator (Supplemental Table S1).^758^ We used random effects models with the inverse variance method for pooling and the logit transformation for proportions.^146,185^ For ease of interpretation we back transformed and rescaled proportions to events per 100 observations. Analysis of the heterogeneity across studies was done using the Maximum-likelihood estimator, Higgin’s I^2^ and Cochran’s Q method.^146,337^ Publication bias was assessed by visual inspection of funnel plots.^209^ In all cases, significance was defined at α = 0.05.

Subgroup analysis was conducted by ethnicity. Specifically, ethnicity was coded as a categorical variable with 13 levels: Caucasian, Native American, Inuit/Eskimo, Latino/Hispanic, African American, West Africans, all other Africans, Persians, all other Middle Easterners, South Asian (Northwest and Southeast separately), East Asian, and Indigenous Oceanian. To further quantify differences in ET and XT prevalence between the 13 ethnic groups, we followed up our meta-analysis with pairwise Pearson chi square tests for independence. To avoid a bias toward very large studies, we adjusted study size in the top 10% of studies to the 90th percentile prior to conducting our chi square tests. Because only two ethnicities, Caucasians and East Asians, have been examined and reported with sufficient numbers of studies over multiple generations in regard to ET and XT prevalence, we used separate chi square tests to evaluate prevalence differences between generations for these ethnic groups in a longitudinal analysis. We defined a generation as 30 years based on the average number of years between the birth of a parent and their children.^782^

#### 3. Estimation of Global Prevalences and Preparation of a World Map

Prevalence of horizontal strabismus differs between ethnicities; ethnicity, therefore, must be taken into account when global prevalence is estimated. The data from Supplemental Table S1 was used to calculate the prevalence of ET and XT for each of the major ethnicities. Populations of the same ethnicity residing in different countries and continents were combined, and a pooled prevalence for each major ethnicity was calculated as explained above, with 95% confidence intervals, and plotted in a histogram, arranged by a decreasing ET/XT ratio. Our quantitative prevalence analysis considered 288 population-based studies from 76 countries around the world (after excluding 13 population-based studies on subjects two years of age or less and 14 additional studies reporting on duplicate cohorts). Based on the prevalences of each ethnicity, and the approximate total population size of ethnicities worldwide, we estimated the global prevalence for ET and for XT. We achieved this by extrapolation and weighing the population size of each of the major ethnicities (United Nations Report: List of countries by population).^H^ Because of apparent generational (secular) changes in Caucasians and in East Asians, we separately estimated the prevalence of ET and XT in the *current* generation, based on studies published after 1995, and also in *previous generations*, based on studies published from 1846–1875, 1876–1905, 1906–1935, 1936–1965, 1966–1995, and 1996–2025, taking into account that some data from previous generations were reported with a long delay.^147,164,700,I^

To generate a world map of ET and XT prevalence ratios, we considered those same prevalence data, but plotted them based on geography (country or region of residence). This map displays three “basic” types – prevalence of ET greatly exceeding XT (ET/XT ratio >1.5, blue), ET prevalence much lower than XT (<0.67, red), and ET having approximately the same prevalence as XT (ET/XT ratios of 0.67–1.5, yellow). Data from clinic-based studies were used to corroborate the patterns, using slightly lighter colors for countries where population-based studies are currently lacking, but clinic-based information was available.

#### 4. Measurement of Equitability and Impact of Studies on Different Ethnicities

To assess whether different ethnicities are represented in an equitable fashion in studies of strabismus prevalence, and to quantify any existing imbalances, we calculated the number of studies per ethnicity and the percentage of the population that had been sampled, as a tool to assess whether ethnicities have been represented proportionally. We compiled the total number of subjects that had been screened, separately for each major ethnicity. We calculated the total number of subjects in population-based cohorts for each ethnicity and the number of subjects with medical records (clinic-based studies), again for each major ethnicity. We also calculated the number of cohorts per ethnicity, and those numbers relative to each major ethnicity’s contribution to the world population.

To assess whether studies of different ethnicities have had commensurate impact, according to their contribution to the world population, and to assess whether certain ethnicities have been neglected, we measured the percentage of publications’ citations as a fraction of the total citations. Citations were obtained on October 4-6, 2025 from the Web of Science Core Collection, Cited Reference Search,^F^ as a measure of their impact.

#### 5. Effect of Latitude

We used simple linear regressions to assess the effect of latitude/geography, by plotting the ET and XT prevalences (and the ET/XT ratio) against latitude. Four ethnic groups (Native Americans, East Asians, Caucasians in Europe, Caucasians in Australia) were eligible for this analysis, because these populations extend geographically over a significant north-south expansion, and there were sufficient data and numbers of studies available across latitudes. We performed our analysis separately for ET and XT, to determine which type of strabismus is mostly influenced, if at all, by latitude. We also investigated differences in prevalence between continents for Caucasians.

#### 6. Effect of Age

We plotted the age ranges for cohorts to assess whether there were any apparent age differences between sampled ethnicities, and we calculated ethnic differences with and without studies that were on adults only. For all these analyses, we excluded the studies with exclusive focus on ages of 2 years or less, as explained in section 2.3. Prevalence and frequency of ET vs XT was compared between studies that included children and those that focused entirely on adults or late teenage years. We also examined changes in ET or XT prevalence of ET/XT ratio changes in older adults, by reviewing studies that reported exclusively or specifically on subjects 55 years of age and older (n=5).^178,218,231,320,444^

#### 7. Comparison of Data from Population-based and Clinic-based Studies

We compared the ratios of ET and XT prevalence from 288 population-based studies (which excluded 14 studies that reported on duplicate cohorts, and 13 studies that focused on ages ≤2 years) with the relative frequencies of ET and XT from 360 clinic-based studies (after excluding 12 reporting on duplicate cohorts and two studies reporting on ages 2 years and younger), weighted for cohort size as explained in section 2.4.3. This was done separately for nine major ethnicities: Caucasians, South Asians (Northwest), South Asians (Southeast), West Africans, all other Africans, Persians, all other Middle Easterners, Latinos/Hispanics, and East Asians. Because of small numbers of clinic-based studies (n≤1) and/or high heterogeneity, this comparison was not possible for African Americans, Native Americans, indigenous Oceanians, or Inuit/Eskimos. Nevertheless, this approach informs whether clinic-based ratios can be used as a proxy for population-based studies, a question that has been discussed for over 150 years.^149,173,174,179,390,753,773,811,841,857^

#### 8. Changes in Prevalence between Generations: Comparison of 2–5 Generations

Several reviews comparing clinic-based percentages of ET and XT have reported changes (decreased numbers of strabismus surgeries) over recent decades.^62,102,126,149,150,158,161,195,236,332,366,376,433,448,458,464,498,663,719,720,824,857^ To determine whether prevalences of ET and/or XT have changed over time, we compared prevalences in two major ethnicities that had sufficient numbers of population-based studies (data points) distributed over a longer time period (several decades), thus making them suitable for such a longitudinal analysis: Caucasians in Europe and North America, and East Asians.

### 2.5 Evaluation of Biases: Examiner Expertise, Study Design (Sampling), and Publication Bias

Bias can affect prevalence estimates at multiple levels and through diverse mechanisms.^182^ To address potential biases in the estimation of ET and XT prevalence, we pursued the following approaches.

#### 1. Variability due to diverse Methodology/Classification

Different studies apply different methodology and classifications, and the expertise of examiners can vary as well. For this reason, some investigators have proposed to consider only so-called “gold standard studies”.^A,B^ To determine whether such “gold standard studies” indeed yield results that differ significantly from those when all studies are considered, we compared them with studies examining cohorts of comparable age and ethnicity, and we used unpaired two sample t-tests to determine any statistically significant differences.

#### 2. Sampling Bias

A potentially profound concern is that population-based studies are mostly school- or preschool-based, and such studies generally assume that all children have the same chance of being included in the sampled institutions. However, children with severe disabilities often do not attend normal schools and, therefore, may be missed.^110,113,122,255,287,333,369,575,582,808,A^ Since infants and children with disabilities have an increased prevalence of horizontal strabismus (see section 3.11.ii), studies based on sampling of “regular” schools underestimate the true prevalence of strabismus. To estimate the effect size of this bias, we compared age- and ethnicity-matched studies with the rare studies that obtained data from birth cohorts (a more time-consuming strategy)^413^ and therefore collect information from the entire (i.e., representative) population, not only of those children attending regular schools. There were thirteen such birth-cohort studies on Caucasians (on children older than 2 years),^65,147,287,394,420,428,491,575,624,649,732,748,833^ three in the Middle East,^256,257,564^ and one on African Americans.^147^ Accordingly, this comparative analysis was possible only for Caucasians. We compared the means of ET and XT prevalence of the Caucasian birth-cohort studies with those from school-based studies with similar age ranges to determine the approximate effect size of any sampling bias.

#### 3. Publication Bias and Meta-analysis Bias

Publication bias can be evaluated and recognized with funnel plots.^208,209,750,752^ To evaluate publication bias and other meta-analysis biases, we compared prevalence numbers as a function of cohort size and prepared funnel plots in an ethnicity-specific design to detect and assess such biases.

## 3 Results

### 3.1 Literature Search Results

Our systematic search for relevant literature identified 689 eligible studies: 315 population-based studies from 76 countries (Supplemental Table S1; Fig. 2), and 374 clinic-based studies from 72 countries (Supplemental Table S2), most of them peer-reviewed journal publications or book chapters (n=645), but also government reports and other reports on websites, conference proceedings (n=10) and doctoral dissertations (n=27). We compiled many more studies than had previously been considered – including 137 studies published in languages other than English. We sorted the studies and entered data by continent, country, region or town, by ethnicity, and in chronological order within the following regions: Europe, North America, Latin America, Africa, Middle East, South Asia, East Asia, and Oceania. We provide in Table 3 the total number of subjects that were screened in the studies we compiled (Supplemental Table S1), and also the total number of patients whose medical records were analyzed (studies compiled in Supplemental Table S2). The percentage of the total population size for each ethnicity examined in population-based studies differed, and the mean cohort sizes also differed between ethnicities, ranging from 0.0012% (South Asians) to 1.24% (indigenous Australians), and mean cohort sizes ranged from 301 (Eskimo/Inuit) to 9,490 (Caucasians) (Table 3). Overall heterogeneity was high (*I*^2^ = 96.7% for ET and 98.5% for XT, τ^2^ = 0.9984 for ET, and 0.9867 for XT; for details, see Supplemental Table S3). Heterogeneity was reduced in most subgroups for specific ethnicities, indicating that ethnicity is a major contributor to heterogeneity. The only subgroups where heterogeneity was not reduced were Caucasians (ET) and East Asians (XT), likely due to generational changes described in section 3.12.ii.

**TABLE 3.**
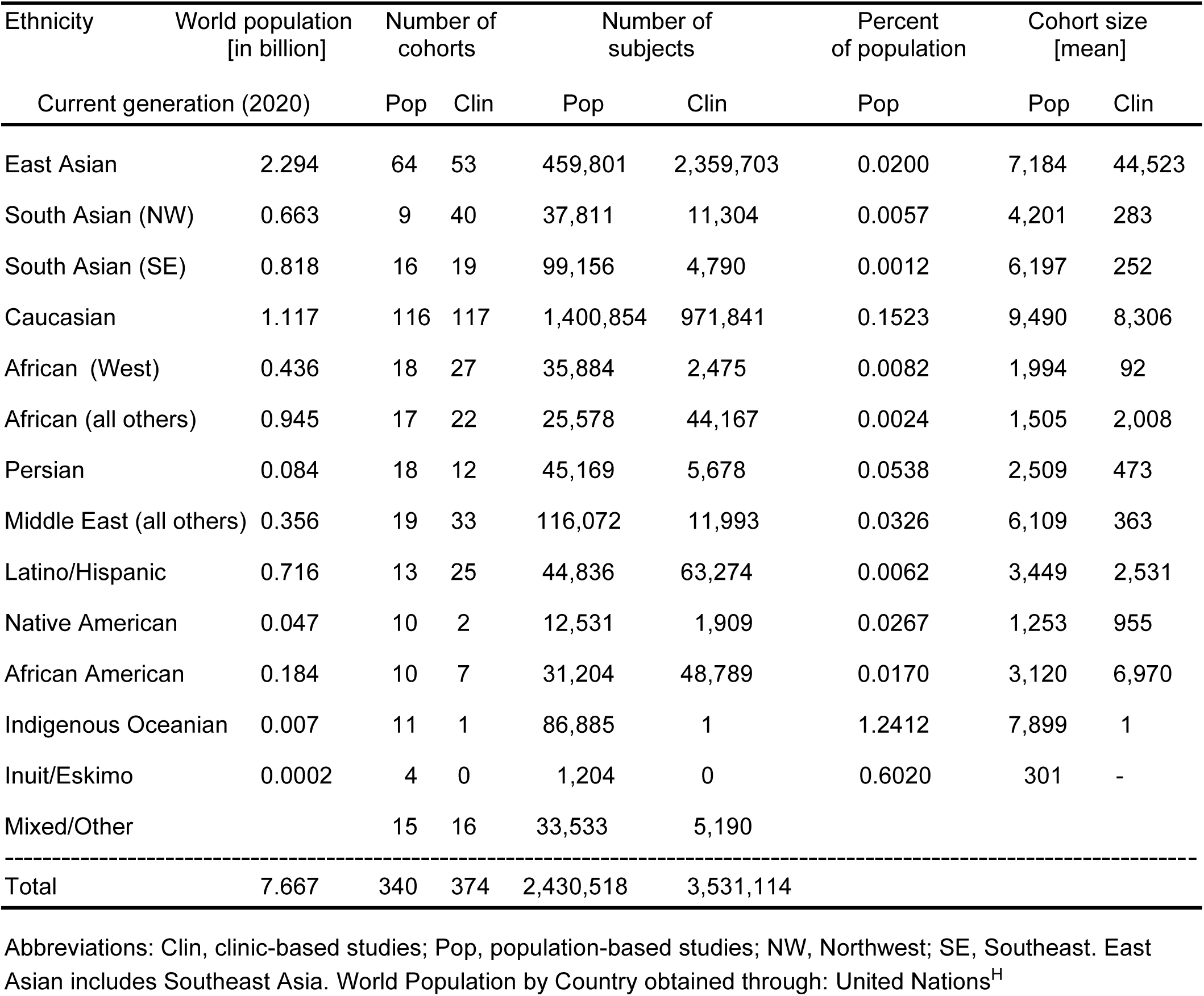
Numbers of subjects and cohort sizes for studies on prevalence and frequencies of horizontal strabismus types in major ethnicities.

### 3.2 Geographical and Ethnic Distribution of ET and XT Prevalence

#### 1. Europe

The ET and XT prevalence was examined in 82 population-based studies (84 cohorts) from 24 European countries, published between 1867 and 2025, in Austria,^24,32,692^ Belgium,^800^ Bosnia,^627^ Czechoslovakia,^515^ Croatia,^394^ Denmark,^250,251,356,684^ Finland,^387,388,429,645,785^ France,^731,743^ Germany,^163,164,179,240,241,258,303,395,678,693,695,702,744,775^ Greenland,^201,J^ Hungary,^809^ Ireland,^196^ Moldova,^599^ Netherlands,^K^ Lithuania,^539^ Norway,^65,343,L^ Poland,^M^ Portugal,^435,N^ Romania,^331^ Russia,^474,506^ Spain,^181,491^ Sweden,^10,61,226,227,266,293,342,416,428,439,446,573,575,582,649^ Switzerland,^249,268,806^ and the UK.^55,112,113,287,347,354,565,581,624,742,748,779,787,833,839^ The cohort-weighted ET prevalence for Caucasians (over 2 years of age) in Europe was 2.065% (1.7917– 2.3771%, 95% CI), the XT prevalence was 0.6247% (0.5045–0.7733, 95% CI), resulting in an ET/XT ratio of 3.31. The prevalence of ET in Caucasians residing in Europe (2.065%) was significantly higher than in Caucasians who reside in continents other than Europe (Random effects model, p=0.0001), with ET prevalences of 1.553% in North America, 0.725% in Latin America, and 1.676% in Oceania (see below).

#### 2. North America and Caribbean

We found 35 population-based studies, reporting on 45 cohorts in Canada, the USA and Caribbean, published between 1925 and 2013, with 22 studies on Caucasians: five in Canada,^380,420,421,667,837^ and 17 studies in the USA.^59,105,147,166,170,200,255,330,414,510,536,568,741,772,D,O,P^ Five studies were on Native Americans,^152,271,437,489,830^ and two studies on Inuit/Eskimos.^380,835^ There were nine studies on African Americans,^147,170,187,255,259,545,631,632,D^ three studies on Latinos/Hispanics,^155,170,545^ one on Asians,^507^ and two on multiple ethnicities.^657,I^ Some of these studies examined different ethnicities separately (Caucasian, Native American, Inuit/Eskimo, African American, Latino/Hispanic, and Asian). Caucasians and Inuit/Eskimos had the highest prevalences of ET (1.553%, 0.9381–2.1063% CI, and 1.997%, 1.7741–5.0557% CI, respectively), while African Americans had a lower ET prevalence (1.348%, 0.9933–1.8257% CI), and Native Americans (in all the Americas) had a very low ET prevalence (0.357%, 0.2018–0.6302% CI), and a very high XT prevalence (1.32015, 0.6793–2.5499%). African Americans had a higher XT prevalence than Caucasians in North America (1.2658%, 0.9527–1.6800% CI vs. 0.9381%, 0.6339–1.3862% CI). African Americans had a much larger overall prevalence of horizontal strabismus (3.006%) than Africans in West Africa (0.9004%) or in other parts of Africa (0.902%, see below).

#### 3. Latin America

We found 20 population-based studies that reported on 23 cohorts in Latin America, published between 1961 and 2024. Six studies reported on mostly Caucasians.^95,168,186,690,694,726^ Ten studies reported on cohorts of multiple ethnicities, mostly Latinos/Hispanics,^171,172,238,270,384,490,505,537,583,Q^ three reported exclusively on Native Americans,^99,272,790^ and one reported on East Asians.^97^ Latinos/Hispanics (in North and South America combined) had a prevalence of 1.7479% for ET (0.809%, 0.5826–1.122% CI), and 0.9389% for XT (0.5554–1.583% CI). Caucasians in Latin America had an ET prevalence of 0.725% (0.5681–0.9258% CI) and an XT prevalence of 0.5577% (0.3206–0.9684 CI). Native Americans in Latin America had a low ET/XT ratio – XT considerably exceeding ET – similar to Native Americans in North America (described above), and therefore these studies were combined because of the relatively small number of studies.

#### 4. Africa

We found 37 studies reporting on 35 cohorts in 16 African countries, published between 1898 and 2024. The studies in northeast, central and south Africa are from Egypt,^9,214^ Ethiopia,^242,275,768^ Kenya,^R,S^ Lesotho,^280^ Madagascar,^71^ Namibia,^252^ Rwanda,^843^ Somalia,^11^ South Africa,^551^ Sudan,^49,760,863^ and Tanzania.^410^ The studies in West Africa are from Congo,^663^ Gabon,^341^ Ghana,^13,423,557^ Liberia,^335^ and Nigeria.^2,15,22,33,34,41,70,76,79,80,86,153,225,T^ The prevalence of horizontal strabismus in most populations in Africa is low, much lower than in major populations in Europe or in North America – the prevalence is 0.9004% in West Africa and 0.902% in other parts of Africa. While West Africans have a lower ET prevalence with 0.366% (0.238–0.5609% CI), and a somewhat higher prevalence of XT with 0.5344% (0.3295–0.8654% CI), this is opposite for people in the other parts of Africa where XT prevalence is significantly lower than ET prevalence: XT prevalence is 0.3692% (0.2232–0.61% CI), while ET prevalence is 0.541% (0.3182–0.9184 CI). The ET/XT ratio in West Africa (Niger-Congo-Bantu populations) is 0.68, while it is more than twice, 1.47, in the North, East and South of Africa, a significant difference (p=0.018, t-test). The reduced ET prevalence largely correlates with the genetically distinct Niger/Congo/Bantu-speaking populations in the western and central regions of Africa.^121,329,610,774^

#### 5. Middle East

We found 36 population-based studies (reporting on 36 cohorts) from ten countries in the Middle East that were published between 1971 and 2024. One study was from Azerbaijan,^12^ 17 from Iran^184,228,229,248,308,316,317,319,320,369,594,643,846–849,U^ – some of these were recently reviewed,^403,679^ three each were from Iraq,^8,307,544^ and Israel,^256,257,564^ one each from Jordan,^463^ Oman,^457^ and Palestine,^430^ three from Saudi Arabia,^16,81,89^ one from Syria,^281^ and five from Turkey.^66,122,132,301,784^ As is apparent in Supplemental Table 1, most Middle Eastern populations (Arabs, Turks, Jews, Kurds) have a higher ET prevalence (1.038%, 0.7086–1.5167% CI) than XT prevalence (0.6634%, 0.4156–1.0573% CI), while this is reversed in Azerbaijanis and Persians, who have a much higher XT prevalence (1.488%, 0.9236–2.3889% CI) than ET prevalence (0.577%, 0.477–0.69695 CI). The ET/XT ratio in Persians and Azerbaijanis is 0.39, while it is 1.56 in all other populations of the Middle East – a highly significant difference (p=0.000026, t-test).

#### 6. South Asia

We found 25 population-based studies (reporting on 25 cohorts) from South Asia, published between 1961 and 2025, from three countries: Bangladesh,^63^ India,^26,29,35,68,94,176,177,296,300,328,393,400,427,509,530,546,552,652,688,733,796^ and Pakistan, including Afghan refugees in Pakistan.^72,73,716^ Thus, the information on the South Asian ethnicity is relatively sparse.^861^ In South Asians, horizontal strabismus has a remarkably low prevalence of 1.0358% in the Northwest of the Indian subcontinent and 0.9028% in the Southeast. However, similar to the horizontal strabismus in Africa and the Middle East, two distinct patterns are present in South Asia, with the Northwest having more ET than XT, while the Southeast has more XT than ET. In the Northwest of the Indian subcontinent, the prevalence of ET was 0.578% (0.3812–0.8761% CI), and the prevalence of XT was 0.4578% (0.2361–0.8859% CI), with an ET/XT ratio of 1.26, while in the Southeast, the prevalence of ET was 0.267% (0.1596-0.4464% CI), and the prevalence of XT was 0.6358% (0.3328–1.2112% CI), with an ET/XT ratio of 0.42 – less than one third of the ET/XT ratio in the Northwest (a borderline significant difference (p=0.0597, t-test), likely due to the small number of studies in the Northwest (n=6). Figure 4 shows how the distribution of ET and XT prevalence on the Indian subcontinent differs between the Northwest and the Southeast.

**Fig. 3.**
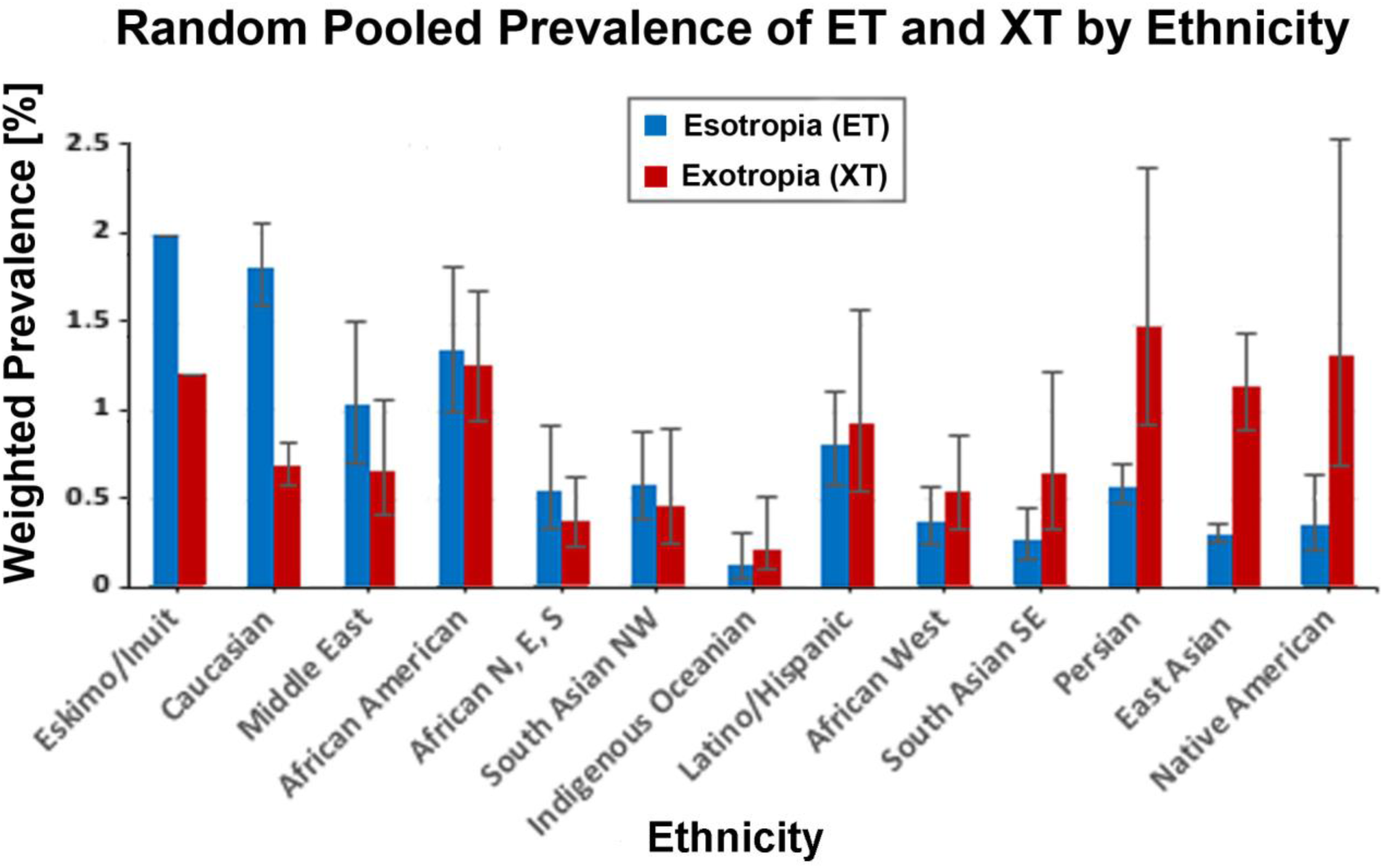
The graph shows the estimated random prevalence of esotropia (ET, blue) and exotropia (XT, red) for 13 different major ethnicities (all generations, when data are available) as indicated. Bars represent 95% confidence intervals, except for Eskimo/Inuit because n=2 for that ethnicity. Data were obtained from 295 population-based cohorts with subjects older than 2 years, and restricted to studies reporting distinct ethnicities. The numbers of studies and details about cohort sizes are summarized in Table 5. The data for Persians includes one study on Azerbaijanis. Abbreviations: E, East, N, North, S, South.

**Fig. 4.**
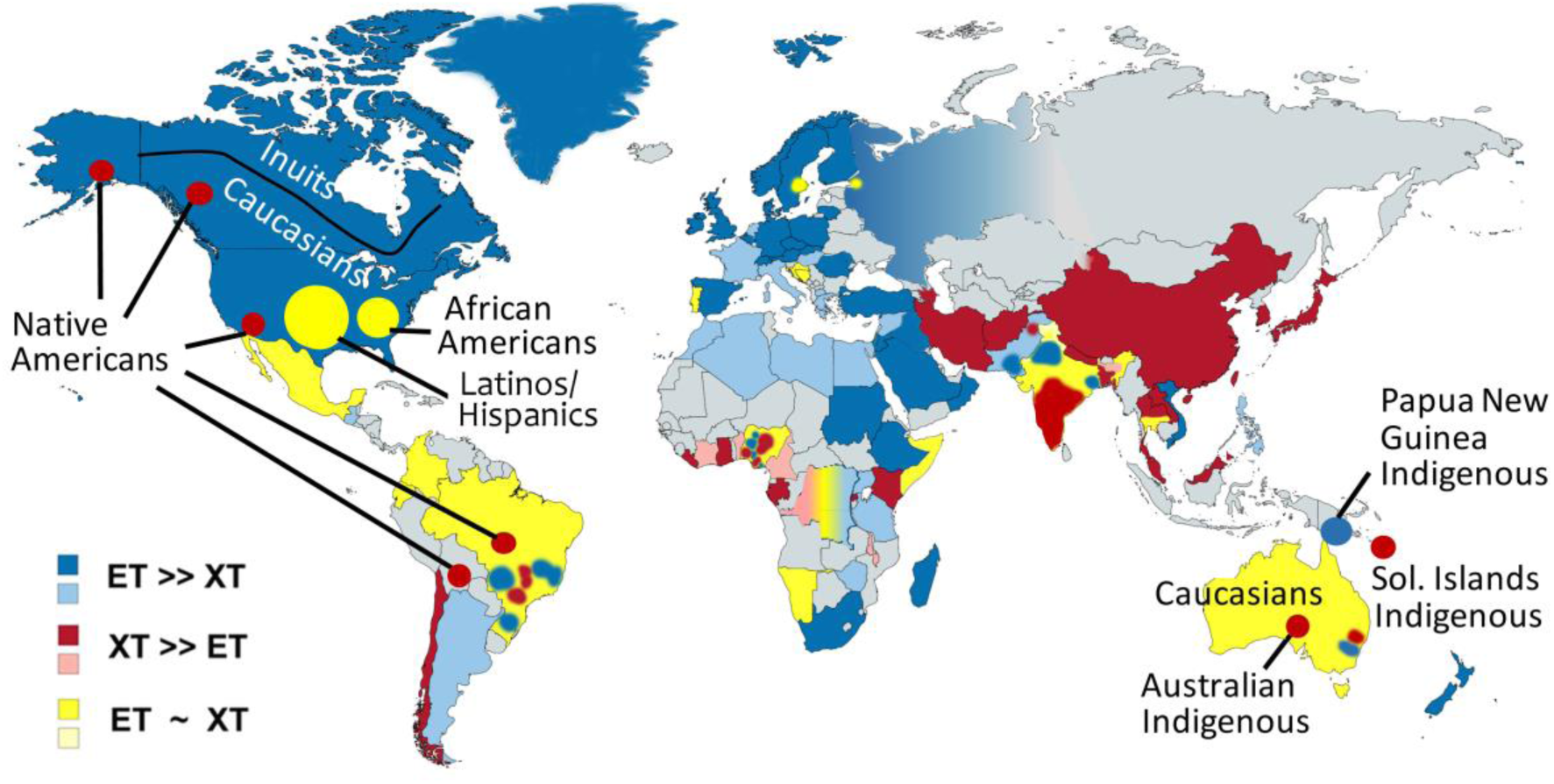
World map of the esotropia/exotropia ratio (ET/XT ratio) among countries. These data were compiled from 295 population-based cohorts in 76 countries (Supplemental Table S1) and 360 clinic-based studies from 72 countries (Supplemental Table S2). ET prevalence much larger than XT prevalence (ET/XT ratio >1.5) is shown in blue; ET prevalence much smaller than XT (ET/XT ratio <0.67) is shown in red; ET similar to XT prevalence (ET/XT ratio of 0.67 – 1.5) is shown in yellow. The relative frequencies of ET and XT from clinic-based studies are indicated by light blue, pink, and light yellow, corresponding to the prevalence ratios and color scheme. For countries with multiple ethnicities, only ethnicity-specific data are shown. Note that ET predominates in countries with mostly Caucasian populations, while XT predominates in East Asia, Southeast India, West Africa and Persia. Sol. Islands, Solomon Islands. Map lines do not necessarily depict accepted national boundaries.

#### 7. East Asia

We found 61 studies that reported on East Asians (63 cohorts), published between 1954 and 2021. We included the Nepalis among the East Asian category, because gene flow into Nepal occurred primarily via Tibet rather than via northeast India.^374,820^ Most studies (n=24) in East Asia were from China,^142,144,206,230,261,323–325,375,378,432,449,454–456,459,604,620,705,821,822,864,866,867^ followed by twelve studies from Japan,^282,314,360,473,493,501–503,554,555,687,844^ six from Nepal,^563,607,724,725,727,728^ six from South Korea,^311,443,444,639,855,856^ four from Malaysia,^148,278,630,769^ four from Thailand,^371,419,470,763^ two from Singapore,^150,151^ one from from Laos,^130^ and one from Vietnam.^611^ East Asians have an ET prevalence of 0.309% (0.2595–0.3674% CI), and an XT prevalence of 1.1395% (0.8976–1.4456% CI). East Asians have an ET/XT ratio (0.27) that is ten times smaller than that in Caucasians (2.83). East Asians thus have one of the highest XT prevalences and lowest ET prevalences when compared with other ethnicities. Recent upward trends of XT prevalence in China and Nepal will be discussed in section 3.12.ii.

#### 8. Oceania

We found 19 population-based studies (reporting on 30 cohorts) from Australasia and Melanesia, published between 1955 and 2016 in five countries: Australia,^110,345,447,465,481,484,665,666,677,A,V,W^ Fiji,^G^ New Zealand,^56,485,732^ Papua-New Guinea,^X^ Tokelau Islands,^216^ and Solomon Islands.^802^ Because of the multiple ethnicities involved (and the differences between these ethnicities), the prevalences are compiled separately by ethnicity in Supplemental Table S1. The prevalence of horizontal strabismus is very low in indigenous populations in Australia and South-East Asia (Philippines, Malaysia). The prevalence of XT (0.222%, 0.0944–0.5214% CI) is larger than that of ET (0.134%, 0.0585–0.3065% CI). A single study of indigenous people in Papua-New Guinea revealed a larger ET prevalence (0.1%) than XT prevalence (0.04%). Caucasians in Oceania (n=7 studies) have an ET prevalence that is lower (1.676%) than in Europe (2.064%), but similar to that of Caucasians in North America (1.553%). East Asians in Oceania have prevalences very similar to those of East Asians residing in East Asian countries.

### 3.3 Comparison of Ethnic Differences in ET and XT Prevalences

The results of the ET and XT prevalences for the different ethnicities from Supplemental Table S1 are summarized in Figure 3. We compare the cohort-size weighted ET and XT prevalences for the 13 main ethnicities in a bar graph, arranged by decreasing ET/XT ratio. Some ethnicities have much more ET than XT, some populations have a similar ratio, and some ethnicities have much more XT than ET (Fig. 3).

Note that ethnicities do not only differ in the relative differences in ET and XT prevalence and ratios, but also in the absolute percentages of horizontal strabismus. Africans, South Asians, and most indigenous people of Oceania have much lower horizontal strabismus prevalences, especially of ET, than the other ethnicities. A low horizontal strabismus prevalence is further supported by studies that did not distinguish ET and XT prevalences, such as for Africans in Tanzania,^825^ Egypt,^214^ Ethiopia,^252,516,712^ Ghana,^576^ Nigeria,^42,759^ Rwanda,^706^ African Americans in North America,^781^ and also for South Asians.^74,140,188,297–299,405,424,425,467,556,567,598,629,721,722,729,734,788,789,791^ Latinos/Hispanics and Middle Eastern populations have an intermediate prevalence of horizontal strabismus, higher than the above-mentioned populations, but lower than in Caucasians and East Asians. This conclusion is further supported by studies that, due to a lack of distinction between ET and XT prevalences, did not qualify for inclusion in the meta-analysis.^180,181,670,812,829,853^

### 3.4 World Map of the Horizontal Strabismus Prevalence (ET/XT Ratio)

As mentioned above, the studies on ET and XT prevalence can be sorted into three types based on the ET/XT ratio: Those with a much larger ET than XT prevalence (type 1 – blue: >1.5 ET/XT ratio), those with a much larger XT than ET prevalence (type 2 – red: <0.67 ratio), and an intermediate type where XT and ET are approximately equal (type 3 – yellow: ratio of 0.67–1.5). Using the geographic data compiled in Supplemental Tables S1 and S2, we generated a world map of horizontal strabismus prevalence. The highest ET/XT ratio is in countries with mostly Caucasians, but also in parts of Africa, the Middle East, Pakistan, and Northwest India, while the lowest ET/XT ratio is among East Asians, Native Americans, Persians, South Asians in the Southeast of India, and populations in West Africa (Fig. 4).

### 3.5 Geographic Distribution of ET and XT Prevalence – Is there an Effect of Latitude?

Previous work suggested that geography/latitude may be responsible for, or contribute to, the differences in prevalence between studies, presumably because the amount and pattern of sunlight can affect various ocular parameters.^69,75,100,109,254,344,372,436,461,481,497,499,512,566,673,810^ Therefore, we examined whether latitude is associated with the ET/XT ratio. To minimize the confounding variable of ethnicity, we compared latitude within those ethnicities that distribute over a major north-south expansion. Latitudinal ranges of the ethnicities we were able to assess were from 60.2° N to 38.7° N (Europeans), 37.7° N to 3.1° N (East Asians), and 64.3° N to 12.7° S (Native Americans). We did not find any significant trends or correlations: R^2^ values of 0.0003 and 0.0004 for Europeans and East Asians, respectively, (Fig. 5A-B) and an R^2^ value of 0.0596 for Native Americans (but this reduced to 0.0004 when one outlier study was removed from the regression analysis, data not shown). We also confirmed the latitude data (from 27° S to 46° S) collected by Lance and Mitchell^436^ for Australians – nearly all of them Caucasians. We conclude that latitude has no effect on the ET/XT ratio. For a plausible alternative explanation for increased XT in Caucasians living outside of Europe, see section 4.2.

**Fig. 5A-B.**
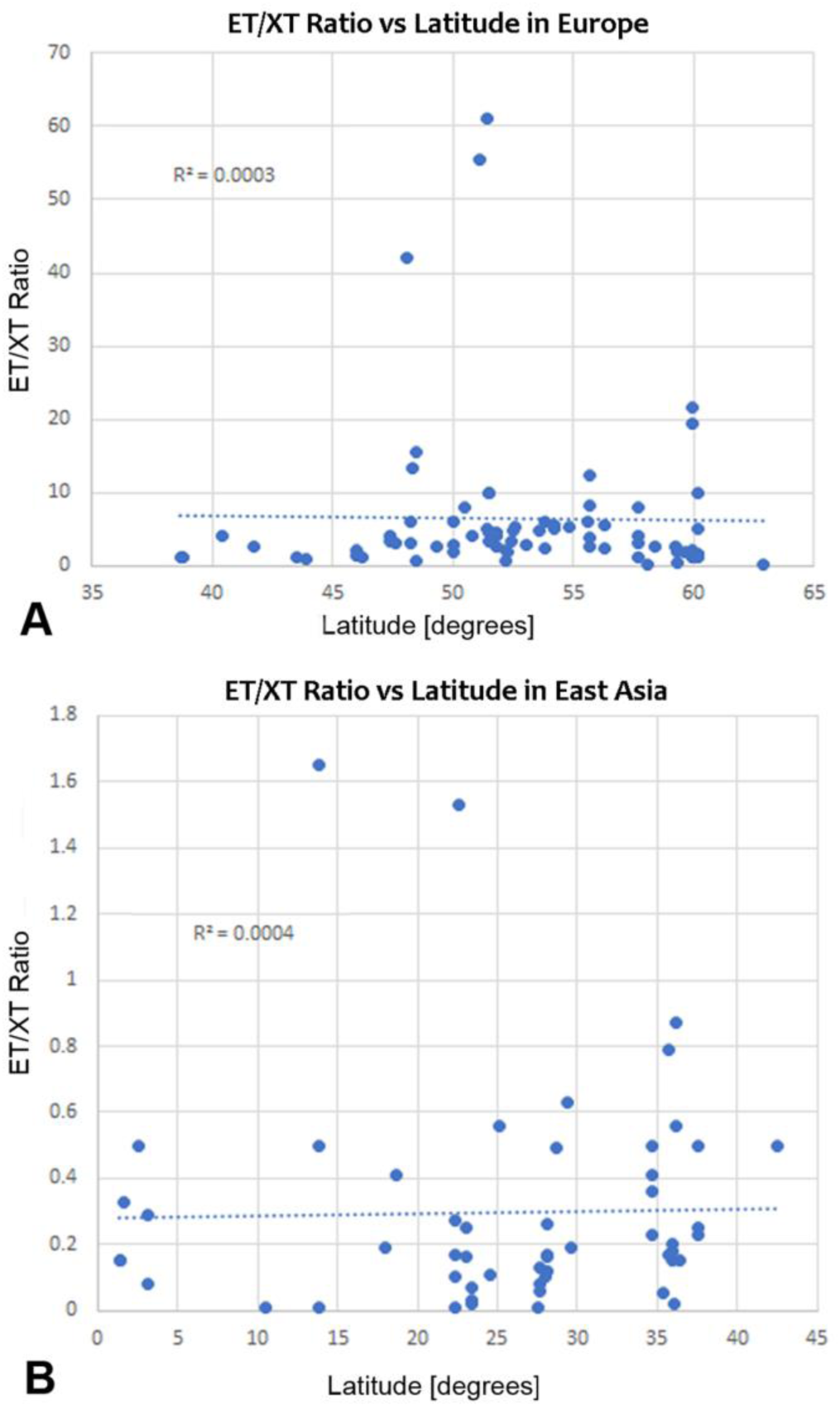
The graphs show that there is no association between the esotropia/exotropia (ET/XT) ratio and latitude. Panel A shows the data and flat trend line for Caucasians in Europe, panel B shows a flat trendline for East Asians. The coefficient of determination (R^2^ values of 0.0003-0.0004) is indicated on the graphs.

### 3.6 Changes in ET and XT Prevalence and Frequency with Age

Previous investigators suggested that differences in the ages of cohorts may explain, to some extent, the variations in results between studies and populations.^251,287,318,575,A^ To determine the extent to which age may be responsible for the prevalence differences between studies on different ethnicities, we compiled the age ranges for study cohorts and plotted them for several of the major ethnicities (Fig. 6A-F). The large majority of studies we reviewed concentrated on the age range of 3-12 years (n=273/288, 94.8%), which includes the age when the peak of prevalences has been reached, and it is prior to the drop-off of ET prevalence around puberty/adolescence.^90,166,251,268,429,444,826^ When the studies that focused only on adults, from various ethnicities, were removed from the ethnicity analysis (n=13/288), our results did not change, indicating that the (relatively few) studies reporting on adults-only are not responsible for the differences between ethnicities.

**Fig. 6A-F.**
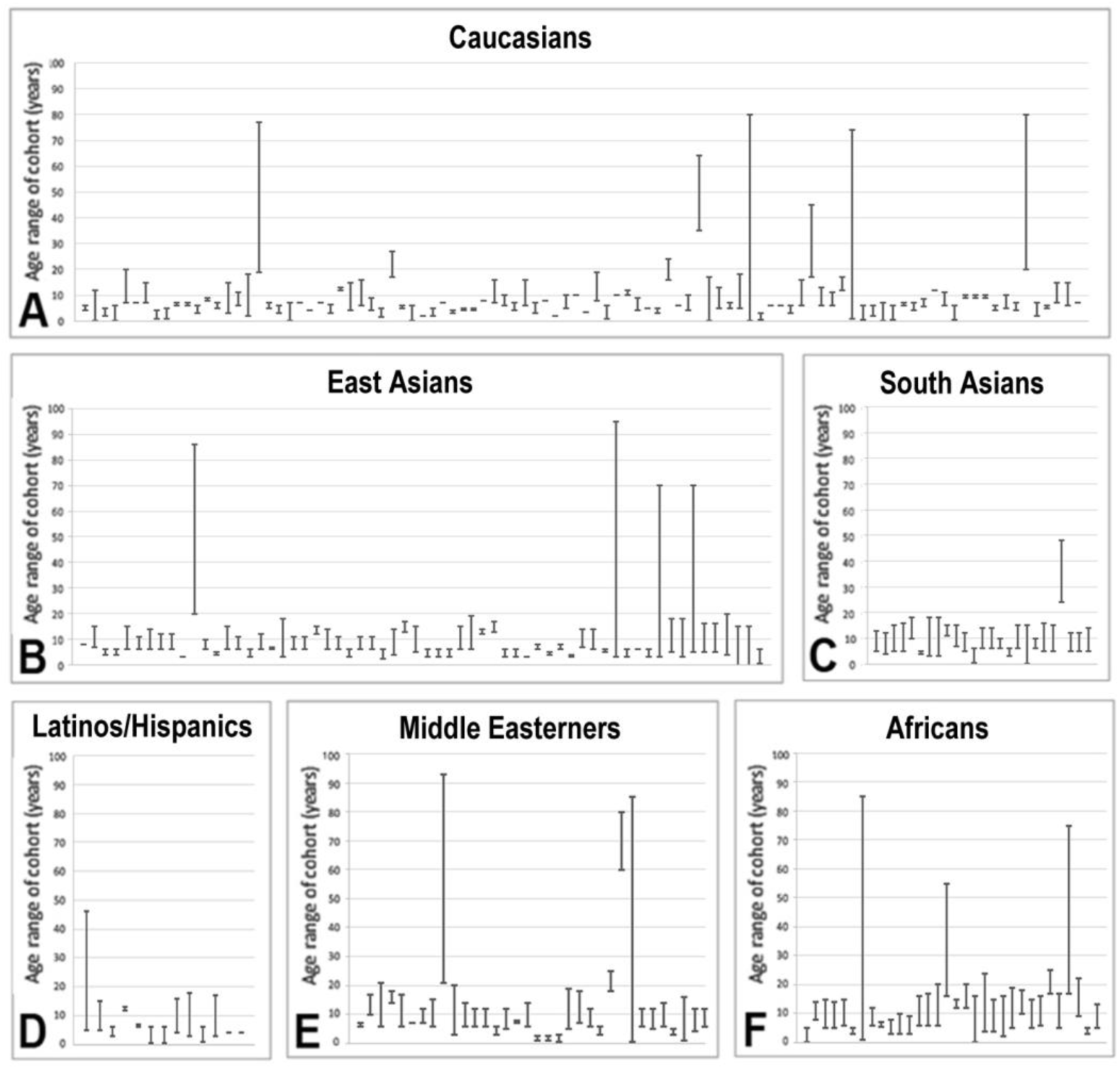
The panels show the age ranges for each of the population-based studies compiled for the major ethnicities in Supplemental Table 1. The large majority of studies focused on the age range of 3-12 years when children have maximal prevalences of either esotropia (ET) or exotropia (XT), and this was confirmed by conducting the meta-analysis with and without the “adults only” studies. Studies in Latin America from regions with mostly Caucasian ancestry were not included in the panel D (Latinos/Hispanics).

Our review of studies that compared multiple age groups showed that the prevalence of horizontal strabismus differs with age in distinct patterns.^90,142,166,170,250,251,268,286,287,318,429,443,444,502,503,510,574,575,771,857,D,I^ Prevalence of both ET and XT is lower in the first three years of life, increases subsequently, and most studies report that ET prevalence decreases during or after puberty.^907,166,251,268,429,444,826^ In Caucasians in Europe and North America, ET prevalence was 1.32% in ages 0.5 to 3 years (n=8 studies), which is about half of the ET prevalence of 2.26% at 4 years and older (n=89, p=0.014, unpaired t-test). Likewise, XT prevalence was 0.47% in the younger age group (n=8), compared to 0.95% in the older age group (n=88, p=0.012). The likely reasons for some of these changes with age are discussed in section 4.2.

Most studies report that XT prevalence increases in the elderly,^178,218,231,251,282,356,469,658,671,D^ but there is some discrepancy between studies whether the prevalence of ET or XT changes already in middle-aged adults, or whether XT is more frequent (and ET less frequent) only in older adults. In East Asians, there is only a small further increase of XT in older age.^444^ In our review, we found a total of 15 studies that reported ET and XT prevalences specifically in adults or in adults and adolescents, in several ethnicities: Caucasians, African Americans, West Africans, East Asians, South Asians, and Persians.^12,68,97,200,216,241,282,319,320,331,341,356,742,744,D^ In 11 of the 15 adult cohorts, ET prevalence was lower, while it was higher in three studies, and there was no difference in one. In eight cohorts, XT prevalence was lower in adults, while in six other cohorts, it was higher, and in one there was no difference when compared with the prevalence in children of the same ethnicity.

In the population-based studies that specifically examined adults 55-60 years of age or older, for Caucasians^356,D^ and African Americans,^D^ and for Persians,^317,320^ the XT prevalence was 2-3fold higher than in cohorts of younger-age adults (p=0.0146, paired t test). Furthermore, when we examined the ratio of ET/XT in adults 60 years or older from five relevant clinic-based studies on Caucasians,^178,218,231,469,658^ we found that in those five studies, the proportion of XT was much higher (49.3%) than would have been expected based on the XT proportion at younger ages in the same ethnicity (26.8%, n=103), and this difference of the proportion of XT was highly significant (p=0.00040, unpaired t-test).

### 3.7 Gender Differences in Horizontal Strabismus

Can gender differences in the prevalence of strabismus explain differences between reports? While there are some reports of a gender difference in horizontal strabismus, mostly of a female bias towards XT,^128,157,170,173,286,291,340,382,404,422,578,579,815^ nearly all of these studies were clinic-based and had relatively small cohorts. Our recent systematic review and meta-analysis of gender in horizontal strabismus (based on 73 population-based studies)^441^ found no difference in the prevalence (2.56% for males, 2.58% for females) – confirming the conclusions from large-cohort population-based studies^147,343,443^ that there is no gender difference in prevalence of strabismus, although there is a significant female bias in the clinic.^441^

### 3.8 The ET/XT Ratio according to Clinic-based Studies: Comparison with Population-based Studies

While clinic-based studies do not inform about the prevalence,^390^ they allow investigators to obtain information about the numbers of patients that seek and receive medical attention at a clinical facility – thus providing information about the relative frequency of ET and XT seen in the clinic. We compiled such information from 360 studies conducted in 72 countries (after excluding 12 studies that reported on the same cohort and two studies that reported exclusively on subjects two years of age or younger, Supplemental Table S2). In Europe, we found 72 clinic-based studies; from Austria,^699^ Bosnia,^415,654^ Denmark,^251,292,648,778^ Finland,^494,646^ France,^165,355,622,634,638,640^ Germany,^193,340,391,703^ Greece,^154,805,868^ Hungary,^746^ Ireland,^359,628,Y^ Italy,^127,265,754^ Netherlands,^173,382,399,792,Z^ North Macedonia,^AA^ Poland,^47,422^ Portugal,^253,284,538^ Romania,^527,BB^ Russia,^794^ Spain,^519,520,CC^ Sweden,^18,315,417,618^ and the UK.^23,112,128,131,178,224,273,344,363,401,464,513,542,559,621,747,753,771,828,840^ There were 59 studies from North America, with 39 studies on Caucasians,^17,28,85,93,108,119,169,218,221,231,239,247,286,289,321,361,362,451,469,492,532–534,619,623,644,658,660,676,696,697,704,772,824,832,859,DD^ one study on Native Americans,^25^ six on African Americans,^279,451,623,644,691,859^ four on Asians,^361,362,623,644^ six on mixed or multiple ethnicities,^100,260,361,362,540,707^ and four on Latinos/Hispanics.^346,623,644,EE^ We found 27 studies in Latin America, on Native Americans,^574,FF^ Caucasians,^517,543,FF^ Latino/Hispanic or mixed race,^21,191,198,294,389,431,468,521,528,529,587,597,600,617,668,669,671,799,869,GG,HH,II^ and populations of African descent.^460^

There were 24 studies from North/East/South Africa on Africans/Arabs,^3,50,98,212,213,215,264,304,488,517,541,569,577,681,723,773,860,FF,JJ,KK,LL,MM^ on Caucasians,^109^ and mixed ethnicities,^773^ and 28 studies from West Africa.^20,75,86,103,106,160,194,199,203–205,406,547–550,570,579,580,585,589,591,613,NN,OO,PP,QQ,RR^ We found 45 studies from the Middle East.^1,3,4,30,36,37,43–45,46,48,51–53,57,83,88,123,124,136,174,175,184,222,234,281,302,352,358,366,397,404,514,553,571,588,637,643,651,661,682,850,851,SS,TT^

There were 41 studies from the Northwest of South Asia, with one study from Afghanistan,^54^ twelve studies from Northwest India,^38,277,471,495,518,525,680,686,689,711,735,797^ and 28 studies from Pakistan.^4,5,58,64,77–78,107,139,141,338,364–365,385–386,472,478–480,664,708–710,713–715,717–718,730^ We found 19 studies on South Asians in Southeast India.^60,69,96,133,149,157,189,192,267,283,300,328,558,561,641,784,795,804,823^

In East Asia, we found 17 studies from China,^102,135,143,145,190,207,262,322,376,377,438,448,783,819,852,857,858^ two studies from Indonesia,^233,636^ twelve from Japan,^263,426,476,498,500,503–504,517,526,584,761,FF^ seven from South Korea,^137,156,310,373,408–409,453^ and eleven studies from Malaysia, Phillipines, Singapore, Thailand or Vietnam.^118,138,148,149,367,392,452,606,653,770,816^ In Oceania, we found an additional nine studies.^56,202,274,344,436,482,601–602,W^

Our analysis shows that the estimated population-based ET frequency was lower (overall by 13.8%) than the frequency obtained from the clinic-based studies (Fig. 7). Conversely, the population-based XT prevalence was higher than the clinic-based estimate (by 13.8%). In other words, clinic-based studies underestimate XT frequency, compared with population-based studies (Fig. 7A). This pattern was consistent throughout all ethnicities, although the margins varied: The ET frequency in clinics was only slightly higher than in population-based studies when the population had a large ET/XT ratio, while the deficit for XT in the clinic was substantial in ethnicities with large fractions of XT (23.3% difference in Persians, 17.0% in Latinos/Hispanics, and 15.2% difference in West Africans, Fig. 7B). This suggests that people with XT present less in clinics than people with ET; patients with XT (or their parents) may feel less of an urgency to visit clinics than patients with ET.

**Fig. 7A-B.**
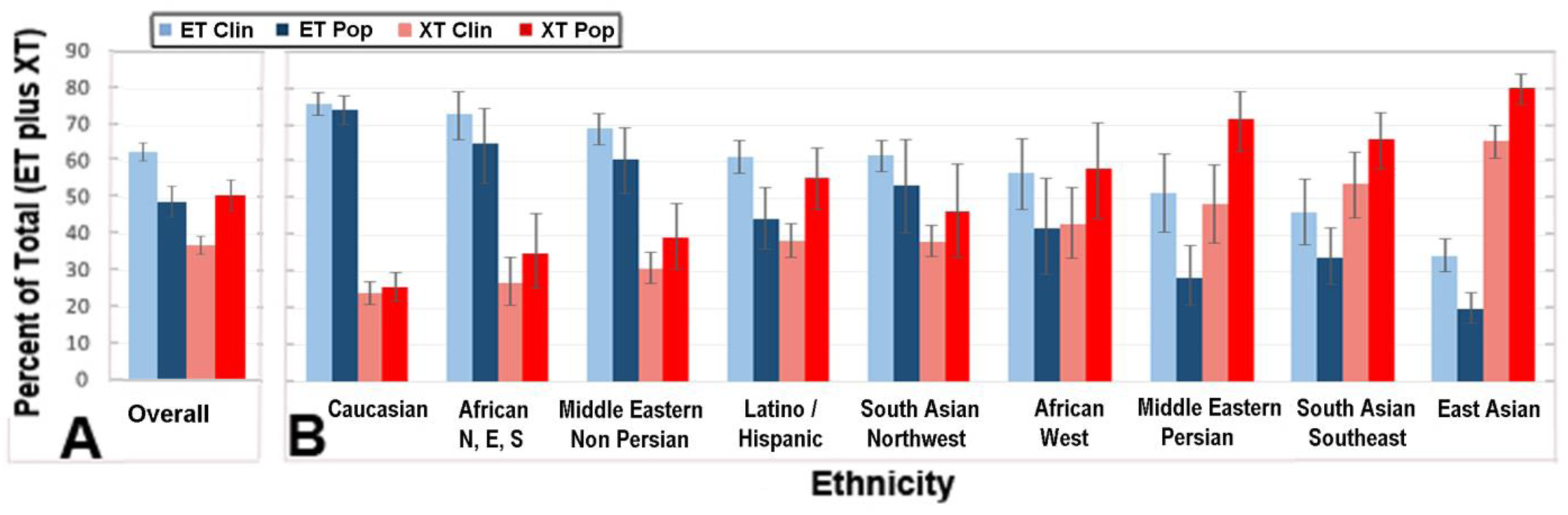
The bar graphs compare the contribution of esotropia (ET) and exotropia (XT) to the total horizontal strabismus. A. overall (for all ethnicities, weighted by population size – see Table 3); and B. for each of the major ethnicities with sufficient data for comparison. Each subpanel shows the percentages of total horizontal strabismus for different ethnicities due to ET (blue) and XT (red), obtained from population-based studies. The graph furthermore compares the percentages obtained through population-based studies (darker color) with those from clinic-based studies (light blue and pink colors). The error bars are 95% confidence intervals. Overall, the clinic-based studies underestimate by about 14% the frequency of XT, suggesting reduced XT presentation in the clinic compared to ET. Abbreviations: N, North; E, East, S, South.

### 3.9 Impact of Studies reporting Prevalence in Different Ethnicities

We found many more population-based studies on Caucasians (n=115), most of them in Europe, than on any of the other major ethnicities (for which n ranged from 2-61 studies, Fig. 8A). Only Caucasians have been consistently examined over several (five) generations, and East Asians for the last two generations (Supplemental Table S1, Fig. 8A). We also found that the population-based studies on Caucasians (especially in Europe and North America) have had significantly more impact – being cited disproportionally, approximately 3-fold more ∼ 40% of all citations) than their equitable share of the world population (which is ∼15% for Caucasians, United Nations).^H^ Nearly all other ethnicities were underrepresented in the number of studies and their impact, especially populations in Africa and in South Asia (Fig. 8).

**Fig. 8A-B.**
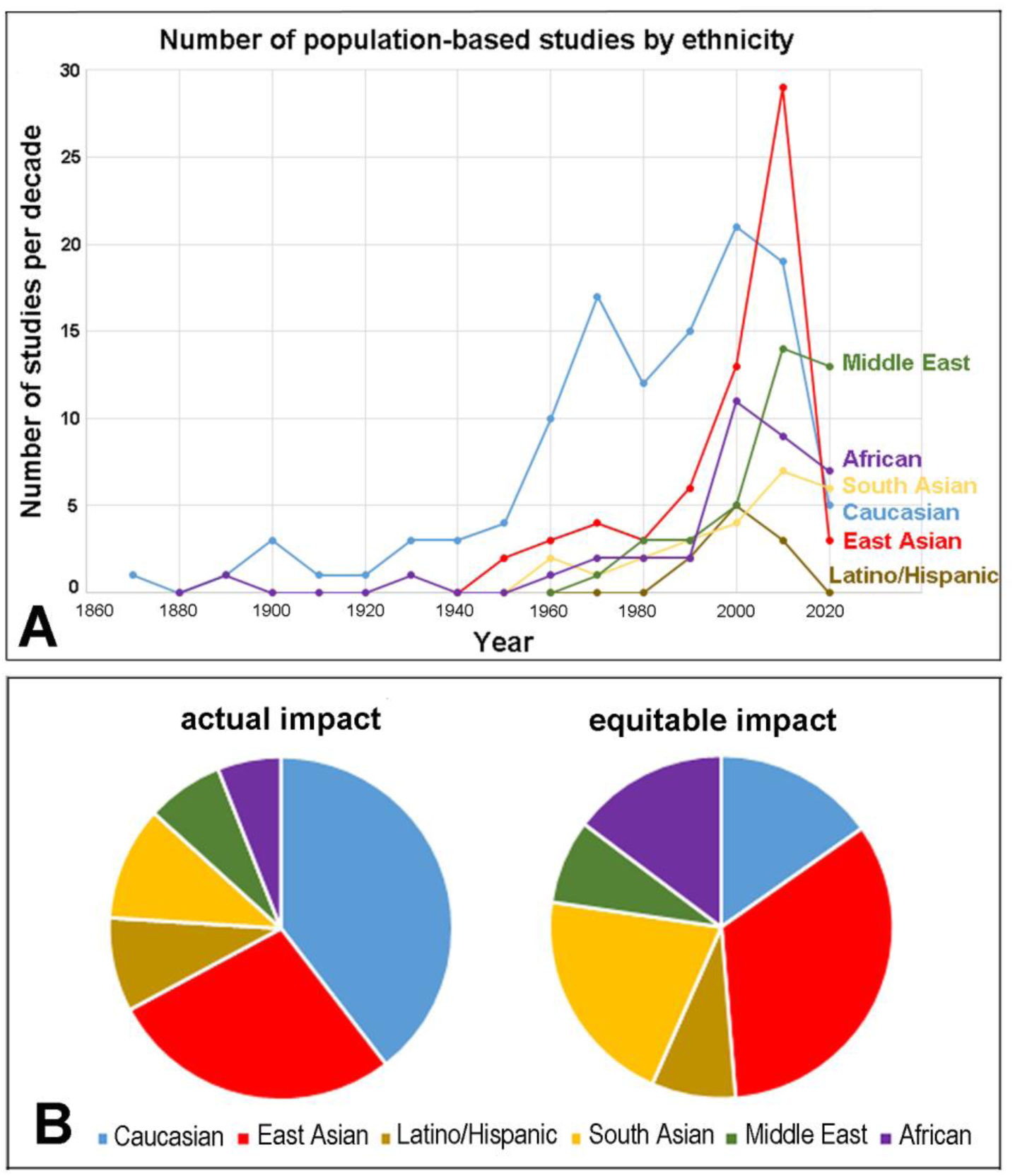
Number of studies over time and their impact by ethnicity. A. The number of population-based studies per decade is indicated between 1860 and 2025, for the largest six ethnicities (Caucasian, East Asian, South Asian, African, Middle East and Latino/Hispanics). Note that only two ethnicities (Caucasians and East Asians) have sufficiently large numbers of studies per decade to allow longitudinal analyses over several generations (see Fig. 10A,B). B. Citations of studies surveying different ethnicities were compiled on October 4-5, 2025 based on the Web of Science, Core collection.^F^ The pie chart shows that the Caucasian impact is overrepresented by three-fold compared with the actual contribution to the world population, which explains, in part, why the prevalence of horizontal strabismus, and especially the prevalence of esotropia (ET), was significantly overestimated. Only 1/3 of the world population has more ET than exotropia (XT), but based on citations of studies, about 2/3 of the impact was from studies that examined populations with ET exceeding XT.

Specifically, Caucasian’s impact is at 313.0% of equitable impact, Hispanics/Latinos are at 117.9%, East Asians at 73.3% of equitable impact, South Asians at 52.9%, Middle Easterns at 51.3%, and Africans trail at 34.5% (Fig. 8B), when citations are measured as a proxy for the relative impact (equitable impact = 100% of the contribution of that ethnicity to the world population). As can be seen in Figure 8A, the number of studies for neglected ethnicities (non-Caucasians) has been increasing in the last 1-2 decades, so the current imbalance is likely to improve in the future.

### 3.10 Role of Socioeconomic Status (SES)

We found several studies that attempted to explore whether SES is associated with horizontal strabismus, but results were inconsistent: some investigators reported a higher strabismus prevalence or frequency of diagnosis/treatment with lower SES,^210,317,443,462,466,522,609,771,A,UU^ while others found no association between SES and horizontal strabismus prevalence or frequency,^163,287,311,443,665,700,805^ and one study reported very low strabismus prevalence with low SES.^177^ When ET and XT were assessed separately, results also varied: one study reported a *higher* ET prevalence with lower SES,^771^ while another one found a *lower* ET prevalence with lower SES,^833^ and several studies reported a higher XT prevalence with lower SES.^147,318,A,B^ The potential confounding of SES and ethnicity in the context of ET and XT prevalence will be discussed in Section 4.4.

### 3.11 Estimation of the Effect Size of Biases

To assess potential biases in the prevalence analyses, we followed three strategies, including a comparison with so-called “gold standard” (expert) studies, a comparison with birth-cohort studies, and an analysis of funnel plots for publication bias and meta-analysis bias.

#### i. Comparison with “Gold Standard” Studies according to Sharbini (2015)

^A^ Some investigators have argued that population-based studies in which a highly qualified examiner administers the tests for strabismus would be more accurate or valid, and they recommend considering only those studies with such a “gold standard” status.^A,B^ When we compared the results of “gold standard” studies with the overall results of ethnicity-matched studies, we did not find any statistically significant differences between the two groups using unpaired Student’s t-tests.

<u>For Caucasians</u> in Europe, ten “gold standard” studies were identified by Sharbini (2015)^A^: ^10,61,287,394,416–417,428,429,787^ These gold standard studies had an ET prevalence of 2.26 ± 0.26% (SEM) and an XT prevalence of 0.76 ± 0.19%. The non-gold standard studies for Caucasians (n=51) had an ET prevalence of 2.64 ± 0.22%, and an XT prevalence of 0.67 ± 0.08%. We found no statistically significant difference between the two types of studies (Student’s t-test, in both cases p > 0.10).

<u>For East Asians</u>, there are eight “gold standard” studies according to Sharbini (2015)^A^:^150,323–324,432,459,501,563,705^ They have an ET prevalence of 0.31 ± 0.06% and an XT prevalence of 1.25 ± 0.28%. The non-gold standard studies we compiled for East Asians (n=37) had an ET prevalence of 0.31 ± 0.04%, and an XT prevalence of 1.16 ± 0.17%. There was no statistically significant difference between the two types of studies (Student’s t-test, in both cases p > 0.40).

<u>For Middle Eastern (non-Persian) Populations</u>, Sharbini (2015)^A^ identified four “gold standard” studies,^16,89,256,457^ with an ET prevalence of 0.96 ± 0.35% and an XT prevalence of 0.44 ± 0.20%. The non-gold standard studies for Middle Eastern populations (n=7) had an ET prevalence of 1.08 ± 0.15%, and an XT prevalence of 1.01 ± 0.36%. Like the other ethnicities tested, we found no statistically significant difference between the two types of studies (Student’s t-test, in both cases p > 0.10).

<u>For all other ethnicities</u> (Africans, South Asians, Latinos/Hispanics and Native Americans), there are either too few “gold standard” studies or too few control studies to allow formal statistical analyses (in all cases n≤3). Nevertheless, based on the three ethnicities where such an analysis was possible, we conclude that there is no significant difference between gold standard and non-gold standard studies, and therefore one should not restrict analyses exclusively to so-called gold standard studies.

#### ii. Potential Sampling Biases: Neglect of subjects with major disabilities

One potential problem of prevalence studies (as explained in section 2.5.2) is the concern that children with major disabilities may be missed in school-based studies, because they attend special schools or no schools at all, resulting in an undercount of strabismus cases. This bias is not present in birth-cohort studies where all children are tracked. When we compared the strabismus prevalence in birth-cohort studies in Caucasians (n=13),^65,147,287,394,420,428,491,575,624,649,732,748,833^ with otherwise comparable school-based studies, there was indeed a 15% higher prevalence in the birth-cohort studies (data not shown), suggesting that about 20 million children worldwide may be missed in estimates derived from school-based studies. Such numbers are supported by studies that included subjects from schools for disabled children^187,251^ and by recent estimates of the global number of strabismic children with cerebral palsy^333^ or Down syndrome.^808^ These are just two of the many disabilities that may prevent normal school attendance. Accordingly, current estimates of the global number of strabismus cases may be too low by 15%.

#### iii. Bias in Very Large Cohort Studies?

In general, studies with the largest cohorts are considered the most reliable for clinical trials, while those with very small cohorts are more likely to generate outliers.^751^ However, we noticed that in some of the ethnicities (South Asians, Caucasians, African Americans), the largest cohort studies reported a strabismus prevalence that was significantly lower than the pooled prevalence estimate. For example, total horizontal strabismus in South Asians in Southeast India was 0.26% according to the three largest cohort studies,^177,400,733^ while the pooled estimate was 0.90%, a significant difference (p=0.02). Likewise, the largest cohort studies in Europe^343,599^ and North America^200^ reported 2.15%, 1.29%, and 2.07% horizontal strabismus for Caucasians, while the pooled estimates for Caucasians in Europe and North America were 2.69% and 2.49%, respectively, again a significant difference (p=0.008), and similarly for African Americans where Traboulsi et al.^781^ reported 2.1%, while the pooled estimate was 2.61%. Therefore, unlike most other types of clinical studies,^751^ in prevalence studies of horizontal strabismus, the largest cohorts may not necessarily provide the most reliable results.

#### iv. Potential Publication Bias and Meta-Analysis Bias

*Analysis of Funnel Plots*. Since one needs a sufficient number of studies to interpret funnel plots,^440^ we focused on Caucasians and East Asians – the ethnicities with the largest numbers of studies. We first plotted the data from Caucasians in Europe for ET prevalence in a funnel plot (Fig. 9A). The plot shows a fairly symmetric distribution around the pooled estimate of 2.1%. There was no major or obvious lack of data points in the funnel, indicating that publication bias was minimal. When we plotted the data from East Asians for XT prevalence (Fig. 9B), we noticed an anomaly: there were groups of data points with abnormally high prevalences that had in common that they were published in 2014 to 2021, from China^142,144,261,325,456,604,822,864,866^ and from Nepal.^607,725^ These eleven studies showed a much higher prevalence for XT than those of the previous two generations (Fig. 10B, see section 3.12). The recent studies from China and Nepal apparently form a separate sub-group that requires exclusion from the original funnel plot (Fig. 9B).^752^ Consequently, the funnel plot showed that XT prevalence in China’s current generation differs from previous generations, and that this trend is not shared by other East Asian countries except for Nepal (see next section).

**Fig. 9A-B.**
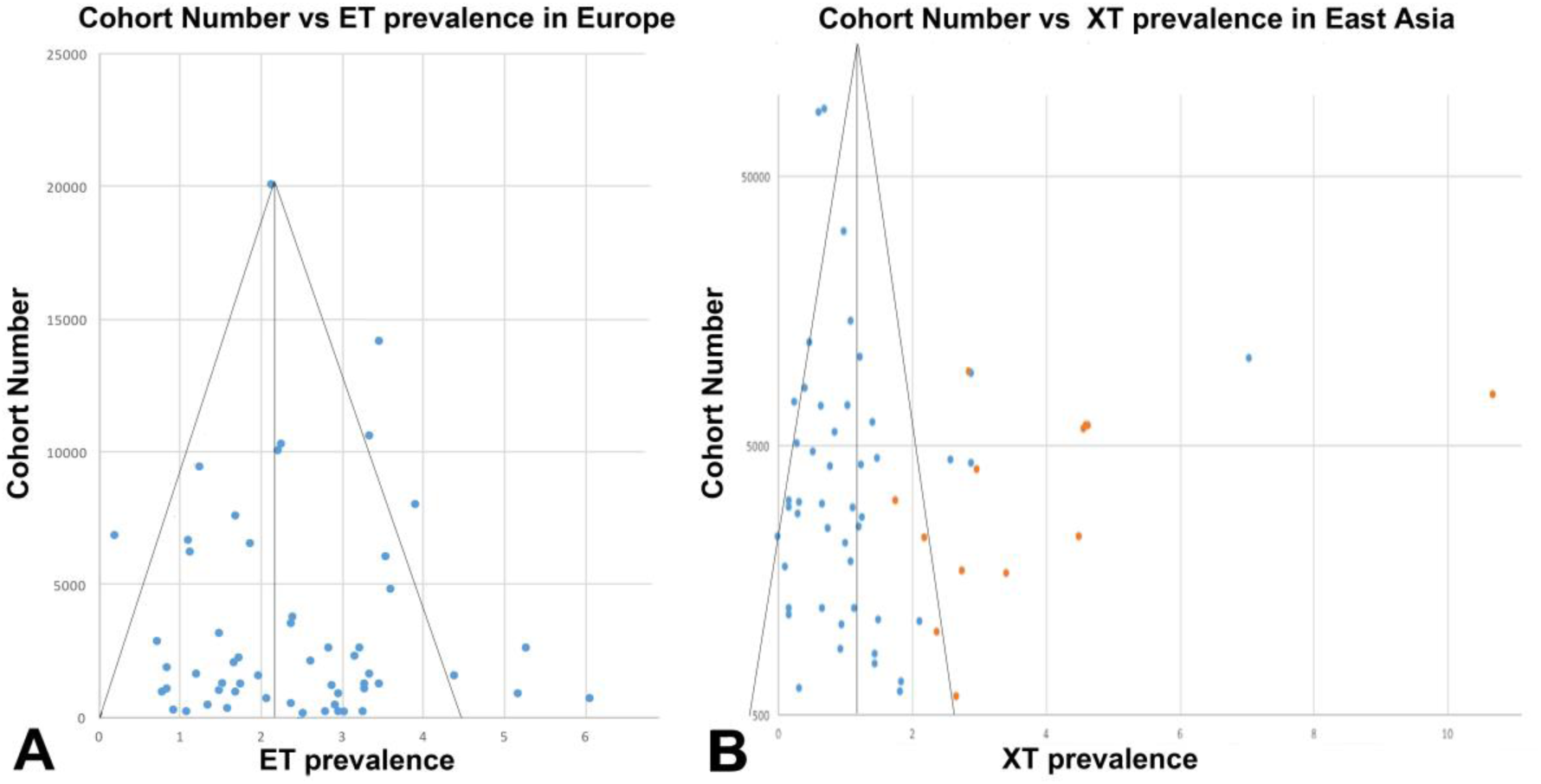
Funnel plots for population-based studies on esotropia (ET) in Caucasians (A) and for exotropia (XT) in East Asians (B). The funnel for ET in Caucasians is symmetric, with very few outliers, suggesting no publication bias. The funnel for XT in East Asians shows the presence of a distinct subgroup: the most recent data (after 2010) from China and Nepal form an apparent subgroup, indicated by orange dots (2011-2025), when compared with studies from other East Asian countries.

**Fig. 10A-B.**
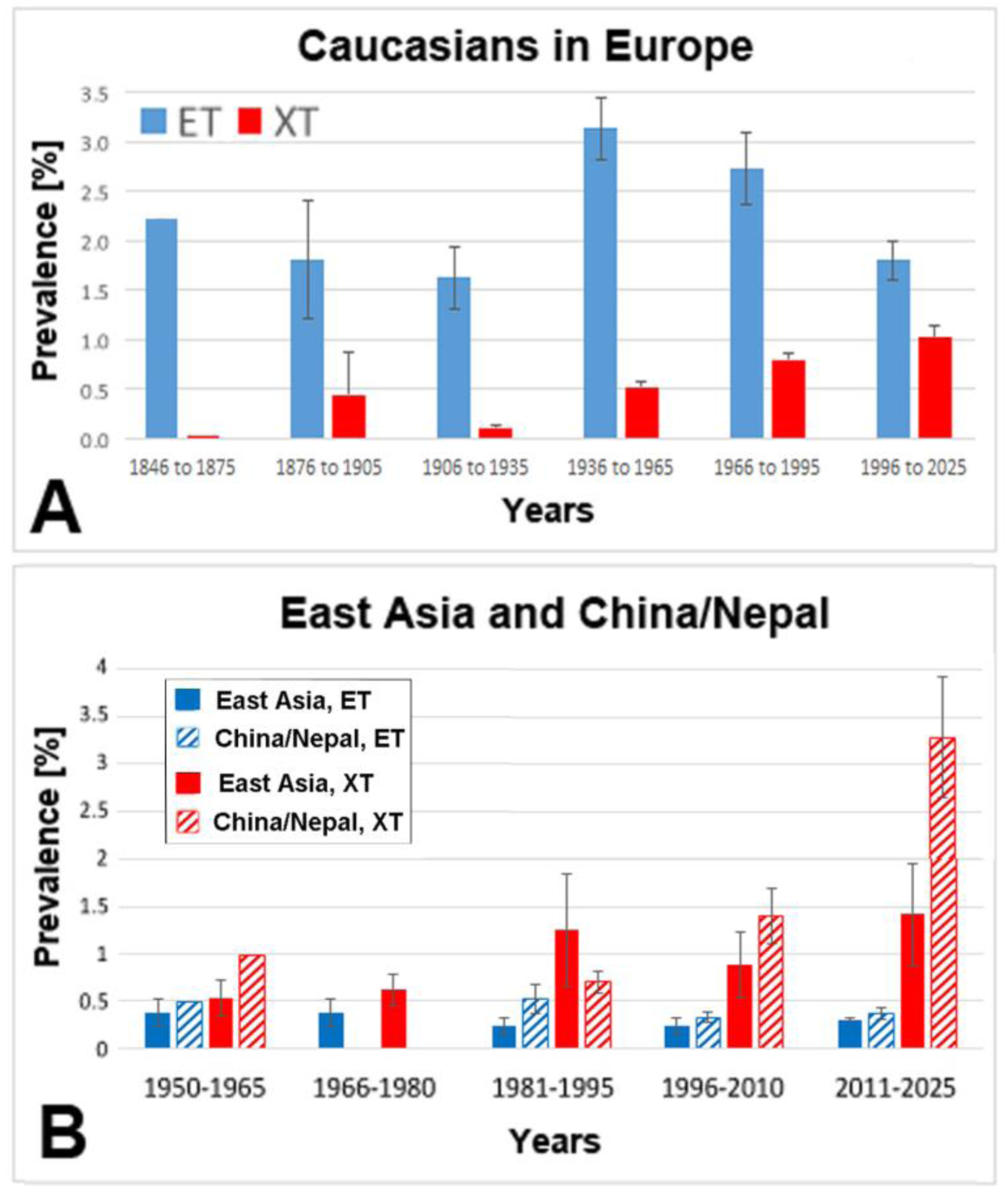
Bar graphs showing secular (generational) differences in esotropia (ET) and exotropia (XT) prevalence among Caucasians in Europe (A) and among East Asians (B). A. Caucasians in Europe had a consistent prevalence of ET between 1.5% and 2% from the 1860s until about 1930, but underwent an ET epidemic from approximately 1960 until 1990, with a doubling of the ET prevalence (blue), and with a subsequent decline to the original base level in the most recent generation (1990-2025); XT prevalence (red), on the other hand, gradually increased from 1860 to 2025. One outlier for XT was removed for the sparse data set of 1876 to 1905 (Tukey’s Hinges with k=1.5). B. All East Asian studies other than those in China and Nepal show a gradual increase in XT prevalence (red) over the last few decades, with a modest decrease in ET prevalence (blue) after 1995. Studies from China and Nepal (hatched bars) in 2011-2025 show a statistically highly significant increase of XT, that is not seen in the other East Asian populations. Error bars are standard errors of the mean (SEM).

### 3.12 Generational Changes in Frequencies and Prevalences of ET and XT

According to clinic-based studies that used longitudinal designs (medical records of clinic visits and surgeries performed in Europe, Canada, USA, Israel, Japan and China), the frequency of ET surgeries has been declining for several decades (from the 1970s to the early 2000s), in some regions by more than 50%,^62,126,158,236,253,332,366,464,596,663,719,824^ while in other regions, especially in East Asia, the frequency of XT diagnoses or surgeries appears to have been increasing.^149–150,376,433,448,498,720,824,855,857^ It has remained unclear whether these trends over decades are due to changes in prevalence of ET, to changes in prevalence of XT, simultaneous changes (but in opposite directions) in both ET and XT prevalences, or whether they are due to other parameters, such as changes in patient behavior, changes in effectiveness of screenings and/or treatments.^62,720,B^ To answer the question about changes of prevalences over time, we compared prevalences in ethnicities with longitudinal data from population-based studies.

i. In Caucasians within Europe, the prevalence of XT gradually increased over the last 2-3 generations (Fig. 10A). The prevalence of ET was stable from the 1860s to 1935, nearly doubled in the generations of 1935-1995, and then decreased back to base levels in the current generation (1996-2025) (Fig. 10A). The number of studies for the period of 1846 to 1935 is relatively sparse, but the ET prevalence of approximately 1.5% to 2.2% is supported by several studies from the UK that did not meet our inclusion criteria because they reported total horizontal strabismus and/or ET/XT ratios for different cohorts.^VV^ Accordingly, there appears to have been an epidemic of ET in Caucasians in Europe between 1935 and 1995, particularly pronounced in the 1950s and 1960s. The ET prevalence then stabilized between the 1990s and 2025 in Caucasians.^332^
ii. In East Asians, ET prevalence does not differ significantly between generations, but in the Chinese and Nepalis, XT prevalence increased dramatically from 1996-2010 to the subsequent period of 2011-2025 (Fig. 10B), suggesting a current XT epidemic among Chinese and Nepalis. In East Asians other than Chinese and Nepalis, the increase in XT prevalence was much more gradual and appears to have stabilized between 1980 and 2025 (Fig. 10A).^426^ Strabismus surgeries already increased in China prior to 2014,^376,377,448^ but there was an acceleration with large increases from 2016 to 2019 (and a COVID-related decrease in 2020).^102,819^ Possible reasons for these two epidemics will be discussed in section 4.2.

### 3.13 Use of Ethnicity-Weighted Contributions for the Estimation of the Global Prevalence of ET and XT

Previous studies have quoted a large range for prevalences of horizontal strabismus, between 0.13% and 9.9% (compiled in Table 1, illustrated in Fig. 1). Generally, the global prevalence is thought to be about 2-4%, but most of these ranges did not take into account ethnic differences or differences between generations. We prepared estimates of the global prevalence of ET and XT – using the cohort-size adjusted % prevalence – by weighing populations (ethnicities) according to their contribution to the world population as published by the United Nations.^H^ We could calculate generational changes for only two ethnicities, Caucasians and East Asians, because there were not enough studies to reliably detect secular differences in other ethnicities. For this reason, only Caucasians and East Asians were assessed differentially for the previous and the current generation, while the prevalences for the other ethnicities are only adjusted for population growth, but we do not distinguish between two different generations – basically assuming no change (Tables 4, 5).

**TABLE 4.**
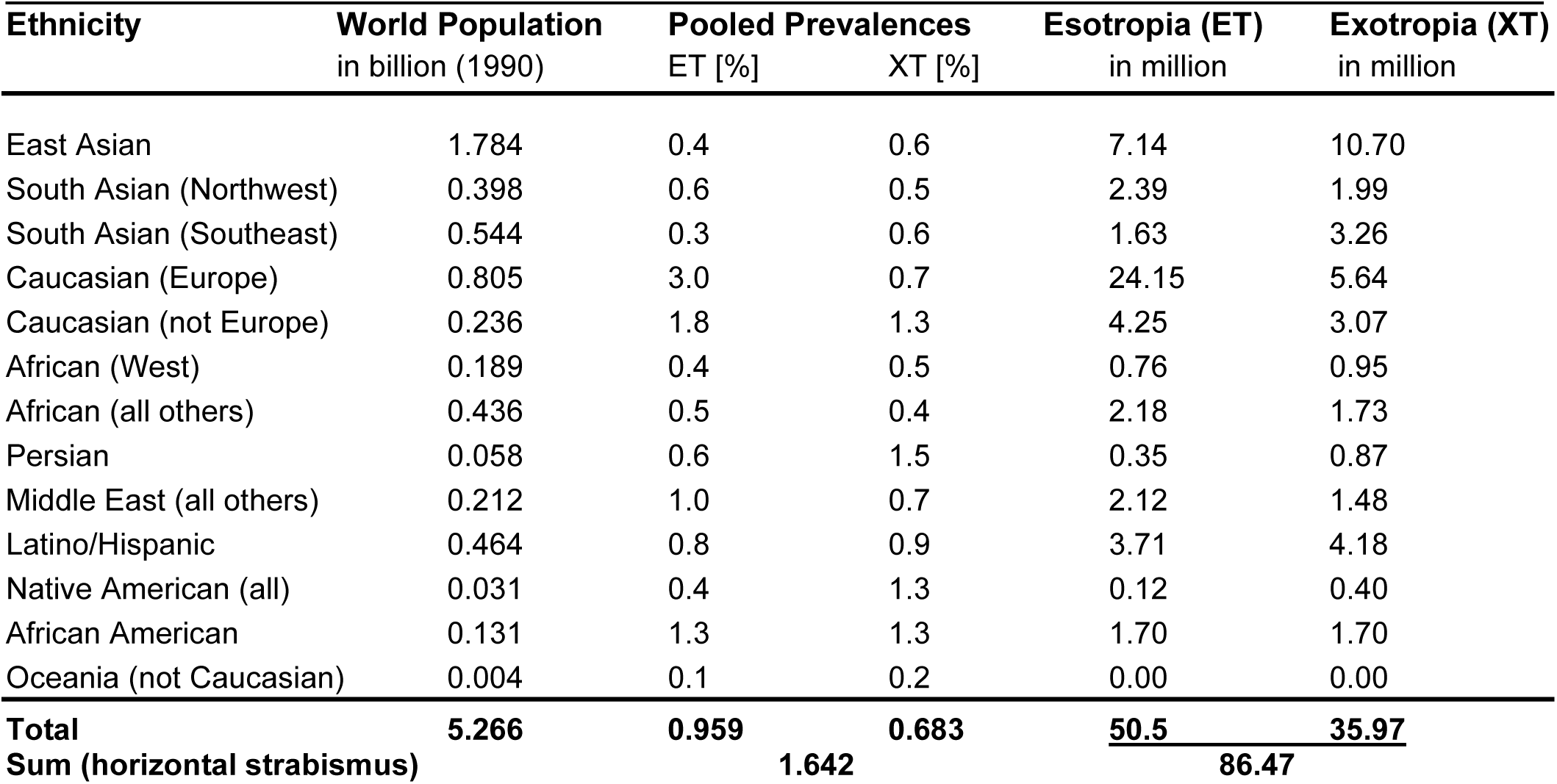
Estimates of esotropia (ET) and exotropia (XT) prevalences and the total numbers of people in the world with ET and XT based on ethnicities – previous generation(s) (1960-1990).

**TABLE 5.** Estimates of esotropia (ET) and exotropia (XT) prevalences and the total numbers of people in the world with ET and XT based on ethnicities – current generation (1990-2025).

| Ethnicity | World Population<br>in billion (2020) | Pooled Prevalences |  | Esotropia (ET)<br>in million | Exotropia (XT)<br>in million |
| --- | --- | --- | --- | --- | --- |
|  |  | ET [%] | XT [%] |  |  |
| East Asian (others) | 0.853 | 0.3 | 1.15 | 2.56 | 9.81 |
| China, Nepal | 1.412. 0.029 | 0.4, 0.1 | 2.7, 1.9 | 5.68 | 38.67 |
| South Asian (Northwest) | 0.663 | 0.6 | 0.5 | 3.98 | 3.32 |
| South Asian (Southeast) | 0.818 | 0.3 | 0.6 | 2.45 | 4.91 |
| Caucasian (Europe) | 0.675 | 1.8 | 1.0 | 12.15 | 6.75 |
| Caucasian (not Europe) | 0.442 | 1.2 | 1.1 | 5.30 | 4.86 |
| African (West) | 0.436 | 0.4 | 0.5 | 1.74 | 2.18 |
| African (all others) | 0.945 | 0.5 | 0.4 | 4.73 | 3.78 |
| Persian | 0.084 | 0.6 | 1.5 | 0.50 | 1.26 |
| Middle East (all others) | 0.356 | 1.0 | 0.7 | 3.56 | 2.49 |
| Latino/Hispanic | 0.716 | 0.8 | 0.9 | 5.73 | 6.44 |
| Native American (all) | 0.047 | 0.4 | 1.3 | 0.19 | 0.61 |
| African Americans | 0.184 | 1.3 | 1.3 | 2.39 | 2.39 |
| Oceania (not Caucasian) | 0.007 | 0.1 | 0.2 | 0.01 | 0.01 |
| **Total** | **7.667** | **0.665** | **1.141** | **50.97** | **87.48** |
| **Sum (horizontal strabismus)** |  | **1.806** |  | **138.45** |  |

*In the previous generation* (1965 to 1995), we estimate that 86.5 million people in the world had horizontal strabismus, 50.5 million with ET, and 36.0 million with XT. This results in a global ET prevalence of 0.96%, and a global XT prevalence of 0.69% for the previous generation (Table 4). The ET prevalence was 3.0% in Caucasians in Europe and 1.8% in Caucasians outside of Euope. In East Asians, the XT prevalence was 0.6%. Due to the higher prevalence of ET in Caucasians (peak of the ET epidemic), but the not yet increased XT prevalence of East Asians (still at 0.6%), the global prevalence of horizontal strabismus was slightly lower (at 1.64%) in the previous generation than it is today (1.81%).

In the *current generation* (1996 to 2025), we estimate that a total of 138.5 million people have horizontal strabismus worldwide, with 51 million having ET, and 87.5 million having XT (Table 5). This results in a global prevalence of horizontal strabismus of 1.81%, with ET at 0.67%, and XT prevalence at 1.14% in the current generation. Thus, compared to the previous generation the ET prevalence decreased, but the XT prevalence increased substantially, due to the decrease in ET in Caucasians, and the large increase of XT in East Asians (mostly in China). For these estimates, we did not take into account the bias caused by neglect of children with major disabilities (revealed by comparison with birth-cohort studies), and this prevalence estimate therefore may be an underestimate.

## 4 Discussion

In the discussion of our findings, we first review potential sources of bias when estimating prevalences, and then comment on the ethnic patterns of horizontal strabismus, many of which are reported here for the first time. We also review the history of the discovery of ethnic variations in strabismus, including why most of these patterns have eluded recognition for many decades. We discuss implications for understanding how such remarkably robust patterns evolved, apparently through emergence of divergent gene pools due to gene admixtures and human migrations. Finally, we present a new theoretical model of how intrinsic factors (genetics) determine a disposition for horizontal strabismus types, but this disposition can be modified by extrinsic risk factors.

### 4.1 Technical Considerations: Sources of Bias in Prevalence Studies

Previous investigators have recognized some of the types and sources of potential biases that need to be considered when attempts are made to estimate the prevalence of horizontal strabismus. Such potential biases include the following: methodology used for examination, classification of strabismus types, study design with possibly biased sampling, age of cohorts, effects of socioeconomic status, geography (latitude), and ethnicity.

Different *methodology* may be used in different studies for sampling, screening and examination,^23,61,174,293,356,395,A^ and different *terminology* may be used to describe and sort the results,^261,287,583,697^ different studies may apply varying definitions of manifest strabismus (strabismus angle), and some may have included microstrabismus^491^ or convergence insufficiency.^345,352,465,697^ During screening, different types of *diagnostic measures* may be used, and small deviations or intermittent *types of strabismus may be missed*,^142,149,200,255,286,303,421,604,631,697^ or may be deliberately excluded.^18^ Some children are too young to cooperate during examination.^399,604^ Depending on the screening design, some studies may have *low (parental) response rates*.^453,491,501,545,582–583,666,A,W^ Some studies excluded subjects whose strabismus had already been treated,^200,201,255,261,417,J^ and school-based studies may miss those children who are unable, due to *disabilities*, to attend normal schools or preschools and child care centers.^110,113,122,255,333,369,575,582,808,A^ Such an underrepresentation of children with disabilities is a concern, because children with disabilities are known to have substantially increased prevalences of ET and XT.^39,159,223,251,295,312,333,515,745,783,808,813,818,836^ Another potential source of bias regarding sampling is that some populations, due to differences in cultural beliefs, may hide people with ET or XT from investigators so they may be excluded from surveys, while other populations may try to find especially those subjects with ocular misalignments,^280,482^ either of which may introduce bias. Cohorts of different *ages* were examined (Fig. 6), and studies examined populations of different *ethnicities*.

Additional concerns are publication bias, because data not supporting the predominant notion of a 2-4% prevalence of horizontal strabismus may not be submitted or accepted for publication. This is a particular concern for studies in countries where the prevalence of horizontal strabismus apparently is lower (Africa, South Asia) than in other regions, or where the type of strabismus pattern differs from the Caucasian “norm”, as is the case in East Asia, West Africa, and in Persians (Fig. 3; Supplemental Table S1).^41,290,341,419,554,641,846^ Since authors usually compare their results with previously published studies, they may have doubts about the validity of their results and therefore may be more hesitant to publish seemingly discrepant results.

### 4 2. Measurement, Evaluation and Minimization of Biases

We here discuss how we quantifed and, when relevant, controlled for effects of age, latitude, ethnicity, method of sampling, and publication bias in the estimation of prevalence of horizontal strabismus.

#### Age

We identified age as an unlikely factor for explaining differences in prevalences between ethnicities. Studies of different ethnicities overall mostly examine children of similar ages (see Fig. 6A-F). While age clearly has an effect on prevalence, it does not explain the distinct ethnic patterns. Several studies reported that Caucasian patients “grow out of ET in or after puberty” – likely due to a widening of the interpupillary distance and growth-related changes of the orbital angles around the time of puberty, thereby reducing the disposition and risk for ET.^572,603,605,771,810,815,826–827,862^ Another – trivial – explanation for the decreased prevalence of horizontal strabismus in adults may be that in a significant fraction of cases the misalignment was corrected by surgery or by botulinum toxin treatment, although nationwide studies suggest that only about 2% to 5% of strabismus cases actually are treated with surgery.^165,451,660,663^

When we compared studies on older adults with studies on younger adults, XT prevalence or frequency was higher in the older adults, and this was apparent in both population-based and clinic-based studies, and in diverse ethnicities, including Caucasians,^178,218,231,251,356,469,658,D^ African Americans,^D^ East Asians,^282^ Hispanics/Latinos,^671^ and Persians.^320^ The shift towards XT in older age likely is due to age-related fat absorption in the orbit which causes a decrease in propotosis, which increases the risk for XT.^31,67,862^

#### Latitude

We quantified the effect size of latitude for the ET/XT ratio independent of ethnicity and found it to have no effect. The coefficient of determination (R^2^) was 0.003-0.004 –insignificant when compared with the effect size of the width of the orbit in different ethnicities.^862^ We conclude that latitude has no effect on horizontal strabismus. In this context it should be noted that an alternative, more plausible explanation for increased XT in Caucasians residing in regions such as South Africa and Australia has been proposed: the process of emigration will select families with a tendency to XT.^357^

#### Ethnicities

The distinct patterns of horizontal strabismus in the major ethnicities have not previously been appreciated – this is the main novel finding of our systematic review. Our comprehensive quantitative analysis and comparison of multiple ethnicities provide a wealth of new insights, detailed in sections 4.3 and 4.5, below. Our findings allow us to predict prevalences, recognize trends, and allow us to dissect the contributions of intrinsic and extrinsic risk factors that may differ between regions and at different epochs.

#### Lack of Data for Some Countries or Populations – Gaps in Knowledge Base

We identified for which countries ET and XT information appears to be lacking – see Fig. 4, our world map. In addition, we identify which populations, countries and continents are under represented in data on strabismus prevalence, and we point out which populations should be prioritized for gaining a better understanding of true prevalences and for providing a more reliable global estimate. There are relatively few studies from certain regions in South America, South Asia, Africa, and Central Asia.

#### Effect of Methodology/Sampling/Study Design

We addressed this issue and roughly quantified its effect. We compared Sharbini’s (2015)^A^ so-called gold standard studies (conducted by “experts”) with the results obtained by comparable studies with the same ethnicity, and we conclude that these potential biases are small, as they were not statistically significant. When we quantified the potential bias due to *disabled children not attending normal schools*, by comparing birth-cohort studies with relevant school-based studies, we found a surprisingly large difference – indicating that as many as 15% of cases with strabismus may be missed (section 3.11.ii). This suggests that there is a substantial sampling bias, consistent with recent conclusions.^333,808^ Some of the studies with the largest cohorts were found to report a lower strabismus prevalence than the pooled strabismus estimate, possibly because reduced time was expended for examining each subject,^496^ and therefore some forms of strabismus may have been missed (sections 3.11.iii and 4.1).

Because of the possibility of missing intermittent types of strabismus, under-reporting of some treated cases, and missing some children who cannot attend normal schools, we consider our prevalence estimates to be at the lower end, and the true prevalence may be somewhat higher, possibly requiring an adjustment of current estimates, although this must be confirmed in non-Caucasian ethnicities. Nevertheless, it is now possible to estimate ET and XT prevalences globally, and for individual countries, as long as their ethnic make-up is known – even when there are no empirical studies yet that have been performed in those particular countries.

### 4.3. Comparison of Results obtained from Population-based and Clinic-based Studies

Regarding the epidemiology of strabismus, two main types of studies must be distinguished:^390^ community- or population-based vs. clinic-based (patients presenting at, or being referred to, clinics). Many investigators are aware of the difference in the types of conclusions that can be drawn from population-based vs. clinic-based studies.^135,149,170,173,207,219,282,390,455,473,575,748,773,778,811,841,857,868,A^ Some investigators^753^ have stated that clinic-based studies appeared to yield results that are similar to those of population-based studies, especially for the collection and analysis of data regarding the ET/XT ratio. Clinic-based studies have also been justified as being “more consistent and reliable.”^174^ Others have cautioned that these two types of studies can not be directly compared,^149,179,390,473,841^ and, indeed, comparison of six clinic-based studies with five population-based studies, all on Caucasians, has shown significant differences (more than three-fold) in the ET/XT ratio between clinic-based and population-based studies (16.6 vs 4.7),^179^ yet it has remained unclear by how much the ET/XT ratio differs between clinic-based data and data obtained from the general population. Our meta-analysis allows us to quantify the effect size and to determine whether such bias is present only in some ethnicities or whether it is present similarly in all major ethnicities. We show that this bias in clinic-based studies is considerable, overall about 14%, although it varies in size between ethnicities and is smaller in populations with large fractions of ET (Fig. 7). We conclude that clinic-based studies underestimate the relative frequency of XT versus ET. This has important consequences when clinic-based data are used to estimate relative frequencies of ET and XT, e.g., in countries lacking population-based studies (Fig. 4).

The reasons for the reduced appearance (or frequency) of XT in the clinic are not known, but may be a reflection of the fact that XT often starts with an intermittent course,^370,562,586,810^ although others disagree.^672,697^ Such intermittent appearance of symptoms may result in less urgency, by the patient, to consult with a clinician. Forms of strabismus such as intermittent XT may be tolerated by the child and his or her parents more readily and therefore the subject may not seek medical attention.^69,149,173,255,382,646,773,778,841^ ET causes amblyopia more often than XT does.^170,666,735,850^ This may also be a cause for delay in XT treatment, because the ophthalmologist may be less inclined to initiate treatment,^158,288,434,672,697,749^ but there is some disagreement.^83,854^ Furthermore, because clinic visits are more cumbersome for people living in the countryside (patients must travel larger distances), clinic-based studies may miss cases in rural populations.^646^ This would lead to an underestimate of horizontal strabismus, and possibly affect XT more than ET.

### 4.4. The Role of Socioeconomic Status (SES) vs. Ethnicity

Studies have reported inconsistent results regarding the question whether SES contributes to ET or XT prevalence, as detailed in Section 3.10. Ethnicity can be strongly associated with SES,^834^ but the predominant effect of ethnicity on horizontal strabismus was not appreciated until recently. The correlation of SES with ET or XT in some studies may have been due to ethnic differences: i.e., there could have been biological rather than socio-economic reasons for the differences in strabismus prevalence. Biological differences between ethnicities can be mis-interpreted as differences in access to health care – “health disparities,” and vice versa. When fewer cases of strabismus are diagnosed or treated in a population, this can be due to health disparities, but, in the case of Hispanics/Latinos, Africans and some Asian ethnicities, it may be mostly or entirely explained by biological differences between ethnicities rather than being due to health disparities/access to care, as has been frequently assumed.^162,210,244,269,396,450,658,812,853,WW^

Another, “simple,” explanation has been offered to explain an association of lower SES with strabismus: it has been reported that parents of the upper class more swiftly recognize eye deviations in their children, and they have the time and means to procure prompt clinical attention and treatment.^771^ Regarding the reason for an association between low SES and strabismus, it was commented that “children [from a lower social class] are more difficult to treat effectively rather than that they more commonly develop strabismus”,^287,746^ which is in addition to possible health disparities^210^ and compliance issues for children with a low SES.^92,632,740,810^ Future studies on strabismus will have to take into account ethnicity as a confounding variable when assessing effects of SES.

### 4.5. Global Distribution of Horizontal Strabismus – Ethnic Patterns

The most intriguing finding of our study is the emergence of surprisingly distinct and consistent ethnic patterns in the prevalences of ET and XT. We show that ethnic differences and prevalence patterns affect many more populations and are much more robust than was previously realized. These findings are important for several reasons: First, they show that a global prevalence can only be established when ethnic differences are taken into consideration and weighed appropriately. In order to estimate strabismus prevalence within a population, the ethnic make-up of the population needs to be known. Second, to interpret trends, one must compare differences and trends *within* ethnicities, not between ethnicities – this is necessary to recognize changes in strabismus prevalence between generations. When prevalences are compiled globally or compared between different ethnicities, existing trends may be diluted or cancel out and therefore may become invisible. Third, the underlying mechanism appears to be variation in the orbital anatomy (and thus genetics),^341,815,862^ indicating an intrinsic disposition for horizontal strabismus prevalences and patterns that can be modified by extrinsic risk factors (such as maternal smoking, premature birth, Cesarean section, refractive error). Fourth, ethnic differences in strabismus are important clinically and from a public health perspective because of different frequencies of co-morbidities (e.g., amblyopia, mental health). Ethnic differences need to be taken into account to identify true health disparities, and to adequately plan for care of strabismus in the future.^659,857^ Such care needs to be tailored to the needs of local populations, but also to prepare adequately for sequelae such as increased mental health issues, especially increased schizophrenia, that may follow increased XT rates decades later.^27,507,780^ Finally, for the first time, total numbers of esotropes and exotropes regionally and worldwide can be estimated, and prevalence changes tracked longitudinally – opening new avenues for assessing roles of major known risk factors for horizontal strabismus which may differ between regions and ethnicities (see sections 4.8 and 4.9).

Some ethnicities (Africans, South Asians, indigenous people in Oceania) have much lower overall strabismus prevalences than the other major ethnicities (Fig. 3). Two possible reasons for this difference, not mutually exclusive, must be considered. First, some populations may have a low kurtosis in the anatomical parameters that impact on strabismus disposition: Interpupillary distance, proptosis and orbital angle. If some populations have fewer extremes in these orbital parameters, then there will be fewer individuals at risk, resulting in lower prevalences. Second, it is possible that babies or children with disabilities (e.g., low birth weight, premature birth, hypoxia during birth – which makes them higher risk for strabismus) do not survive in societies with less advanced health care systems, and this would reduce the ultimate strabismus prevalence in the population.^80,317,333,815^ If the second reason is true, then one would expect that, as health care facilities improve in developing countries, the prevalence of horizontal strabismus increases. Based on the currently available data, there do not seem to be such trends (yet). Of course, changes in multiple risk factors for strabismus play a role, making it difficult to adequately explore this possibility.

### 4.6. Ethnic Differences in ET and XT Prevalences: Evolutionary Implications

Our quantitative summary (Fig. 3) and the world map (Fig. 4) show notable differences in prevalences between ethnicities. Populations residing in regions with frequent migrations and/or frequent gene admixture in historic and prehistoric times (Middle East, Mediterranean, North and West Africa, Northwest India, Latin America)^593^ tend to have intermediate ratios of ET/XT prevalence, while those populations that have been more isolated (remote), or in extreme locations with less opportunity of gene admixture, have more extreme ET/XT patterns (Western Europeans, Native Americans, East Asians, indigenous people of Australia, Eskimos/Inuit).

Northwestern Europe was considered a geographical “cul de sac” in terms of ancient human migrations.^608,765^ The distribution of ethnic strabismus prevalences and ratios provides strong clues of how such patterns likely have evolved. Essentially, to permanently alter the ET/XT patterns within a population, admixture between populations has to happen. Several studies have shown that admixture between ethnicities leads to intermediate ratios and patterns of ET/XT,^361–362,517,FF^ with other examples showing that ET-XT-related parameters such as interpupillary distance and interorbital width also become intermediate in ethnically admixed offspring.^245,334^ This leads to intriguing questions of how the ethnic ET, XT diversity originated in anatomically modern humans – possibly due to gene admixture when anatomically modern humans encountered and interbred with archaic hominins^111,276,524,592,593,798,803^ who had a wider orbital width (Neanderthals and Denisovans).^524,614,XX^ A wider orbital width associates with increased inclination towards XT.^406,675,862^ Current patterns of ET/XT ratios in different countries and ethnicities are not only compatible with conclusions from population genetics, but they are best understood in the context of ancient human gene pools, gene admixture between populations, and ancient and more recent human migrations.

### 4.7. History of the Discovery of Ethnic Patterns

Given the distinct differences in strabismus patterns between ethnicities, it may appear surprising that most of these patterns have remained undetected for so long. In this section, we will examine the reasons for this delay. There seems to be a combination of factors responsible, including: Small number of studies for many ethnicities, neglect of key existing studies by researchers, and variability of results. There has been a considerable focus on Caucasians and neglect of other ethnicities, especially prior to the 1950s. Some key early studies, e.g., on African Americans, Africans, and East Asians were overlooked, apparently because they were not published in mainstream journals,^285^ or were published in non-English languages.^341,554,FF^ Additionally, many investigators were unaware of the importance of reporting data separately for different ethnicities.^841^ The African patterns remained hidden, despite first reports, more than 80 years ago, of prevalences in Africans that differed from those in Caucasians.^341^ Asian patterns remained unknown to most investigators despite multiple studies in the 1950s and 1960s^473,554,555,G^ – reviewed in 1972^419^ – apparently because most of them were published locally in non-English journals. Mosaic patterns such as those in Africa, the Arctic, the Middle East, or South Asia were difficult to discern early on, when data from only a handful of studies were available.

#### i. Africans and African Americans

It was noted at the beginning of the last century that Africans and African Americans “rarely, if at all” have strabismus^114,120,418,431,523,616^ – reviewed by multiple authors.^19,341,696,815^ However, these initial reports were merely anecdotal impressions suggesting that some Africans have a lower prevalence of strabismus than Caucasians, and other investigators emphasized that strabismus does exist in Africans.^14,183,341,418,815^ The first systematic, population-based studies were conducted by Holm^341^ in Africa, and by Gover and Yaukey^285^ in the USA, and both revealed a low prevalence of horizontal strabismus among Africans and African Americans. However, the study in the USA^285^ is often overlooked: For example, Hansen (1971)^313^ says, incorrectly, that “no information is available on the comparative occurrence of strabismus in blacks and whites in the United States.” The lower prevalence of horizontal strabismus in West Africans^341^ was confirmed by additional studies (Supplemental Table S1).^15,86,335^ Some practitioners took issue with the statement “squints are extremely rare in coloured races” (Squint Editorial in the British Medical Journal)^746^ – Taylor^766^ countered that “squints are very common in East Africa.” Indeed, several population-based studies from East Africa confirmed relatively high prevalences (e.g., Kenya: 2.86%,^R^ Ethiopia: 3.1% and 4.4%,^242,838^ Sudan: 2.73%).^760^ Yet, overall, most studies agree on a lower strabismus prevalence, and especially a low prevalence of ET, in Africans and African Americans when compared with Caucasians.^80,162,174,252,313,341,396,815,843,S^

Besides the issue of a low overall prevalence of horizontal strabismus among Africans and African Americans, there is a second parameter of interest: the ET/XT ratio. The ET/XT ratio was initially reported to be very low in Africans,^341^ and this was supported by subsequent studies in Africa,^15,33,76,225,335,423,843,R,S^ and two studies in the USA.^545,D^ Some authors state that Gover and Yaukey^285^ were the first to report that XT is more frequent than ET in African Americans.^220,612,696^ This is incorrect and due to a misread of Schlossman and Priestley’s review^696^ by Abrahamsson et al.^19^ Roberts and Rowland^D^ and the Multi-ethnic Pediatric Eye Disease Study Group^545^ were the first to report a greater XT than ET prevalence in African Americans. However, other studies in Africa, and most of the ones in the USA, reported approximately equal ratios of ET and XT (USA and Caribbean,^170,187,255,632^ Africa,^252^ or even higher ET than XT prevalences (USA,^147,259,631^ Africa,^49,71,86,103,275,551,760^). How can such seemingly discrepant results be explained? Two possible, not mutually exclusive, explanations are (1) that the ancestors of current day African Americans have admixed with Caucasians to dilute the original pattern (indeed African Americans have, on average, only 70-80% original African genes,^84,115–116^) and (2) the ancestries within in Africa are not uniform, but constitute a mosaic of patterns.

Our study is the first to show that there are indeed two major patterns of horizontal strabismus in Africa. Many of the early studies happened to examine Niger-Congo-Bantu populations in West Africa, where in general XT prevalence exceeds ET prevalence (Fig. 4; Supplemental Table S1). Friedman et al.^255^ suggested that ET/XT differences between cohorts of African Americans may reflect the fact that they originated from different sites and populations in Africa and thus reflect genetic differences – a notion which our review and other studies support.^84,115–116,121,167,774^ Indeed, a smaller fraction of African Americans has origins among the Niger-Congo-Bantu in West Africa, while the majority of African Americans appears to originate from other populations in Africa.^115^ With additional reports from the last 12 years, significant ET/XT differences between Niger-Congo-Bantu and non-Niger-Congo-Bantu populations have now emerged – this mosaic pattern on the African continent would have been difficult if not impossible to recognize just one or two decades ago due to the scarcity of studies (Fig. 4; Supplemental Table S1).

#### ii. East Asians

East Asians are known to have a higher XT than ET prevalence. Such differences, when compared with Caucasians, were first documented for the Japanese in 1954,^554^ and for Chinese living in Fiji.^G^ These initial reports were supported by subsequent studies in the 1950s, 1960s, and 1970s in Japan;^314,473,493,517,555,584,844,FF^ – for a review of the early literature, see Konyama, 1972.^419^ This divergent pattern – East Asians having patterns distinct from Caucasians – was further confirmed in the 1970s, 1980s and 1990s in Thais,^419,470^ in Koreans,^639,856^ in Malaysians,^769^ and eventually in Chinese populations (Taiwan,^449,705,821^ Hong Kong,^206^ Kazakhstan,^375^). However, none of these studies attracted much attention, and therefore the difference between East Asians and Caucasians regarding the ET/XT ratio remained hidden. The ethnic difference was effectively communicated to a wider audience only relatively recently, by Yu et al., 2002,^857^ based on clinic data.^433^

Some articles and reviews, including recent ones, group all “Asians” in the XT >> ET category,^220,282,290,317,318,442,615,855^ however, our work shows that this is incorrect, because a sizeable fraction of South Asian populations does not share the East Asian pattern. To our knowledge, the difference in strabismus patterns between populations in the Northwest and the Southeast of the Indian subcontinent has not previously been noticed (see below, section 4.5.vi).

#### iii. Pacific Islanders

The work of early investigators^216,482,484,486,802,G,X^ showed that the indigenous people in the South Pacific (including Australia) generally have very low prevalences of strabismus, and an especially low, almost non-existing prevalence of ET. This was confirmed by more recent studies.^345,590,767^ Mann^482^ realized that the global variation of horizontal strabismus was primarily due to differences in the prevalence of ET, not in the prevalence of XT.

#### iv. Native Americans

Initially, the strabismus pattern of Native Americans was difficult to recognize. Apparently, none of the authors conducting studies in North America^25,152,271,437,489,830^ were aware of reports from South America,^99,790^ and one of the first studies in North America combined data from Eskimos/Inuit and Native Americans,^841^ thus “diluting” the diametrical patterns of these two ethnicities. The recent study from Brazil by Germano et al.^272^ was not aware of the earlier Latin American literature on Native Americans.^99,483,517,790,FF^ When one considers all available studies, including the more recent ones from the last 20 years,^152,271,272,437^ together with some of the classic older studies,^25,99,489,517,830,790,FF^ it emerges that Native Americans, overall, have less horizontal strabismus than Caucasians,^853^ and that their pattern (with significantly more XT than ET) is remarkably similar to that of East Asians (Fig. 3). The finding of a lower prevalence of strabismus in Native Americans is supported by qualitative observations such as “no Indians…ever get concomitant strabismus unless white blood is present”^129^ and “Indians have less muscular problems [than Caucasians].”^309^

#### v. Eskimos/Inuit

Ida Mann emphasized the high frequency of ET in Eskimos/Inuit based on clinical cohorts,^486^ but a subsequent population-based study in Canada reported only on the combined data from Eskimos/Inuit and Native Americans.^841^ This obscured the distinct pattern in Eskimos/Inuit. A high ET prevalence among Eskimos, as reported by Mann in 1972,^486^ was corroborated.^380,835^ Apparently unaware of Mann’s and Wyatt and Boyd’s work,^486,841^ Woodruff and Samek^835^ commented on the fact that the frequent ET in Eskimos/Inuit was not due to hypermetropia; they found “no reasonable explanation of cause” for the frequent ET in Eskimos/Inuit. Apparently, the authors missed the works of Skeller (1954)^736^ and Waardenburg (1954)^815^ who had pointed out a narrow interorbital width and a particularly short interpupillary distance that is unique to the Eskimo/Inuit skull,^125,353^ thus providing a plausible explanation for the frequent ET in Eskimos/Inuit.^815,862^

#### vi. South Asians

The first population-based studies from South Asia generated a confusing picture, since the studies from the Northwest of India indicated an ET/XT ratio that was approximately equal or showed much more ET than XT,^26,296,546^ while population-based studies from Southeast India indicated a much larger prevalence of XT than ET.^427,652^ Nearly all subsequent studies from the Northwest of the Indian subcontinent confirmed a similar prevalence of ET and XT or more ET than XT,^94,530,716^ with only one study showing significantly more XT than ET.^72^ In the Southeast of India, nearly all studies report a much lower ET/XT ratio.^29,68,176,300,393,400,552,688,733^ Despite the relatively sparse number of population-based studies in South Asia,^861^ thus, the evidence for differences between these populations is accumulating. Accordingly, there is no single “South Asian” pattern, because the pattern in Northwest India and in Pakistan is more similar to that of most populations in the Middle East, which differs significantly from that of East Asians, while South Indians’ and Persians’ patterns may reflect their common Dravidian ancestry.^91,232,383,475,655^

#### vii. Middle East

The first studies from the Middle East were on Jews and Arabs, with later studies on Turks and Persians. It turned out that prevalences from countries with Jews, Arabs, Turks and Kurds were very similar, with ET typically exceeding XT prevalences, similar to Caucasians and North Africans, while Persians consistently show a higher prevalence of XT over ET, more similar to Southeast Indians and East Asians. Insights into differences among Middle Eastern populations were made possible only in the last decade, since countries such as Iran do not seem to have any population-based studies in the 20th century that distinguished between ET and XT (Supplemental Table S1).^318,403,679^

In summary, recognition of differences between ethnicities and patterns of strabismus prevalence requires adequate numbers of population-based studies, and for many countries and populations, such studies have become available only in recent years. Some insights into ethnic differences remained hidden due to neglect of reports published in languages other than English. As with a giant jig saw puzzle, a sufficient number of prominently placed pieces of the puzzle is needed to recognize patterns, and initially, these pieces were too few and too far between. However, clear patterns are now emerging that not only provide a convincing account of the current status, but also intriguing indications on how these distinct patterns of strabismus prevalence must have evolved.

The basis and mechanism of the ethnic strabismus patterns likely relate to the known differences between ethnicities in orbital anatomy: interorbital width, interpupillary distances, and proptosis.^86,104,341,406,431,487,517,633,635,756–757,815,862,FF^ Such differences between ethnic groups shift the balance between enhanced disposition (risks) for ET vs. XT.^406,431,815,857^ This notion is supported by the well-known fact that clinical syndromes with narrow interpupillary distances (e.g. Down syndrome) have much more ET,^336,368,808^ while syndromes with extremely wide interpupillary distances (e.g., Crouzon or Apert syndrome) have much more XT associated.^675^ In addition to orbital parameters, the potential role of refractive errors needs to be considered.

Numerous studies have shown associations between refractive errors (hyperopia, myopia, astigmatism and anisometropia) and different types of horizontal strabismus.^170,444,764,866^ Esotropia is associated with hyperopia, and exotropia is associated with myopia. However, the causal relationship between refractive error and strabismus is unclear, and it has been suggested that exotropia may cause or worsen myopia rather than the refractive error causing the strabismus.^211,866^ Since the prevalence and types of refractive error differ between ethnicities, it is tempting to speculate whether the current myopia epidemic may, in part, explain the recent XT surge in China. However, the fact that the myopia epidemic involves all of East Asia, including Korea, Japan, and Indonesia,^801^ but the XT surge is only seen in China (and Nepal), indicates that other factors must play a role.

It goes beyond the scope of our review to discuss the extensive literature on refractive errors and strabismus, or the above-mentioned craniofacial dysostosis syndromes, or the evolutionary history of diverse orbital patterns in different archaic and modern human populations, or to discuss the probable molecular and genetic bases of orbital diversity – which all are fascinating topics for further study.

### 4.8. The Global Prevalence of Horizontal Strabismus: Intrinsic Factors, Extrinsic Factors, and Generational Changes

Estimation of the global prevalence of horizontal strabismus is challenging because the global prevalence is composed of the sum of all the ethnically distinct prevalences among its constituent populations. Furthermore, the ethnically distinct prevalences are determined by two types of factors: (1) intrinsic (genetic) factors that govern the shape of the orbit, and (2) extrinsic factors that impact on the anatomically defined intrinsic “baseline.” The occurrence of two epidemics, in populations (Europeans and Chinese) whose ethnic make-up has changed minimally, if at all, suggests that the intrinsic, genetically determined disposition for horizontal strabismus can be modified by extrinsic (environmental) factors. The relevance of extrinsic factors becomes apparent when one analyzes generational changes of strabismus prevalence within the same population. The most likely relevant extrinsic factors (risk factors) are cultural and health factors such as maternal smoking and Cesarean section,^147,237,306,326,609,777,842,I^ but there may be others: low birth weight (in Japan^762^) premature birth, some types of refractive errors, and reduced risks of childhood diseases due to vaccinations.^126,243^ Both types of factors (intrinsic/genetic and extrinsic/environmental) need to be considered for the global prevalence. Some eye conditions have both intrinsic (genetic) and extrinsic (environmental) contributions, as exemplified by the phenomenon of myopia.^801^

Different estimates of global strabismus prevalence will be obtained when they are based on fractions of the world population that are not representative of the entire population, and different estimates will also be obtained when data from different generations are used for calculations, because exposure to extrinsic risk factors may differ between generations. In at least two major populations (Europeans and Chinese), strabismus prevalence has changed significantly between generations (Fig. 10A,B). For meaningful estimates and comparisons, it should be specified for which generation the prevalence is estimated.

Until now, many regional patterns of horizontal strabismus were not apparent, because differences between most ethnicities were not known, and because the changes in prevalence over time within the same population were not appreciated. Accordingly, postulated or assumed prevalences often were mixtures from different ethnicities, with contributing data from different epochs, sometimes going back several generations. Our meta-analysis with a global and historical focus makes possible meaningful analyses of both, global prevalence, and local prevalence, for different generations, and can reveal secular trends – for those ethnicities where sufficient numbers of studies are available. These new insights will allow investigators to determine with much greater precision the effects of known risk factors and to predict effects of such risk factors in current and future generations.

Our estimates of the global prevalence of ET and XT in the current generation, at 0.67% and 1.14%, respectively, are lower than those from some previous studies. Adinanto et al.^B^ estimated horizontal strabismus at 2.9% in 2010-2015, while Hashemi et al.^318^ estimated a 2.00% prevalence in 1986-2018, with 0.77% esotropia, and 1.23% exotropia. One major reason for the differences between estimates is that none of the previous studies accounted for the substantial ethnic differences, with very low prevalences in Africa and South Asia, and did not account for such effects on a global scale. Prevalences of past generations are not necessarily appropriate for estimating the *current* global prevalence, especially when current and past generations may have been exposed at different rates to extrinsic risk factors (such as maternal smoking, low birth weight, premature birth, Cesarean section).^101^ The prevalence of ET in Europeans, with many studies from the peak of the ET epidemic, was taken to be representative for other ethnicities, and this seems to be largely responsible for the broadly incorrect estimates of global strabismus prevalences as summarized in Table 1 and Figure 1.

### 4.9. Implications for Global Health beyond Ophthalmology

Strabismus has detrimental consequences not only for visual function such as possible amblyopia, loss of binocular vision and defects of stereopsis,^434,810^ but also for psychosocial development^117,662^ and development of mental illness, schizophrenia in particular.^27–28,379,445,507,511,535,560,698,780^ The substantially increased risk of schizophrenia after childhood XT, and especially constant XT, is particularly concerning.^27,780,YY^ Indeed, in countries such as China, where XT has been increasing in the last three decades, it has been reported that schizophrenia prevalence has more than doubled between 1990 and 2010.^134^ Since XT prevalence has continued to increase in China, this trend and the delayed impact on schizophrenia, can be expected to continue for the next two decades, assuming that XT is indeed a significant risk factor for schizophrenia.^27^ If true, then reducing modifiable risk factors for XT, such as Cesarean section^609,776^ or certain types of refractive errors (severe myopia, astigmatism, anisometropia),^170,444,764,866^ may decrease XT prevalence, and with that, eventually, schizophrenia prevalence. It is not known whether prompt treatment of XT may prevent schizophrenia from developing,^27,407^ but this should be explored. Nevertheless, these considerations illustrate how important it is to know the prevalence of horizontal strabismus, to monitor trends, and to advocate preventative steps as well as early interventions in individuals with ET or XT.

## 5 Conclusions

Our work reveals that the preeminent factor that explains differences in prevalence of the different types of horizontal strabismus is ethnicity, rather than socioeconomic status, methodology, or geography. The underlying reasons for ethnic differences are likely due to differences in orbital anatomy,^862^ but this, and the effects of extrinsic (environmental) risk factors require additional investigation. We also show longitudinal, generational changes in strabismus prevalence. Monitoring strabismus prevalence between generations shows that extrinsic factors (risk factors) play a role, but their identification awaits future confirmation. With more accurate estimates of the global and local prevalence of esotropia and exotropia, and their trends, more targeted measures can be taken to mitigate the downstream consequences and adverse effects of horizontal strabismus on individuals and society. By facilitating access to a large body of previously neglected literature, our work provides a more appropriate context and comparison for future studies. This should stimulate further research to better understand horizontal strabismus and its evolutionary history.

## 6 Method of Literature Search

PubMed and Google Scholar were searched for relevant studies. Our review protocol was registered in PROSPERO (CRD420251080354).^E^ The following search terms were used: “strabismus”, “esotropia”, “exotropia”, “prevalence.” Inclusion criteria were quantitative information on the prevalence of esotropia and exotropia or the ratio of esotropia/exotropia. There were no language restrictions as long as references or English titles or relevant key words appeared in searches. We accepted all studies that report original and quantitative data on frequencies of esotropia (ET) and exotropia (XT) in human subjects, either obtained from population-based surveys, or from medical records in clinical settings. Studies were included only when they provided *quantitative* information about both ET and XT separately (not just combined ET and XT prevalence or prevalence of horizontal plus vertical strabismus). We accepted both, population-based studies, as well as clinic-based studies (surveys of medical records from patients seeking attention at a clinic). Case reports were not included. All relevant studies were sorted into either population-based studies or clinic-based studies. Our search was not restricted by year of publication or language. The last search was conducted on 1 July, 2025. For a detailed description of inclusion and exclusion criteria, quality and risk for bias assessment, data collection, and statistical analyses, please refer to the Methods section.

## Key References

As key references, we selected Hermann Cohn, 1867 (Breslau, Germany),^163^ because his was the first study reporting the prevalence of horizontal strabismus. We also include Stig Holm, 1939, (Lambarene, Gabon and Lund, Sweden)^341^ because his work was the first to report on ethnic differences in the type and prevalence of horizontal strabismus. We acknowledge the pioneering work of Ida Mann, 1966, (Perth, Australia)^482^ who provided the first global analysis and insight into ophthalmologic disease, including strabismus. The work of Emily Chew and colleagues (1994) (Bethesda, USA)^147^ provided not only comprehensive data on strabismus prevalence, but also on extrinsic (risk) factors that modify strabismus prevalence. The final work that we acknowledge is by Hassan Hashemi and colleagues (2019) (Tehran, Iran),^318^ because it was the first systematic review that estimated the global prevalence of strabismus, including esotropia and exotropia. All five works laid the foundation for significant progress in the epidemiology of strabismus and pioneered a better understanding of this disorder.

## Disclosure

The authors have no conflicts of interest to declare, except that our study was supported by the National Institutes of Health, grant numbers EY031729 (C.S.v.B. and W.Y.) and GM103554 (C.S.v.B.).

## CRediT authorship contribution statement

**Christopher von Bartheld:** Conceptualization, Data curation, Supervision, Project administration, Writing – original draft, review & editing. **Molly Hagen:** Methodology, Software, Formal analysis, Writing – review & editing, Methodology. **Jiayi Jiang:** Software, Methodology, Formal analysis. **Wei Yang:** Supervision, Conceptualization, Methodology, Writing – review & editing. **Andrea Agarwal:** Data curation, Investigation, Conceptualization, Writing – review & editing.

## Declaration of Generative AI and AI-assisted technologies in the writing process

No such technologies were used in the writing process.

## Declaration of Competing Interest

The authors state that they have no financial interests or personal relationships that could have influenced the work presented in this paper, other than receiving grant funding to conduct the work. The sponsors had no role in relation to the study design, collection, analysis and interpretation of data, writing of the review and decision to submit for publication.

The authors report no proprietary or commercial interest in any product mentioned in this article.

## Ethics

Our review is based on previous research, and therefore does not require approval from local Ethics Committees.

## Data Availability

All data produced in the present work are contained in the manuscript and Supplemental Tables or are available upon reasonable request to the authors

## Acknowledgments

We thank Norman Huckle and Jenny Costa (University of Nevada, Reno), Chengyuan Feng (Johns Hopkins University), and Carina Vetye-Maler (Munich/Buenos Aires) for retrieval of literature. For translations, we thank Larisa Baryshnikova-Wiggins and Rafal Butowt (University of Nevada, Reno), Chengyuan Feng (Johns Hopkins University), Koji Matsuda (Matsuda Eye Clinic, Osaka, Japan), and Sineenart Sengyee, Elham Taheri, and Haifeng Zheng (University of Nevada, Reno). We are grateful to Aderonke Baiyeroju (University of Ibidan, Nigeria) and Napaporn Tananuvat (Chiang Mai University Thailand) for making literature available. We also thank Alan Johnson (Sierra Eye Associates, Reno, Nevada) and Koji Matsuda (Matsuda Eye Clinc, Osaka, Japan) for helpful comments on earlier versions of our manuscript. This work is dedicated to the memory of three pioneers of strabismus prevalence: Hermann Cohn (Breslau, Germany), Stig Holm (Lambarene, Gabon and Lund, Sweden), and Ida Mann (Perth, Australia).

## SUPPLEMENTAL MATERIALS

**Supplemental Table 1.** Esotropia/exotropia (ET/XT) ratio in different regions. Population-based, 315 studies with 340 cohorts. * 0.5 was added to each value for the ET/XT ratio when the value was 0, to avoid a zero in the denominator or enumerator, according to Sweeting et al., 2004.^758^ Cohort type: C, community; S, School or Pre-school; B, Birth cohort. Yrs, years; Hor. Strab, horizontal strabismus; ET, esotropia, XT, exotropia. Duplicate publication of the same cohort is indicated in *italics*. Studies reporting exclusively on ages 2 years or younger are gray-shaded.

| First Author | Year | Ethnicity/Region | Cohort size | Cohort type | Age yrs | Hor. Strab % | ET % | XT % | ET/XT Ratio |
| --- | --- | --- | --- | --- | --- | --- | --- | --- | --- |
| **EUROPE** |  |  |  |  |  |  |  |  |  |
| Adler | 1876 | Vienna, Austria | 100 | S | 8-19 | 4.00 | 3.00 | 1.00 | 3.00 |
| Schenk | 1969 | Vienna, Austria | 67 | C | 4-7 | 10.45 | 8.96 | 1.49 | 6.00 |
| Aichmair | 1992 | Vienna, Austria | 843 | S | ~8 | 3.91 | 2.97 | 0.95 | 3.13 |
| Vereecken | 1966 | Belgium | 1,215 | C | 6-10 | 3.70 | 3.29 | 0.41 | 8.00 |
| Popovic-Beganovic | 2018 | Bosnia | 997 | S | 7-16 | 2.91 | 1.50 | 1.40 | 1.07 |
| Medkova | 1959 | Opava, Czechoslovakia | 148 | C? | 4-6 | 4.73 | 4.05 | 0.68 | 6.00 |
| Karlica | 2008 | Croatia | 20,045 | B | ~2-5 | 3.97 | 2.14 | 1.84 | 1.16 |
| *Frandsen* | 1958 | *Denmark* | *16,146* | *S* | *0.5-18* | *~5* | *~4* | *~1* | *~4* |
| *Frandsen* | 1960 | *Denmark* | *13,107* | *S* | *0-20* | *4.39* | *3.47* | *0.92* | *3.77* |
| Frandsen | 1960 | Denmark | 2,570 | S | 0-6 | 6.03 | 5.29 | 0.74 | 7.16 |
| Frandsen | 1960 | Denmark | 10,537 | S | 7-20 | 4.41 | 3.36 | 1.05 | 3.19 |
| Sandfeld | 2018 | Roskilde, Denmark | 445 | S | 4-7 | 1.57 | 1.35 | 0.22 | 6.00 |
| Hultman | 2019 | Denmark | 3,785 | C | 20-80+ | 1.08 | 0.77 | 0.32 | 2.42 |
| Rantanen | 1971 | Helsinki, Finland | 2,100 | S | 7 | 4.76 | 2.62 | 2.14 | 1.22 |
| Laatikainen | 1980 | Helsinki, Finland | 411 | S | 7-15 | 4.62 | 2.92 | 1.70 | 1.71 |
| Kaakinen | 1981 | Helsinki, Finland | 182 | C? | 1-4 | 1.10 | 1.10 | 0 | 5* |
| Kaakinen | 1986 | Helsinki, Finland | 169 | C? | 1-5 | 3.55 | 2.96 | 0.59 | 5.00 |
| Tuppurainen | 1993 | Kuopio, Finland | 56 | C | 5 | 1.79 | 0 | 1.79 | 0.33* |
| Speeg-Schatz | 2004 | Alsace, France | 2,318 | S | 1-6 | 3.11 | 2.89 | 0.22 | 13.4 |
| Sigronde | 2020 | Burgundy, France | 1,236 | S | 3-5 | 3.24 | 1.54 | 1.70 | 0.90 |
| *Cohn* | 1867 | *Breslau, Germany* | *10,060* | *S* | *6-16* | *2.25* | *2.21* | *0.04* | *55.5* |
| *Cohn* | 1904 | *Breslau, Germany* | *10,000* | *S* | *children* | *-* | *2.22* | *-* | *-* |
| Schleich | 1905 | Tübingen, Germany | 2,098 | S | ~6-18 | 1.48 | 1.38 | 0.10 | 15.5 |
| Speidel | 1905 | Tübingen, Germany | 566 | S | ~16-24 | 2.83 | 1.06 | 1.77 | 0.60 |
| Schildwächter | 1972 | Hessia, Germany | 2,660 | S | ~4-9 | 4.10 | 4.10 | 0 | 219* |
| De Decker | 1973 | Kiel, Germany | 1,525 | S | 6-7 | 5.25 | 4.39 | 0.85 | 5.15 |
| Schütte | 1976 | Aachen, Germany | 4,229 | S | 2-7 | 4.26 | 3.43 | 0.83 | 4.14 |
| Toppel | 1978 | Munich, Germany | 1,212 | S | 0-12 | 3.55 | 3.47 | 0.08 | 42.0 |
| Haase | 1979 | Hamburg, Germany | 830 | S | 6-7 | 5.18 | 5.18 | 1.08 | 4.78 |
| Friedrich | 1987 | Kiel, Germany | 1,024 | B | 0 | 0.1 | 0.1 | 0 | 3* |
| Friedrich | 1987 | Kiel, Germany | 180 | B | 0.1-0.3 | 55.56 | 5.56 | 50? | 0.11 |
| Rüssmann | 1990 | Cologne, Germany | 254 | S | 5-6 | 1.97 | 1.18 | 0.79 | 1.5 |
| Kasman-Keller | 1998 | Saar, Germany | 939 | S | 3-6 | 2.34 | 1.70 | 0.64 | 2.67 |
| Fiess | 2017 | Wiesbaden, Germany | 264 | C | 4-10 | 1.52 | 1.14 | 0.38 | 3.00 |
| Fiess | 2020 | Mainz, Germany | 14,700 | C | 35-74 | 2.40 | 1.60 | 0.80 | 2.00 |
| Von Csapody-Mocsy | 1934 | Budapest, Hungary | 8,000 | S | children | - | 3.94 | - | - |
| Donnelly | 2005 | Ulster, N. Ireland | 1,582 | C | 8-9 | 3.98 | 3.35 | 0.63 | 5.30 |
| Montvilaite | 2015 | Lithuania | 40 | C | 5-7 | 2.50 | 2.50 | 0 | 3* |
| Paduca | 2020 | Moldova | 861,682 | C | 0-17 | 1.29 | 1.00 | 0.29 | 3.41 |
| Polling | 2015 | Rotterdam, Netherlands | 6,690 | S? | Mean=6 | 1.48 | 1.11 | 0.37 | 2.96 |
| Gunnufsen | 1938 | Oslo, Norway | 50,000 | S | 9-10 | - | 1.8 | - | - |
| Holst | 1962 | Oslo, Norway | 21,484 | S | 9-10 | 2.25 | 2.16 | 0.1 | 21.6 |
| Holst | 1962 | Oslo, Norway | 31,330 | S | 9-10 | 2.05 | 1.95 | 0.1 | 19.5 |
| Aslaksen | 2023 | Sorlandet, Norway | 63 | B | 5-13 | 4.76 | 0.00 | 4.76 | 0.14* |
| Dalz | 2009 | Poznan, Poland | 3,025 | S | 7-15 | 1.79 | 1.39 | 0.40 | 3.50 |
| Quental | 2013 | Portugal (South) | 885 | S | 3-15 | 1.47 | 0.79 | 0.68 | 1.17 |
| Lanca | 2014 | Lisbon, Portugal | 672 | S | 6-11 | 3.87 | 2.08 | 1.79 | 1.16 |
| Hendrickson | 2001 | Sibiu, Romania | 676 | S | 2-18 | 9.03 | 6.07 | 2.96 | 2.05 |
| Hendrickson | 2001 | Sibiu, Romania | 184 | S | 19-77 | 5.43 | 3.26 | 2.17 | 1.50 |
| Mazepa | 1967 | Saratov, Russia | 1,507 | S? | 3-7 | 1.99 | 1.99 | 0 | 61* |
| Maimulov | 1971 | Leningrad, Russia | 2,034 | S? | 5-7 | 3.05 | 1.67 | 1.38 | 1.21 |
| Delgado-Molina | 1991 | Madrid, Spain | 1,852 | S | ~8 | 1.08 | 0.86 | 0.22 | 4.00 |
| Martinez | 1997 | Valladolid, Spain | 1,179 | B | 3-6 | 3.99 | 2.88 | 1.10 | 2.62 |
| Nordlöv | 1944 | Gotaland, Sweden | 2,271 | S | 7 | 3.57 | 3.17 | 0.40 | 8.00 |
| Nordlöv | 1964 | Gotaland, Sweden | 6,004 | B | 0-7 | 3.98 | 3.55 | 0.43 | 8.19 |
| *Fabian* | *1966* | *Vasteras, Sweden* | *1,200* | *B* | *2* | *2.08* | *1.75* | *0.33* | *5.25* |
| Köhler | 1973 | Lund, Sweden | 2,397 | C | 4 | 3.80 | 2.89 | 0.92 | 3.17 |
| *Fabian* | *1974* | *Vasteras, Sweden* | *1,200* | *B* | *2* | *2.08* | *1.75* | *0.33* | *5.25* |
| *Lennerstrand* | *1989* | *Stockholm, Sweden* | *1,047* | *C* | *5-10* | *1.43* | *0.86* | *0.57* | *1.50* |
| *Gallo* | *1991* | *Stockholm, Sweden* | *1,047* | *C* | *5-10* | *1.34* | *0.76* | *0.76* | *1.33* |
| Rassmussen | 2000 | Uppsala, Sweden | 3,493 | B | 3-6.5 | 3.5 | 2.38 | 1.12 | 2.13 |
| Kvarnström | 2001 | Lund/Linköping, Sweden | 3,126 | B | <10 | 2.08 | 1.50 | 0.58 | 2.59 |
| Ohlsson | 2001 | Göteborg, Sweden | 1,046 | S | 12-13 | 1.53 | 0.86 | 0.67 | 1.29 |
| *Aring* | *2005* | *Göteborg, Sweden* | *143* | *S* | *4-15* | *3.50* | *2.80* | *0.70* | *4.00* |
| *Gronlund* | *2006* | *Göteborg, Sweden* | *143* | *S* | *4-15* | *3.50* | *2.80* | *0.70* | *4.00* |
| Holmström | 2006 | Stockholm, Sweden | 217 | C | 10 | 3.23 | 0.92 | 2.30 | 0.40 |
| Abdi | 2008 | Stockholm, Sweden | 216 | S | 6-16 | 1.39 | 0.93 | 0.46 | 2.00 |
| Larsson | 2015 | Stockholm, Sweden | 217 | S | 10 | - | - | 2.30 | - |
| Voirol | 1912 | Basel, Switzerland | 939 | S | 6-15 | 0.85 | 0.64 | 0.21 | 3.04 |
| Franceschetti | 1966 | Geneva, Switzerland | 314 | S | 4-6 | 2.87 | 1.59 | 1.27 | 1.25 |
| Gansner | 1968 | Zürich Switzerland | 9,384 | S | 4-9 | 1.55 | 1.25 | 0.30 | 4.19 |
| Worth | 1906 | London, UK | 10,239 | S | children | 2.47 | 2.26 | 0.21 | 10.50 |
| Tyser | 1949 | London, UK | 460 | S | 1.5-5 | 2.39 | 2.39 | 0 | 23* |
| Sorsby | 1960 | UK | 1,033 | S | 17-27 | 3.97 | 3.29 | 0.68 | 4.86 |
| Graham | 1974 | Cardiff, UK | 4,784 | B | 5-6 | 4.39 | 3.62 | 0.77 | 4.70 |
| Bruce | 1991 | Bradford, UK | 339 | C | 3 | 4.13 | 2.95 | 1.18 | 2.50 |
| *Horwood* | *1993* | *London, UK* | *75* | *C* | *0.5* | *4.00* | *2.67* | *1.33* | *2.00* |
| Stayte | 1993 | Berkshire, UK | 6,483 | B | 2-5 | 2.28 | 1.88 | 0.40 | 4.70 |
| Newman | 1996 | Cambridge, UK | 6,794 | C | 3.5 | 0.50 | 0.21 | 0.29 | 0.70 |
| O'Connor | 2002 | East Midlands, UK | 169 | S | 10-12 | 2.37 | 1.78 | 0.59 | 3.00 |
| Anker | 2003 | Cambridge, UK | 5,142 | B | 0.6-0.8 | 0.25 | 0.18 | 0.08 | 2.25 |
| Williams | 2008 | Avon, UK | 7,538 | B | 7 | 2.19 | 1.70 | 0.49 | 3.46 |
| Hu | 2012 | Walsall, UK | 2,830 | S | 3-4 | 0.88 | 0.74 | 0.14 | 5.25 |
| Toufeeq | 2014 | Chesterfield, UK | 3,726 | S | 4-5 | 3.72 | 2.41 | 1.31 | 1.84 |
| Bruce | 2016 | Bradford, UK | 1,558 | S | 4-5 | 2.12 | 1.22 | 0.90 | 1.36 |
| Plotnikov | 2019 | Avon, UK | 5,172 | B | 7 | 3.37 | 2.83 | 0.54 | 5.11 |
| **INUIT/ESKIMO** |  |  |  |  |  |  |  |  |  |
| Duelund | 2024 | Multiple sites, Greenland (Inuit) | 462 | S | 6 | 0.21 | 0.21 | 0 | 3.00* |
| Duelund | 2025 | Multiple sites, Greenland (Inuit) | 274 | S | 6 | >0.36 | >0.36 | 0 | >3.00* |
| **NORTH AMERICA** |  |  |  |  |  |  |  |  |  |
| **CAUCASIAN** |  |  |  |  |  |  |  |  |  |
| Kornder | 1974 | British Columbia, Canada | 1,074 | B | 0.5-3 | 3.07 | 2.05 | 1.02 | 2.00 |
| Kornder | 1974 | British Columbia, Canada | 2,619 | S | 6 | 4.43 | 2.71 | 1.72 | 1.59 |
| Johnson | 1984 | Nain, Labrador, Canada USA | 80 | C |  | 1.25 | 0 | 1.25 | 0.33* |
| Woodruff | 1986 | New Brunswick, Canada | 6,080 | S | 6 | 3.98 | 2.70 | 1.28 | 2.11 |
| Robinson | 1999 | Ontario, Canada | 3,434 | S | 3-6 | 1.05 | 0.84 | 0.20 | 4.14 |
| Collins | 1925 | NY, IN, SC, USA | 12,134 | S | 6-16 | 0.91 | 0.73 | 0.17 | 4.29 |
| Knapp | 1931 | Duluth, MN, USA | 4,424 | C | adults | 0.34 | 0.14 | 0.20 | 0.67 |
| Knapp | 1931 | Duluth, MN, USA | 12,253 | S | children | 2.35 | 1.77 | 0.58 | 3.06 |
| Downing | 1945 | USA | 60,000 | S | 17-45 | 2.07 | 1.43 | 0.65 | 2.20 |
| Blum | 1959 | Oakland, USA | 1,163 | S | 6-13 | 4.64 | 3.18 | 1.46 | 2.18 |
| Roberts | 1972 | Multiple sites, USA | 7,119 | S | 6-11 | 2.38 | 1.12 | 0.60 | 1.88 |
| Roberts | 1975 | Multiple sites, USA | 6,768 | S | 12-17 | 3.95 | 1.46 | 2.47 | 0.59 |
| Roberts | 1978 | Multiple sites, USA | 10,126 | S, C | 1-74 | 3.3 | 1.3 | 2.0 | 0.65 |
| Nixon | 1985 | Indiana, USA | 1,219 |  | 0 | 34.78 | 2.13 | 32.65 | 0.07 |
| Sondhi | 1988 | Indiana, USA | 2,271 |  | 0 | 47.34 | 0.70 | 46.63 | 0.02 |
| Archer | 1989 | Indiana, USA | 3,324 |  | 0 | ~77 | ~3 | ~74 | 0.04 |
| Helveston | 1993 | Indiana, USA | 1,219 |  | 0 | 36.2 | 3.20 | 33.0 | 0.97 |
| Thorn | 1994 | Boston, MA, USA | 34 |  | 0 | 8.82 | 2.94 | 5.88 | 0.50 |
| Chew | 1994 | Multiple sites, USA | 17,931 | B | 0-7 | 5.40 | 4.13 | 1.27 | 3.25 |
| Friedman | 2009 | Baltimore, MD, USA | 259 | C | <2 | 2.32 | 1.54 | 0.77 | 1.50 |
| Friedman | 2009 | Baltimore, MD, USA | 771 | C | 2-6 | 3.50 | 1.43 | 2.08 | 0.83 |
| Cotter | 2011 | Baltimore, MD, USA | 1,861 | C | 0.5-6 | 2.85 | 1.77 | 1.07 | 1.65 |
| McKean-Cowdin | 2013 | Riverside, CA, USA | 1,514 | C | 0.5-6 | 3.04 | 2.31 | 0.73 | 3.16 |
| Mohney | 2021 | Rochester, MN, USA | 1,000 | S | 5-18 | 5.40 | 2.00 | 3.40 | 0.59 |
| **NATIVE AMERICAN** |  |  |  |  |  |  |  |  |  |
| Wick | 1976 | South Dakota, USA | 398 | S | 5-10 | 3.76 | 0.25 | 3.51 | 0.07 |
| Maples | 1990 | Oklahoma, USA | 792 | S | ~3-14 | 2.27 | 1.14 | 1.14 | 1.00 |
| Maples | 1990 | Minnesota, USA | 5,133 | S | ~3-14 | 0.45 | 0.21 | 0.23 | 0.92 |
| Lang | 2007 | Alaska, USA | 80 | C | 3-11 | 6.25 | 1.25 | 5.00 | 0.25 |
| Garvey | 2010 | Arizona, USA | 909 | S | 3-9 | 1.21 | 0.33 | 0.88 | 0.38 |
| Chiarelli | 2013 | Ontario, Canada | 146 | S | 2-13 | 4.11 | 0.68 | 3.42 | 0.20 |
| *ESKIMO / INUIT* |  |  |  |  |  |  |  |  |  |
| Woodruff | 1976 | Belcher Island, Canada | 138 | C | 0-79 | 5.80 | 3.62 | 2.17 | 1.67 |
| Johnson | 1984 | Nain, Labrador, Canada | 330 | C | 0-85 | 1.21 | 0.91 | 0.30 | 3.00 |
| *AFRICAN AMERICAN (including Caribbean)* |  |  |  |  |  |  |  |  |  |
| Friendly | 1978 | Washington DC, USA | 633 | S | 2.5-6 | 1.11 | 0.95 | 0.16 | 6.00 |
| Roberts | 1978 | Multiple sites, USA | 1,205 | S, C | 1-74 | 3.3 | 0.6 | 2.7 | 0.22 |
| De Vries | 1984 | Curacao, Caribbean | 905 | S | 9-17 | 1.88 | 0.99 | 0.88 | 1.13 |
| Chew | 1994 | Multiple sites, USA | 19,619 | B | 0-7 | 3.62 | 2.28 | 1.34 | 1.71 |
| Preslan | 1996 | Baltimore, MD, USA | 680 | C | 4-7 | 3.09 | 2.79 | 0.29 | 9.50 |
| Preslan | 1998 | Baltimore, MD, USA | 285 | C | 4-6 | 3.16 | 1.75 | 1.40 | 1.25 |
| Multi-ethnic | 2008 | Los Angeles, CA, USA | 3,005 | C | 0.5-6 | 2.46 | 1.10 | 1.36 | 0.80 |
| Friedman | 2009 | Baltimore, MD, USA | 274 | C | 0.5-2 | 0.36 | 0.36 | 0.00 | 3* |
| Friedman | 2009 | Baltimore, MD, USA | 994 | C | 2-6 | 2.41 | 1.21 | 1.21 | 1.00 |
| Cotter | 2011 | Baltimore, MD, USA | 3,604 | C | 0.5-6 | 2.28 | 1.11 | 1.11 | 1.00 |
| *ASIAN* |  |  |  |  |  |  |  |  |  |
| McKean-Cowdin | 2013 | Riverside, CA, USA | 1,522 | C | 0.5-6 | 3.48 | 1.38 | 2.10 | 0.66 |
| *LATINO/HISPANIC* |  |  |  |  |  |  |  |  |  |
| Choi | 1995 | Los Angeles, CA, USA | 2,192 | S | 6-7 | 1.23 | 0.59 | 0.64 | 0.92 |
| Multi-ethnic | 2008 | Los Angeles, CA, USA | 3,003 | C | 0.5-6 | 2.33 | 0.87 | 1.47 | 0.59 |
| Cotter | 2011 | Baltimore, MD, USA | 3,026 | C | 0.5-6 | 2.28 | 0.93 | 1.35 | 0.68 |
| *MULTIPLE ETHNICITIES* |  |  |  |  |  |  |  |  |  |
| Repka (a) | 2012 | Baltimore, MD, USA | 2,546 | S | 0.5-6 | 2.51 | 1.26 | 1.26 | 1.00 |
| Carpiuc | 2013 | Oakland, CA, USA | 14,050 | B | 0-10 | 2.20 | 1.60 | 0.60 | 2.68 |
| *LATIN AMERICA* |  |  |  |  |  |  |  |  |  |
| *NATIVE AMERICAN* |  |  |  |  |  |  |  |  |  |
| Urrets-Zavalía | 1961 | La Paz, Bolivia | 2,089 | S | ~6 | 2.01 | 0.29 | 1.72 | 0.17 |
| Urrets-Zavalía | 1961 | La Paz, Bolivia | 2,526 | S | ~6 | 0.28 | 0.08 | 0.20 | 0.40 |
| Belfort-Mattos | 1970 | Amazon, Brazil | 81 | C | 2-78 | 3.70 | 0 | 3.70 | 0.14* |
| Germano | 2017 | Avai City, Brazil | 377 | C | 0-100 | 1.19 | 0 | 1.19 | 0.11* |
| **CAUCASIAN** |  |  |  |  |  |  |  |  |  |
| Costa | 1979 | Sao Paulo, Brazil | 569 | S | 2-9 | 1.41 | 0.88 | 0.53 | 1.67 |
| Schimiti | 2001 | Ibipora, Brazil | 13,471 | S | 6-12 | 0.84 | 0.54 | 0.25 | 2.16 |
| Beer | 2003 | Sao Paulo, Brazil | 476 | C | 0.3-6 | 3.36 | 0.84 | 2.52 | 0.33 |
| Beer | 2003 | Sao Paulo, Brazil | 2,164 | S | 4-6 | 1.43 | 0.92 | 0.51 | 1.82 |
| de Sousa | 2012 | Botucatu City, Brazil | 500 | S | 5-12 | 0.80 | 0.20 | 0.60 | 0.33 |
| Shimauti | 2012 | Sao Paulo, Brazil | 10,994 | C | ? | 1.15 | 0.63 | 0.52 | 1.21 |
| Schaal | 2018 | South East Brazil | 1,852 | C | 1-11 | 1.62 | 1.13 | 0.32 | 3.50 |
| **LATINO/HISPANIC OR MIXED RACE** |  |  |  |  |  |  |  |  |  |
| Garcia | 2004 | Natal, Brazil | 1,015 | S | 5-46 | 2.76 | 0.59 | 2.17 | 0.27 |
| Couto Jr | 2007 | Rio de Janeiro, Brazil | 1,800 | S | 4-? | 1.39 | 0.78 | 0.61 | 1.27 |
| Couto Jr | 2010 | Rio de Janeiro, Brazil | 1,800 | S | 4-? | 0.33 | 0.22 | 0.11 | 2.00 |
| Fernandes | 2024 | Sao Paulo, Brazil | 17,973 | S | 3-17 | 1.89 | 1.05 | 0.83 | 1.26 |
| Maul | 2000 | Santiago, Chile | 5,303 | C | 5-15 | 9.86 | 2.34 | 7.52 | 0.24 |
| Vasquez | 2014 | Bogota, Colombia | 855 | S | 3-18 | 3.04 | 1.64 | 1.40 | 1.17 |
| Márquez Galvis | 2017 | Pereira, Colombia | 348 | S | 1-6 | 1.72 | 0.86 | 0.86 | 1.00 |
| Molinari | 2005 | Sierra, Ecuador | 6,143 | S | 4-16 | 0.63 | 0.36 | 0.28 | 1.29 |
| Juárez-Muñoz | 1996 | Mexico City, Mexico | 343 | S | 3-6 | 1.17 | 0.58 | 0.58 | 1.00 |
| Ohlsson | 2003 | Monterrey, Mexico | 1,035 | S | 12-13 | 1.26 | 0.68 | 0.58 | 1.17 |
| **EAST ASIAN** |  |  |  |  |  |  |  |  |  |
| Beiguelman | 1964 | Sao Paulo, Brazil | 296 | C | >18 | 5.41 | 4.05 | 1.35 | 3.00 |
| **AFRICA (NORTH, EAST, SOUTH)** |  |  |  |  |  |  |  |  |  |
| Elsahn | 2014 | Alexandria, Egypt | 6,029 | S | 6-12 | 0.13 | 0.12 | 0.02 | 7.0 |
| Abdelrahman | 2020 | Sohag City, Egypt | 584 | S | 5.5-7 | 0.51 | 0.51 | 0 | 7* |
| Giorgis | 2001 | Addis Abeba, Ethiopia | 1,894 | C | <5 | 1.43 | 1.06 | 0.37 | 2.86 |
| Tegegne | 2021 | Bahir Dar City, Ethiopia | 611 | C | 6-18 | 5.07 | 4.09 | 0.98 | 4.17 |
| Fikre | 2022 | Gondar City, Ethiopia | 848 | S | 3-8 | 2.81 | 1.79 | 1.02 | 1.75 |
| Onsomu | 2003 | Nairobi, Kenya | 559 | S | 3-5 | 2.86 | 0.18 | 2.68 | 0.07 |
| Mutie | 2008 | Nairobi (East), Kenya | 602 | S | 3-10 | 0.33 | 0.33 | 0 | 5* |
| Gordon | 1982 | Maseru, Lesotho | 1,296 | S | 3-9+ | 0.69 | 0.54 | 0.15 | 3.50 |
| Auzemery | 1995 | Madagascar | 1,081 | S | 8-14 | 1.11 | 0.69 | 0.42 | 1.64 |
| Freedman | 1973 | Namibia | 680 | C | 1-85 | 0.29 | 0.15 | 0.15 | 1.00 |
| Yassur | 1972 | Rwanda | 1,550 | S | 10-18 | 0.84 | 0.32 | 0.52 | 0.62 |
| Abdi Ahmed | 2020 | Hargeisa, Somalia | 1,204 | S | 6-15 | 0.75 | 0.33 | 0.42 | 0.80 |
| Naidoo | 2003 | Durban, South Africa | 4,890 | C | 5-15 | 1.23 | 0.88 | 0.33 | 2.69 |
| Zeidan | 2007 | Khartoum City, Sudan | 916 | C | <16 | 0.33 | 0 | 0.33 | 0.2* |
| Taha | 2015 | Khartoum City, Sudan | 768 | S | 5-14 | 2.73 | 2.21 | 0.39 | 5.67 |
| Alrasheed | 2016 | South Darfur, Sudan | 1,666 | S | 6-15 | 0.30 | 0.18 | 0.12 | 1.50 |
| Kingo | 2009 | Dar es Salaam, Tanzania | 400 | S | 6-17 | 0 | 0 | 0 | 1* |

| AFRICA (WEST) |  |  |  |  |  |  |  |  |  |
| --- | --- | --- | --- | --- | --- | --- | --- | --- | --- |
| Pergens | 1898 | Congo | 100 | C |  | 0 | 0 | 0 | 1* |
| Holm | 1939 | Gabon | 1,931 | C | 16-55 | 0.62 | 0.10 | 0.52 | 0.20 |
| Kumah | 2013 | Ashanti, Ghana | 2,435 | S | 12-15 | 1.77 | 0 | 1.77 | 0.01* |
| Nartey | 2016 | Accra, Ghana | 811 | S | 6-16 | 0 | 0 | 0 | 1* |
| Abdul-Kabir | 2017 | Kumasi, Ghana | 67 | S | 17-25 | 1.49 | 0 | 1.49 | 0.33* |
| Hertz | 1964 | Liberia | 1,020 | S | 6-20 | 1.37 | 0 | 1.37 | 0.34* |
| Abiose | 1980 | Kaduna, Nigeria | 5,220 | S | 12-20 | 0.67 | 0.3 | 0.44 | 0.52 |
| Baiyerou-Agbeja | 1998 | Ibadan, Nigeria | 759 | S | 0-16 | 0.26 | 0.26 | 0 | 5* |
| Abah | 2001 | Zaria, Nigeria | 327 | S | 5-17 | 0.31 | 0.31 | 0 | 3* |
| *Ajaiyeoba* | 2005 | *Ilesa, Nigeria* | *1,144* | *S* | *4-24* | *0.26* | *0.09* | *0.17* | *0.50* |
| *Ajaiyeoba* | 2006 | *Ilesa, Nigeria* | *1,144* | *S* | *4-24* | *0.26* | *0.09* | *0.17* | *0.50* |
| Atamah | 2005 | Benin City, Nigeria | 600 | C | 17-75 | 1.17 | 0.33 | 0.83 | 0.40 |
| *Azonobi* | 2009 | *Ilorin, Nigeria* | *7,288* | *S* | *2-16* | *0.44* | *0.30* | *0.14* | *2.20* |
| *Azonobi* | 2009 | *Ilorin, Nigeria* | *7,288* | *S* | *2-16* | *0.44* | *0.30* | *0.14* | *2.20* |
| Ayanniyi | 2010 | Ilorin, Nigeria | 1,393 | S | 4-15 | 0.36 | 0.14 | 0.22 | 0.66 |
| Akpe | 2014 | Benin City, Nigeria | 2,139 | S | 5-19 | 0.89 | 0.56 | 0.33 | 1.70 |
| Ezinne | 2018 | Anambra, Nigeria | 998 | S | 5-15 | 2.81 | 0.70 | 2.10 | 0.33 |
| Chibueze | 2020 | Ogun State, Nigeria | 360 | S | 9-22 | 1.39 | 0.83 | 0.56 | 1.50 |
| Adejumo | 2021 | Ijebu, SW Nigeria | 560 | S | 3-5 | 0.89 | 0.71 | 0.18 | 4.00 |
| Atuanya | 2024 | Benin City, Nigeria | 300 | S | 5-13 | 3.67 | 2.00 | 1.67 | 1.20 |
| MIDDLE EAST |  |  |  |  |  |  |  |  |  |
| Abdiyeva | 2021 | Baku, Azerbaijan | 2,700 | S | 5-19 | 3.00 | 0.6 | 2.4 | 0.23 |
| Fotouhi | 2007 | Dezful, Iran | 5,544 | S | 7-18 | 0.72 | 0.51 | 0.22 | 2.33 |
| Ostadi Moghaddam | 2008 | Mashhad, Iran | 2,150 | S | 6-12 | 3.12 | 0.88 | 2.19 | 0.40 |
| Jamali | 2009 | Shahrood, Iran | 815 | S | 6-7 | 1.10 | 0.61 | 0.49 | 1.24 |
| Yekta | 2010 | Shiraz, Iran | 2,683 | S | ~10-17 | 1.89 | 0.56 | 1.16 | 0.48 |
| Faghihi | 2011 | Mashhad, Iran | 2,150 | S | 6-21 | 2.97 | 0.88 | 2.09 | 0.42 |
| Kwak | 2011 | Alborz Mountains, Iran | 243 | C | ? | 13.58 | 1.65 | 11.93 | 0.14 |
| Faghihi | 2012 | Varamin, Iran | 1,133 | S | 14-18 | 1.32 | 0.44 | 0.88 | 0.50 |
| Yekta | 2012 | Bojnurd, Iran | 1,551 | S | 6-17 | 1.87 | 0.52 | 1.35 | 0.39 |
| Hashemi | 2015 | Iran (multiple sites) | 3,675 | S | 7 | 1.71 | 0.44 | 1.27 | 0.34 |
| Rajavi | 2015 | Tehran, Iran | 2,410 | S | 7-12 | 2.28 | 1.00 | 1.29 | 0.77 |
| *Yekta* | 2016 | *Dezful, Iran* | *1,130* | *S* | *6-15* | *1.94* | *0.71* | *1.24* | *0.57* |
| *Yekta* | 2017 | *Dezful, Iran* | *1,130* | *S* | *6-15* | *1.94* | *0.71* | *1.24* | *0.57* |
| Hashemi | 2017 | Tehran (North), Iran | 1,414 | C | 3-93 | 4.74 | 0.30 | 4.44 | 0.68 |
| Hashemi | 2017 | Dezful, Iran | 1,834 | C | 3-93 | 3.97 | 0.53 | 3.44 | 0.15 |
| Hamidi | 2019 | Bojnurd, Iran | 6,600 | S | 3-6 | 0.36 | 0.24 | 0.12 | 2.0 |
| Hashemi | 2020 | Shahrekord (Isfahan), Iran | 726 | S | 18-25 | 1.52 | 0.28 | 1.24 | 0.18 |
| Hashemi | 2022 | Tehran, Iran | 2,227 | C | >60 | 5.19 | 0.59 | 4.60 | 0.13 |
| Derakhshan | 2024 | Mashhad, Iran | 5,054 | C | 0.5-85 | 1.97 | 0.51 | 1.46 | 0.35 |
| Abd | 2022 | Karbala, Iraq | 800 | S | 6-12 | 2.50 | 0.50 | 2.00 | 0.25 |
| Halboos | 2023 | Babylon, Iraq | 1,016 | S | 5-12 | 5.33 | 3.16 | 2.17 | 1.45 |
| Mukhaizer | 2023 | Baghdad, City, Iraq | 8,850 | S | 6-14 | 0.53 | 0.37 | 0.16 | 2.36 |
| Neumann | 1971 | Haifa, Israel | 6,400 | B | 1-2.5 | 2.09 | 1.41 | 0.61 | 2.31 |
| Friedman | 1980 | Haifa, Israel | 38,000 | B | 1-2.5 | 1.25 | 0.95 | 0.30 | 3.13 |
| Friedmann | 1980 | Negev, Israel | 3,375 | B | 0.5-3 | 4.56 | 0.71 | 3.08 | 0.23 |
| Maaaita | 2003 | Jordan | 1,725 | C | 6-14 | 0.52 | 0.41 | 0.12 | 3.50 |
| Lithander | 1998 | Oman | 6,292 | S | 6-7, 11-12 | 0.65 | 0.41 | 0.24 | 1.71 |
| Labadi | 2022 | Nablus, Palestine | 727 | S | 3-5 | 0.83 | 0.69 | 0.14 | 5.0 |
| Badr | 1981 | Al Majma'ah, Saudi Arabia | 833 | S | children | 1.80 | 1.08 | 0.72 | 1.50 |
| Abolfotouh | 1994 | Abha, Saudi Arabia | 971 | S | 6-12 | 2.99 | 1.96 | 1.03 | 1.90 |
| Bardisi | 2002 | Jeddah, Saudi Arabia | 609 | S | 3-6 | 0.70 | 0.53 | 0.18 | 2.94 |
| Gorham | 2021 | Syria | 91 | C | 1-16 | 13.2 | 9.9 | 3.3 | 3.00 |
| Turacli | 1995 | Ankara, Turkey | 23,810 | S | 5-12 | 2.28 | 1.37 | 0.91 | 1.51 |
| Aslan | 2013 | Kahramanmaras, Turkey | 116 | C | 6-12 | 2.59 | 1.72 | 0.86 | 2.00 |
| Caca | 2013 | Diyarbakir, Turkey | 21,062 | S | 6-14 | 2.09 | 1.20 | 0.89 | 1.35 |
| Gursoy | 2013 | Eskisehir, Turkey | 709 | S | 7-8 | 2.12 | 1.41 | 0.71 | 2.00 |
| Celikay | 2016 | Ankara, Turkey | 686 | S | 4-12 | 0.73 | 0.44 | 0.29 | 1.50 |
| **SOUTH ASIA (NORTHWEST)** |  |  |  |  |  |  |  |  |  |
| McLaren | 1961 | Gujarat, India (migrants, Tanzania) | 359 | S | 6-14 | 1.11 | 1.11 | 0 | 9* |
| Agarwal | 1966 | Delhi, India | 4,201 | S | 3~18 | 1.12 | 0.52 | 0.60 | 0.88 |
| Gupta | 2000 | Aligarh, India | 310 | S | 4-12 | 2.90 | 1.94 | 0.97 | 2.0 |
| Murthy | 2002 | New Delhi, India | 6,447 | C | 5-15 | 0.51 | 0.29 | 0.22 | 1.36 |
| Bedi | 2016 | Rajasthan, India | 2,754 | S | 10-18 | 0.26 | 0.11 | 0.15 | 0.75 |
| Mohan | 2017 | Rajasthan, India | 16,168 | S | 6-14 | 1.78 | 0.95 | 0.83 | 1.14 |
| Awan | 1995 | Pakistan (North) | 1,306 | S | ~6-10 | 2.45 | 0.61 | 1.84 | 0.33 |
| Shaikh | 2005 | Karachi, Pakistan | 5,110 | C | 5-15 | 0.57 | 0.49 | 0.08 | 6.25 |
| Awan | 1998 | Afghan refugees | 1,156 | C | 0-60+ | 1.38 | 0.52 | 0.87 | 0.60 |
| **SOUTH ASIA (SOUTHEAST)** |  |  |  |  |  |  |  |  |  |
| Asaduzzaman | 2024 | Dhaka, Bangladesh | 200 | S | 3-6 | 2.00 | 0 | 2.00 | 0.11* |
| Kuruville | 1978 | Udupi, Karnataka, India | 8,496 | S | 5-12 | 1.51 | 0.38 | 1.13 | 0.33 |
| Datta | 1983 | Calcutta, India | 24,007 | S | 5-13 | 0.22 | 0.13 | 0.09 | 1.44 |
| Reddy | 1987 | Kakinada, India | 3,675 | S | 11-15 | 0.27 | 0.08 | 0.19 | 0.43 |
| Kalikivayi | 1997 | Hyderabad, India | 4,029 | S | 3-18 | 0.74 | 0.22 | 0.52 | 0.42 |
| Dandona | 2002 | Mahabubnagar District, Telangana, India | 4,074 | C | 7-15 | 1.87 | 0.54 | 1.33 | 0.41 |
| Singh | 2011 | Bhopal, India | 20,800 | S | 5-16 | 0.11 | 0.04 | 0.06 | 0.69 |
| Naik | 2013 | Ahmednagar, India | 1,095 | S | 6-15 | 1.55 | 0 | 1.55 | 0.03* |
| Kemmanu | 2016 | Karnataka, India | 23,100 | C | 0-15 | 0.45 | 0.11 | 0.34 | 0.33 |
| Akarkar | 2019 | Goa, India | 817 | S | 6-10 | 1.10 | 0.86 | 0.24 | 3.50 |
| Saxena | 2019 | Indore District, Madhya Pradesh, India | 1,322 | S | 5-16 | 2.80 | 0.76 | 2.04 | 0.37 |
| Agrawal | 2020 | Raipur, India | 1,557 | S | 5-15 | 0.19 | 0 | 0.19 | 0.14* |
| Atiya | 2020 | Chennai, India | 75 | S | 24-48 | 12.00 | 0 | 12.00 | 0.53* |
| Gupta | 2021 | Surat, India | 1,747 | S | 5-12 | 0.37 | 0.63 | 1.43 | 0.44 |
| Vasantha | 2021 | Kollam, Kerala, India | 3,100 | S | 5-12 | 1.35 | 0.74 | 0.61 | 1.21 |
| Hegde | 2025 | Satara District, India | 1,062 | S | 5-14 | 1.04 | 0.47 | 0.56 | 0.83 |

| EAST ASIA |  |  |  |  |  |  |  |  |  |
| --- | --- | --- | --- | --- | --- | --- | --- | --- | --- |
| Liang | 1984 | Taiwan, China | 5,507 | S | 6-11 | 1.69 | 0.83 | 0.86 | 0.97 |
| Wang | 1989 | Taiwan, China | 11,806 | S | 6-11 | 0.81 | 0.34 | 0.47 | 0.72 |
| Edwards | 1991 | Hong Kong, China | 122 |  | 0.4-0.8 | 1.64 | 0.00 | 1.64 | 0.23* |
| Ji | 1994 | Xinjiang, China | 4,125 | S | 4-14 | 1.16 | 0.39 | 0.78 | 0.50 |
| See | 1996 | Taiwan, China | 862 | S | 6-11 | 1.39 | 0.46 | 0.93 | 0.50 |
| He | 2004 | Guangzhou, China | 4,364 | C | 5-15 | 3.00 | 0.42 | 2.58 | 0.16 |
| He | 2007 | Yangxi, China | 2,454 | S | 13-17 | 1.69 | 0.37 | 1.22 | 0.30 |
| Lai | 2009 | Taiwan, China | 618 | S | 3-6 | 0.81 | 0.49 | 0.32 | 1.53 |
| Fan | 2011 | Hong Kong, China | 601 | S | 3-6 | 2.33 | 0.50 | 1.83 | 0.27 |
| Fan | 2011 | Hong Kong, China | 823 | S | 3-6 | 1.70 | 0.24 | 1.46 | 0.17 |
| Jin | 2011 | Nanchang, China | 4,376 | S | 3-6 | 2.22 | 0.73 | 1.49 | 0.49 |
| Pi | 2012 | Yongchuan, China | 3,079 | C | 6-15 | 0.26 | 0.10 | 0.16 | 0.63 |
| Lin | 2013 | Shantou, China | 7,464 | S | 6-19 | 2.90 | 0.19 | 2.71 | 0.07 |
| Fu | 2014 | Anyang, China | 2,260 | S | ~12-14 | 4.60 | 0.08 | 4.51 | 0.02 |
| Lin | 2014 | Shantou, China | 7,464 | S | 6-17 | 6.78 | 0.19 | 6.59 | 0.03 |
| Zhu | 2015 | Nanjing, China | 5,831 | S | 3-6 | 5.40 | 0.77 | 4.63 | 0.17 |
| Chen | 2016 | Nanjing, China | 5,667 | S | 3-6 | 5.33 | 0.76 | 4.57 | 0.16 |
| Lin | 2016 | Shantou, China | 7,464 | S | 6-17 | 10.88 | 0.19 | 10.69 | 0.02 |
| Pan | 2017 | Yunnan, China | 9,263 | S | 6-14 | 3.15 | 0.30 | 2.85 | 0.11 |
| Zhu | 2019 | Yunnan, China | 3,050 | S | 7-8, 13-14 | 1.90 | 0.16 | 1.74 | 0.09 |
| Chen | 2021 | Nanjing, China | 2,018 | S | 3 | 2.43 | 0.20 | 2.23 | 0.09 |
| Chen | 2021 | Nanjing, China | 1,766 | ? | 4-5 | 3.57 | 0.34 | 3.23 | 0.11 |
| Lu | 2008 | Maqin County, Tibet | 1,084 | S | 6-14 | 2.49 | 0.37 | 2.12 | 0.17 |
| He H | 2020 | Lhasa, Tibet, China | 1,856 | S | 6-8 | 2.75 | 0.4 | 2.3 | 0.19 |
| Wang | 2021 | Nanjing, China | 1,986 | S | 4-5 | 5.40 | 0.56 | 4.84 | 0.12 |
| Zhang | 2021 | Hong Kong, China | 4,273 | S | 6-8 | 3.02 | 0.28 | 2.74 | 0.10 |
| Nakagawa | 1954 | Sapporo, Japan | 4,171 | S | 6-15 | 1.51 | 0.26 | 1.25 | 0.21 |
| Majima | 1960 | Tokyo, Japan | 3,033 | S | 6-10 | 0.56 | 0.23 | 0.33 | 0.70 |
| Nakajima | 1960 | Shizuoka, Japan | 2,874 | S | 6~12 | 0.28 | 0.10 | 0.17 | 0.60 |
| Nakajima | 1960 | Shizuoka, Japan | 1,159 | S | ~6-12 | 0.60 | 0.43 | 0.17 | 2.50 |
| Nakajima | 1960 | Shizuoka, Japan | 3,033 | S | 6-11 | 0.56 | 0.23 | 0.33 | 0.70 |
| Harada | 1961 | Tokyo, Japan | 1,058 | S | 4-5 | 1.98 | 1.04 | 0.95 | 1.10 |
| Yazawa | 1973 | Hoya City, Japan | 7,989 | S | 6-12 | 0.65 | 0.24 | 0.41 | 0.58 |
| Yazawa | 1973 | Hoya City, Japan | 2,700 | S | ~12-15 | 0.48 | 0.04 | 0.44 | 0.08 |
| Inatomi | 1976 | Tokyo, Japan | 4,963 | S | 6-15 | 0.56 | 0.28 | 0.28 | 1.00 |
| Maruo | 1977 | Tokyo, Japan | 10,512 | S | 6-14 | 2.18 | 0.95 | 1.23 | 0.79 |
| Matsuo | 2005 | Okayama, Japan | 86,531 | S | 6-12 | 0.98 | 0.28 | 0.69 | 0.41 |
| Matsuo (a) | 2007 | Okayama, Japan | 84,619 | S | 6-12 | 0.84 | 0.22 | 0.62 | 0.36 |
| Matsuo (b) | 2007 | Okayama, Japan | 33,929 | ? | 1.5 | 0.06 | 0.02 | 0.04 | 0.50 |
| Goseki | 2017 | Tokyo, Japan | 1,214 | C | 20-86 | 1.32 | 0.16 | 1.15 | 0.14 |
| Satou | 2018 | Tochigi, Japan | 760 | S | 3-6 | 1.71 | 0.26 | 1.45 | 0.15 |
| Yu | 1991 | Seoul, South Korea | 1,211 | S | 6 | 0.99 | 0.33 | 0.66 | 0.50 |
| Rah | 1997 | Seoul, South Korea | 9,054 | S | 5-6 (?) | 3.56 | 0.66 | 2.89 | 0.23 |
| Yoon | 2011 | South Korea | 14,464 | C | 3-95 | 1.30 | 0.20 | 1.10 | 0.18 |
| Yoon | 2011 | South Korea | 3,416 | C | 3-18 | 1.49 | 0.23 | 1.26 | 0.19 |
| Lee | 2017 | South Korea | 30,538 | C | 3-70+ | 1.20 | 0.20 | 0.99 | 0.20 |
| Han | 2018 | South Korea | 5,935 | C | 5-18 | 1.63 | 0.22 | 1.42 | 0.15 |
| Lee | 2021 | South Korea | 13,411 | C | 5-70+ | 7.53 | 0.40 | 7.13 | 0.06 |
| Casson | 2012 | Vientiane, Laos | 2,842 | S | 6-11 | 1.34 | 0.21 | 1.13 | 0.19 |
| Teoh | 1982 | Petaling Jaya, Malaysia | 650 | S | 8 | 2.00 | 0.15 | 1.85 | 0.08 |
| Goh | 2005 | Kuala Lumpur, Malaysia | 4,634 | C | 7-15 | 0.67 | 0.15 | 0.52 | 0.29 |
| Premseenthil | 2013 | Kuching, Malaysia | 400 | S | 4-6 | 0.25 | 0 | 0.25 | 0.33* |
| Chew | 2018 | Segamat, Malaysia | 1,287 | S | 4-6 | 0.23 | 0.08 | 0.16 | 0.50 |
| Nepal | 2003 | Kathmandu, Nepal | 1,100 | S | 5-16 | 1.64 | 0.09 | 1.55 | 0.06 |
| Shrestha | 2006 | Kathmandu, Nepal | 1,816 | S | 5-16 | 1.21 | 0.11 | 1.10 | 0.10 |
| Sherpa | 2011 | Dhulikhel, Nepal | 466 | S | 0-15 | 3.01 | 0 | 0.43 | 0.20* |
| Shrestha | 2011 | Kathmandu, Nepal | 4,228 | S | 4-20 | 3.12 | 0.24 | 2.89 | 0.08 |
| Pant | 2014 | Kathmandu, Nepal | 569 | C | <18 | 2.99 | 0.35 | 2.64 | 0.13 |
| Sherpa | 2014 | Chitwan District, Nepal | 332 | S | 0-15 | 3.01 | 0 | 3.01 | 0.48* |
| Chia | 2010 | Singapore (STARS) | 2,992 | C | 2.5-6 | 0.77 | 0.10 | 0.67 | 0.15 |
| Chia | 2013 | Singapore | 2,992 | C | 0.5-6 | 0.77 | 0.10 | 0.67 | 0.15 |
| Konyama | 1972 | Bangkok, Thailand | 2,415 | S | 3-18 | 1.12 | 0.37 | 0.75 | 0.50 |
| Mahachaiyaku I | 1997 | Bangkok, Thailand | 1,219 | S | 6-12 | 0.16 | 0 | 0.16 | 0.20* |
| Tananuvat | 2004 | Chiang Mai, Thailand | 1,084 | S | 6-7 | 0.90 | 0.26 | 0.64 | 0.41 |
| Jenchitr | 2012 | Bangkok, Thailand | 1,780 | S | 6-11 | 2.53 | 1.57 | 0.96 | 1.65 |
| Paudel | 2014 | Ba Ria (Vung Tau), Vietnam | 2,238 | S | 12-15 | 0.13 | 0.13 | 0 | 7* |
| **OCEANIA (AUSTRALIA AND MELANESIA)** |  |  |  |  |  |  |  |  |  |
| **INDIGENOUS** |  |  |  |  |  |  |  |  |  |
| Mann | 1966 | Australia (Northwest) | 1,680 | C? |  | 0.30 | 0 | 0.30 | 0.09* |
| Mann | 1968 | Australia (Desert) | 805 | C |  | 0.37 | 0 | 0.37 | 0.14* |
| Royal Austral College | 1980 | Australia | 62,116 | C | 0-60+ | 0.73 | 0.18 | 0.55 | 0.33 |
| Hopkins | 2014 | Queensland, Australia | 181 | S | 5-13 | 0 | 0 | 0 | 1.00 |
| Hopkins | 2016 | Queensland, Australia | 181 | S | 5-13 | 0 | 0 | 0 | 1.00 |
| Ward | 1955 | Fiji | 4,000 | C | 0-60 | 0 | 0 | 0 | 1.00 |
| Mann | 1969 | New Zealand, Maori | 333 | C |  | 2.10 | 1.50 | 0.6 | 2.50 |
| Mann | 1955 | Papua-New Guinea | 13,751 | C |  | 0.15 | 0.11 | 0.04 | 3.0 |
| Elliott | 1965 | Tokelau Islands | 1,862 | C |  | 0 | 0 | 0 | 1.00 |
| Elliott | 1965 | Tokelau Islands | 476 | C | children | 0 | 0 | 0 | 1.00 |
| Elliott | 1965 | Tokelau Islands | 884 | C | adults | 0 | 0 | 0 | 1.00 |
| Verlee | 1968 | Solomon Islands | 616 | C | 0-89 | 1.95 | 0.16 | 1.79 | 0.09 |
| **CAUCASIAN** |  |  |  |  |  |  |  |  |  |
| Brown | 1976 | Sydney, Australia | 5,426 | S | 4-7 | 3.43 | 1.79 | 1.64 | 1.09 |
| Macfarlane | 1987 | Brisbane, Australia | 877 | S | 6-11 | 2.51 | 1.60 | 0.91 | 1.75 |
| Robaei (a) | 2006 | Sydney, Australia | 1,107 | S | 5.5-8.5 | 3.07 | 1.99 | 1.08 | 1.83 |
| Robaei (b) | 2006 | Sydney, Australia | 1,406 | S | 12 | 1.99 | 1.14 | 0.85 | 1.34 |
| Sharbini | 2015 | Sydney, Australia | 1,131 | C | 0.5-6 | 3.36 | 1.24 | 2.12 | 0.58 |
| Ward | 1955 | Fiji | 438 | C | 0-60 | 0.46 | 0.46 | 0 | 5* |
| Simpson | 1984 | New Zealand (West) | 988 | B | 6-7 | 2.63 | 2.02 | 0.61 | 3.33 |

| *MULTIPLE ETHNICITIES* |  |  |  |  |  |  |  |  |  |
| --- | --- | --- | --- | --- | --- | --- | --- | --- | --- |
| *Robaei* | 2006 | *Sydney, Australia* | 1,739 | S | 5.5-8.5 | 2.30 | 1.50 | 0.81 | 1.86 |
| *Robaei* | 2006 | *Sydney, Australia* | 2,353 | S | 12 | 2.76 | 0.89 | 1.15 | 0.78 |
| Sharbini |  |  |  |  |  |  |  |  |  |
| *Leone* | 2009 | *Sydney, Australia* | 1,740 | S | 6 | 2.30 | 1.49 | 0.80 | 1.86 |
| *Leone* | 2009 | *Sydney, Australia* | 2,353 | S | 12 | 2.04 | 0.89 | 1.15 | 0.78 |
| Pai | 2013 | Sydney, Australia | 1,188 |  |  |  | 0.34 | 0.08 | 4.00 |
| Sharbini | 2015 | Sydney, Australia | 491 | C | 0.5-6.0 | 3.05 | 1.22 | 1.83 | 0.67 |
| Sharbini | 2015 | Sydney, Australia | 2,353 | C | 11-14 | 2.68 | 1.23 | 1.44 | 0.85 |
| Ward | 1955 | Fiji (South Asian, others) | 1,447 | C | 0-60 | 0 | 0 | 0 | 1.00 |
| Anstice | 2012 | New Zealand | 3,273 | S, C | ~4 | 0.24 | 0.18 | 0.06 | 3.00 |
| *EAST ASIAN* |  |  |  |  |  |  |  |  |  |
| Robaei | 2006 | Sydney, Australia | 632 | S | 5.5-8.5 | 2.06 | 0.79 | 1.27 | 0.63 |
| Robaei | 2006 | Sydney, Australia | 947 | S | 12 | 2.11 | 0.53 | 1.58 | 0.34 |
| Sharbini | 2015 | Sydney, Australia | 516 | C | 0.5-6.0 | 2.13 | 0.39 | 1.74 | 0.22 |
| Ward | 1955 | Fiji (Chinese) | 205 | C | 0-60 | 1.46 | 0.49 | 0.98 | 0.50 |

**Supplemental Table 2.**
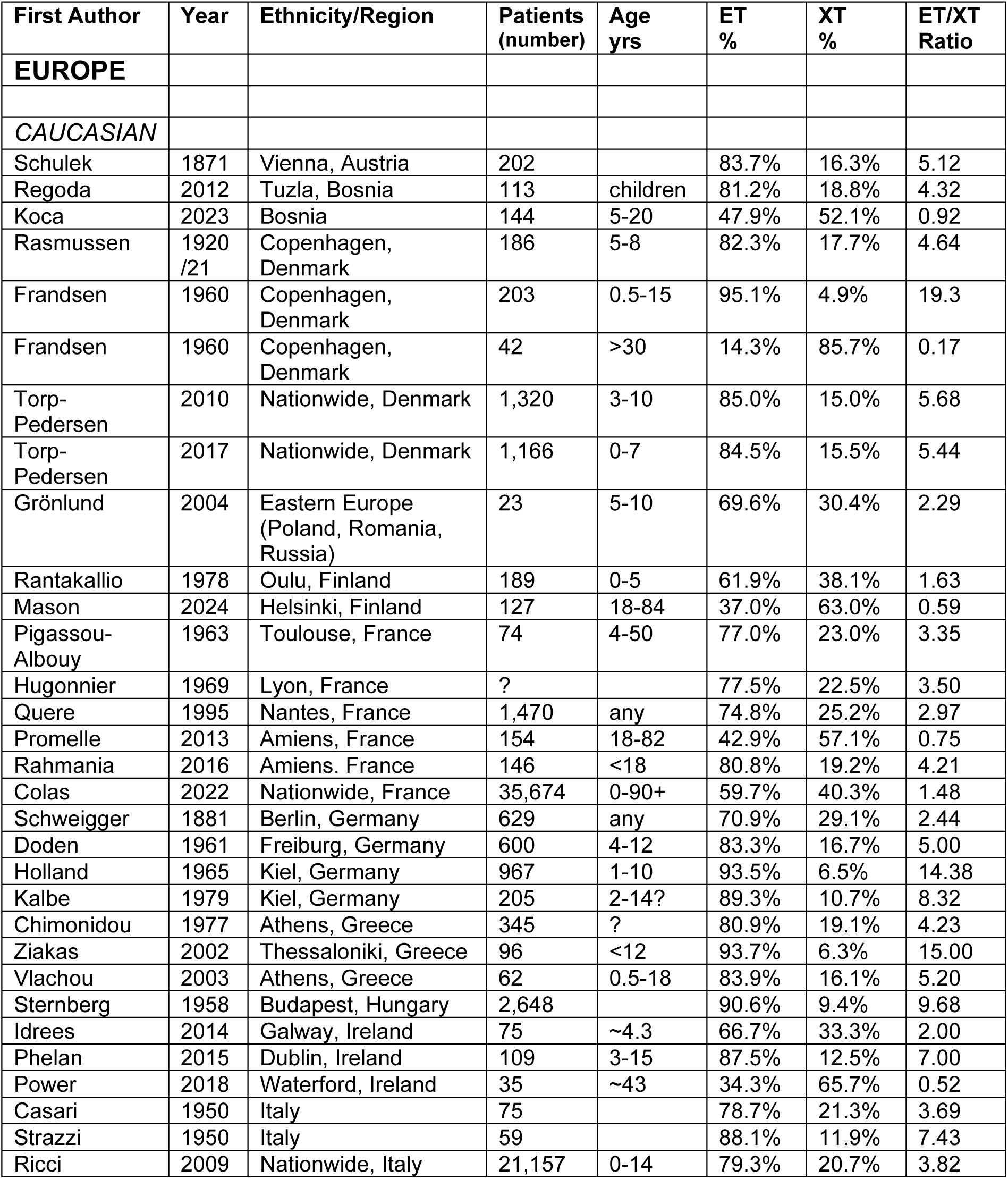
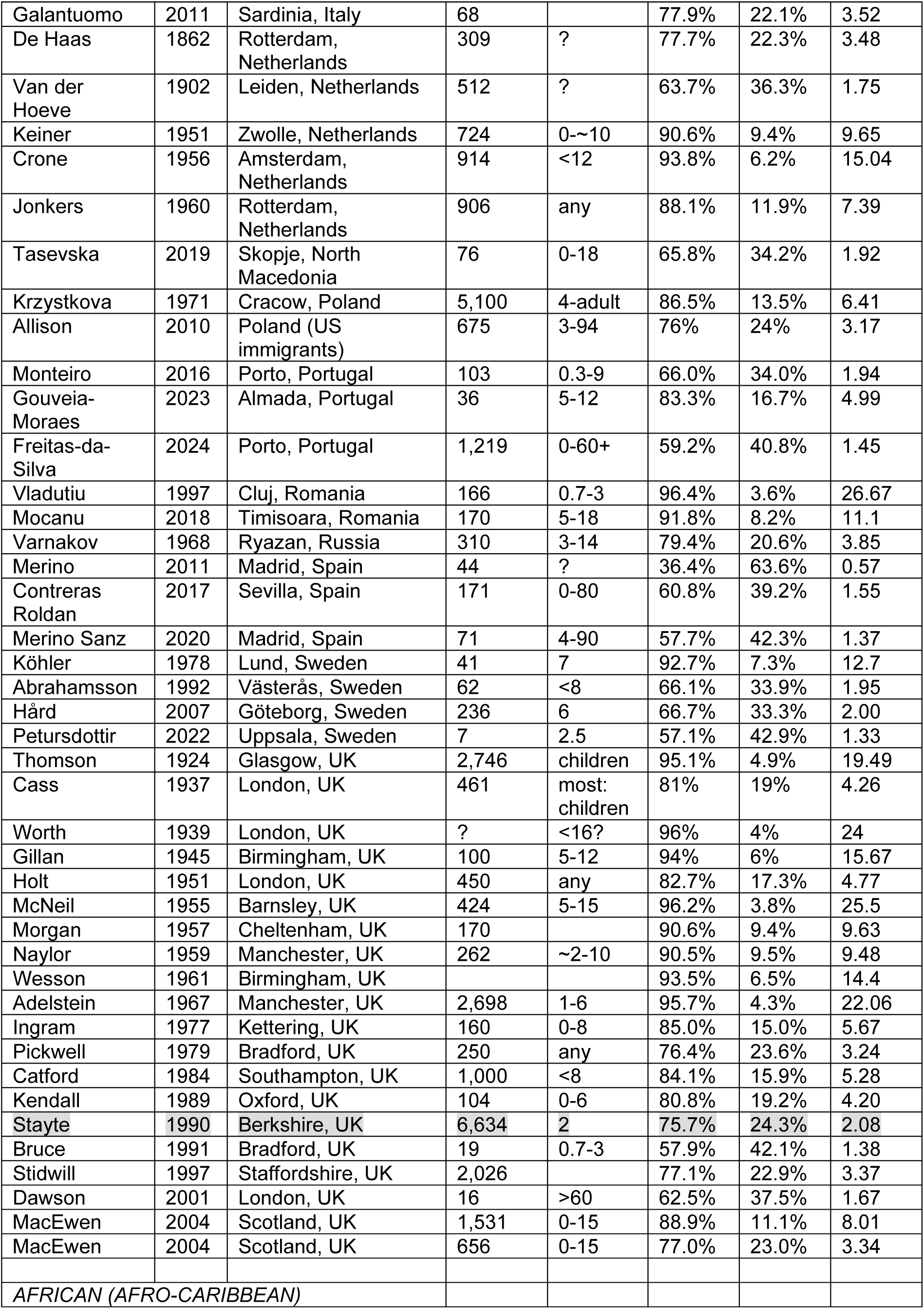
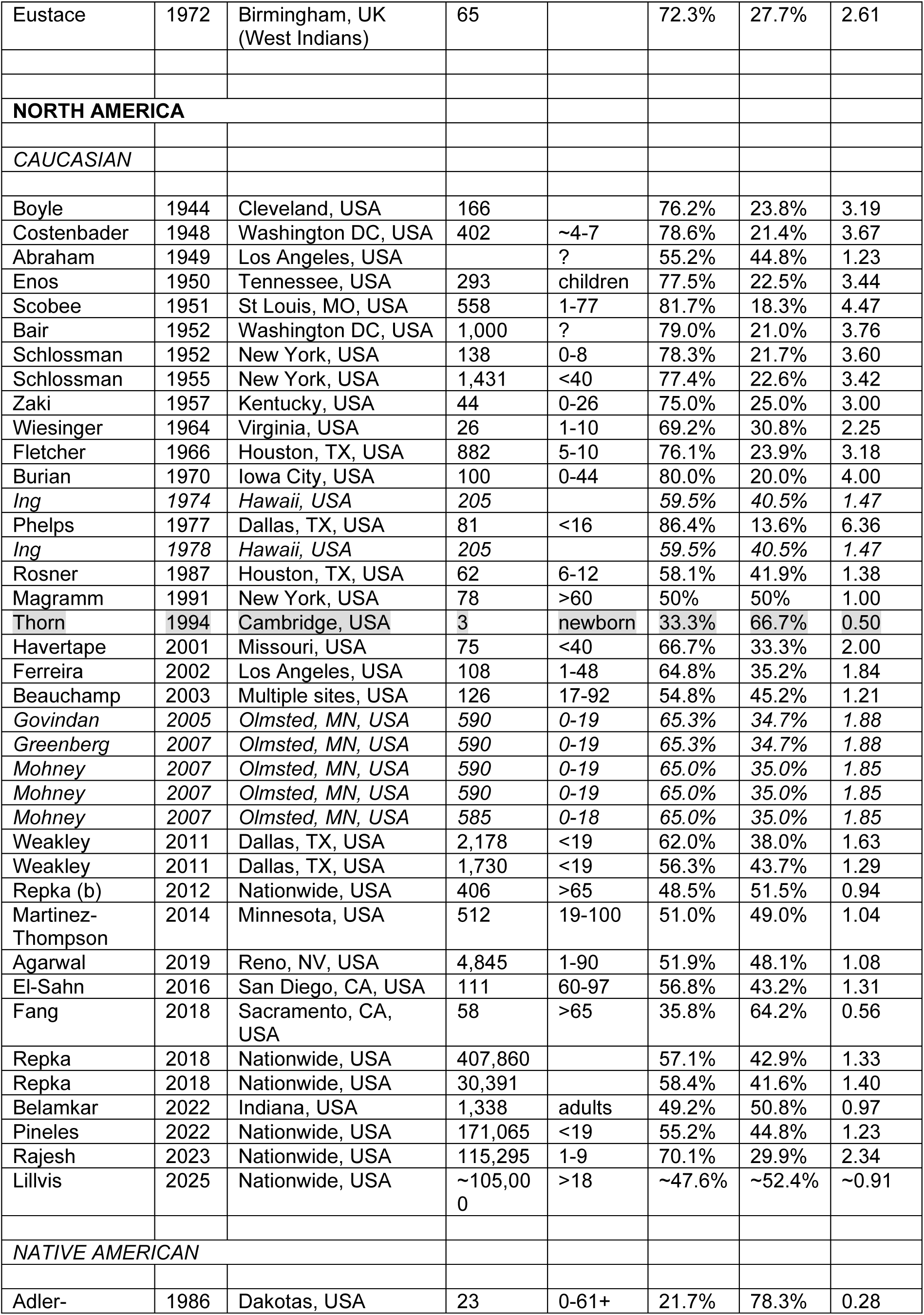
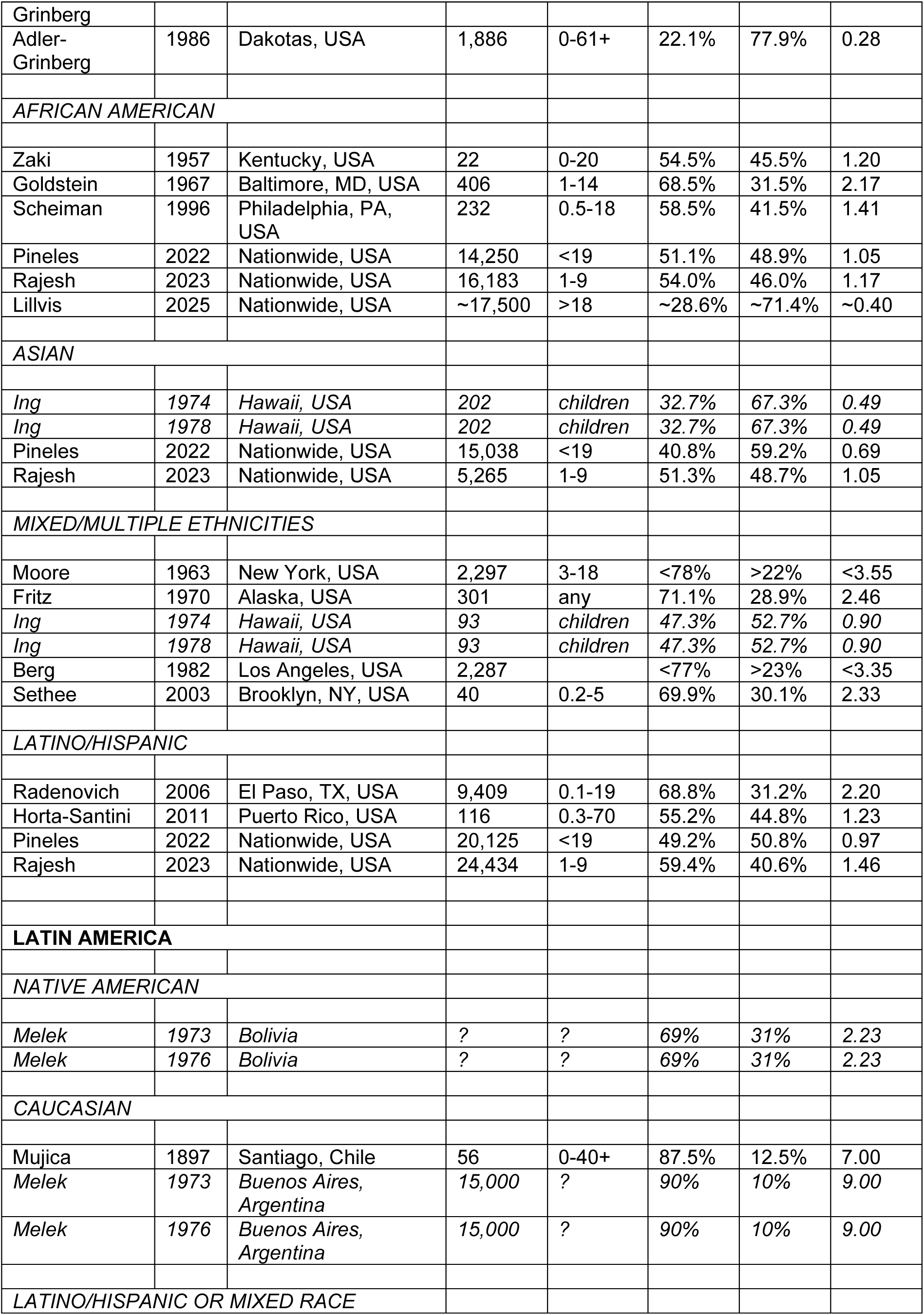
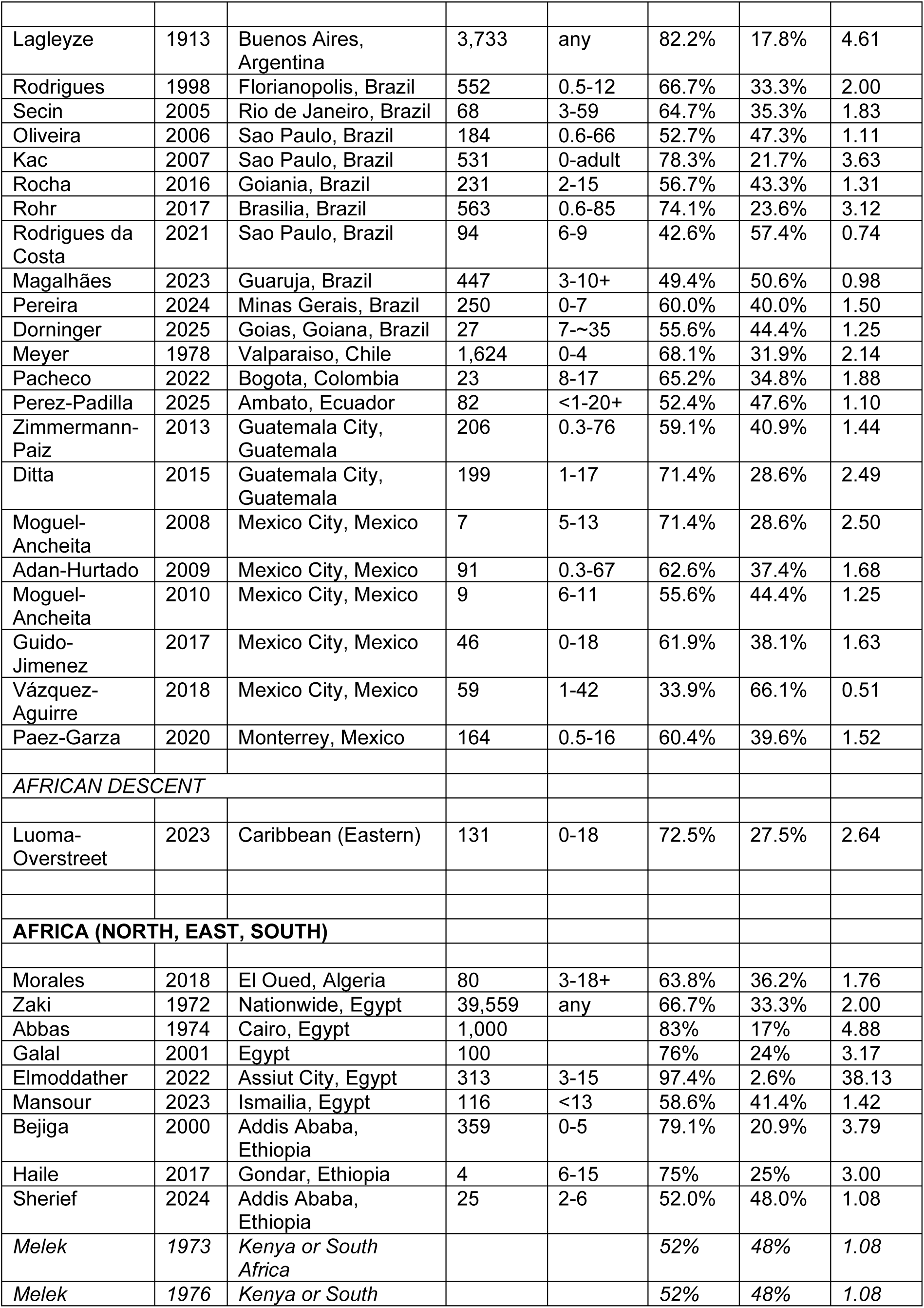
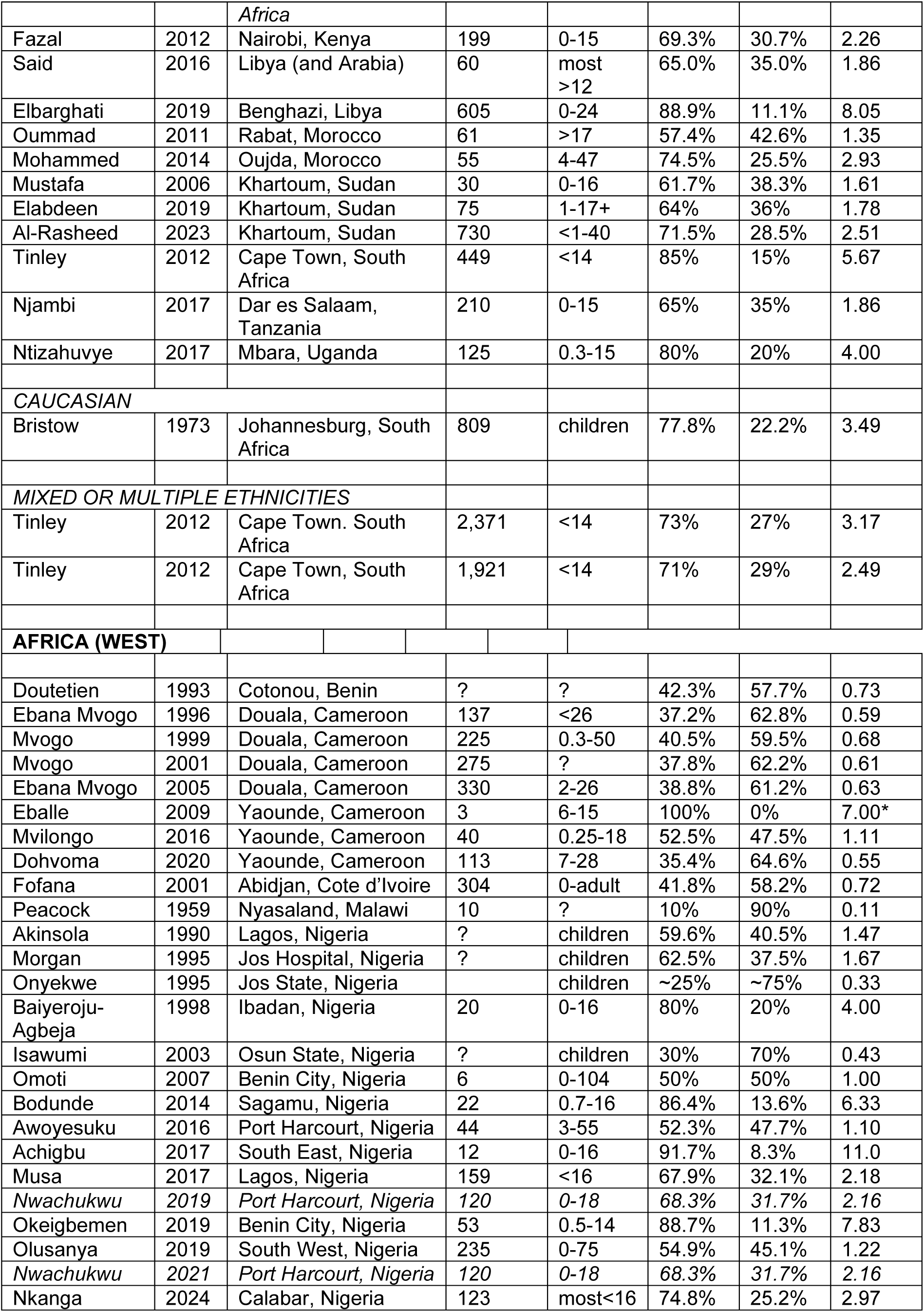
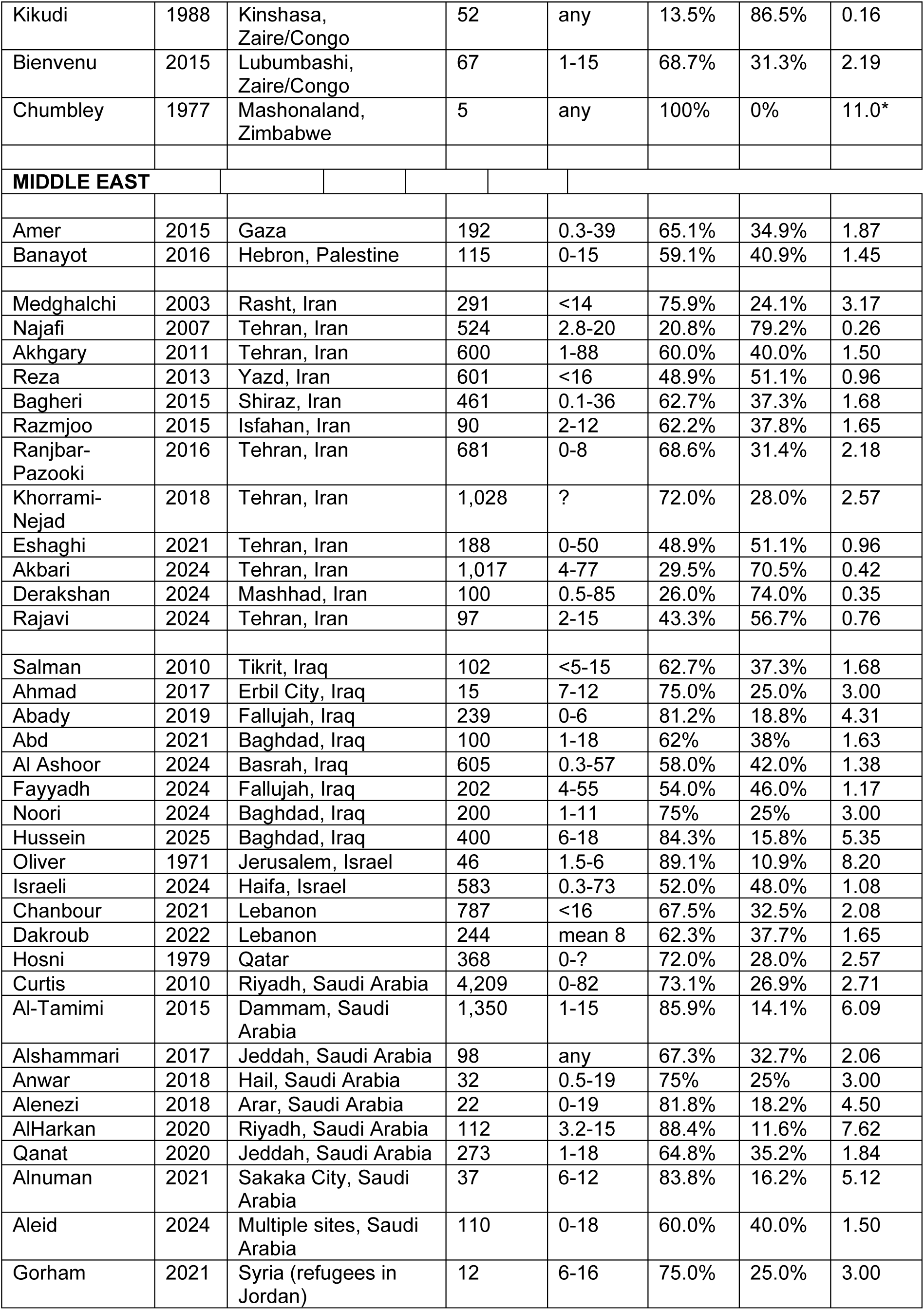
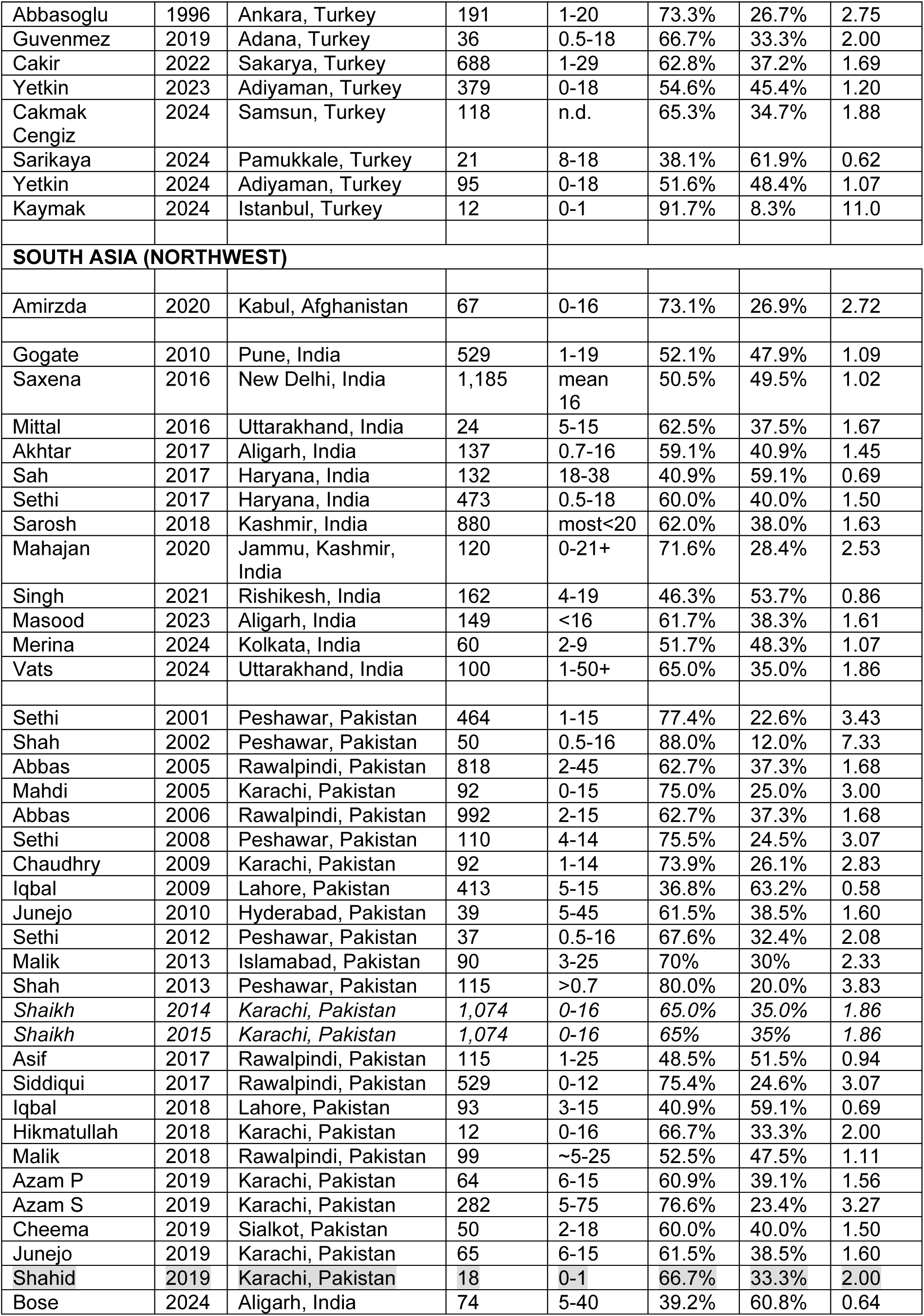
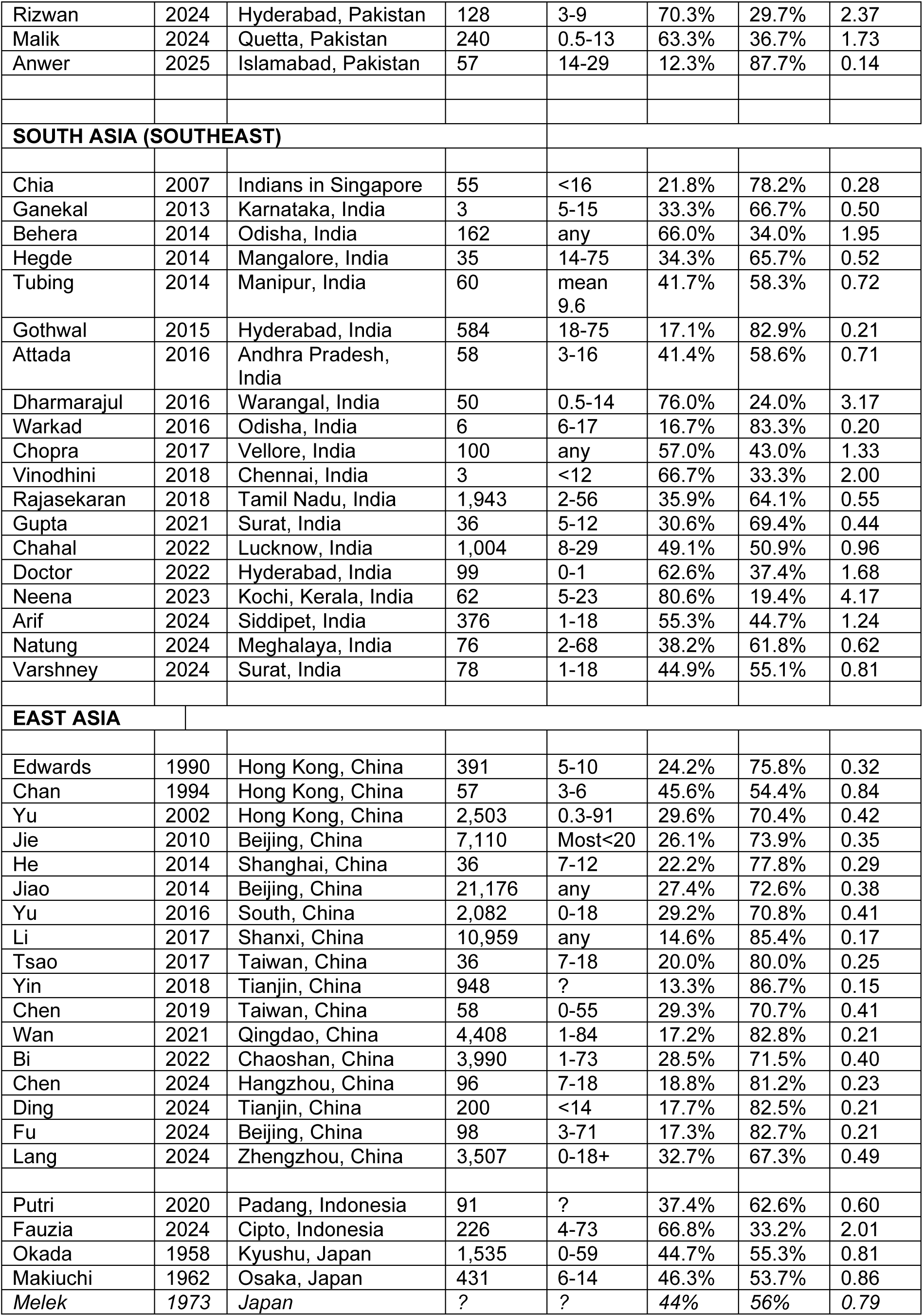
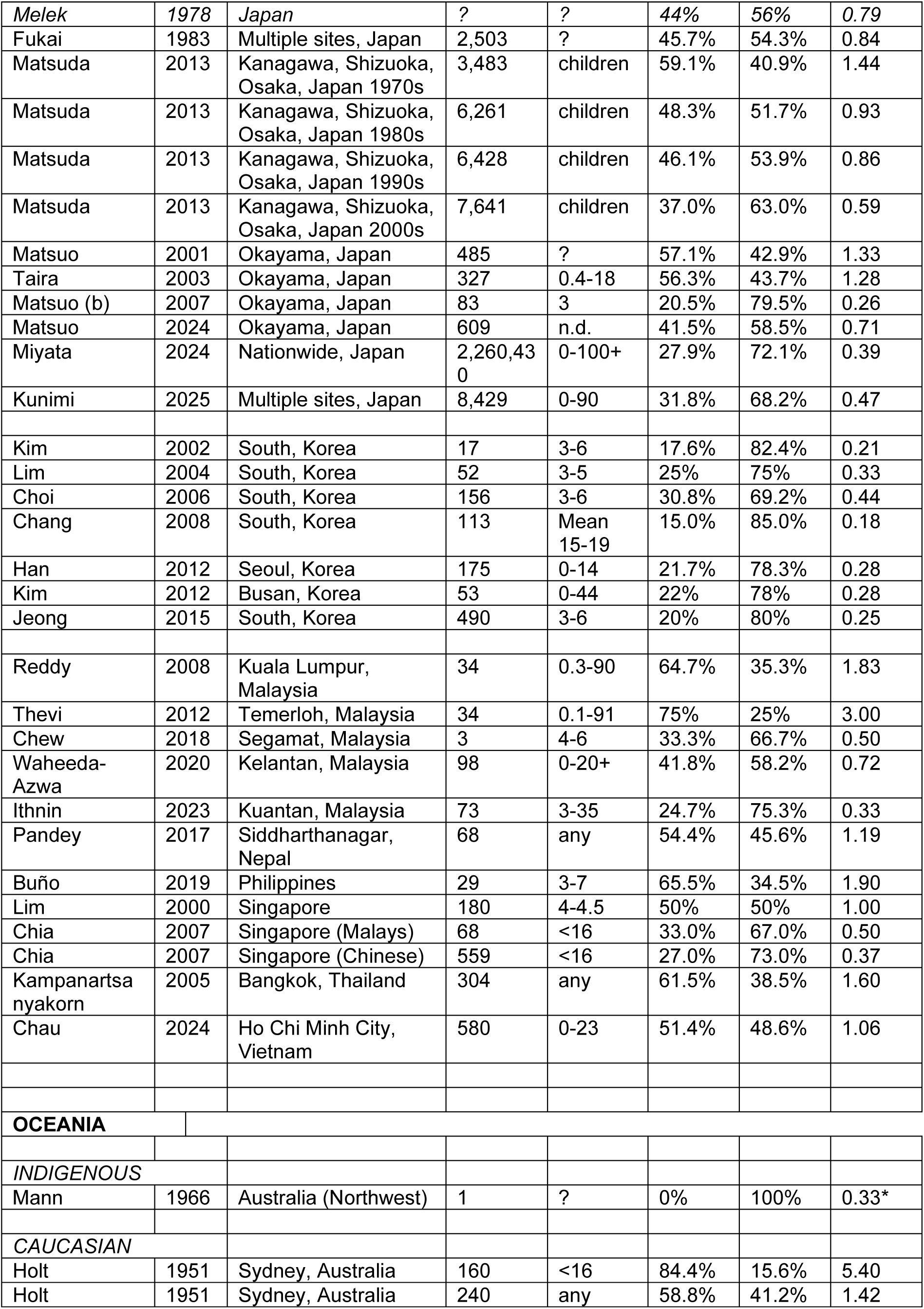
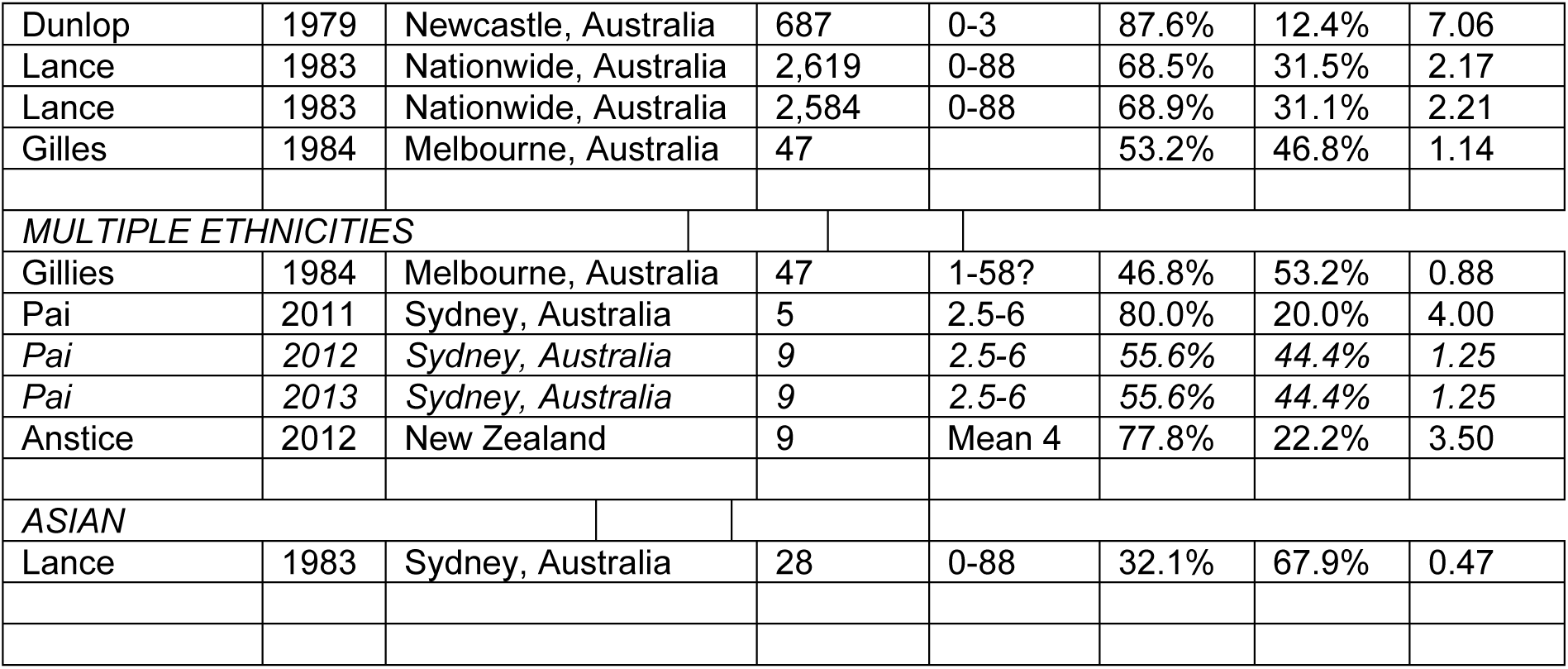
Esotropia/exotropia (ET/XT) ratio in different regions. Clinic-based, 374 studies with 374 cohorts. * 0.5 was added to each value for the ET/XT ratio when the value was 0, to avoid a zero in the denominator or enumerator, according to Sweeting et al., 2004.^758^ Abbreviations: yrs, years; ET, esotropia, XT, exotropia. Duplicate publication of the same cohort is indicated in *italics*.

**Supplemental Table S3.** Analysis of Heterogeneity. Esotropia.

Esotropia
| Population | n | $I^2$ | $I^2$ 95% CI | $\tau^2$ | $\tau^2$ 95% CI |
| --- | --- | --- | --- | --- | --- |
| Overall | 292 | 96.7% | [96.4%; 96.9%] | 0.9984 | [0.8631; 1.2992] |
| Caucasian | 89 | 97.7% |  | 0.3492 |  |
| Native American | 10 | 60.0% |  | 0.3661 |  |
| African American | 9 | 86.2% |  | 0.1491 |  |
| Latino/Hispanic | 13 | 90.4% |  | 0.2699 |  |
| Eskimo/Inuit | 2 | 72.7% |  | 0.2167 |  |
| Africa East, North | 17 | 88.6% |  | 0.9394 |  |
| Africa West | 18 | 59.7% |  | 0.4045 |  |
| Middle East | 18 | 91.5% |  | 0.5861 |  |
| Persian | 18 | 53.7% |  | 0.0784 |  |
| South Asia NW | 9 | 84.4% |  | 0.3002 |  |
| South Asia SE | 16 | 89.6% |  | 0.8190 |  |
| East Asia | 62 | 86.9% |  | 0.3105 |  |
| Oceania | 11 | 71.2% |  | 1.0001 |  |

CI, confidence interval; NW, Northwest; SE, Southeast. The total number of studies exceeds the number given in Fig. 2, because some studies report on cohorts of different ethnicities,<sup>147,170,255,510,545,D</sup> while others do not distinguish multiple ethnicities<sup>657,I</sup> and therefore are not included in this analysis.

Exotropia
| Population | n | $I^2$ | $I^2$ 95% CI | $\tau^2$ | $\tau^2$ 95% CI |
| --- | --- | --- | --- | --- | --- |
| Overall | 292 | 98.5% | [98.5%; 98.6%] | 0.9867 | [0.8545; 1.2818] |
| Caucasian | 89 | 96.4% |  | 0.6141 |  |
| Native American | 10 | 89.7% |  | 1.0098 |  |
| African American | 9 | 72.6% |  | 0.1060 |  |
| Latino/Hispanic | 13 | 98.6% |  | 0.8246 |  |
| Eskimo/Inuit | 2 | 66.0% |  | 0.1612 |  |
| Africa East, North | 17 | 79.3% |  | 0.7261 |  |
| Africa West | 18 | 85.9% |  | 0.7137 |  |
| Middle East | 18 | 96.0% |  | 0.8349 |  |
| Persian | 18 | 95.7% |  | 1.0365 |  |
| South Asia NW | 9 | 89.6% |  | 0.8338 |  |
| South Asia SE | 16 | 96.3% |  | 1.6493 |  |
| East Asia | 62 | 99.1% |  | 0.8482 |  |
| Oceania | 11 | 86.1% |  | 1.3948 |  |

CI, confidence interval; NW, Northwest; SE, Southeast. The total number of studies exceeds the number given in Fig. 2, because some studies report on cohorts of different ethnicities,<sup>147,170,255,510,545,D</sup> while others do not distinguish multiple ethnicities<sup>657,I</sup> and therefore are not included in this analysis.

## References

1. Abady NH, Al-jumaili AA, Fayyadh RA. Association between strabismus and refractive errors among preschool children in Fallujah, Iraq. Indian J Public Health Res Dev. 2019; 10(5):508–513.

2. Abah ER, Oladigbolu KK, Samaila E, Gani-Ikilama A. Ocular disorders in children in Zaria children’s school. Niger J Clin Pract. 2011; 14(4):473–476. doi: 10.4103/1119-3077.91759

3. Abbas S, Said A, Riad S. Incidence of the various types of squint: 1000 cases examined in the squint clinic Kasr El Aini Hospital. Bull Ophthalmol Soc Egypt. 1974; 67:311–314.

4. Abbas M, ur Rahman H, Butt IA, Ghani N. Prevalence and mode of presentation of vertical deviations in squint patients. Rawal Med J. 2005; 30:79–81.

5. Abbas M, Qureshi N, Ishaq N, Chaudhry MM, Ismail MAH. Refractive error study in strabismic children: Results from Al-Shifa Trust, Rawalpindi, Pakistan. Med Forum Monthly. 2006; 17(10):13–17.

6. Abbasoglu OE, Sener EC, Sanac AS. Factors influencing the successful outcome and response in strabismus surgery. Eye (Lond). 1996; 10(Pt 3):315–320. doi: 10.1038/eye.1996.66

7. Abd ZA, Salman HA. Demographic Data of 100 Strabismus Cases. Ann Trop Med Pub Health 2021; 24(6):463–468.

8. Abd FN, Fezea JF, Ahmed NS. The prevalence of amblyopia and heterotropia in school children aged 6 - 12 years old in Karbala, Iraq. Int J Pharm Pharmaceut Res. 2022; 26(1):225–233.

9. Abdelrahman A, Abdellah M, Alsamman A, Radwan G. The prevalence of strabismus in children at school age in Sohaf City. Egypt J Clin Ophthalmol 2020; 3(1):11–17.

10. Abdi S, Lennerstrand G, Pansell T, Rydberg A. Orthoptic findings and asthenopia in a population of Swedish schoolchildren aged 6 to 16 years. Strabismus. 2008; 16(2):47–55. doi: 10.1080/09273970802020243

11. Abdi Ahmed Z, Alrasheed SH, Alghamdi W. Prevalence of refractive error and visual impairment among school-age children of Hargesia, Somaliland, Somalia. East Mediterr Health J. 2020; 26(11):1362–1370. doi: 10.26719/emhj.20.077

12. Abdul-Kabir M, Abdul-Sadik A, Ansah DO and Ofosu-Koranteng L. Prevalence of anisometropia, strabismus and amblyopia among first year optometry students in Kwame Nkrumah University of Science and Technology, Ghana. Mathews J Ophtalmol. 2017; 2(2):018.

13. Abdiyeva Y. The level of prevalence and risk factors of strabismus among children and adolescents, depending on the type of settlements in the Ganja-Gazakh economic district (Azerbaijan). Ophthalmol Reports. 2021;14(4):45–53. Russian

14. Abelsdorff G. [Über Augenbefunde bei Malayen, Mongolen und Negern.] Bericht 27. Vers Ophthalmol Gesellsch. (Heidelberg) 1898; Bergmann, Wiesbaden 27:263–271. German

15. Abiose A, Bhar IS, Allanson MA. The ocular health status of postprimary school children in Kaduna, Nigeria: report of a survey. J Pediatr Ophthalmol Strabismus. 1980; 17(5):337–340. doi: 10.3928/0191-3913-19800901-17

16. Abolfotouh MA, Badawi I, Faheem Y. Prevalence of amblyopia among schoolboys in Abha city, Asir Region, Saudi Arabia. J Egypt Public Health Assoc. 1994; 69(1-2):19–30.

17. Abraham SV. A new classification of nonparalytic strabismus. Am J Ophthalmol. 1949; 32(1):93–98. doi: 10.1016/0002-9394(49)91113-7

18. Abrahamsson M, Fabian G, Sjöstrand J. Refraction changes in children developing convergent or divergent strabismus. Brit J Ophthalmol. 1992; 76(12):723–727. doi: 10.1136/bjo.76.12.723

19. Abrahamsson M, Magnusson G, Sjöstrand J. Inheritance of strabismus and the gain of using heredity to determine populations at risk of developing strabismus. Acta Ophthalmol Scand. 1999; 77(6):653–657. doi: 10.1034/j.1600-0420.1999.770609.x

20. Achigbu EO, Oguego NC, Achigbu K. Spectrum of eye disorders seen in a pediatric eye clinic South East Nigeria. Niger J Surg. 2017; 23(2):125–129. doi: 10.4103/njs.NJS_37_16

21. Adan-Hurtado EE, Arroyo-Yllanes ME. [Frecuencia de los diferentes tipos de estrabismo.] Rev Mex Oftalmol. 2009; 83(6):340–348. Spanish

22. Adejumo OO, Olusanya BA, Ajayi BG. Ocular Disorders among Preschool Children in Southwest Nigeria. Middle East Afr J Ophthalmol. 2021;28(1):23–28. doi: 10.4103/meajo.MEAJO_191_19

23. Adelstein AM, Scully J. Epidemiological aspects of squints. Br Med J. 1967; 3(5561):334-338. doi: 10.1136/bmj.3.5561.334

24. Adler H. [Beobachtungen und Bemerkungen uber das Sehen der Taubstummen.] Klin Monatsbl Augenheilk. 1876; 14:65–96. German

25. Adler-Grinberg D. Need for eye and vision care in an underserved population: refractive errors and other ocular anomalies in the Sioux. Am J Optom Physiol Opt. 1986; 63(7):553–558. doi: 10.1097/00006324-198607000-00009

26. Agarwal LP, Prakash P, Mahur A, Pathak MM. Visual defects in school children. Orient Arch Ophthal. 1966; 4:1–8.

27. Agarwal AB, Christensen AJ, Feng CY, Wen D, Johnson LA, von Bartheld CS. Expression of schizophrenia biomarkers in extraocular muscles from patients with strabismus: an explanation for the link between exotropia and schizophrenia? PeerJ. 2017; 5:e4214. doi: 10.7717/peerj.4214

28. Agarwal AB, Cassinelli K, Johnson LA, et al. Seasonality of births in horizontal strabismus: comparison with birth seasonality in schizophrenia and other disease conditions. J Dev Orig Health Dis. 2019; 10(6):636–644. doi: 10.1017/S2040174419000102

29. Agrawal D, Sahu A, Agrawal D. Prevalence of ocular morbidities among school children in Raipur district, India. Indian J Ophthalmol. 2020; 68(2):340–344. doi: 10.4103/ijo.IJO_1454_19

30. Ahmad MA, Al-Khayat ZA, Shakir NF. Prevalence of refractive errors and other ocular disorders among students of the primary schools in urban of the Erbil city. Polytechnic J. 2017; 7:1-10.

31. Ahmadi H, Shams PN, Davies NP, Joshi N, Kelly MH. Age-related changes in the normal sagittal relationship between globe and orbit. J Plast Reconstr Aesthet Surg. 2007; 60(3):246–250. doi: 10.1016/j.bjps.2006.07.001

32. Aichmair H, Grossmann W, Aichmair M, et al. [Randomized field study of the etiology of strabismus concomitans]. Wien Klin Wochenschr. 1992; 104(19):600–606. German

33. Ajaiyeoba AI, Isawumi MA, Adeoye AO, Oluleye TS. Prevalence and causes of blindness and visual impairment among school children in south-western Nigeria. Int Ophthalmol. 2005; 26(4-5):121–125. doi: 10.1007/s10792-005-4836-4

34. Ajaiyeoba AI, Isawumi MA, Adeoye AO, Oluleye TS. Prevalence and causes of eye diseases amongst students in south-western Nigeria. Ann Afr Med. 2006; 5:197–203.

35. Akarkar SO, Naik PG, Cacodcar JA. Prevalence and distribution of ocular morbidities among primary school children in Goa. J Clin Ophthalmol Res. 2019; 7:61–64.

36. Akbari MR, Alghurab A, Khorrami-Nejad M, Azizi E, Masoomian B. Sensory exotropia versus sensory esotropia: A comparative clinical features study. J Optom. 2024; 17(3):100516. doi: 10.1016/j.optom.2024.100516.

37. Akhgary M, Ghassemi-Broumand M, Amiri MA, Seyed MT. Prevalence of strabismic binocular anomalies, amblyopia and anisometropia. Rehabilitation Faculty of Shahid Beheshti Medical University. J Optom. 2011; 4(3):110–114. doi: 10.1016/S1888-4296(11)70050-4

38. Akhtar N, Gupta S. Prevalence of types of strabismus in pediatric patients in a tertiary centre of North India. J Dent Med Sci. 2017; 16(6):61–64.

39. Akinci A, Oner O, Bozkurt OH, Guven A, Degerliyurt A, Munir K. Refractive errors and ocular findings in children with intellectual disability: a controlled study. J AAPOS. 2008; 12(5):477–481. doi: 10.1016/j.jaapos.2008.04.009

40. Akowuah PK, Adade S, Nartey A, et al. Strabismus and amblyopia in Africa - a systematic review and meta-analysis. Strabismus. 2023; 31(1):31–44. doi: 10.1080/09273972.2022.2157023

41. Akpe BA, Dawodu OA, Abadom EG. Prevalence and pattern of strabismus in primary school pupils in Benin City, Nigeria. Nigerian J Ophthalmol. 2014; 22:38-43.

42. Alabi AS, Aribaba OT, Alabi AO, Ilo O, Onakoya AO, Akinsola FB. Visual impairment and ocular morbidities among schoolchildren in Southwest, Nigeria. Niger Postgrad Med J. 2018; 54:25(3):166-171. doi: 10.4103/npmj.npmj_85_18

43. Al Ashoor MA, Al Taha HM. Prevalence of Strabismus among Patients Attending Basrah Teaching Hospital, Basrah, Iraq. Al-Anbar Med J. 2025; 21(1):43–48. doi: 10.33091/amj.2024.153108.1901

44. Aleid KM, Alhawsawi KI, Abutalib YB, et al. Cross-sectional Study on Strabismus Prevalence and Risk Factors in Saudi Arabian Children. J Adv Trends Med Res. 2024; 1(2):519–525. doi: 10.4103/ATMR.ATMR_52_24

45. Alenezi HM, El-Fetoh NMA, Alruwaili AS, et al. Squint in children and adolescents, Arar, Northern Saudi Arabia. Egypt J Hosp Med. 2018; 70(2):298–302.

46. AlHarkan DH. Profile of Children with Strabismus Managed without Surgery at a Tertiary Eye Hospital of Central Saudi Arabia. Majmaah J Heal Sci. 2020; 8(1): 1–8. doi:10.5455/mjhs.2020.01.002

47. Allison CL. Proportion of refractive errors in a Polish immigrant population in Chicago. Optom Vis Sci. 2010; 87(8):588–592. doi: 10.1097/OPX.0b013e3181e61beb

48. Alnuman RWS, Alhablani FSN, Alruwaili EMA, et al. Prevalence of squint in primary school children and its associated sociodemographic factors in Sakaka City, Aljouf Region, Saudi Arabia. Int J Med Devel Countr. 2021; 5(4):1085–1091.

49. Alrasheed SH, Naidoo KS, Clarke-Farr PC. Prevalence of visual impairment and refractive error in school-aged children in South Darfur State of Sudan. Afr Vision Eye Health. 2016; 75(1),a355.

50. Al-Rasheed SH. Clinical Characteristics of Horizontal Strabismus in Sudanese Patients. Pak J Ophthalmol. 2023; 39(3):202–207.

51. Alshammari M, Alhibshi N, Almusallam A, Badawood E, Abdulwassi H. Risk factors for developing different subtypes of strabismus in a Saudi population. Int J Med Health Res. 2017; 3(11):116–120.

52. Al-Tamimi ER, Shakeel A, Yassin SA, Ali SI, Khan UA. A clinic-based study of refractive errors, strabismus, and amblyopia in pediatric age-group. J Family Community Med. 2015; 22(3):158–162. doi: 10.4103/2230-8229.163031

53. Amer A. Relative Prevalence of Various Types of Strabismus in Patients Attending NGO’s Medical Centers in Gaza Strip. Sci J Public Health. Special Issue: Health Behavior and Public Health. 2015; 3(1-1):1–5.

54. Amirzda SM. Prevalence of horizontal strabismus in pediatric patients at University Eye Hospital. J Opht Res Rev Rep. 2020; 1(1):1-2.

55. Anker S, Atkinson J, Braddick O, et al. Identification of infants with significant refractive error and strabismus in a population screening program using noncycloplegic videorefraction and orthoptic examination. Invest Ophthalmol Vis Sci. 2003; 44(2):497–504. doi: 10.1167/iovs.02-0070

56. Anstice N, Spink J, Abdul-Rahman A. Review of preschool vision screening referrals in South Auckland, New Zealand. Clin Exp Optom. 2012; 95(4):442–448. doi: 10.1111/j.1444-0938.2012.00713.x

57. Anwar AA, Albalawi AMA, Alharbi AAH, et al. Pattern of strabismus in children and adolescents in Hail, KSA. J Health Med Nursing. 2018; 54:28–33.

58. Anwer N, Anwer A, Akhtar R, Ahmed S. Frequency of Types of Squint and Gender Distribution Presenting to a Tertiary Care Hospital in Islamabad. Al-Shifa J Ophthalmol. 2025; 21(1):17–22.

59. Archer SM, Sondhi N, Helveston EM. Strabismus in infancy. Ophthalmology. 1989; 96(1):133-137. doi: 10.1016/s0161-6420(89)32932-0

60. Arif NM, Nathani AA. Assessment and Management of Strabismus Among Paediatric at Tertiary Care Teaching Hospitals. Res J Med Sci. 2024; 18:444–448. doi: 10.36478/makrjms.2024.7.444.448

61. Aring E, Grönlund MA, Andersson S, Hård AL, Ygge J, Hellström A. Strabismus and binocular functions in a sample of Swedish children aged 4-15 years. Strabismus. 2005; 13(2):55–61. doi: 10.1080/09273970590922664

62. Arora A, Williams B, Arora AK, McNamara R, Yates J, Fielder A. Decreasing strabismus surgery. Brit J Ophthalmol. 2005; 89(4):409-412. doi: 10.1136/bjo.2004.053678

63. Asaduzzaman M. Pattern of Ocular Morbidity among Children Referred Ophthalmology Dept. at Mugda Medical College Hospital, Mugda, Dhaka, Bangladesh. J Med Sci. 2024; 4(3):66-89.

64. Asif M, Habiba U, Khan MW, Kawish AB, Rehman A-u. Frequency of major types of manifest strabismus among patients of age group 1 to 25 years presented to Benazir Bhutto Hospital Rawalpindi. Pak J Med Biol Sci. 2017; 1:30-34.

65. Aslaksen AK, Vikesdal GH, Voie MT, Rowlands M, Skranes J, Haugen OH. Visual function in Norwegian children aged 5-13 years with prenatal exposure to opioid maintenance therapy: A case-control study. Acta Ophthalmol. 2024; 102(4):409–420. doi: 10.1111/aos.15764

66. Aslan L, Aslankurt M, Aksoy A, Altun H. Preventable visual impairment in children with nonprofound intellectual disability. Eur J Ophthalmol. 2013; 23(6):870–875. doi: 10.5301/ejo.5000304

67. Athanasiov PA, Prabhakaran VC, Selva D. Non-traumatic enophthalmos: a review. Acta Ophthalmol. 2008; 86(4):356–364. doi: 10.1111/j.1755-3768.2007.01152.x

68. Atiya A, Hussaindeen JR, Kasturirangan S, Ramasubramanian S, Swathi K, Swaminathan M. Frequency of undetected binocular vision anomalies among ophthalmology trainees. J Optom. 2020; 13(3):185–190. doi: 10.1016/j.optom.2020.01.003

69. Attada TR, Deepika M, Laxmi S. Strabismus in paediatric age (3-16 year): a clinical study. Int J Res Med Sci. 2016; 4(6):1903–1909.

70. Atuanya GN, Suzan OI. Visual Problems Affecting Reading and Learning Among Primary School Children in Benin City, Nigeria. Optom Vis Perform. 2024; 12(2):91–98.

71. Auzemery A, Andriamanamihaja R, Boisier P. [A survey of the prevalence and causes of eye disorders in primary school children in Antananarivo]. Sante. 1995; 5(3):163–166. French

72. Awan AA, Aftab M, Arif S. Refractive error and squint in primary school children. J Ayub Med Coll Abbotabad Pak. 1995; 7(2):26–27.

73. Awan HR, Ihsan T. Prevalence of visual impairment and eye diseases in Afghan refugees in Pakistan. E Mediterr Health J. 1998; 4(3):560–566.

74. Awan AR, Jamshed J, Khan MM, Latif Z. Prevalence and causes of visual impairment and blindness among school children in Muzaffarabad, Pakistan. Int J Sci Rep. 2018; 4(4):93–98.

75. Awoyesuku EA, Fiebai B, Onua AA. Pattern of strabismus in a tertiary hospital in Nigeria: a six-year review. Port Harcourt Med J. 2016; 10:14–17.

76. Ayanniyi AA, Mahmoud AO, Olatunji FO. Causes and prevalence of ocular morbidity among primary school children in Ilorin, Nigeria. Niger J Clin Pract. 2010; 13(3):248–253.

77. Azam P, Nausheen N, Fahim MF. Prevalence of strabismus and its type in pediatric age group 6-15 years in a tertiary eye care hospital, Karachi. Biom Biostat Int J. 2019; 8(1):24–28.

78. Azam S, Priyanka, Qasim M, Frequency of neurogenic strabismus in Al-Ibrahim eye hospital, Karachi. Pak. J. Ophthalmol. 2019; 35 (2): 122–126.

79. Azonobi IR, Olatunji FO, Addo J. Prevalence and pattern of strabismus in Ilorin. West Afr J Med. 2009a; 28(4):253–256.

80. Azonobi IR, Adido J, Olatunji FO, Bello A, Mahmoud AO. Risk factors of strabismus in Southwestern Nigeria. Pak J Ophthalmol. 2009b; 25(3):129–134.

81. Badr I, Qureshi I. Ocular status of school children in the town of Al-Majma’ah, Central Province, Saudi Arabia. Saudi Med J. 1981; 2:221–224.

82. Bagheri M, Farvardin M, Saadat M. A study of consanguineous marriage as a risk factor for developing comitant strabismus. J Community Genet. 2015; 6(2):177–180. doi: 10.1007/s12687-015-0213-9

83. Bagheri M, Farvardin M. The clinical effect of surgical timing in infantile exotropia. J AAPOS. 2018; 22(3):167–169.e1. doi: 10.1016/j.jaapos.2017.12.004

84. Baharian S, Barakatt M, Gignoux CR, et al. The great migration and African-American genomic diversity. PLoS Genet. 2016; 27;12(5):e1006059. doi: 10.1371/journal.pgen.1006059

85. Bair DR. Intermittent exotropia; diagnosis and incidence. Am Orthopt J. 1952; 2:12–17.

86. Baiyeroju-Agbeja AM, Owoeye JFA. Strabismus in children in Ibadan. Niger J Ophthalmol. 1998; 6(1):31–33.

87. Balduzzi S, Rücker G, Schwarzer G. How to perform a meta-analysis with R: a practical tutorial. Evid Based Ment Health. 2019; 22(4):153–160. doi: 10.1136/ebmental-2019-300117

88. Banayot RG. A retrospective analysis of eye conditions among children attending St. John Eye Hospital, Hebron, Palestine. BMC Res Notes. 2016; 9:202. doi: 10.1186/s13104-016-2011-9

89. Bardisi WM, Bin Sadiq BM. Vision screening of preschool children in Jeddah, Saudi Arabia. Saudi Med J. 2002; 23:445–449.

90. Barrie TS. The incidence of squint at various ages. Trans Ophthalmol Soc UK. 1922; 42:273–278.

91. Basu A, Mukherjee N, Roy S, et al. Ethnic India: a genomic view, with special reference to peopling and structure. Genome Res. 2003; 13(10):2277–2290. doi: 10.1101/gr.1413403

92. Beardsell R, Clarke S, Hill M. Outcome of occlusion treatment for amblyopia. J Pediatr Ophthalmol Strabismus. 1999; 36(1):19–24. doi: 10.3928/0191-3913-19990101-05

93. Beauchamp GR, Black BC, Coats DK, et al. The management of strabismus in adults--I. Clinical characteristics and treatment. J AAPOS. 2003; 7(4):233–240. doi: 10.1016/s1091-8531(03)00112-5

94. Bedi R, Bedi DK, Dudule CN, Nizamuddin, Keswani M, Saxena P. Prevalence of ocular morbidities among school children: a comparative study across social categories in Ajmer City. Natl J Commun Med. 2016; 7(3):184–188.

95. Beer SMC, Scarpi MJ, Minello A. [Ocular findings in children between 0 and 6 years of age, residing in the city of Sao Caetano do Sul, SP]. Arq Bras Oftalmol. 2003; 66(6):839–845. Portuguese

96. Behera S, Dutta BK, Chowdhury RK, Sar M. A clinic-anatomical study of strabismus in a tertiary care hospital. IOSR J Dent Med Sci. 2014; 13(1):32–35.

97. Beiguelman B. A survey on genetical and anthropological traits among Japanese immigrants in Brazil. Z Morphol Anthropol. 1964; 55(1):46–59.

98. Bejiga A. Unilateral blindness and low vision due to strabismic amblyopia. Ethiop J Health Dev. 2000; 14(1):109–112.

99. Belfort Mattos R. [Estudo oftalmologico dos Indios do Medio Xingu.] Arq Bras Oftalmol. 1970; 33(20):33–43. Portuguese

100. Berg PH. Effect of light intensity on the prevalence of exotropia in strabismic populations. Brit Orthopt J. 1982; 39:55–56.

101. Betrán AP, Ye J, Moller AB, Zhang J, Gülmezoglu AM, Torloni MR. The increasing trend in Caesarean section rates: Global, regional and national estimates: 1990-2014. PLoS One. 2016; 11(2):e0148343. doi: 10.1371/journal.pone.0148343

102. Bi Y, Yam JC, Lin S. A retrospective study of strabismus surgery in a tertiary eye hospital in the Chaoshan area in China from 2014 to 2020. BMC Ophthalmol. 2022; 22(1):246. doi: 10.1186/s12886-022-02479-8

103. Bienvenu YA, Angel MN, Sebastien MM, et al. [Study of strabismus in children 0-15 years followed in Lubumbashi, Democratic Republic of Congo: Analysis of epidemiological and clinical aspects]. Pan Afr Med J. 2015; 22:66. French. doi: 10.11604/pamj.2015.22.66.5324

104. Blake CR, Lai WW, Edward DP. Racial and ethnic differences in ocular anatomy. Int Ophthalmol Clin. 2003; 43(4):9–25. doi: 10.1097/00004397-200343040-00004.

105. Blum HL, Peters HB, Bettman JW. Vision Screening for Elementary Schools. The Orinda Study. Univ of California Press, Berkeley and Los Angeles, 1959, 146 pages.

106. Bodunde OT, Onabolu OO, Fakolujo VO. Pattern of squint presentations in children in a tertiary institution in Western Nigeria. J Dent Med Sci. 2014; 13(5):29–31.

107. Bose A, Amitava AK, Gupta Y, et al. To compare horizontal strabismus deviation as assessed from photographs with that in the strabismus clinic using the prism bar. Indian J Ophthalmol. 2024; 72(8):1199–1203. doi: 10.4103/IJO.IJO_409_23

108. Boyle MO. The frequency of squint. Am J Ophthalmol. 1944; 27(12):1413–1416.

109. Bristow JH, Douglas WHG, McCartney D, Swartz J, Armstrong EB, Schwyzer E. Divergent squint in Caucasoids. Cases seen in a paediatric hospital outpatients department. S Afr Med J. 1973; 47(40):1925–1926.

110. Brown S, Jones D. A survey of the incidence of defective vision and strabismus in kindergarten age children – Sydney, 1976. Aust Orthopt J. 1976; 15:24-28.

111. Browning SR, Browning BL, Zhou Y, Tucci S, Akey JM. Analysis of Human Sequence Data Reveals Two Pulses of Archaic Denisovan Admixture. Cell. 2018; 173(1):53–61.e9. doi: 10.1016/j.cell.2018.02.031

112. Bruce A, Hurst M, Abbott H, Harrison H. The incidence of refractive error and anomalies of binocular vision in infants. Br Orthoptic J. 1991; 48:32-35.

113. Bruce A, Santorelli G. Prevalence and risk factors of strabismus in a UK multi-ethnic birth cohort. Strabismus. 2016; 24(4):153–160. doi: 10.1080/09273972.2016.1242639

114. Bruns HD. Certain Eye Affections in the Negro (Discussion). Trans Am Ophthalmol Soc. 1910; 12(Pt 2):462–463.

115. Bryc K, Auton A, Nelson MR, et al. Genome-wide patterns of population structure and admixture in West Africans and African Americans. Proc Natl Acad Sci U S A. 2010; 107(2):786–791. doi: 10.1073/pnas.0909559107

116. Bryc K, Durand EY, Macpherson JM, Reich D, Mountain JL. The genetic ancestry of African Americans, Latinos, and European Americans across the United States. Am J Hum Genet. 2015; 96(1):37–53. doi: 10.1016/j.ajhg.2014.11.010

117. Buffenn AN. The impact of strabismus on psychosocial health and quality of life: a systematic review. Surv Ophthalmol. 2021; 66(6):1051–1064. doi: 10.1016/j.survophthal.2021.03.005

118. Buño II B, Monzon-Pajarillo AK. Effects of visual impairment on quality of life in children aged 3-7 years. Philip J Ophthalmol. 2019; 44:14-18.

119. Burian HM, Luke NE. Sensory retinal relationships in 100 consecutive cases of heterotropia. A comparative clinical study. Arch Ophthalmol. 1970; 84(1):16–20. doi: 10.1001/archopht.1970.00990040018005

120. Burnett SM. International Medical Congress: Ametropia. Am J Ophthalmol. 1887; 4:299.

121. Busby GB, Band G, Si Le Q, et al. Admixture into and within sub-Saharan Africa. Elife. 2016; 5. pii: e15266. doi: 10.7554/eLife.15266

122. Caca I, Cingu AK, Sahin A, et al. Amblyopia and refractive errors among school-aged children with low socioeconomic status in southeastern Turkey. J Pediatr Ophthalmol Strabismus. 2013; 50(1):37–43. doi: 10.3928/01913913-20120804-02

123. Çakır B, Aksoy NÖ, Bursalı Ö, Özmen S. Non-ocular risk factors in Turkish children with strabismus and amblyopia. Turk J Pediatr. 2022;64(2):341–349. doi: 10.24953/turkjped.2021.204

124. ÇAKMAK CENGİZ E, NİYAZ ŞAHİN L. Ezotropya ve Ekzotropya Tanılı Hastalarda Hidrosefali, Serebral Palsi, Epilepsi Sıklığının Saptanması ve Cerrahi Dışı Tedavi Sonrası Klinik Özelliklerin Karşılaştırılması. MN Opthalmol. 2024;31(4):197-202. Turkish

125. Cameron J. The interorbital width. A new cranial dimension. Its significance in modern and fossil man and in lower mammals. Am J Biol Anthropol. 1931; 15(3):509–519. doi: 10.1002/ajpa.1330150308

126. Carney CV, Lysons DA, Tapley JV. Is the incidence of constant esotropia in childhood reducing? Eye (Lond). 1995; 9 (Pt 6 Su):40–41.

127. Casari G. [Corrispondenza retinica e terapia ortottica dello strabismo concomitante.] Rass ital ottal. 1950; 19:101–139. Italian

128. Cass EE. Divergent Strabismus. Brit J Ophthalmol. 1937; 21(10):538-559. doi: 10.1136/bjo.21.10.538

129. Cass E. A decade of northern ophthalmology. Can J Ophthalmol. 1973; 8(2):210–217.

130. Casson RJ, Kahawita S, Kong A, Muecke J, Sisaleumsak S, Visonnavong V. Exceptionally low prevalence of refractive error and visual impairment in schoolchildren from Lao People’s Democratic Republic. Ophthalmology. 2012; 119(10):2021–2027. doi: 10.1016/j.ophtha.2012.03.049

131. Catford JG, Absolon MJ, Millo A. Squints: a sideways look. In: Progress in Child Health. Ed: Macfarlane JA. Edinburgh, Churchill-Livingstone, 1984. pp. 38-50.

132. Çelikay O, Çalışkan S, Acar M, et al. [The results of an eye health screening in a primary school children.] Turkiye Klinikleri J Ophthalmol. 2016; 25. 10.5336/ophthal.2015-48981. Turkish

133. Chahal V, Singh V, Singh J, Kaur G, Srivastava RM, Agrawal S. Profile of orthoptic clinic patients at a tertiary care Government Medical University in North India: A 6-year review. J Clin Ophthalmol Res. 2022; 10:27–32. doi: 10.4103/jcor.jcor_123_21

134. Chan KY, Zhao FF, Meng S, et al. Prevalence of schizophrenia in China between 1990 and 2010. J Glob Health. 2015; 5(1):010410. doi: 10.7189/jogh.05.010410

135. Chan OY, Edwards M. Refraction referral criteria for Hong Kong Chinese preschool children. Ophthalmic Physiol Opt. 1994; 14(3):249–256. doi: 10.1111/j.1475-1313.1994.tb00005.x

136. Chanbour H, Bsat A, Chanbour W, Cherfan C. Geographic Variation in Strabismus Pattern Among Pediatric Age Group in Lebanon: A Single-Centre Five-Year Observational Study. Cureus. 2021; 13(6):e15957. doi: 10.7759/cureus.15957

137. Chang YS, Kim SY. [Clinical study of A-V pattern strabismus in Korea.] J Korean Ophthalmol Soc. 2008; 49(12):1974–1980. Korean

138. Chau VTB, Trí TQ, Hai LT, Nhung DT. RESEARCH ON THE CORRELATION BETWEEN REFRACTIVE ERROR AND STRABISMUS AT HCM EYE HOSPITAL. Vietnam J Commun Med. 2024; 65(Special Issue 8):134–142. Vietnamese

139. Chaudhry TA, Khan A, Khan MB, Ahmad K. Gender differences and delay in presentation of childhood squint. J Pak Med Assoc. 2009; 59(4):229–231.

140. Chauhan H, Dohare S, Khandekar J, Pradhan SK, Jain R, Mahajan H. Prevalence of ocular morbidity in 5 to 14 years children in an urban resettlement colony of New Delhi, India. Indian J Pub Health Res Dev. 2013; 4(4):25–28.

141. Cheema MN, Anwar S, Hussain A, Bangash MT, Afzal MFB, Shamas F. Frequency of Esotropia Among the Patients Presented with Ocular Misalignments between the Age Group of 2-18 Years in Ophthalmology Outdoor Department at Islam Teaching Hospital. Pak J Med Health Sci. 2019; 13(4):830–833.

142. Chen X, Fu Z, Yu J, et al. Prevalence of amblyopia and strabismus in Eastern China: results from screening of preschool children aged 36-72 months. Brit J Ophthalmol. 2016; 100(4):515–519. doi: 10.1136/bjophthalmol-2015-306999

143. Chen YW, Lin SA, Lin PW, Huang HM. The difference of surgical outcomes between manifest exotropia and esotropia. Int Ophthalmol. 2019; 39(7):1427–1436. doi: 10.1007/s10792-018-0956-5

144. Chen D, Li R, Li X, et al. Prevalence, incidence and risk factors of strabismus in a Chinese population-based cohort of preschool children: the Nanjing Eye Study. Br J Ophthalmol. 2021; 105(9):1203–1210. doi: 10.1136/bjophthalmol-2020-316807

145. Chen F, Lou L, Yu X, et al. Evaluation and application of a Chinese version symptom questionnaire for visual dysfunctions (CSQVD) in school-age children. Adv Ophthalmol Pract Res. 2024; 4(3):134–141. doi: 10.1016/j.aopr.2024.05.001

146. Cheung MW, Ho RC, Lim Y, Mak A. Conducting a meta-analysis: basics and good practices. Int J Rheum Dis. 2012; 15(2):129–135. doi: 10.1111/j.1756-185X.2012.01712.x

147. Chew E, Remaley NA, Tamboli A, Zhao J, Podgor MJ, Klebanoff M. Risk factors for esotropia and exotropia. Arch Ophthalmol. 1994; 112(10):1349–1355. doi: 10.1001/archopht.1994.01090220099030

148. Chew FLM, Thavaratnam LK, Shukor INC, et al. Visual impairment and amblyopia in Malaysian pre-school children - The SEGPAEDS study. Med J Malaysia. 2018; 73(1):25–30.

149. Chia A, Roy L, Seenyen L. Comitant horizontal strabismus: an Asian perspective. Brit J Ophthalmol. 2007; 91(10):1337–1340. doi: 10.1136/bjo.2007.116905

150. Chia A, Dirani M, Chan YH, et al. Prevalence of amblyopia and strabismus in young Singaporean Chinese children. Invest Ophthalmol Vis Sci. 2010; 51:3411–3417. doi: 10.1167/iovs.09-4461

151. Chia A, Lin X, Dirani M, et al. Risk factors for strabismus and amblyopia in young Singapore Chinese children. Ophthalmic Epidemiol. 2013; 20(3):138–147. doi: 10.3109/09286586.2013.767354

152. Chiarelli CA, Chris AP. Visual status of First Nations children: The Sagamok First Nation Vision Care Project. Can J Optom. 2013; 75(4):39–46.

153. Chibueze OS, Oluwatosin OJ, Ebele EG. PREVALENCE AND PATTERN OF STRABISMUS AMONG SECONDARY SCHOOL CHILDREN IN OGUN STATE, NIGERIA. Int J Med Sci Appl Biosci. 2020; 5(3):14–22.

154. Chimonidou E, Palimeris G, Koliopoulos J, Velissaropoulos P. Family distribution of concomitant squint in Greece. Br J Ophthalmol. 1977; 61(1):27–29. doi: 10.1136/bjo.61.1.27

155. Choi TB, Lee DA, Oelrich FO, Amponash D, Bateman JB, Christensen RE. A retrospective study of eye disease among first grade children in Los Angeles. J Am Optom Assoc. 1995; 66(8):484–488.

156. Choi KW, Koo BS, Lee HY. Preschool vision screening in Korea: Results in 2003. J Korean Ophthalmol Soc. 2006; 47:112–120. Korean

157. Chopra V, Balasubramanian P. Clinical study of concomitant squint. J Evid Based Med Healthc. 2017; 4(54):3294–3297.

158. Chou MR, Malik AN, Suleman M, Gray M, Yeates D, Goldacre MJ. Time trends over five decades, and recent geographical variation, in rates of childhood squint surgery in England. Brit J Ophthalmol. 2013; 97(6):746–751. doi: 10.1136/bjophthalmol-2012-302932

159. Christian LW, Nandakumar K, Hrynchak PK, Irving EL. Visual and binocular status in elementary school children with a reading problem. J Optom. 2018; 11(3):160–166. doi: 10.1016/j.optom.2017.09.003

160. Chumbley LC. Impressions of eye diseases among Rhodesian Blacks in Mashonaland. S Afr Med J. 1977; 52(8):316–318.

161. Clarke LC, Fraser SG. Hospital Episode Statistics and trends in ophthalmic surgery 1998-2004. BMC Ophthalmol. 2006; 6:37. doi: 10.1186/1471-2415-6-37

162. Cohen J. Screening results on the ocular status of 651 prekindergarteners. Am J Optom Physiol Opt. 1981; 58(8):648–662.

163. Cohn H. [Untersuchungen der Augen von 10060 Schulkindern nebst Vorschlaegen zur Verbesserung der den Augen nachtheiligen Schuleinrichtungen.] Leipzig, Verlag von Friederich Fleischer, 1867. https://archive.org/details/b2163693x

164. Cohn H. [Ueber Vererbung und Behandlung des Einwaerts-Schielens.] Berl Klin Wochenschr. 1904; 41:1047–1051. German

165. Colas Q, Capsec J, Arsène S, Pisella PJ, Grammatico-Guillon L, Khanna RK. Strabismus outcomes after surgery: the nationwide SOS France study. Graefes Arch Clin Exp Ophthalmol. 2022; 260(6):2037–2043. doi: 10.1007/s00417-021-05541-1

166. Collins SD. Strabismus and defective color sense among school children. Pub Health Rep. (Washington) 1925; 40:1515–1523.

167. Coon CS. The Origin of Races. Oxford, England: Knopf, 1962.

168. Costa MN, Jose NK, Filho NM, et al. [Estudo da incidência de ambliopia, estrabismo e anisometropia em pré-escolares.] Arq Bras Oftal. 1979; 42(6):249–252. Portuguese

169. Costenbader F, Bair D, McPhail A. Vision in strabismus; a preliminary report. Arch Ophthal. 1948; 40(4):438–453. doi: 10.1001/archopht.1948.00900030450005

170. Cotter SA, Varma R, Tarczy-Hornoch K, et al. Risk factors associated with childhood strabismus: the multi-ethnic pediatric eye disease and Baltimore pediatric eye disease studies. Ophthalmology. 2011; 118(11):2251–2261. doi: 10.1016/j.ophtha.2011.06.032

171. Couto Jr AS, Pinto GR, de Oliveira DA, et al. [Prevalence of the ametropias and eye diseases in preschool and school children of Alto da Boa Vista favelas, Rio de Janeiro, Brazil]. Rev Bras Oftalmol. 2007; 66(5):304–308. Portuguese

172. Couto Jr AS, Jardim JL, de Oliveira DA, Gobetti TC, Portes AJF, Neurauter R. [Eye diseases in preschool and school children in the city of Duque de Caxias, Rio de Janeiro, Brazil]. Rev Bras Oftalmol. 2010; 69(1):7–11. Portuguese

173. Crone RA, Velzeboer CM. Statistics on strabismus in the Amsterdam youth; researches into the origin of strabismus. AMA Arch Ophthalmol. 1956; 55(4):455–470. doi: 10.1001/archopht.1956.00930030459002

174. Curtis TH, McClatchey M, Wheeler DT. Epidemiology of surgical strabismus in Saudi Arabia. Ophthalmic Epidemiol. 2010; 17(5):307–314. doi: 10.3109/09286586.2010.508351

175. Dakroub M, El Hadi D, El Moussawi Z, Ibrahim P, Al-Haddad C. Characteristics and long-term surgical outcomes of horizontal strabismus. Int Ophthalmol. 2022; 42(5):1639–1649. doi: 10.1007/s10792-021-02159-4

176. Dandona R, Dandona L, Srinivas M, et al. Refractive error in children in a rural population in India. Invest Ophthalmol Vis Sci. 2002; 43(3):615–622.

177. Datta A, Choudhury N, Kundu K. An epidemiological study of ocular condition among primary school children of Calcutta Corporation. Indian J Ophthalmol. 1983; 31(5):505–510.

178. Dawson E, Bentley C, Lee J. Squint surgery in the over sixties. Strabismus. 2001; 9(4):217–220. doi: 10.1076/stra.9.4.217.690

179. De Decker W, Tessmer J. [Occurrence of squint and efficiency of treatment in Schleswig-Holstein]. Klin Monbl Augenheilkd. 1973; 162(1):34–42. German

180. de Figueiredo RM, dos Santos EC, de Jesus IA, Castilho RM, dos Santos EV. [Proposal for a procedure for the detection of ophthalmic disorders in school children.] Rev Saúde Públ. 1993; 27(3):204-209. Portuguese. doi: 10.1590/s0034-89101993000300008

181. Delgado Molina A, Zato Gomez de Liano MA. [Estereogramas y prevencion visual escolar.] Arch Soc Esp Oftal. 1991; 60:445–450. Spanish

182. Delgado-Rodríguez M, Llorca J. Bias. J Epidemiol Community Health. 2004; 58(8):635-641. doi: 10.1136/jech.2003.008466

183. Deplanche M. Deux annees de pratique ophtalmologique en Cote d’Ivoire. Boll Soc Path Exot. 1931; 24(5):406–415. French

184. Derakhshan A, Sabermoghaddam A, Abdolahian M, et al. The Strabismus Prevalence in a Random Sample of Iran’s Northeastern Population. Pak J Ophthalmol. 2024; 40(1):34–39. doi: 10.36351/pjo.v40i1.1688

185. DerSimonian R, Laird N. Meta-analysis in clinical trials. Control Clin Trials. 1986; 7:177–188. doi: 10.1016/0197-2456(86)90046-2.

186. De Sousa RLF, Funayama BS, Cataneo L, Padovini CR, Schellini SA. [Comparison between visual acuity and photoscreening used like visual screening methods for scholar aged children.] Rev Bras Oftalmol. 2012; 71(6):358–363. Portuguese

187. De Vries B, Houtman WA, Van Bochove J. Prevalence of squint and amblyopia on the island of Curacao. Brit Orthopt J. 1984; 41:53-57.

188. Dey AK, Nath AB. Prevalence of ocular morbidities among school children in a rural block of Cachar, Assam. J Evolut Med Dent Sci. 2017; 6(55):4124–4127.

189. Dharmaraju B, Vijayasree S, Mythili K. Study of aetiological factors contributing to paediatric strabismus. J Evid Based Med Healthc. 2016; 3(65):3520–3523.

190. Ding J, Wang W, Zhang W, Li Y. Parenting Psychological Distress and Its Association with Demographic and Clinical Characteristics in Strabismus Children: A Cross-Sectional Study. Authorea. [Preprint] posted January 04, 2024. doi: 10.22541/au.170438407.75629142/v1

191. Ditta LC, Pereiras LA, Graves ET, et al. Establishing a surgical outreach program in the developing world: pediatric strabismus surgery in Guatemala City, Guatemala. J AAPOS. 2015; 19(6):526–530. doi: 10.1016/j.jaapos.2015.09.005

192. Doctor MB, Sachadeva V, Kekunnaya R. Profile of infantile strabismus at a tertiary eye care center in India. Indian J Ophthalmol. 2022; 70:3056–3060. doi: 10.4103/ijo.IJO_543_22

193. Doden W. [A study of strabismus]. Buch Augenarzt. 1961; 38:69–115. German

194. Dohvoma VA, Ebana Mvogo SR, Mvilongo CT, Epee E, Ebana Mvogo C. Strabisme de l’enfance négligé: aspects épidémiologiques, cliniques et thérapeutiques [Neglected childhood strabismus: Epidemiological, clinical and therapeutic aspects]. J Fr Ophtalmol. 2020; 43(8):774–778. French. doi: 10.1016/j.jfo.2019.11.040

195. Dombrow M, Engel HM. Rates of strabismus surgery in the United States: implications for manpower needs in pediatric ophthalmology. J AAPOS. 2007; 11(4):330–335. doi: 10.1016/j.jaapos.2007.05.010

196. Donnelly UM, Stewart NM, Hollinger M. Prevalence and outcomes of childhood visual disorders. Ophthalmic Epidemiol. 2005; 12(4):243–250. doi: 10.1080/09286580590967772

197. Donnelly UM. Horizontal strabismus worldwide - what are the risk factors? Ophthalmic Epidemiol. 2012; 19(3):117–119. doi: 10.3109/09286586.2012.681002

198. Dorninger IE, Isaac DL, Taleb AC, Carvalho KM, Ávila MP. Quality of life in patients with strabismus: Assessment of the preoperative and postoperative psychosocial and functional aspects. Arq Bras Oftalmol. 2025; 88(3):e2024–0118. Portuguese. doi: 10.5935/0004-2749.2024-0118

199. Doutetien C, Oussa G, Babagbetou M, Bassabi SK. Epidemiologie du strabisme au C.N.H.U de Cotonou. Premieres journees Senegalo-Saoudiennes d’ophthalmologie, Dakar, decembre 1993. French

200. Downing AH. Ocular defects in sixty thousand selectees. Arch Ophthalmol. 1945; 33(2):137–143.

201. Duelund N, Nisted I, Jørgensen ME, Heegaard S, Jensen H. Visual profiling and vision screening of preschool children in Greenland. Int J Circumpolar Health. 2025; 84(1):2489194. doi: 10.1080/22423982.2025.2489194

202. Dunlop P, Dunlop D. Infant strabismus: A 25-year review of 750 cases. Med J Aust. 1979; 1:111–113. doi: 10.5694/j.1326-5377.1979.tb112041.x

203. Ebana Mvogo C, Bella-Hiag AL, Epesse M. [Strabismus in Cameroon]. J Fr Ophtalmol. 1996; 19(11):705-709. French

204. Ebana Mvogo C, Ellong A, Owona D, Luma H, Bella LA. [Amblyopia and strabismus in our environment]. Bull Soc Belge Ophtalmol. 2005; 297:39–44. French

205. Eballe AO, Bella LA, Owono D, Mbome S, Mvogo CE. [Eye disease in children aged 6 to 15 years: a hospital-based study in Yaounde]. Sante. 2009; 19(2):61–66. French. doi: 10.1684/san.2009.0158

206. Edwards M. The refractive status of Hong Kong Chinese infants. Ophthalmic Physiol Opt. 1991; 11(4):297–303.

207. Edwards M, Yap M. Visual problems in Hong Kong primary school children. Clin Exp Optometr. 1990; 73(2):58–63.

208. Egger M, Smith GD, Schneider M, Minder C. Bias in meta-analysis detected by a simple, graphical test. Brit Med J. 1997a; 315:629–634. doi: 10.1136/bmj.315.7109.629

209. Egger M, Smith GD, Phillips AN. Meta-analysis: principles and procedures. Brit Med J. 1997b; 315:1533-1537. doi: 10.1136/bmj.315.7121.1533

210. Ehrlich JR, Anthopolos R, Tootoo J, et al. Assessing geographic variation in strabismus diagnosis among children enrolled in Medicaid. Ophthalmology. 2016; 123(9):2013–2022. doi: 10.1016/j.ophtha.2016.05.023

211. Ekdawi NS, Nusz KJ, Diehl NN, Mohney BG. The development of myopia among children with intermittent exotropia. Am J Ophthalmol. 2010; 149(3):503–507. doi: 10.1016/j.ajo.2009.10.009

212. Elabdeen RHZ, Ibrahim SM. Clinical study of vertical strabismus among patients attending squint clinic- Makkah Eye Hospital – Khartoum. Int Res Med Health Sci. 2019; 2(5):1–19.

213. Elbarghathi AM, Elbarghathi MF, Abdullah RM. Epidemiology of Strabismus among Patients Visiting Squint Clinic in Benghazi. Appl Cell Biol. 2019; 7(1):6–11.

214. El-Bayoumy BM, Saad A, Choudhury AH. Prevalence of refractive error and low vision among schoolchildren in Cairo. East Mediterr Health J. 2007; 13(3):575–579.

215. Elmoddather M. Prevalence of strabismus and its types in pediatric population and the outcomes of different treatment modalities: A 3-year prospective study in a referral eye center in Upper Egypt. Egypt J Hosp Med. 2022; 86:826–830.

216. Elliott R. An ophthalmic survey of the Tokelau Islands. Trans Ophthalmol Soc NZ. 1965; 17:25-35.

217. Elsahn, M. International Vision Screening: Results from Alexandria, Egypt. Curr Ophthalmol Rep. 2014; 2:137–141. doi: 10.1007/s40135-014-0055-3

218. El-Sahn MF, Granet DB, Marvasti A, Roa A, Kinori M. Strabismus in adults older than 60 years. J Pediatr Ophthalmol Strabismus. 2016; 53(6):365–368. doi: 10.3928/01913913-20160722-02

219. Emmrich K. [Results of eye examinations in the schools]. Dtsch Gesundheitsw. 1954; 9(20):622–626. German

220. Engle EC. Genetic basis of congenital strabismus. Arch Ophthalmol. 2007; 125(2):189–195. doi: 10.1001/archopht.125.2.189

221. Enos MV. Anomalous correspondence. Am J Ophthalmol. 1950; 33(12):1906-1914. doi: 10.1016/0002-9394(50)90011-0

222. Eshaghi M, Arabi A, Banaie S, Shahraki T, Eshaghi S, Esfandiari H. Predictive factors of stereopsis outcomes following strabismus surgery. Ther Adv Ophthalmol. 2021; 13:25158414211003001. doi: 10.1177/25158414211003001

223. Etezad Razavi M, Sharifi M, Vahed E. Evaluation of visual problems in children with pervasive developmental disorder. Bina J Ophthalmol. 2012; 18(2):186–190.

224. Eustace P. Myopia and divergent squint in West Indian children. Brit J Ophthalmol. 1972; 56(7):559–564. doi: 10.1136/bjo.56.7.559

225. Ezinne NE, Mashige KP. Refractive error and visual impairment in primary school children in Onitsha, Anambra State, Nigeria. Afr Vision Eye Health. 2018; 77(1), a455.

226. Fabian G. [Augenaerztliche Reihenuntersuchung von 1,200 Kindern im 2. Lebensjahr.] Acta Ophthalmol. 1966; 44(3):473–479. German. doi: 10.1111/j.1755-3768.1966.tb08061.x

227. Fabian G, Wendell ME. Ophthalmologic and orthoptic examination of 1,200 children up to age 2. Am Orthopt J. 1974; 24:86–90.

228. Faghihi M, Ostadimoghaddam H, Yekta AA. Amblyopia and strabismus in Iranian schoolchildren, Mashhad. Strabismus. 2011; 19(4):147–152. doi: 10.3109/09273972.2011.622341

229. Faghihi M, Ostadi moghaddam H, Fatemi A, Heravian Shandiz J, Yekta A. Strabismus and amblyopia in schoolboys of Varamin, Iran, in 2010. Iranian J Ophthalmol. 2012; 24:32-38.

230. Fan DSP, Lai C, Lau HH, Cheung EY, Lam DS. Change in vision disorders among Hong Kong preschoolers in 10 years. Clin Exp Ophthalmol. 2011; 39(5):398–403. doi: 10.1111/j.1442-9071.2010.02470.x

231. Fang SY, Gandhi N, Satterfield D, O’Hara M. Strabismus surgery for Medicare-aged patients: more than a decade of insights. J AAPOS. 2018; 22(3):170–173. doi: 10.1016/j.jaapos.2017.12.017

232. Farjadian S, Sazzini M, Tofanelli S, et al. Discordant patterns of mtDNA and ethno-linguistic variation in 14 Iranian ethnic groups. Hum Hered. 2011; 72(2):73–84. doi: 10.1159/000330166

233. Fauzia FM, Bani AP. SURGICAL MANAGEMENT IN ESOTROPIA AT CIPTO MANGUNKUSUMO HOSPITAL: A 4-YEAR OBSERVATION ON CHARACTERISTIC AND RESULT. Ophthalmol Indones. 2024; 50(1):28–37.

234. Fayyadh RA, Abdulelah AB, Yosif WAS. The outcome of horizontal strabismus surgery in Anbar governorate in Iraq: an interventional study. J Emergency Med Trauma Acute Care. 2024; 2024(1):1–7.

235. Ferede AT, Alemu DS, Gudeta AD, Alemu HW, Melese MA. Visual Impairment among Primary School Children in Gondar Town, Northwest Ethiopia. J Ophthalmol. 2020; 2020:6934013. doi: 10.1155/2020/6934013

236. Ferguson JA, Goldacre MJ, Henderson J, Bron AJ. Ophthalmology in the Oxford region: analysis of time trends from linked statistics. Eye (Lond). 1991; 5 (Pt 3):379–384. doi: 10.1038/eye.1991.61

237. Fernandes M, Yang X, Li JY, Cheikh Ismail L. Smoking during pregnancy and vision difficulties in children: a systematic review. Acta Ophthalmol. 2015; 93(3):213–223. doi: 10.1111/aos.12627

238. Fernandes AG, Vianna RG, Gabriel DC, et al. Refractive error and ocular alignment in school-aged children from low-income areas of São Paulo, Brazil. BMC Ophthalmol. 2024; 24(1):452. doi: 10.1186/s12886-024-03710-4

239. Ferreira RC, Oelrich F, Bateman B. Genetic aspects of strabismus. Arc Bras Oftalmol. 2002; 65:171–175. doi: 10.1590/S0004-27492002000200004

240. Fiess A, Kölb-Keerl R, Schuster AK, et al. Prevalence and associated factors of strabismus in former preterm and full-term infants between 4 and 10 years of age. BMC Ophthalmol. 2017; 17(1):228. doi: 10.1186/s12886-017-0605-1

241. Fiess A, Elflein HM, Urschitz MS, et al. Prevalence of strabismus and its impact on vision-related quality of life: Results from the German population-based Gutenberg Health Study. Ophthalmology. 2020; 127(8):1113–1122. doi: 10.1016/j.ophtha.2020.02.026

242. Fikre B, Ferede AT, Tefera TK. Strabismus Prevalence and Associated Factors among Kindergarten School Children, Northwest Ethiopia. Int J Ophthalmol Vis Sci. 2022; 7(2):45–50.

243. Finlay R. Unexpected beneficial effects of measles immunisation. Number of squint operations has decreased. Brit Med J. 2000; 320(7239):939; author reply 939-940.

244. Fischbach LA, Lee DA, Englehardt RF, Wheeler N. The prevalence of ocular disorders among Hispanic and Caucasian children screened by the UCLA Mobile Eye Clinic. J Community Health. 1993; 18(4):201–211. doi: 10.1007/BF01324431

245. Fischer E. Die Rehobother Bastards und das Bastardierungsproblem beim Menschen. Jena: Gustav Fischer, 1913. 327 pp.

246. Flegontov P, Altınışık NE, Changmai P, et al. Palaeo-Eskimo genetic ancestry and the peopling of Chukotka and North America. Nature. 2019; 570(7760):236-240. doi: 10.1038/s41586-019-1251-y

247. Fletcher MC, Silverman SJ. Strabismus. I. A summary of 1,110 consecutive cases. Am J Ophthalmol. 1966; 61(1):86–94.

248. Fotouhi A, Hashemi H, Khabazkhoob M, Mohammad K. The prevalence of refractive errors among schoolchildren in Dezful, Iran. Br J Ophthalmol. 2007; 91(3):287–292. doi: 10.1136/bjo.2006.099937

249. Franceschetti A, Franceschetti AT, Hudson-Shaw S. [Detection of eye disorders in children of pre-school age]. Klin Monbl Augenheilkd. 1966; 149(5):657–662. German

250. Frandsen AD. Some results from a clinical-statistical survey on strabismus among Copenhagen children. Acta Ophthalmol (Copenh). 1958; 36(3):488–498. doi: 10.1111/j.1755-3768.1958.tb00825.x

251. Frandsen AD. Occurrence of squint: a clinical-statistical study on the prevalence of squint and associated signs in different groups and ages of the Danish population. Acta Ophthalmol Suppl. 1960; 62:9–157.

252. Freedman J. Survey of ocular disease among the Nama people of South West Africa. Brit J Ophthalmol. 1973; 57(9):681–687. doi: 10.1136/bjo.57.9.681

253. Freitas-da-Costa P, Falcão-Reis F, Magalhães A. Trends and patterns in pediatric ophthalmology and strabismus surgeries: a decade review from a leading Portuguese university hospital. Strabismus. 2024; 32(1):54–62. doi: 10.1080/09273972.2024.2317221

254. French AN, O’Donoghue L, Morgan IG, Saunders KJ, Mitchell P, Rose KA. Comparison of refraction and ocular biometry in European Caucasian children living in Northern Ireland and Sydney, Australia. Invest Ophthalmol Vis Sci. 2012; 53(7):4021–4031. doi: 10.1167/iovs.12-9556

255. Friedman DS, Repka MX, Katz J, et al. Prevalence of amblyopia and strabismus in white and African American children aged 6 through 71 months: The Baltimore Pediatric Eye Disease Study. Ophthalmology. 2009; 116:2128–2134. doi: 10.1016/j.ophtha.2009.04.034

256. Friedman Z, Neumann E, Hyams SW, Peleg B. Ophthalmic screening of 38,000 children, age 1 to 2 1/2 years, in child welfare clinics. J Pediatr Ophthalmol Strabismus. 1980; 17(4):261–267. doi: 10.3928/0191-3913-19800701-16

257. Friedmann L, Biedner B, David R, Sachs U. Screening for refractive errors, strabismus and other ocular anomalies from ages 6 months to 3 years. J Pediatr Ophthalmol Strabismus. 1980; 17(5):315–317. doi: 10.3928/0191-3913-19800901-11

258. Friedrich D, de Decker W. Prospective study of the development of strabismus during the first six months of life. In: Lenk-Schafer M (ed) Orthoptic Horizons: Transactions of the Sixth International Orthoptic Congress. Harrogate. British Orthoptic Society, London, 1987, pp 21-28.

259. Friendly DS. Preschool visual acuity screening tests. Trans Am Ophthalmol Soc. 1978; 76:383–480.

260. Fritz MH. The eye and adnexal disease of the Alaska native-1970. Alaska Med. 1970; 12(3):70–75.

261. Fu J, Li SM, Liu LR, et al. Prevalence of amblyopia and strabismus in a population of 7th-grade junior high school students in Central China: The Anyang Childhood Eye Study (ACES). Ophthalmic Epidemiol. 2014; 21(3):197–203. doi: 10.3109/09286586.2014.904371

262. Fu J, Wang Y, Liu J, Chen W, Jiang M. Repeatability and reproducibility of VR in automated measurement and diagnosis of strabismus. Res Square [Preprint] posted Feb 16, 2024 doi: 10.21203/rs.3.rs-3823812/v1

263. Fukai S, Kawase Y, Fukuda M, et al. [Incidence of esotropia and exotropia in Japan.] Jap Orthopt J. 1983; 11(1):29–35. Japanese

264. Galal AH; Nowier SR; Rizk A. Heritability of strabismus among Egyptian population. Ain-Shams Med J. 2001; 52 (1-2-3):229-239.

265. Galantuomo MS, Zucca I, Cuccu A, Fossarello M. Familial strabismus in Sardinia. Invest Ophthalmol Vis Sci. 2011; 52(14):6357–6357.

266. Gallo JE, Lennerstrand G. A population-based study of ocular abnormalities in premature children aged 5 to 10 years. Am J Ophthalmol. 1991; 111(5):539–547. doi: 10.1016/s0002-9394(14)73695-5

267. Ganekal S, Jhanji V, Liang Y, Dorairaj S. Prevalence and etiology of amblyopia in Southern India: results from screening of school children aged 5-15 years. Ophthalmic Epidemiol. 2013; 20(4):228–231. doi: 10.3109/09286586.2013.809772

268. Gansner J. [On the incidence of strabismic amblyopia. Statistical survey of the preschool children of an urban population]. Ophthalmologica. 1968; 155(3):234–244. German. doi: 10.1159/000305286

269. Ganz ML, Xuan Z, Hunter DG. Prevalence and correlates of children’s diagnosed eye and vision conditions. Ophthalmology. 2006; 113(12):2298–2306. doi: 10.1016/j.ophtha.2006.06.015

270. Garcia CA, de Sousa AB, Mendonca MB, Andrade LL, Orefice F. Prevalence of strabismus among students in Natal/RN – Brazil. Arq Bras Oftalmol. 2004; 67(5):791–794. doi: 10.1590/S0004-27492004000500018

271. Garvey KA, Dobson V, Messer DH, Miller JM, Harvey EM. Prevalence of strabismus among preschool, kindergarten, and first-grade Tohono O’odham children. Optometry. 2010; 81(4):194–199. doi: 10.1016/j.optm.2009.10.010

272. Germano RAS, Kawai RM, de Souza BL, Germano FAS, Germano CS, Germano JE. Frequency of ocular conditions in native Brazilians from Avai City, Sao Paulo state. Rev Bras Oftalmol. 2017; 76(5):227–231. doi: 10.5935/0034-7280.20170047

273. Gillan RU. An analysis of one hundred cases of strabismus treated orthoptically. Brit J Ophthalmol. 1945; 29(8):420–428. doi: 10.1136/bjo.29.8.420

274. Gillies WE, Hughes A. Results in 50 cases of strabismus after graduated surgery designed by A scan ultrasonography. Brit J Ophthalmol. 1984; 68:790–795. doi: 10.1136/bjo.68.11.790

275. Giorgis AT, Bejiga A. Prevalence of strabismus among pre-school children community in Butajira Town. Ethiop J Health Dev. 2001; 15(2):125–130.

276. Gittelman RM, Schraiber JG, Vernot B, Mikacenic C, Wurfel MM, Akey JM. Archaic hominin admixture facilitated adaptation to Out-of-Africa environments. Curr Biol. 2016; 26(24):3375–3382. doi: 10.1016/j.cub.2016.10.041

277. Gogate PM, Rishikeshi N, Taras S, Aghor M, Deshpande MD. Clinical audit of horizontal strabismus surgery in children in Maharashtra, India. Strabismus. 2010; 18(1):13–17. doi: 10.3109/09273970903567618

278. Goh PP, Abqariyah Y, Pokharel GP, Ellwein LB. Refractive error and visual impairment in school-age children in Gombak District, Malaysia. Ophthalmology. 2005; 112(4):678–685. doi: 10.1016/j.ophtha.2004.10.048

279. Goldstein H, Henderson M, Goldberg ID, Benitez E, Hawkins CM. Perinatal factors associated with strabismus in Negro children. Am J Public Health Nations Health. 1967; 57(2):217–228.

280. Gordon YJ, Mokete M. Screening of pre-school and school children for ocular anomalies in Lesotho. J Trop Med Hyg. 1982; 85(4):135–137.

281. Gorham JP, Behshad S, Weil NC. Comparison of Two Photoscreeners in a Population of Syrian Refugee Children. J Pediatr Ophthalmol Strabismus. 2021; 58(6):396–400. doi: 10.3928/01913913-20210428-01

282. Goseki T, Ishikawa H. The prevalence and types of strabismus, and average of stereopsis in Japanese adults. Jpn J Ophthalmol. 2017; 61(3):280–285. doi: 10.1007/s10384-017-0505-1

283. Gothwal VK, Bharani S, Kekunnaya R, et al. Measuring Health-Related Quality of Life in Strabismus: A Modification of the Adult Strabismus-20 (AS-20) Questionnaire Using Rasch Analysis. PLoS One. 2015; 10(5):e0127064. doi: 10.1371/journal.pone.0127064

284. Gouveia-Moraes F, Barros S, Vide Escada A, et al. Strabismus and Health Related Quality of Life in a pediatric Portuguese population. Strabismus. 2023; 31(4):262–270. doi: 10.1080/09273972.2023.2276138

285. Gover M, Yaukey JB. Physical impairments of members of low-income farm families; 11, 490 persons in 2,477 Farm Security Administration borrower families, 1940; extent of immunization against smallpox, diptheria, and typhoid fever. Public Health Rep. 1946; 61:97-109.

286. Govindan M, Mohney BG, Diehl NN, Burke JP. Incidence and types of childhood exotropia: a population-based study. Ophthalmology 2005; 112:104–108. doi: 10.1016/j.ophtha.2004.07.033

287. Graham PA. Epidemiology of Strabismus. Brit J Ophthalmol. 1974; 58(3):224-231. doi: 10.1136/bjo.58.3.224

288. Green AS, Green LD. Squint: when shall we operate? JAMA. 1921; 77(13):1003–1007.

289. Greenberg AE, Mohney BG, Diehl NN, Burke JP. Incidence and types of childhood esotropia: A population based study. Ophthalmology. 2007; 114(1):170–174. doi: 10.1016/j.ophtha.2006.05.072

290. Green-Simms AE, Mohney BG. Epidemiology of pediatric strabismus. In: Pediatric Ophthalmology, Neuro-Ophthalmology, Genetics. Lorenz B, Brodsky MC (eds), pages 1-9. Berlin, Springer, 2010.

291. Gregersen E. The polymorphous exo patient. Analysis of 231 successive cases. Acta Ophthalmol (Copenh). 1969; 47(3):579-590. doi: 10.1111/j.1755-3768.1969.tb08144.x

292. Grönlund MA, Aring E, Hellström A, Landgren M, Strömland K. Visual and ocular findings in children adopted from eastern Europe. Brit J Ophthalmol. 2004; 88(11):1362–1367. doi: 10.1136/bjo.2004.042085

293. Grönlund MA, Andersson S, Aring E, Hård AL, Hellström A. Ophthalmological findings in a sample of Swedish children aged 4-15 years. Acta Ophthalmol Scand. 2006; 84(2):169–176. doi: 10.1111/j.1600-0420.2005.00615.x

294. Guido Jiméneza MA, Pérez Pérez JF, Arroyo Yllanes ME. [Características clínicas del estrabismo en pacientes con catarata congénita.] Rev Mex Oftalmol. 2017; 91(3):122–126. Spanish. doi: 10.1016/j.mexoft.2016.04.011

295. Gummel K, Ygge J. Ophthalmologic findings in Russian children with fetal alcohol syndrome. Eur J Ophthalmol. 2013; 23(6):823–830. doi: 10.5301/ejo.5000296

296. Gupta M, Gupta Y. A survey on refractive error and strabismus among children in a school at Aligarh. Indian J Public Health. 2000; 44(3):90–93.

297. Gupta V, Nanda R. Prevalence of eye diseases in primary school students in Nagrota. J Med Sci Clin Res. 2017; 5(7):25156–25159. doi: 10.18535/jmscr/v5i7.139

298. Gupta M, Gupta BP, Chauhan A, Bhardwaj A. Ocular morbidity prevalence among school children in Shimla, Himachal, North India. Indian J Ophthalmol. 2009; 57(2):133–138. doi: 10.4103/0301-4738.45503

299. Gupta N, Arya SK, Walia D, Mallik A, Sood S. Ocular morbidity among school-going children in the Union Territory of Chandigarh. Int Ophthalmol. 2014; 34(2):251–257. doi: 10.1007/s10792-013-9825-4

300. Gupta PK, Caculo DU. The attitude towards strabismus and barriers for its treatment in parents from rural and urban areas. Indian J Clin Exp Ophthalmol. 2021; 7(1):54–61. doi: 10.18231/j.ijceo.2021.012

301. Gursoy H, Basmak H, Yaz Y, Colak E. Vision screening in children entering school: Eskisehir, Turkey. Ophthalmic Epidemiol. 2013; 20(4):232–238. doi: 10.3109/09286586.2013.808672

302. Guvenmez O, Kayiklik A. Strabismus in Pediatric Age: A Single Center Experience in Turkey. Ulutas Med J. 2019; 5(3):184–188. doi: 10.5455/umj.20191127100430

303. Haase W, Mühlig HP. [The incidence of squinting in school beginners in Hamburg]. Klin Monbl Augenheilkd. 1979; 174(2):232–235. German

304. Haile WA, Ayanaw TF, Alemayehu DG, Destaye SA. Prevalence and types of amblyopia among primary school children in Gondar town, Northwest Ethiopia. Open Access J Ophthalmol. (OAJO) 2017; 2(3):000124.

305. Hainline L, Riddell PM. Eye alignments and convergence in young infants. In: Vital-Durand F, Atkinson J, Braddick OJ, eds: Infant Vision. 1996. Oxford: Oxford Univ Press, pp. 221–247.

306. Hakim RB, Tielsch JM. Maternal cigarette smoking during pregnancy. A risk factor for childhood strabismus. Arch Ophthalmol. 1992; 110(10):1459–1462. doi: 10.1001/archopht.1992.01080220121033

307. Halboos MT, Mohammed MH, Al Jenabi ZK, Hamad N. Determination of Refractive Error and Strabismus in Two Primary Schools. J Opht Res Rev Rep. 2024; 5(1):1-5.

308. Hamidi A, Heravian Shandiz J, Jalalifar S, Boomi Quchan Atigh S, Darvishi A, Lashkardoost H. The Prevalence of Strabismus and Heterophoria in 3–6 Years Old Children in Bojnurd, Iran, in 2016-2017. J Paramed Sci Rehabil. 2020; 9(1):59-67. Persian

309. Hamilton JE. Vision anomalies of Indian school children: the Lame Deer study. J Am Optom Assoc. 1976; 47(4):479–487.

310. Han KE, Lim KH. Discrepancies between parental reports and clinical diagnoses of strabismus in Korean children. J AAPOS. 2012; 16(6):511–514. doi: 10.1016/j.jaapos.2012.07.005

311. Han KE, Baek SH, Kim SH, Lim KH; Epidemiologic Survey Committee of the Korean Ophthalmological Society. Prevalence and risk factors of strabismus in children and adolescents in South Korea: Korea National Health and Nutrition Examination Survey, 2008-2011. PLoS One. 2018; 13(2):e0191857. doi: 10.1371/journal.pone.0191857

312. Hanman K, Suda K, Koklanis K, Georgievski Z. Vision screening of individuals with mild intellectual disability. Aust Orthoptic J. 2009; 41(2):17–19.

313. Hansen AC. Comparative ophthalmology: black and white. J Natl Med Assoc. 1971; 63(6):445-449, 454.

314. Harada M, Yamamoto H. [Strabismus and amblyopia mass screening of preschool children.] Ganrin. 1961; 55(4): 374–377. Japanese

315. Hård AL. Results of vision screening of 6-year-olds at school: a population-based study with emphasis on screening limits. Acta Ophthalmol Scand. 2007; 85(4):415–418. doi: 10.1111/j.1600-0420.2006.00865.x

316. Hashemi H, Yekta A, Jafarzadehpur E, et al. The prevalence of strabismus in 7-year-old schoolchildren in Iran. Strabismus. 2015; 23(1):1–7. doi: 10.3109/09273972.2014.999795

317. Hashemi H, Nabovati P, Yekta A, Ostadimoghaddam H, Behnia B, Khabazkhoob M. The prevalence of strabismus, heterophorias, and their associated factors in underserved rural areas of Iran. Strabismus. 2017; 25(2):60–66. doi: 10.1080/09273972.2017.1317820

318. Hashemi H, Pakzad R, Heydarian S, et al. Global and regional prevalence of strabismus: a comprehensive systematic review and meta-analysis. Strabismus. 2019; 27(2):54–65. doi: 10.1080/09273972.2019.1604773

319. Hashemi H, Pakzad R, Nabovati P, et al. The prevalence of tropia, phoria and their types in a student population in Iran. Strabismus. 2020; 28(1):35–41. doi: 10.1080/09273972.2019.1697300

320. Hashemi H, Nabovati P, Yekta AA, et al. Binocular vision disorders in a geriatric population. Clin Exp Optom. 2022; 105(5):539–545. doi: 10.1080/08164622.2021.1922065

321. Havertape SA, Cruz OA, Chu FC. Sensory strabismus--eso or exo? J Pediatr Ophthalmol Strabismus. 2001; 38(6):327–30; quiz 354-355. doi: 10.3928/0191-3913-20011101-05

322. He J, Lu L, Zou H, et al. Prevalence and causes of visual impairment and rate of wearing spectacles in schools for children of migrant workers in Shanghai, China. BMC Public Health. 2014; 14:1312. doi: 10.1186/1471-2458-14-1312

323. He MG, Zeng J, Liu Y, Xu J, Pokharel GP, Ellwein LB. Refractive error and visual impairment in urban children in Southern China. Invest Ophthalmol Vis Sci. 2004; 45(3):793–799. doi: 10.1167/iovs.03-1051

324. He MG, Huang W, Zheng Y, Huang L, Ellwein LB. Refractive error and visual impairment in school children in rural southern China. Ophthalmology. 2007; 114(2):374–382. doi: 10.1016/j.ophtha.2006.08.020

325. He H, Fu J, Meng Z, Chen W, Li L, Zhao X. Prevalence and associated risk factors for childhood strabismus in Lhasa, Tibet, China: a cross-sectional, school-based study. BMC Ophthalmol. 2020; 20(1):463. doi: 10.1186/s12886-020-01732-2

326. He W, van der Most PJ, Ong JS, et al. Large-scale GWAS of strabismus identifies risk loci and provides support for a link with maternal smoking. Nat Commun. 2025; 16(1):7890. doi: 10.1038/s41467-025-62456-9

327. Hegde S, Mendonca N, Sripathi Kamath B, Vinay PG. Assessment of awareness and psychosocial impact of strabismus in rural India. Int J Biomed Res. 2014; 5(12):744–747. doi: 10.7439/ijbr.v5i12.849

328. Hegde SA, Joshi BS, Karambelkar VH. Prevalence of strabismus among school going children in India. Bioinformation. 2025; 21(3):410–413. doi: 10.6026/973206300210410

329. Heine B, Nurse D. African Languages. An Introduction. Cambridge Univ Press, Cambridge, 2000.

330. Helveston EM. 19th annual Frank Costenbader Lecture--the origins of congenital esotropia. J Pediatr Ophthalmol Strabismus. 1993; 30(4):215–232. doi: 10.3928/0191-3913-19930701-03

331. Hendrickson K, Bleything W. The visual profile of Romanian children and adults assessed through vision screenings. Optometry. 2001; 72(6):388–396.

332. Heng SJ, MacEwen CJ. Decrease in the rate of esotropia surgery in the United Kingdom from 2000 to 2010. Brit J Ophthalmol. 2013; 97(5):598–600. doi: 10.1136/bjophthalmol-2012-302726

333. Herron MS, Wang L, von Bartheld CS. Prevalence and Types of Strabismus in Cerebral Palsy: A Global and Historical Perspective Based on a Systematic Review and Meta-Analysis. Ophthalmic Epidemiol. 2025; 32(2):125–142. doi: 10.1080/09286586.2024.2331537

334. Herskovits MJ. Growth of interpupillary distance in American Negroes. Am J Phys Anthropol. 1926; 9(4):467–470.

335. Hertz J, Gombosh G, Avshalom A. Vision screening of students in Liberia. A preliminary report. J Pediatr Ophthalmol Strabismus. 1964; 1(3):33–36.

336. Hestnes A, Sand T, Fostad K. Ocular findings in Down’s syndrome. J Ment Defic Res. 1991; 35 (Pt 3):194–203. doi: 10.1111/j.1365-2788.1991.tb01052.x

337. Higgins JPT, Thomas J; Cochrane Collaboration. Cochrane Handbook for Systematic Reviews of Interventions. 2^nd^ edition. Hoboken: Wiley-Blackwell; 2020.

338. Hikmatullah BVS. Factors responsible for delayed presentation of strabismus in patients aging up to 16 years. Ophthalmol Update. 2018; 16(4):835–837.

339. Hirschberg J: The History of Ophthalmology, vol 1. Translated by Blodi FC. Bonn, Wayenbergh, 1982, p 110.

340. Holland G. [On the time of onset and the cause of strabismus in early childhood]. Klin Monbl Augenheilkd. 1965; 147(4):498–508. German

341. Holm S. [Le strabisme concomitant chez les palénégrides au Gabon, Afrique Equatoriale Française. Contribution å la question de race et de strabisme.] Acta Ophthalmol. 1939; 17:367–387. French

342. Holmström G, Rydberg A, Larsson E. Prevalence and development of strabismus in 10-year-old premature children: a population-based study. J Pediatr Ophthalmol Strabismus. 2006; 43(6):346–352. doi: 10.3928/01913913-20061101-04

343. Holst JC, Tjaland J. [Some figures from the ophthalmological department of the schools in Oslo.] Tidsskr Nor Laegeforen. 1962; 82:1291–1293. Norwegian

344. Holt R. A study of divergent strabismus in Australia. Brit Orthopt J. 1951; 8:95-100.

345. Hopkins S, Sampson GP, Hendicott PL, Wood JM. A Visual Profile of Queensland Indigenous Children. Optom Vis Sci. 2016; 93(3):251–258. doi: 10.1097/OPX.0000000000000797

346. Horta-Santini JM, Vergara C, Colón-Casasnovas JE, Izquierdo NJ. Strabismus surgery at the Puerto Rico Medical Center: A brief report. Puerto Rico Health Sci J. 2011; 30(4):203–205.

347. Horwood AM. Maternal observations of ocular alignment in infants. J Pediatr Ophthalmol Strabismus. 1993; 30(2):100–105. doi: 10.3928/0191-3913-19930301-09

348. Horwood A. Too much or too little: neonatal ocular misalignment frequency can predict later abnormality. Brit J Ophthalmol. 2003a; 87(9):1142–1145. doi: 10.1136/bjo.87.9.1142

349. Horwood A. Neonatal ocular misalignments reflect vergence development but rarely become esotropia. Brit J Ophthalmol. 2003b; 87(9):1146–1150. doi: 10.1136/bjo.87.9.1146

350. Horwood A, Williams B. Does neonatal ocular misalignment predict later abnormality? Eye (Lond). 2001; 15(Pt 4):485–491. doi: 10.1038/eye.2001.160

351. Horwood AM, Riddell PM. Can misalignments in typical infants be used as a model for infantile esotropia? Invest Ophthalmol Vis Sci. 2004; 45(2):714–720. doi: 10.1167/iovs.03-0454

352. Hosni FA, Green J. Orthoptics in the Gulf. Brit Orthopt J. 1979; 36:63-69.

353. Howells WW. Cranial Variation in Man. Peabody Museum of Archeology and Ethnology, Harvard University, Cambridge, MA, 1973.

354. Hu VH, Starling A, Baynham SN, Wager H, Shun-Shin GA. Accuracy of referrals from an orthoptic vision screening program for 3- to 4-year-old preschool children. J AAPOS. 2012; 16(1):49–52. doi: 10.1016/j.jaapos.2011.06.013

355. Hugonnier R, Hugonnier SC. 1969. In: Veronneau Troutman S (Ed.) Oculomotor Paralysis. Chapter 13, St Louis: CV Mosby, page 215.

356. Hultman O, Beth Høeg T, Munch IC, et al. The Danish Rural Eye Study: prevalence of strabismus among 3785 Danish adults - a population-based cross-sectional study. Acta Ophthalmol. 2019; 97(8):784–792. doi: 10.1111/aos.14112

357. Humphriss D. Divergent squint in Caucasoids. S Afr Med J. 1974; 48(8):286.

358. Hussein WA, Al-Gailani A, Al-Jammal AM. Psychological impacts on parents of children with strabismus in Baghdad: A cross-sectional study at Ibn Al-Haitham Eyes Teaching Hospital, 2023. Iraqi New Med J. 2025; 11(21):1-9.

359. Idrees Z, Dooley I, Fahy G. Horizontal strabismus surgical outcomes in a teaching hospital. Ir Med J. 2014; 107(6):176–178.

360. Inatomi M, Futenma M, Hayashi M, et al. [Natural course of uncorrected visual acuity and refraction among school children.] Jap J Clin Ophthal. (Rinsho Ganka) 1976; 30:1387–1397. Japanese

361. Ing MR, Pang SW. The racial distribution of strabismus. A statistical study. Hawaii Med J. 1974; 33(1):22–23.

362. Ing MR, Pang SWL. The racial distribution of strabismus. In: Reinecke RD, ed. Strabismus: Proceedings of the Third Meeting of the International Strabismological Association, May 10-12, 1978, Kyoto, Japan. New York: Grune & Stratton, 1978:107-109.

363. Ingram RM. The problem of screening children for visual defects. Brit J Ophthalmol. 1977; 61(1):4–7. doi: 10.1136/bjo.61.1.4

364. Iqbal Y, Niazi FK, Niazi MAK. Frequency of eye diseases in school age children. Pak J Ophthalmol. 2009; 25(4):185–190.

365. Iqbal S, Shafiq M, Zeeshan M, Nadeem HA, Jamshed M. Type of horizontal deviation in consanguinity. Pak J Ophthalmol. 2018; 34(2):103–106.

366. Israeli A, Bar-Asher T, Mezer E. Characteristics and Trends of Strabismus Surgeries at a Tertiary Hospital Over 2 Decades - What Can Be Learned for Years to Come? J Binoc Vis Ocul Motil. 2024; 74(2):84–90. doi: 10.1080/2576117X.2024.2364946

367. Ithnin MH, Othman SF, Ramli CN. RETROSPECTIVE ANALYSIS OF BINOCULAR VISION ANOMALIES (STRABISMUS AND NON-STRABISMUS) IN A HIGHER EDUCATION OPTOMETRY CLINIC, KUANTAN, MALAYSIA FROM 2018 UNTIL 2021. Int J Allied Health Sci. 2023; 7(5):371-380. doi: 10.31436/ijahs.v7i5.867

368. Jaeger EA. Ocular findings in Down’s syndrome. Trans Am Ophthalmol Soc. 1980; 78:808–845.

369. Jamali P, Fotouhi A, Hashemi H, Younesian M, Jafari A. Refractive errors and amblyopia in children entering school: Shahrood, Iran. Optom Vis Sci. 2009; 86(4):364–369. doi: 10.1097/OPX.0b013e3181993f42

370. Jampolsky A. Ocular deviations. Int Ophthalmol Clin. 1964; 4:567–701.

371. Jenchitr W, Pongprayoon C, Vongkittiruk, Imsuwan Y. [Ocular health in private school girls in Sukhumvit area of Bangkok.] J Health Sci. 2012; 21(3):489–498. Thai

372. Jenkins HR. Demographics: Geographic variations in the prevalence and management of exotropia. Am Orthopt J. 1992; 42:82–87.

373. Jeong SH, Kim US. Ten-Year Results of Home Vision-Screening Test in Children Aged 3-6 Years in Seoul, Korea. Semin Ophthalmol. 2015; 30(5-6):383–388. doi: 10.3109/08820538.2014.912335

374. Jeong C, Ozga AT, Witonsky DB, et al. Long-term genetic stability and a high-altitude East Asian origin for the peoples of the high valleys of the Himalayan arc. Proc Natl Acad Sci U S A. 2016; 113(27):7485–7490. doi: 10.1073/pnas.1520844113

375. Ji G. [An investigation of 4,125 cases of Kazak childhood strabismus and amblyopia]. Yan Ke Xue Bao. 1994; 10(4):193–196. Chinese

376. Jiao Y, Zhu Y, Zhou Z, et al. Strabismus surgery distribution during 10-year period in a tertiary hospital. Chin Med J (Engl). 2014; 127(16):2911–2914.

377. Jie Y, Xu Z, He Y, et al. A 4 year retrospective survey of strabismus surgery in Tongren Eye Centre Beijing. Ophthalmic Physiol Opt. 2010; 30(3):310–314. doi: 10.1111/j.1475-1313.2010.00716.x

378. Jin H, Yi JL, Xie H, et al. [A study on visual development among preschool children]. Zhonghua Yan Ke Za Zhi. (Chin J Ophthalmol) 2011; 47(12):1102–1106. Chinese

379. Jin K, Aboobakar IF, Whitman MC, Oke I. Mental Health Conditions Associated with Strabismus in a Diverse Cohort of US Adults. JAMA Ophthalmol. 2024; 142(5):472–475. doi: 10.1001/jamaophthalmol.2024.0540

380. Johnson GJ, Green JS, Paterson GD, Perkins ES. Survey of ophthalmic conditions in a Labrador community: II. Ocular disease. Can J Ophthalmol. 1984; 19(5):224–233.

381. Johnson GJ, Minassian DC, Weale RA, West SK, Gower EW, Kuper H, Lindfield R. Epidemiology of Eye Disease. 3^rd^ ed. 2012. Imperial College Press, 645 pp.

382. Jonkers GH. Statistics on deviations of binocular imbalance. Ophthalmologica. 1960; 140:180–192. doi: 10.1159/000303829

383. Joseph T. Early Indians: The Story of Our Ancestors and Where We Came From. Juggernaut, New Delhi, 2018.

384. Juárez-Muñoz IE, Rodríguez-Godoy ME, Guadarrama-Sotelo ME, Guerrero-Anaya M, Mejía-Arangúre JM, Sciandra-Rico M. [Incidence of common ophthalmological disorders in preschool children in Mexico City]. Salud Publica Mex. 1996; 38(3):212–216. Spanish

385. Junejo SA, Ansari MA. Outcome of monocular surgery for horizontal strabismus in Hyderabad. Clin Ophthalmol. 2010; 4:269–273. doi: 10.2147/opth.s8892

386. Junejo AY, Hassan M ul. Strabismus and its Types in Children of Age 6 to 15 Years Presenting at a Public Sector Hospital of Karachi. J Dow Univ Health Sci. 2019; 13:24–29.

387. Kaakinen K. Photographic screening for strabismus and high refractive errors of children aged 1-4 years. Acta Ophthalmol (Copenh). 1981; 59(1):38–44. doi: 10.1111/j.1755-3768.1981.tb06708.x

388. Kaakinen K, Kaseva H, Kause ER. Mass screening of children for strabismus or ametropia with two-flash photoskiascopy. Acta Ophthalmol (Copenh). 1986; 64(1):105–110. doi: 10.1111/j.1755-3768.1986.tb06882.x

389. Kac MJ, de Freitas Junior MB, Kac SI, de Andrade EP. [Frequency of ocular deviations at the strabismus sector of the Hospital do Servidor Publico Estadual de Sao Paulo.] Arq Bras Oftalmol. 2007; 70(6):939–942. Portuguese. doi: 10.1590/s0004-27492007000600010

390. Kahn HA. Letter: Ocular disease in South West Africa. Brit J Ophthalmol. 1974; 58(6):634. doi: 10.1136/bjo.58.6.634

391. Kalbe U, Berndt K, de Decker W. [Strabismus bei zerebralparetischen und ungesch*ä*digten Kindern. Vergleich der motorischen Symptome.] Klin Monbl Augenheilkd. 1979; 175(3):367–374. German

392. Kampanartsanyakorn S, Surachatkumtonekul T, Dulayajinda D, Jumroendararasmee M, Tongsae S. The outcomes of horizontal strabismus surgery and influencing factors of the surgical success. J Med Assoc Thai. 2005; 88 Suppl 9:S94–99.

393. Kalikivayi V, Naduvilath TJ, Bansal AK, Dandona L. Visual impairment in school children in Southern India. Indian J Ophthalmol. 1997; 45:129–134.

394. Karlica D, Galetović D, Znaor L, Bucat M. Strabismus incidence in infants born in Split-Dalmatia County 2002-2005. Acta Clin Croat. 2008; 47(1):5–8.

395. Käsmann-Kellner B, Heine M, Pfau B, Singer A, Ruprecht KW. [Screening for amblyopia, strabismus and refractive abnormalities in 1,030 kindergarten children]. Klin Monbl Augenheilkd. 1998; 213(3):166–173. German. doi: 10.1055/s-2008-1034968

396. Kattouf VM, Scharre J, McMahon J, Morrissey C, Korajczyk D, Beatty R. Comprehensive vision care in urban communities: the pediatric outreach program. Optometry. 2009; 80(1):29–35. doi: 10.1016/j.optm.2008.10.002

397. Kaymak NZ, Kaplan AT, Yalçın SÖ, Gün RD, Yaprak DÇ, Tanyıldız B. Outcomes of Eye Examination and Vision Screening in Term Infants Presenting to a Tertiary Hospital in Türkiye. Turkish J Ophthalmol. 2025; 55(2):86–91. doi: 10.4274/tjo.galenos.2025.07572

398. Kayser M. The human genetic history of Oceania: near and remote views of dispersal. Curr Biol. 2010; 20(4):R194–201. doi: 10.1016/j.cub.2009.12.004

399. Keiner GBJ. New Viewpoints on the Origin of Squint. A Clinical and Statistical Study on its Nature, Cause and Therapy. Martinus Nijhoff, The Hague, 1951.

400. Kemmanu V, Hegde K, Giliyar SK, Shetty BK, Kumaramanickavel G, McCarty CA. Prevalence of Childhood Blindness and Ocular Morbidity in a Rural Pediatric Population in Southern India: The Pavagada Pediatric Eye Disease Study-1. Ophthalmic Epidemiol. 2016; 23(3):185–192. doi: 10.3109/09286586.2015.1090003

401. Kendall JA, Stayte MA, Wortham C. Ocular defects in children from birth to 6 years of age. Br Orthopt J. 1989; 46:3–6.

402. Kerdoncuff E, Skov L, Patterson N, et al. 50,000 years of evolutionary history of India: Impact on health and disease variation. Cell. 2025; 188(13):3389–3404.e6. doi: 10.1016/j.cell.2025.04.027

403. Keshavarz K, Angha P, Sayehmiri F, Sayemiri K, Yasemi M. The prevalence of visual disorders in Iranian students: A meta-analysis study and systematic review. Electron Physician. 2017; 9(10):5516–5524. doi: 10.19082/5516

404. Khorrami-Nejad M, Akbari MR, Khosravi B. The prevalence of strabismus types in strabismic Iranian patients. Clin Optometr. 2018; 10:19–24. doi: 10.2147/OPTO.S147642

405. Khurana AK, Sikka KL, Parmar IP, Aggarwal SK. Ocular morbidity among school children in Rohtak City. Indian J Public Health. 1984;28(4):217–220.

406. Kikudi Z, Maertens K, Kayembe L. [Strabismus and heterophoria: the situation in Zaire]. J Fr Ophtalmol. 1988; 11(11):765–768. French

407. Kilgore KP, Barraza RA, Hodge DO, McKenzie JA, Mohney BG. Surgical correction of childhood intermittent exotropia and the risk of developing mental illness. Am J Ophthalmol. 2014; 158(4):788–792. doi: 10.1016/j.ajo.2014.06.008

408. Kim K, Ahn S, Koo B, Kim S. [Preschool vision screening for 3 to 6-year old children in Seoul.] J Korean Ophthalmol Soc. 2002; 43(4):714–727. Korean

409. Kim IG, Park JM, Lee SJ. Factors associated with the direction of ocular deviation in sensory horizontal strabismus and unilateral organic ocular problems. Korean J Ophthalmol. 2012; 26(3):199–202. doi: 10.3341/kjo.2012.26.3.199

410. Kingo AU, Ndawi BT. Prevalence and causes of low vision among schoolchildren in Kibaha District, Tanzania. Tanzan J Health Res. 2009; 11(3):111–115. doi: 10.4314/thrb.v11i3.47695

411. Kiorpes L, Boothe RG. Naturally occurring strabismus in monkeys (*Macaca nemestrina*). Invest Ophthalmol Vis Sci. 1981; 20(2):257–263.

412. Kiorpes L, Boothe RG, Carlson MR, Alfi D. Frequency of naturally occurring strabismus in monkeys. J Pediatr Ophthalmol Strabismus. 1985; 22(2):60–64. doi: 10.3928/0191-3913-19850301-07

413. Klebanoff MA. The Collaborative Perinatal Project: a 50-year retrospective. Paediatr Perinat Epidemiol. 2009; 23(1):2–8. doi: 10.1111/j.1365-3016.2008.00984.x

414. Knapp FN. The economic importance of squint in children and its effect in after years. Minnesota Med. 1931; 14(4):324–329.

415. Koca S, Halilbasic M. Results of horizontal strabismus surgery in the period from 2017 to 2023 at the Clinic for Eye Diseases Tuzla. Acta Med Saliniana. 2024; 53(2):34–39. doi: 10.5457/ams.v53i2.748

416. Köhler L, Stigmar G. Vision screening of four-year-old children. Acta Paediatr Scand. 1973; 62(1):17–27. doi: 10.1111/j.1651-2227.1973.tb08060.x

417. Köhler L, Stigmar G. Visual disorders in 7-year-old children with and without previous vision screening. Acta Paediatr Scand. 1978; 67(3):373–377. doi: 10.1111/j.1651-2227.1978.tb16337.x

418. Kollock CW. Certain Eye Affections in the Negro (Discussion). Trans Am Ophthalmol Soc. 1910; 12(Pt 2):463–465.

419. Konyama K. Refractive state and ocular deviation of the school children in Thailand. Orient Arch Ophthal. 1972; 10:229–237.

420. Kornder LD, Nursey JN, Pratt-Johnson JA, Beattie A. Detection of manifest strabismus in young children. I. A prospective study. Am J Ophthalmol. 1974a; 77(2):207–210. doi: 10.1016/0002-9394(74)90674-6

421. Kornder LD, Nursey JN, Pratt-Johnson JA, Beattie A. Detection of manifest strabismus in young children. 2. A retrospective study. Am J Ophthalmol. 1974b; 77(2):211-214. doi: 10.1016/0002-9394(74)90675-8

422. Krzystkowa K, Pajakowa J. The sensorial state in divergent strabismus. In: Orthopt Proc 2nd Int Orthopt Congr, pp. 72-76, Ed: Mein J., Excerpta Medica, Amsterdam, 1971

423. Kumah BD, Ebri A, Abdul-Kabir M, et al. Refractive error and visual impairment in private school children in Ghana. Optom Vis Sci. 2013; 90(12):1456–1461. doi: 10.1097/OPX.0000000000000099

424. Kumar D, Singh JV, Ahuja PC, Agarwal J, Mohan U. Ocular morbidity among school children in Sarojini nagar development block of Lucknow. Indian J Commun Med. 1992;17(3):109–113.

425. Kumar R, Dabas P, Mehra M, Ingle GK, Saha R, Kamlesh. Ocular morbidity amongst primary school children in Delhi. Health Popul Perspect Issues. 2007; 30(3):222–229.

426. Kunimi K, Goseki T, Nishina S, Negishi T, Sato M. 2021 trends in the treatment of patients with strabismus in Japan. Jpn J Ophthalmol. 2025; 69(1):10-16. doi: 10.1007/s10384-024-01144-5

427. Kuruvilla J, Srinivasa Rao PN. Ocular morbidity in school children in rural coastal area of Karnataka. Indian J Ophthalmol. 1978; 26(2):9–12.

428. Kvarnström G, Jakobsson P, Lennerstrand G. Visual screening of Swedish children: an ophthalmological evaluation. Acta Ophthalmol Scand. 2001; 79(3):240–244. doi: 10.1034/j.1600-0420.2001.790306.x

429. Laatikainen L, Erkkila H. Refractive errors and other ocular findings in school children. Acta Ophthalmol Scand. 1980; 58:129–136. doi: 10.1111/j.1755-3768.1980.tb04576.x

430. Labadi L, Shahin R, Eperjesi F, Al-Shanti Y, Shehadeh M, Taha I. Prevalence of Visual Disorders among Urban Palestinian Preschool Children. Open Ophthalmol J. 2022; 16(1):e187436412112241. doi: 10.2174/18743641-v16-e2112241

431. Lagleyze P. [Du Strabisme. Recherches Etiologiques - Pathogenie Mecanisme du Traitement.] Paris: Jules Rousset, 1913, 409 pp.

432. Lai YH, Hsu HT, Wang HZ, Chang SJ, Wu WC. The visual status of children ages 3 to 6 years in the vision screening program in Taiwan. J AAPOS. 2009; 13(1):58–62. doi: 10.1016/j.jaapos.2008.07.006

433. Lambert SR. Are there more exotropes than esotropes in Hong Kong? Brit J Ophthalmol. 2002; 86(8):835–836. doi: 10.1136/bjo.86.8.835

434. Lambert SR. Population-Based Incidence of Strabismus: Why Is It Important? JAMA Ophthalmol. 2017; 135(10):1053–1054. doi: 10.1001/jamaophthalmol.2017.3123

435. Lança C, Serra H, Prista J. Strabismus, visual acuity, and uncorrected refractive error in Portuguese children aged 6 to 11 years. Strabismus. 2014; 22(3):115–119. doi: 10.3109/09273972.2014.932395

436. Lance P, Mitchell R. Australian contribution to international orthoptic association survey. Aust Orthopt J. 1983; 20:59-63.

437. Lang D, Leman R, Arnold AW, Arnold RW. Validated portable pediatric vision screening in the Alaska Bush. A VIPS-like study in the Koyukon. Alaska Med. 2007; 49(1):2–15.

438. Lang L, Guo K, Zhang L, et al. The distribution characteristics of strabismus surgery types in a tertiary hospital in the Central Plains region during the COVID-19 epidemic. BMC Ophthalmol. 2024; 24(1):67. doi: 10.1186/s12886-024-03327-7

439. Larsson E, Holmström G, Rydberg A. Ophthalmological findings in 10-year-old full-term children- a population-based study. Acta Ophthalmol. 2015; 93(2):192–198. doi: 10.1111/aos.12476

440. Lau J, Ioannidis JP, Terrin N, Schmid CH, Olkin I. The case of the misleading funnel plot. BMJ. 2006; 333:597–600. doi: 10.1136/bmj.333.7568.597

441. Laughton SC, Hagen MM, Yang W, von Bartheld CS. Gender differences in horizontal strabismus: Systematic review and meta-analysis shows no difference in prevalence, but gender bias towards females in the clinic. J Glob Health. 2023; 13:04085. doi: 10.7189/jogh.13.04085

442. Lavrich JB. Intermittent exotropia: continued controversies and current management. Curr Opin Ophthalmol. 2015; 26(5):375–381. doi: 10.1097/ICU.0000000000000188

443. Lee JF, Kim CZ, Nam KY, Lee SU, Lee SJ. [An epidemiological survey of strabismus and nystagmus in South Korea: KNHANES V.] J Korean Ophthalmol Soc. 2017; 58(11):1260–1268. Korean. doi: 10.3341/jkos.2017.58.11.1260

444. Lee SH, Jung SJ, Ohn YH, Chang JH. Association between refractive errors and horizontal strabismus: the Korea National Health and Nutrition Examination Survey. J AAPOS. 2021; 25(6):340.e1-340.e7. doi: 10.1016/j.jaapos.2021.07.012

445. Lee YH, Repka MX, Borlik MF, et al. Association of Strabismus with Mood Disorders, Schizophrenia, and Anxiety Disorders among Children. JAMA Ophthalmol. 2022; 140(4):373–381. doi: 10.1001/jamaophthalmol.2022.0137

446. Lennerstrand G, Gallo JE. Prevalence of refractive errors and ocular motility disorders in 5- to 10-year-old Swedish children born prematurely or at full-term. Acta Ophthalmol (Copenh). 1989; 67(6):717–718. doi: 10.1111/j.1755-3768.1989.tb04408.x

447. Leone JF, Cornell E, Morgan IG, et al. Prevalence of heterophoria and associations with refractive error, heterotropia and ethnicity in Australian school children. Br J Ophthalmol. 2010; 94(5):542–546. doi: 10.1136/bjo.2009.163709

448. Li JH, Xie WF, Tian JN, Zhang LJ, Cao MM, Wang L. Changing Strabismus Surgery Distribution at Shanxi Province Eye Hospital in Central China. J Pediatr Ophthalmol Strabismus. 2017; 54(2):112–116. doi: 10.3928/01913913-20161013-01

449. Liang YS, Lai IC, Loke TY, Chen TT. [Preliminary report of ocular examination in school children.] Acta Soc Ophthalmol Sin. 1984; 23:1–7. Chinese

450. Lieu AC, Walker EH, Robbins SL, Granet DB, Rudell JC. Epidemiology of strabismus among adults in the United States: insights from the All of Us database. J AAPOS. 2025 Sep 29:104657. doi: 10.1016/j.jaapos.2025.104657

451. Lillvis JH, Feehan M, Shwani T, et al. Strabismus and Strabismus Surgery in the U.S. Veterans Health Administration: Foundational Analyses of Electronic Health Record Data from 2000 to 2022. J Pers Med. 2025; 15(2):40. doi: 10.3390/jpm15020040

452. Lim HC, Quah BL, Balakrishnan V, Lim HC, Tay V, Emmanuel SC. Vision screening of 4-year-old children in Singapore. Singapore Med J. 2000; 41(6):271–278.

453. Lim HT, Yu YS, Park SH, et al. The Seoul Metropolitan Preschool Vision Screening Programme: results from South Korea. Brit J Ophthalmol. 2004; 88(7):929–933. doi: 10.1136/bjo.2003.029066

454. Lin S, Huang Y, Ma D, et al. [A cross-sectional study of strabismus prevalence in urban and rural primary and secondary school students in Shantou City.] Chin J Strabismus Pediatr Ophthalmol. 2013; 21(3):56–56. Chinese

455. Lin S, Congdon N, Yam JC, et al. Alcohol use and positive screening results for depression and anxiety are highly prevalent among Chinese children with strabismus. Am J Ophthalmol. 2014; 157(4):894–900. doi: 10.1016/j.ajo.2014.01.012

456. Lin S, Gong W, Chen B, Zhang M. Prevalence of intermittent exotropia among primary and secondary school students in Shantou, China. Eye Sci. 2016; 31(1):7–12.

457. Lithander J. Prevalence of amblyopia with anisometropia or strabismus among schoolchildren in the Sultanate of Oman. Acta Ophthalmol Scand. 1998; 76(6):658–662. doi: 10.1034/j.1600-0420.1998.760604.x

458. Louwagie CR, Diehl NN, Greenberg AE, Mohney BG. Is the incidence of infantile esotropia declining? A population-based study from Olmsted County, Minnesota, 1965 to 1994. Arch Ophthalmol. 2009; 127(2):200-203. doi: 10.1001/archophthalmol.2008.568

459. Lu P, Chen X, Zhang W, Chen S, Shu L. Prevalence of ocular disease in Tibetan primary school children. Can J Ophthalmol. 2008; 43(1):95–99. doi: 10.3129/i07-194

460. Luoma-Overstreet GN, Kumar V, Lam K, Brown DD, Couser NL. The epidemiology of strabismus and cataracts within a pediatric population in Saint Vincent and the Grenadines: an analysis of 201 consecutive cases. Int J Mol Epidemiol Genet. 2023; 14(1):11–18.

461. Lyle TK, Wybar K. 1967. Lyle and Jackson’s Practical Orthoptics in the Treatment of Squint, 5^th^ ed, pp 326-329.

462. Lyle TK. 1950. Worth and Chavasses’s Squint. The Binocular Reflexes and the Treatment of Strabismus. 8^th^ ed. The Blakiston Company: Philadelphia. https://babel.hathitrust.org/cgi/pt?id=uc1.b4208491;view=1up;seq=16

463. Maaita JF, Sunna LF, Al-Madani MV, Horrani SM. Eye diseases in children in Southern Jordan. Saudi Med J. 2003; 24(2):154–156.

464. MacEwen CJ, Chakrabarti HS. Why is squint surgery in children in decline? Brit J Ophthalmol. 2004; 88(4):509–511. doi: 10.1136/bjo.2002.016618

465. Macfarlane DJ, Fitzgerald WJ, Stark DJ. The prevalence of ocular disorders in 1000 Queensland primary schoolchildren. Aust N Z J Ophthalmol. 1987; 15(3):161–174. doi: 10.1111/j.1442-9071.1987.tb00066.x

466. MacQueen IAG, Sutherland MS. An investigation of the epidemiology of squint. Health Bull. 1953; 11:65–68.

467. Madhavi MR, Kesuraju V, Nagrale P, Poka A. Ocular morbidity among school aged children in Indian scenario. Int J Res Med Sci. 2015; 3(6):1431–1434.

468. Magalhães AL, Ferreira KM, Prado KF, Salgado LM. Frequência dos desvios oculares no ambulatório de estrabismo em um hospital terciário do interior de São Paulo. Braz J Health Rev. 2023; 6(5):25701–25707. Portuguese. doi: 10.34119/bjhrv6n5-557

469. Magramm I, Schlossman A. Strabismus in patients over the age of 60 years. J Pediatr Ophthalmol Strabismus. 1991; 28(1):28–31.

470. Mahachaiyakul A, Sinpornchai N, Kunavisarut S. [The study of refractive state and strabismus prevalence in school children.] Thai J Ophthalmol. 1997; 11:1–8. Thai

471. Mahajan S, Gupta SK. Clinical study of concomitant strabismus. Perspect Med Res. 2020; 8(3):44–48.

472. Mahdi Z, Munami S, Shaikh ZA, Awan H, Wahab S. Pattern of Eye Diseases in Children at Secondary Level Eye Department in Karachi. Pak J Ophthalmol. 2005; 22(3):145–151.

473. Majima A, Nakajima A, Ichikawa H, Watanabe M. Prevalence of ocular anomalies among school children. Am J Ophthalmol. 1960; 50:139–146. doi: 10.1016/0002-9394(60)90848-5

474. Maĭmulov VG. [State of visual functions of Leningrad preschool children]. Oftalmol Zh. 1971; 26(5):378–381. Russian

475. Majumder PP. People of India: Biological diversity and affinities. Evol Anthropol. 1998; 6(3):100–110.

476. Makiuchi S, Yamaji R, Kozaki M, et al. [On the investigations of amblyopic children and students in compulsory course in Osaka-fu.] Rinsho Ganka (Jpn J Clin Ophthalmol.) 1962; 16:151–165. Japanese

477. Malaspinas AS, Westaway MC, Muller C, et al. A genomic history of Aboriginal Australia. Nature. 2016; 538:207–214. doi: 10.1038/nature18299

478. Malik MA, Habiba U-e. Frequency of esotropia and exotropia among patients between 3 to 25 years of age. Al-Shifa J Ophthalmol. 2013; 9:34-38.

479. Malik S, Sughra U, Altaf S, Kausar S, Ahmad A, Imran M. Strabismus in Patients with Low Vision Visiting a Tertiary Eye Care Setting in Rawalpindi. Al-Shifa J Ophthalmol. 2018; 14(2):92–98.

480. Malik SN, Masud H, Zamir Q. Frequency of Esotropia and its Management in Pediatric Age Group at CMH Quetta. Pak Armed Forces Med J. 2024; 74(Suppl-2):S324-S328. doi: 10.51253/pafmj.v74iSUPPL-2.6037

481. Mandel Y, Grotto I, El-Yaniv R, et al. Season of birth, natural light, and myopia. Ophthalmology. 2008; 115(4):686–692. doi: 10.1016/j.ophtha.2007.05.040

482. Mann I. Culture, Race, Climate and Eye Disease. Springfield, Illinois, Charles C. Thomas, 1966. 580 pages.

483. Mann I. The geographic approach to ophthalmology. Arch Soc Amer Oftal Optom. 1967; 6:191–193.

484. Mann I, Rountree P. Geographic ophthalmology. A report on a recent survey of Australian aboriginals. Am J Ophthalmol. 1968; 66(6):1020–1034.

485. Mann I, Potter D. Geographic ophthalmology. A preliminary study of the Maoris of New Zealand. Am J Ophthalmol. 1969; 67(3):358–369.

486. Mann I. Eye disease in the Eskimo and in the Australian Aboriginal: a brief comparison. Acta Ophthalmol (Copenh). 1972; 50(4):543–548. doi: 10.1111/j.1755-3768.1972.tb05982.x

487. Mannhardt J. [*Muskuläre* Asthenopie und Myopie.] Arch Ophthalmol. 1871; 17(2):69–97. German

488. Mansour OM, Elbassiouny O, Abdelhamid M, Awadalla A, Mohammed MA. A Study of Ophthalmic Problems Among Children Attending Ophthalmic Outpatient Clinic, Suez Canal University Hospital. Suez Canal Univ Med J. 2023; 26(8):20-29.

489. Maples WC, Atchley J, Ashby W, Ficklin T. An epidemiological study of the ocular and visual profiles of Oklahoma Cherokees and Minnesota Chippewas. J Am Optom Assoc. 1990; 61(10):784–788.

490. Márquez Galvis MM, Cáceres Díaz MC. [Visual and ocular health profile of children from two child development centers in Pereira, Colombia.] Cienc Tecnol Salud Vis Ocul. 2017; 15(2):61–70. Spanish

491. Martínez J, Cañamares S, Saornil MA, Almaraz A, Pastor JC. Original papers: Prevalence of amblyogenic diseases in a preschool population sample of Valladolid, Spain. Strabismus. 1997; 5(2):73–80. doi: 10.3109/09273979709057390

492. Martinez-Thompson JM, Diehl NN, Holmes JM, Mohney BG. Incidence, types, and lifetime risk of adult-onset strabismus. Ophthalmology. 2014; 121(4):877–882. doi: 10.1016/j.ophtha.2013.10.030

493. Maruo T, Kubota N, Arimoto H. [The results of examinations for strabismus and amblyopia in elementary and secondary school pupils.] Ganka Rinsho Iho (Jpn Rev Clin Ophthalmol.) 1977; 71:712–714. Japanese

494. Mason A, Lindberg L, Joronen K, Koivisto A-M, Rantanen A. Strabismus is more than a misalignment; a cross-sectional pilot study of HRQOL in Finnish strabismic adults referred to a university hospital. Acta Ophthalmol. 2024; 102:428–434. doi: 10.1111/aos.15813.

495. Masood A, Amitava AK, Khalid A, et al. Ocular morbidity and co-morbidities in children attending a nodal district early intervention center in Uttar Pradesh. Indian J Ophthalmol. 2023; 71(1):203–208. doi: 10.4103/ijo.IJO_1637_22

496. Mathers M, Keyes M, Wright M. A review of the evidence on the effectiveness of children’s vision screening. Child Care Health Dev. 2010; 36(6):756–780. doi: 10.1111/j.1365-2214.2010.01109.x

497. Matsuda K, Yokoyama T, Kozaki M. The relationship between the incidence of esotropia and birth month: Part 2 - Focusing on the seasonal variation of hyperopia. Folia Japon Ophthalmol Clin. 2009; 2(2):156–160. Japanese

498. Matsuda K, Yokoyama T, Ra KE, Kozaki M. [Long-term changes in the ratio of exotropia to esotropia patients]. Nippon Ganka Gakkai Zasshi. 2013; 117(5):427–432. Japanese

499. Matsuda K, Yokoyama T, Fukui M, Morita T, Kozaki M. [Inverse analysis of the relationship between ocular refraction and birth month using matrix equations.] Nippon Ganka Gakkai Zasshi (J Jpn Ophthalmol Soc.) 2016; 120(8):533–539. Japanese

500. Matsuo T, Yamane T, Ohtsuki H. Heredity versus abnormalities in pregnancy and delivery as risk factors for different types of comitant strabismus. J Pediatr Ophthalmol Strabismus. 2001; 38(2):78–82. doi: 10.3928/0191-3913-20010301-08

501. Matsuo T, Matsuo C. The prevalence of strabismus and amblyopia in Japanese elementary school children. Ophthalmic Epidemiol. 2005; 12(1):31–36. doi: 10.1080/09286580490907805

502. Matsuo T, Matsuo C. Comparison of prevalence rates of strabismus and amblyopia in Japanese elementary school children between the years 2003 and 2005. Acta Med Okayama. 2007a; 61(6):329–334. doi: 10.18926/AMO/32877

503. Matsuo T, Matsuo C, Matsuoka H, Kio K. Detection of strabismus and amblyopia at 1.5- and 3-year-old children by preschool vision-screening program in Japan. Acta Med Okayama. 2007b; 61:9–16. doi: 10.18926/AMO/32910

504. Matsuo T, Hamasaki I, Kamatani Y, et al. Genome-Wide Association Study with Three Control Cohorts of Japanese Patients with Esotropia and Exotropia of Comitant Strabismus and Idiopathic Superior Oblique Muscle Palsy. Int J Mol Sci. 2024; 25(13):6986. doi: 10.3390/ijms25136986

505. Maul E, Barroso S, Munoz SR, Sperduto RD, Ellwein LB. Refractive error study in children: results from La Florida, Chile. Am J Ophthalmol. 2000; 129(4):445–454. doi: 10.1016/s0002-9394(99)00454-7

506. Mazepa GM. [Results of a thorough eye examination in kindergarten children of the city of Saratov]. Oftalmol Zh. 1967; 22(2):151–154. Russian

507. McBride A, Arblaster G. Schizophrenia and Orthoptic Conditions: A Literature Review. Br Ir Orthopt J. 2024; 20(1):133–145. doi: 10.22599/bioj.327

508. McLaren DS. Nutrition and eye disease in East Africa: experience in Lake and Central Provinces, Tanganyika. J Trop Med Hyg. 1960; 63:101–122.

509. McLaren DS. The refraction of Indian school children: a comparison of data from East Africa and India. Br J Ophthalmol. 1961; 45(9):604–613. doi: 10.1136/bjo.45.9.604

510. McKean-Cowdin R, Cotter SA, Tarczy-Hornoch K, et al. Prevalence of amblyopia or strabismus in Asian and non-Hispanic white preschool children: multi-ethnic pediatric eye disease study. Ophthalmology. 2013; 120:2117–2124. doi: 10.1016/j.ophtha.2013.03.001

511. McKenzie JA, Capo JA, Nusz KJ, Diehl NN, Mohney BG. Prevalence and sex differences of psychiatric disorders in young adults who had intermittent exotropia as children. Arch Ophthalmol. 2009; 127(6):743–747. doi: 10.1001/archophthalmol.2009.68

512. McMahon G, Zayats T, Chen YP, Prashar A, Williams C, Guggenheim JA. Season of birth, daylight hours at birth, and high myopia. Ophthalmology. 2009; 116(3):468–473. doi: 10.1016/j.ophtha.2008.10.004

513. McNeil NL. Patterns on visual defects in children. Brit J Ophthalmol. 1955; 39(11):688–701. doi: 10.1136/bjo.39.11.688

514. Medghalchi A. [A study on prevalence of horizontal strabismus in patients under 14 years.] J Guilan Univ Med Sci. 2003; 12(47):80–85. Persian

515. Medkova L, Bersky K, Bublikova D, Hajek F. [Appearance of strabismus in children of mothers with late pregnancy toxemia.] Cesk Oftalmol. 1959; 15:254–257. Russian

516. Mehari ZA, Yimer AW. Prevalence of refractive errors among schoolchildren in rural central Ethiopia. Clin Exp Optom. 2013; 96(1):65–69. doi: 10.1111/j.1444-0938.2012.00762.x

517. Melek N. [Estrabismo.] Arch Argent Pediatr 1973; 71(1):14-16. Spanish

518. Merina M, Lav K, Vaibhav S, Sagar B, Maneesh S, Lal MA. Retrospective Analysis of Children Coming for First Eye Check up in a Tertiary Center. Int J Pharmaceut Clinical Res. 2024; 16(2):1149–1156.

519. Merino P, Mateos C, Gómez De Liaño P, Franco G, Nieva I, Barreto A. [Horizontal sensory strabismus: characteristics and treatment results]. Arch Soc Esp Oftalmol. 2011; 86(11):358–362. Spanish. doi: 10.1016/j.oftal.2011.05.028

520. Merino Sanz P, Donoso Torres HE, Gómez de Liaño Sánchez P, Casco Guijarro J. Current trends of strabismus surgery in a tertiary hospital. Arch Soc Esp Oftalmol (Engl Ed). 2020; 95(5):217–222. doi: 10.1016/j.oftal.2020.01.007

521. Meyer FA, Campusano C, Reyes J, Valdenegro JP. [Epidemiologic aspects of strabismus in Valparaíso, Chile]. Rev Med Chil. 1978; 106(9):718–720. Spanish

522. Miller FJW, Court SDM, Walton WS, Knox EG. Growing Up in Newcastle Upon Tyne. New York, Oxford Univ Press, 1960, 369 pp.

523. Minor JL. Some impressions of certain eye affections in the negro as compared with the white race. Trans Am Ophthalmol Soc. 1910; 12(Pt 2):459–465.

524. Mishol N, Herzlinger G, Rak Y, Smilanksy U, Carmel L, Gokhman D. Candidate Denisovan fossils identified through gene regulatory phenotyping. Proc Natl Acad Sci U S A. 2025; 122(35):e2513968122. doi: 10.1073/pnas.2513968122

525. Mittal S, Maitrea A, Dhasmana R. Clinical profile of refractive errors in children in a tertiary care hospital of Northern India. Int J Community Med Public Health. 2016; 3(5):1189–1194.

526. Miyata M, Kido A, Miyake M, et al. Prevalence and Incidence of Strabismus by Age Group in Japan: A Nationwide Population-Based Cohort Study. Am J Ophthalmol. 2024; 262:222–228. doi: 10.1016/j.ajo.2023.11.022

527. Mocanu V, Horhat R. Prevalence and risk factors of amblyopia among refractive errors in an Eastern European population. Medicina (Kaunas). 2018; 54(1):6. doi: 10.3390/medicina54010006

528. Moguel-Ancheita S, Ramírez-Sibaja S, Castellanos-Pérez BC, Orozco-Gómez LP. [Análisis de las funciones sensoriomotoras y depresión en niños con estrabismo. Primera fase.] Cir Cir. 2008; 76(2):101–107. Spanish

529. Moguel-Ancheita S, Ramírez-Sibaja S, Reyes-Pantoja SA, Orozco-Gómez LP. [Funciones visuomotoras e inteligencia posterior al tratamiento del estrabismo. Segunda fase.] Cir Cir. 2010; 78(6):470–475. Spanish

530. Mohan A, Bisht A, Sharma VK, Jamil Z. Epidemiology of ocular morbidity among school-going children. All India Ophthalmol Soc Proc. 2017; FP1242:67-90.

531. Moher D, Liberati A, Tetzlaff J, Altman DG, The PRISMA Group. Preferred reporting items for systematic reviews and meta-analyses: The PRISMA statement. PLOS Med. 2009; 6(7): e1000097. doi: 10.1371/journal.pmed.1000097

532. Mohney BG. Common forms of childhood strabismus in an incidence cohort. Am J Ophthalmol. 2007a; 144(3):465–467. doi: 10.1016/j.ajo.2007.06.011

533. Mohney BG. The development of psychiatric disease in young adults who had childhood strabismus. Acta Ophthalmol. 2007b; 85:(s240).

534. Mohney BG, Greenberg AE, Diehl NN. Age at strabismus diagnosis in an incidence cohort of children. Am J Ophthalmol. 2007c; 144(3):467–469. doi: 10.1016/j.ajo.2007.04.022

535. Mohney BG, McKenzie JA, Capo JA, Nusz KJ, Mrazek D, Diehl NN. Mental illness in young adults who had strabismus as children. Pediatrics. 2008; 122(5):1033–1038. doi: 10.1542/peds.2007-3484

536. Mohney BG, Lepor L, Hodge DO. Subclinical markers of strabismus in children 5-18 years of age. J AAPOS. 2021; 25(3):139.e1-139.e5. doi: 10.1016/j.jaapos.2021.02.008

537. Molinari A, Heede S, Virgili G, Angi M. [Trastornos de la motilidad ocular en una población de niños escolares de la Sierra Ecuatoriana / Ocular motility disorders in a population of school children in the Sierra Ecuatoriana.] Arch Chil Oftalmol. 2005; 63(2):359–362. Spanish

538. Monteiro S, Casal I, Vale C, et al. [Estrabismo em idade ambliogénica: estudo retrospetivo de 12 meses consecutivos de referenciação oftalmológica hospitalar.] Oftalmologia. 2016; 40(4):317–323. Portuguese. doi: 10.48560/rspo.8010

539. Montvilaite D, Grizickaite A, Augyte A, Skvarciany I, Barkus A, Usonis V. Ophthalmological follow-up of prematurely born children in preschool age: prospective study of visual acuity, refractive errors and strabismus. Acta Med Lituanica. 2015; 22(4):205–215.

540. Moore S. Orthoptic treatment for intermittent exotropia. Amer Orthopt J. 1963; 13:14-20.

541. Morales OY, Ghoul S, Muguercia GY, Delfino LRJ, Divasto CG. [Surgical behavior in the Ophthalmological Hospital “Friendship Algeria-Cuba.”] Rev Inf Cient. 2018; 97(1):10–18. Spanish

542. Morgan A. Incidence of divergent squint among fair-haired people. Brit Orthopt J. 1957; 14:111–112.

543. Mujica A, Navarro F. [Estudio estadistico de las affecciones de los ojos.] Rev Med Chile. 1897; 25(4):77–170. Spanish

544. Mukhaiser MH, Al-Khateeb HS, Mohammed IA. Prevalence of Lazy Eyes Among Primary School Students in Rural Areas of Baghdad City. J Adv Sci Nanotechnol. 2023; 2(1):204–214. doi: 10.55945/joasnt.2023.2.1.204-214

545. Multi-ethnic Pediatric Eye Disease Study Group. Prevalence of amblyopia and strabismus in African American and Hispanic children ages 6 to 72 months: the Multi-ethnic Pediatric Eye Disease Study. Ophthalmology. 2008; 115:1229-1236. doi: 10.1016/j.ophtha.2007.08.001

546. Murthy GVS, Gupta SK, Ellwein LB, Muñoz SR, Pokharel GP, Sanga L, Bachani D. Refractive error in children in an urban population in New Delhi. Invest Ophthalmol Vis Sci. 2002; 43(3):623–631.

547. Musa KO, Ikuomenisan SJ, Idowu OO, Salami MO, Olowoyeye AO. Spectrum of childhood strabismus seen at Lagos University Teaching Hospital, Lagos, Nigeria. Nig Qt J Hosp Med. 2017; 27(2):726-732.

548. Mvilongo C, Omgbwa A, Nkidiaka C, Elom A, Hoffman W, Ebana C. Strabismus amblyopia in young Cameroonian at their first visit at Yaounde Hospital Centre-Essos. J Clin Exp Ophthalmol. 2016; 7:613.

549. Mvogo CE, Bella-Hiag AL, Ellong A, Mbarga BM, Epesse M. [Exotropia in black Cameroonians]. Sante. 1999; 9(5):289–292. French

550. Mvogo CE, Ellong A, Bella-Hiag AL, Luma-Namme H. [Hereditary factors in strabismus]. Sante. 2001; 11(4):237–239. French

551. Naidoo KS, Raghunandan A, Mashige KP, et al. Refractive error and visual impairment in African children in South Africa. Invest Ophthalmol Vis Sci. 2003; 44(9):3764–3770. doi: 10.1167/iovs.03-0283

552. Naik R, Gandhi J, Shah N. Prevalence of Ocular Morbidity among School Going Children (6-15years). Sch J App Med Sci. 2013; 1(6):848–851. doi: 10.36347/sjams.2013.v01i06.0044. 848

553. Najafi A. [Prevalence of different types of strabismus in Labbafinejad Hospital.] Med Sci. (J Islam Azad Univ.) 2007; 17(1):33–36. Persian

554. Nakagawa J, Suzuki S. [The incidence of strabismus in schoolchildren.] J Sapporo City Gen Hosp. 1954; 36:60–65. Japanese.

555. Nakajima A, Yoshimoto T, Ito N, Kimura T, Majima A, Awaya S. [Distribution of eye diseases among school children.] Rinsho Ganka (Jpn J Clin Ophthalmol). 1960; 14:1762–1769. Japanese

556. Narayanan A, Krishnamurthy SS, Kumar R K. Status of Eye Health among School Children in South India - Sankara Nethralaya School Children Eye Examination Study (SN-SEES). Ophthalmic Epidemiol. 2021; 28(4):349–358. doi: 10.1080/09286586.2020.1849743

557. Nartey ET, van Staden DB, Amedo AO. Prevalence of ocular anomalies among schoolchildren in Ashaiman, Ghana. Optom Vis Sci. 2016; 93(6):607–611. doi: 10.1097/OPX.0000000000000836

558. Natung T, Devi OS, Thangkhiew L, Paul S. Clinical pattern and burden of strabismus in a teaching institute of Northeast India. J Family Med Prim Care. 2024; 13:5739–5744. doi: 10.4103/jfmpc.jfmpc_1032_24

559. Naylor EJ, Wright AG. Incidence of amblyopia in children with strabismus. Brit Orthopt J. 1959; 16:109–113.

560. Ndlovu D, Nhleko S, Pillay Y, Tsiako T, Yusuf N, Hansraj R. The prevalence of strabismus in schizophrenic patients in Durban, KwaZulu Natal. S Afr Optom. 2011; 70:101–108. doi: 10.4102/aveh.v70i3.106

561. Neena R, Gayathri MS, Prakash N, Anantharaman G. Impact of online classes on eye health of children and young adults in the setting of COVID-19 pandemic: A hospital-based survey. Oman J Ophthalmol. 2023; 16(1):45–50. doi: 10.4103/ojo.ojo_57_22

562. Nelson LB. Diagnosis and management of strabismus and amblyopia. Pediatr Clin North Am. 1983; 30:1003–1014. doi: 10.1016/s0031-3955(16)34498-4

563. Nepal BP, Koirala S, Adhikary S, Sharma AK. Ocular morbidity in schoolchildren in Kathmandu. Brit J Ophthalmol. 2003; 87(5):531–534. doi: 10.1136/bjo.87.5.531

564. Neumann E, Eibschitz N, Hyams S, Friedman Z. Ophthalmic screening in child welfare clinics in Israel with particular reference to strabismus and amblyopia. J Pediatr Ophthalmol Strabismus. 1971; 8:257–260.

565. Newman DK, Hitchcock A, McCarthy H, Keast-Butler J, Moore AT. Preschool vision screening: outcome of children referred to the hospital eye service. Brit J Ophthalmol. 1996; 80(12):1077–1082. doi: 10.1136/bjo.80.12.1077

566. Nickla DL. Ocular diurnal rhythms and eye growth regulation: where we are 50 years after Lauber. Exp Eye Res. 2013; 114:25–34. doi: 10.1016/j.exer.2012.12.013

567. Nirmalan PK, Vijayalakshmi P, Sheeladevi S, Kothari MB, Sundaresan K, Rahmathullah L. The Kariapatti pediatric eye evaluation project: baseline ophthalmic data of children aged 15 years or younger in Southern India. Am J Ophthalmol. 2003; 136(4):703–709. doi: 10.1016/s0002-9394(03)00421-5

568. Nixon RB, Helveston EM, Miller K, Archer SM, Ellis FD. Incidence of strabismus in neonates. Am J Ophthalmol. 1985; 100(6):798–801. doi: 10.1016/s0002-9394(14)73370-7

569. Njambi L, Rita O, Kazim D, Sonia V. Prevalence and pattern of manifest strabismus in paediatric patients at CCBRT, Dar es Salaam, Tanzania. J Ophthalmol East Cent South Afri. (JOECSA) 2017; 21:9–12.

570. Nkanga ED, Okonkwo SN, Oyeniyi TI, et al. Spectrum of Manifest Strabismus in A Tertiary Eye Care Hospital in Calabar, Nigeria: Demographics, Types and Co-morbidities. West Afr J Med. 2024; 41(9):919–926.

571. Noori AR, Taher AH. Association of Horizontal Strabismus and Refractive Errors for Children in Baghdad, Iraq. Med Sci J Adv Res. 2024; 4(4):259–268. doi: 10.46966/msjar.v4i4.160

572. Nordenson JW. Die Stellung der Seitenwände der Augenhöhle beim Menschen in verschiedenen Lebensaltern. Acta Chirurg Scand. 1920; 52:45–54. German

573. Nordlöw W. [Ögonen inom den förebyggande barnavården.] Svenska Läk-tidn. 1944; 41:1385–1395. Swedish

574. Nordlöw W. Age distribution of the onset of esotropia. Br J Ophthalmol. 1953; 37(10):593–600. doi: 10.1136/bjo.37.10.593

575. Nordlöw W. Squint—the frequency of onset at different ages, and the incidence of some associated defects in a Swedish population. Acta Ophthalmol. 1964; 42:1015–1037. doi: 10.1111/j.1755-3768.1964.tb03667.x

576. Ntim-Amponsah CT, Ofosu-Amaah S. Prevalence of refractive error and other eye diseases in schoolchildren in the Greater Accra region of Ghana. J Pediatr Ophthalmol Strabismus. 2007; 44(5):294–297. doi: 10.3928/01913913-20070901-04

577. Ntizahuvye S, Onyango J. Prevalence of strabismus and the outcomes of its management among children attending Ruharo Eye Center, South Western Uganda. J Ophthalmol East Cent South Afr. (JOECSA) 2017; 21:13–15.

578. Nusz KJ, Mohney BG, Diehl NN. Female predominance in intermittent exotropia. Am J Ophthalmol. 2005; 140(3):546–547. doi: 10.1016/j.ajo.2005.03.026

579. Nwachukwu H, Onua AA, Adio AO. Risk factors of strabismus in children in a Southern Nigerian tertiary hospital. World J Opthalmol Vision Res. 2(4): 2019; 2(4):000541.

580. Nwachukwu H, Adio AO, Nathaniel GI, Musa KO. Pattern of Manifest Strabismus in Children Seen in a Tertiary Hospital in Rivers State, Nigeria. Int J Ophthalmol Vis Sci. 2021; 6(4): 209–214.

581. O’Connor AR, Stephenson TJ, Johnson A, Tobin MJ, Ratib S, Fielder AR. Strabismus in children of birth weight less than 1701 g. Arch Ophthalmol. 2002; 120(6):767–773. doi: 10.1001/archopht.120.6.767

582. Ohlsson J, Villarreal G, Sjöström A, Abrahamsson M, Sjöstrand J. Visual acuity, residual amblyopia and ocular pathology in a screened population of 12-13-year-old children in Sweden. Acta Ophthalmol Scand. 2001; 79(6):589–595. doi: 10.1034/j.1600-0420.2001.790609.x

583. Ohlsson J, Villarreal G, Sjöström A, Cavazos H, Abrahamsson M, Sjöstrand J. Visual acuity, amblyopia, and ocular pathology in 12- to 13-year-old children in Northern Mexico. J AAPOS. 2003; 7(1):47–53. doi: 10.1067/mpa.2003.S1091853102420113

584. Okada T. [Statistical study on strabismus.] Igaku kenkyu Acta medica. 1958; 29:1369–1385. Japanese

585. Okeigbemen VW, Momoh N. Ocular comorbidities in children with strabismus in Benin City. Sahel Med J. 2019; 22:13–17. doi: 10.4103/smj.smj_41_17

586. Olitsky SE, Nelson LB. 2005. Chapter 9: Strabismus Disorders. In: Harley’s Pediatric Ophthalmology, 5^th^ Edition, Nelson LB, Olitsky SE (eds). Lippincott Williams & Wilkins, Philadelphia; pp. 143–192.

587. Oliveira BFTd, Bigolin S, BarretoSouza M, Polati M. [Sensorial strabismus: a study of 191 cases.] Arq Bras Oftalmol. 2006; 69(1):71–74. Portuguese. doi: 10.1590/s0004-27492006000100014

588. Oliver M, Nawratzki I. Screening of pre-school children for ocular anomalies. II. Amblyopia. Prevalence and therapeutic results at different ages. Br J Ophthalmol. 1971; 55(7):467–471. doi: 10.1136/bjo.55.7.467

589. Olusanya BA, Ugalahi MO, Ayeni O, Fawole OI, Baiyeroju AM. Common Forms of Strabismus in a Tertiary Eye Clinic in Southwest Nigeria. Niger J Ophthalmol 2019; 27:62-67.

590. Omar R, Wan Abdul WMH, Knight VF. Status of visual impairment among indigenous (Orang Asli) school children in Malaysia. BMC Public Health. 2019; 19(Suppl 4):543. doi: 10.1186/s12889-019-6865-3

591. Omoti AE, Waziri-Erameh MJ. Pattern of neuro-ophthalmic disorders in a tertiary eye centre in Nigeria. Niger J Clin Pract. 2007; 10(2):147–151.

592. Ongaro L, Huerta-Sanchez E. A history of multiple Denisovan introgression events in modern humans. Nat Genet. 2024; 56(12):2612–2622. doi: 10.1038/s41588-024-01960-y

593. Oppenheimer S. Out-of-Africa, the peopling of continents and islands: tracing uniparental gene trees across the map. Philos Trans R Soc Lond B Biol Sci. 2012; 367(1590):770-784. doi: 10.1098/rstb.2011.0306

594. Ostadi Moghaddam H, Fotouhi A, KhabazKhoob M, Heravian J, Yekta A, Javaherforoushzadeh A. Prevalence of amblyopia in school children in Mashhad. Bina J Ophthalmol. 2008; 13(3):289–294.

595. Ostrer H, Skorecki K. The population genetics of the Jewish people. Hum Genet. 2013; 132(2):119–127. doi: 10.1007/s00439-012-1235-6

596. Oystreck DT, Lyons CJ. Comitant strabismus: Perspectives, present and future. Saudi J Ophthalmol. 2012; 26(3):265–270. doi: 10.1016/j.sjopt.2012.05.002

597. Pacheco P, Andrews S, Chaskel R. Strabismus and Quality of Life: The Impact of Surgical Intervention in Children and Adolescents in Colombia. World Soc Psychiatry. 2022; 4:159–163.

598. Padhye AS, Khandekar R, Dharmadhikari S, Dole K, Gogate P, Deshpande M. Prevalence of uncorrected refractive error and other eye problems among urban and rural school children. Middle East Afr J Ophthalmol. 2009; 16(2):69–74. doi: 10.4103/0974-9233.53864

599. Paduca A, Arnaut O, Cardaniuc C, et al. Epidemiology of childhood manifest strabismus in the Republic of Moldova. Strabismus. 2020; 28(3):128–135. doi: 10.1080/09273972.2020.1791912

600. Páez-Garza JH, Rangel-Padilla A, González-Godínez S, de la Rosa-Pacheco S. Strabismus in the north of Mexico: clinical characteristics in a pediatric population at public and private health institutions. [Estrabismo en el norte de México: características clínicas en pacientes pediátricos de instituciones de salud públicas y privadas]. Rev Mex Oftalmol. 2020; 94(3):99–104. Spanish

601. Pai ASI, Wang JJ, Samarawickrama C, et al. Prevalence and risk factors for visual impairment in preschool children. The Sydney paediatric eye disease study. Ophthalmology. 2011; 118(8):1495–1500. doi: 10.1016/j.ophtha.2011.01.027

602. Pai AS, Rose KA, Leone JF, et al. Amblyopia prevalence and risk factors in Australian preschool children. Ophthalmology. 2012; 119(1):138–144. doi: 10.1016/j.ophtha.2011.06.024

603. Pallin P. The biorbital angle and the position of the orbital walls at different ages: their significance to the appearance and disappearance of squint. Acta Ophthalmol. 1937; 15(4):389–405.

604. Pan CW, Chen X, Zhu H, et al. School-based assessment of amblyopia and strabismus among multiethnic children in rural China. Sci Rep. 2017; 7(1):13410. doi: 10.1038/s41598-017-13926-8

605. Panas F. [Paralysies oculaires motrices d’origine traumatique.] Arch Ophthal. 1899; 19:625–641. French

606. Pandey S, Pandey A, Bajracharya K, et al. An assessment of surgical outcome with the influencing factors of horizontal strabismus surgery. Asian J Med Sci. 2017; 8(5)54-57.

607. Pant M, Shrestha GS, Joshi ND. Ocular morbidity among street children in Kathmandu Valley. Ophthalmic Epidemiol. 2014; 21(6):356–361. doi: 10.3109/09286586.2014.964035

608. Parés JM, Duval M, Arnold LJ. New views on an old move: Hominin migration into Eurasia. Quat Int. 2013; 295:5–12. doi: 10.1016/j.quaint.2011.12.015

609. Pathai S, Cumberland PM, Rahi JS. Prevalence of and early- life influences on childhood strabismus. Findings from the Millennium Cohort Study. Arch Pediatr Adolesc Med. 2010; 164:250–257. doi: 10.1001/archpediatrics.2009.297

610. Patin E, Lopez M, Grollemund R, et al. Dispersals and genetic adaptation of Bantu-speaking populations in Africa and North America. Science. 2017; 356(6337):543-546. doi: 10.1126/science.aal1988

611. Paudel P, Ramson P, Naduvilath T, et al. Prevalence of vision impairment and refractive error in school children in Ba Ria - Vung Tau province, Vietnam. Clin Exp Ophthalmol. 2014; 42(3):217–226. doi: 10.1111/ceo.12273

612. Paul TO, Hardage LK. The heritability of strabismus. Ophthalmic Genet. 1994; 15(1):1–18. doi: 10.3109/13816819409056905

613. Peacock OSB. An analysis of ophthalmic cases in Nyasaland. Cent Afr J Med. 1959; 5:347–348.

614. Pearce E, Stringer C, Dunbar RI. New insights into differences in brain organization between Neanderthals and anatomically modern humans. Proc Biol Sci. 2013; 280(1758):20130168. doi: 10.1098/rspb.2013.0168

615. Péchereau A, Denis D, Speeg-Schatz C. [Strabisme-rapport Société Française d’Ophtalmologie,] éd. Elsevier Masson, Issy-les-Moulineaux, 2013, 546 pp.

616. Pergens E. [Les yeux et les fonctions visuelles des Congolais.] Janus. 1898; 2:459–463. French

617. Pérez-Padilla CA, Herrera-Lazo ZdC, Yabor-Labrada MdC, Villacres-Moya SS. Incidencia y características del estrabismo en pacientes de la provincia ecuatoriana de Tungurahua. [Incidence and characteristics of strabismus in patients from the Ecuadorian province of Tungurahua]. Mediciego. 2025; 31:e4132. Spanish

618. Pétursdóttir D, Holmström G, Larsson E. Strabismus, stereoacuity, accommodation and convergence in young adults born premature and screened for retinopathy of prematurity. Acta Ophthalmol. 2022; 100(3):e791–e797. doi: 10.1111/aos.14987

619. Phelps WL, Muir J. Anisometropia and strabismus. Amer Orthopt J. 1977; 27:131–133.

620. Pi LH, Chen L, Liu Q, et al. Prevalence of eye diseases and causes of visual impairment in school-aged children in Western China. J Epidemiol. 2012; 22(1):37–44. doi: 10.2188/jea.je20110063

621. Pickwell LD. Prevalence and management of divergence excess. Am J Optom Physiol Opt. 1979; 56(2):78–81. doi: 10.1097/00006324-197902000-00003

622. Pigassou-Albouy R, Garipuy J. [Comparison between internal and external strabismus]. Arch Ophtalmol Rev Gen Ophtalmol. 1963; 23:575–582. French

623. Pineles SL, Repka MX, Velez FG, et al. Prevalence of pediatric eye disease in the optumlabs data warehouse. Ophthalmic Epidemiol. 2022; 29(5):537–544. doi: 10.1080/09286586.2021.1971261

624. Plotnikov D, Shah RL, Rodrigues JN, et al. A commonly occurring genetic variant within the NPLOC4-TSPAN10-PDE6G gene cluster is associated with the risk of strabismus. Hum Genet. 2019; 138(7):723–737. doi: 10.1007/s00439-019-02022-8

625. Podgor MJ, Remaley NA, Chew E. Associations between siblings for esotropia and exotropia. Arch Ophthalmol. 1996; 114(6):739–744. doi: 10.1001/archopht.1996.01100130731018

626. Pokharel GP, Negrel AD, Munoz SR, Ellwein LB. Refractive error study in children: results from Mechi Zone, Nepal. Am J Ophthalmol. 2000; 129(4):436–444. doi: 10.1016/s0002-9394(99)00453-5

627. Popović-Beganović A, Zvorničanin J, Vrbljanac V, Zvorničanin E. The prevalence of refractive errors and visual impairment among school children in Brčko district, Bosnia and Herzegovina. Semin Ophthalmol. 2018; 33(7-8):858–868. doi: 10.1080/08820538.2018.1539182

628. Power B, Murphy M, Stokes J. The impact of strabismus surgery on Irish adults. Br Ir Orthopt J. 2018; 14(1):6–10. doi: 10.22599/bioj.107

629. Prabhu AV, Ve RS, Talukdar J, Chandrasekaran V. Prevalence of visual impairment in school-going children among the rural and urban setups in the Udupi district of Karnataka, India: A cross-sectional study. Oman J Ophthalmol. 2019; 12:145–149. doi: 10.4103/ojo.OJO_190_2018

630. Premsenthil M, Manju R, Thanaraj A, Rahman SA, Kah TA. The screening of visual impairment among preschool children in an urban population in Malaysia; the Kuching pediatric eye study: a cross sectional study. BMC Ophthalmol. 2013; 13:16. doi: 10.1186/1471-2415-13-16

631. Preslan MW, Novak A. Baltimore Vision Screening Project. Ophthalmology. 1996; 103(1):105–109. doi: 10.1016/s0161-6420(96)30753-7

632. Preslan MW, Novak A. Baltimore Vision Screening Project. Phase 2. Ophthalmology. 1998; 105(1):150-153. doi: 10.1016/s0161-6420(98)91813-9

633. Prieto-Díaz J, Souza-Dias C. [Estrabismo.] 5ed. Ediciones Cientificas Argentinas, 2005.

634. Promelle V, Bremond-Gignac, Milazzo S. Epidemiology of patients undergoing strabismus surgery at adult age: retrospective study of 221 patients. Acta Ophthalmol. 2013; 91:s252.

635. Pryor HB. Objective measurement of interpupillary distance. Pediatrics. 1969; 44(6):973–977.

636. Putri P, Julita. Profil Strabismus Horizontal di RSUP Dr. M Djamil Padang Januari – Desember 2017. Jurnal Kesehatan Andalas. 2020; 9(1):83-87. Indonesian

637. Qanat AS, Alsuheili A, Alzahrani AM, Faydhi AA, Albadri A, Alhibshi N. Assessment of Different Types of Strabismus Among Pediatric Patients in a Tertiary Hospital in Jeddah. Cureus; 2020; 12(12):e11978. doi: 10.7759/cureus.11978

638. Quere MA, Mehel E. 1995. [Epidemiologie. In: Les Strabismes de L’Adolescent et de L’Adulte.] V. 3.1. Cahiers de Sensorio-Motricite XXe Colloque, 1995. pp 3-7. http://www.strabisme.net/strabologie/Telechargement/files/SAA.pdf

639. Rah SH, Jun HS, Kim SH. [An epidemiological survey of strabismus among school-children in Korea.] J Korean Ophthalmol Soc. 1997; 38(12):2195–2199. Korean

640. Rahmania N, van Rompay T, Morfeq H, Promelle V, Milazzo S. Surgical treatment of pediatric strabismus (PS): series of 148 patients. Acta Ophthalmol. 2016; 94:S256.

641. Rajasekaran R, Kumari RM, Balagopal A, Ramesh PV, Mohan K. Prevalence of various types of strabismus among patients attending a tertiary eye care hospital at Tiruchirappalli. J Evolution Med Dent Sci. 2018; 7(52):5484–5487.

642. Rajavi Z, Sabbaghi H, Baghini AS, Yaseri M, Moein H, Akbarian S, Behradfar N, Hosseini S, Rabei HM, Sheibani K. Prevalence of amblyopia and refractive errors among primary school children. J Ophthalmic Vis Res. 2015; 10(4):408–416. doi: 10.4103/2008-322X.176909

643. Rajavi Z, Khorrame Z, Ashrafi S. The effects of refractive status on the outcomes of strabismus surgery in patients with esotropia. BMC Ophthalmol. 2024; 24(1):271. doi: 10.1186/s12886-024-03531-5

644. Rajesh AE, Davidson O, Lacy M, et al. Race, Ethnicity, Insurance, and Population Density Associations with Pediatric Strabismus and Strabismic Amblyopia in the IRIS® Registry. Ophthalmology. 2023; 130(10):1090–1098. doi: 10.1016/j.ophtha.2023.06.008

645. Rantanen A, Tommila V. Prevalence of strabismus in Finland. Acta Ophthalmol. 1971; 49:506–509.

646. Rantakallio P, Krause U, Krause K. The use of the ophthalmological services during the preschool age, ocular findings and family background. J Pediatr Ophthalmol Strabismus. 1978; 15(4):253–258. doi: 10.3928/0191-3913-19780701-16

647. Rao GN, Sabnam S, Pal S, Rizwan H, Thakur B, Pal A. Prevalence of ocular morbidity among children aged 17 years or younger in the eastern India. Clin Ophthalmol. 2018; 12:1645–1652. doi: 10.2147/OPTH.S171822

648. Rasmussen C. [Nachuntersuchung der Schieloperierten an der Augenklinik des Kommunehospitals.] Verhandl Ophthalmol Ges Kopenhagen. 1920/21; 9:1-24. German

649. Rasmussen F, Thorén K, Caines E, Andersson J, Tynelius P. Suitability of the Lang II random dot stereotest for detecting manifest strabismus in 3-year-old children at child health centres in Sweden. Acta Paediatr. 2000; 89(7):824–829.

650. Rasmussen M, Guo X, Wang Y, et al. An Aboriginal Australian genome reveals separate human dispersals into Asia. Science. 2011; 334(6052):94-98. doi: 10.1126/science.1211177

651. Razmjoo H, Haj-Yahya Y, Javic E, Abtahi SM, Mehrabi-Koushki A. Study of 100 children with strabismus admitted to Feyz hospital, Isfahan, Iran, in 2012-2013. J Isfahan Med School. 2015; 32(313):2149-2156.

652. Reddy SC. Ocular morbidity and colour blindness among school children in Kakinada. The Antiseptic. 1987; 84(5):611–616.

653. Reddy SC, Tajunisah I, Low KP, Karmila AB. Prevalence of eye diseases and visual impairment in urban population - a study from University of Malaya medical centre. Malays Fam Physician. 2008; 3(1):25-28.

654. Regoda V, Sefic-Kasumovic S. Role of hereditary factors in strabismus occurrence. Med Arch. 2012; 66(6):418–419. doi: 10.5455/medarh.2012.66.418-419

655. Reich D, Thangaraj K, Patterson N, Price AL, Singh L. Reconstructing Indian population history. Nature. 2009; 461:489–494. doi: 10.1038/nature08365

656. Reich D, Patterson N, Campbell D, et al. Reconstructing Native American population history. Nature. 2012; 488(7411):370-374. doi: 10.1038/nature11258

657. Repka MX, Friedman DS, Katz J, Ibironke J, Giordano L, Tielsch JM. The prevalence of ocular structural disorders and nystagmus among preschool-aged children. J AAPOS. 2012a; 16(2):182–184. doi: 10.1016/j.jaapos.2011.12.156

658. Repka MX, Yu F, Coleman A. Strabismus among aged fee-for-service Medicare beneficiaries. J AAPOS. 2012b; 16(6):495–500. doi: 10.1016/j.jaapos.2012.07.010

659. Repka MX. A close look at pediatric eye disease. Ophthalmology. 2014; 121(3):617–618. doi: 10.1016/j.ophtha.2013.12.038

660. Repka MX, Lum F, Burugapalli B. Strabismus, strabismus surgery, and reoperation rate in the United States: Analysis from the IRIS registry. Ophthalmology. 2018; 125(10)1646-1653. doi: 10.1016/j.ophtha.2018.04.024

661. Reza BM, Reza MM, Mojgan K, Hassan LM. Pediatric strabismus: prevalence and surgical outcomes in Yazd, Iran. Guoji Yanke Zazhi (Int Eye Sci.) 2013; 13(8):1521–1524.

662. Ribeiro GdeB, Bach AG, Faria CM, Anastásia S, Almeida HC. Quality of life of patients with strabismus. Arq Bras Oftalmol. 2014; 77(2):110–113. doi: 10.5935/0004-2749.20140027

663. Ricci B, Coppola G, Ricci V, Ziccardi L. Nationwide study of hospitalization and surgical treatment for childhood strabismus in Italy between 1999 and 2004. Int Ophthalmol. 2009; 29(3):153–156. doi: 10.1007/s10792-008-9209-3

664. Rizwan N, Nizamani N, Talpur A, Nizamani YM, Channa SN. Frequency of Refractive Error in Patients with Squint. Ann Pak Inst Med Sci. 2024; 21(3):256–260. doi: 10.48036/apims.v20i3.1099

665. Robaei D, Rose KA, Kifley A, Cosstick M, Ip JM, Mitchell P. Factors associated with childhood strabismus: findings from a population-based study. Ophthalmology. 2006a; 113(7):1146–1153. doi: 10.1016/j.ophtha.2006.02.019

666. Robaei D, Kifley A, Mitchell P. Factors associated with a previous diagnosis of strabismus in a population-based sample of 12-year-old Australian children. Am J Ophthalmol. 2006b; 142(6):1085–1088. doi: 10.1016/j.ajo.2006.06.053

667. Robinson B, Bobier WR, Martin E, Bryant L. Measurement of the validity of a preschool vision screening program. Am J Public Health. 1999; 89(2):193–198. doi: 10.2105/ajph.89.2.193

668. Rocha MNAM, Sanches A, Pessoa FF, et al. Clinical forms and risk factors associated with strabismus in visual binocularity. Rev Bras Oftalmol. 2016; 75(1):34–39. doi: 10.5935/0034-7280.20160008

669. Rodrigues da Costa D, Debert I, Susanna FN, Falabreti JG, Polati M, Susanna Júnior R. Vision for the Future Project: Screening impact on the prevention and treatment of visual impairments in public school children in São Paulo City, Brazil. Clinics (Sao Paulo). 2021; 76:e3062. doi: 10.6061/clinics/2021/e3062

670. Rodríguez MA, Castro González M. [Visual health of schoolchildren in Medellin, Antioquia, Colombia]. Bol Oficina Sanit Panam. 1995; 119(1):11–14. Spanish

671. Rohr JTD, Isaac CR, Correia CdS. Epidemiology of strabismus surgery in a public hospital of the Brazilian Federal District. Rev Bras Oftalmol. 2017; 76(5):250–254. doi: 10.5935/0034-7280.20170052

672. Romanchuk KG, Dotchin SA, Zurevinsky J. The natural history of surgically untreated intermittent exotropia-looking into the distant future. J AAPOS. 2006; 10(3):225–231. doi: 10.1016/j.jaapos.2006.02.006

673. Romano PE. Editorial: The relationship between light and exotropia. Binocular Vision Q. 1990; 5:11-12.

674. Rose K, Younan C, Morgan I, Mitchell P. Prevalence of undetected ocular conditions in a pilot sample of school children. Clin Exp Ophthalmol. 2003; 31(3):237–240. doi: 10.1046/j.1442-9071.2003.00636.x

675. Rosenberg JB, Tepper OM, Medow NB. Strabismus in craniosynostosis. J Pediatr Ophthalmol Strabismus. 2013; 50(3):140-148. doi: 10.3928/01913913-20121113-02

676. Rosner J, Rosner J. Comparison of visual characteristics in children with and without learning difficulties. Am J Optom Physiol Opt. 1987; 64(7):531–533. doi: 10.1097/00006324-198707000-00008

677. Royal Australian College of Ophthalmologists. National Trachoma & Eye Health Program. 217 pp, Promail Printing Group Pty. Ltd., Artarmon, N.S.W., Sydney, 1980. ISBN 0 9594785 0 7

678. Rüssmann W, König U, Schlimbach K, Pawlowska-Seyda D, Wirbatz B. [Refractive errors, strabismus and amblyopia in pre-school screening--experiences using a vision test in kindergarten]. Offentl Gesundheitswes. 1990; 52(2):77–84. German

679. Saatchi M, Sadeghi T, Peyman M. Amblyopia and Strabismus in Iran. Arch Persian Ophthalmol. 2025; 6(1):264-275. http://apo.tums.ac.ir/article-1-50-en.html

680. Sah SP, Sharma IP, Chaudhry M, Saikia M. Health-Related Quality of Life (HRQoL) in Young Adults with Strabismus in India. J Clin Diagn Res. 2017; 11(2):NC01-NC04. doi: 10.7860/JCDR/2017/24541.9389

681. Said JE, Abdulkader JE. Myectomy technique for horizontal squint surgery. EC Ophthalmology. 2016; 4.2:487–491.

682. Salman MS. Pediatric eye diseases among children attending outpatient eye department of Tikrit Teaching Hospital. Tikrit J Pharm Sci. 2010; 7(1):95–103.

683. Salomão SR, Cinoto RW, Berezovsky A, et al. Prevalence and causes of visual impairment in low-middle income school children in Sao Paulo, Brazil. Invest Ophthalmol Vis Sci. 2008; 49(10):4308–4313. doi: 10.1167/iovs.08-2073

684. Sandfeld L, Weihrauch H, Tubaek G, Mortzos P. Ophthalmological data on 4.5- to 7-year-old Danish children. Acta Ophthalmol. 2018; 96(4):379–383. doi: 10.1111/aos.13650

685. Sanfilippo PG, Hammond CJ, Staffieri SE, et al. Heritability of strabismus: genetic influence is specific to eso-deviation and independent of refractive error. Twin Res Hum Genet. 2012; 15(5):624–630. doi: 10.1017/thg.2012.22

686. Sarosh R, Khan A, Rashid O, Hakak B, un Nisa A, Sarosh P. Profile of strabismus at a tertiary care hospital in Kashmir. Int J Contemp Med Res. 2018; 5(6)F4-F7.

687. Satou T, Takahashi Y, Ito M, Mochizuki H, Niida T. Evaluation of visual function in preschool-age children using a vision screening protocol. Clin Ophthalmol. 2018; 12:339–344. doi: 10.2147/OPTH.S160288

688. Saxena A, Nema N, Deshpande A. Prevalence of refractive errors in school-going female children of a rural area of Madhya Pradesh, India. J Clin Ophthalmol Res. 2019; 7(2):45–49.

689. Saxena R, Singh D, Gantyala SP, Aggarwal S, Sachdeva MM, Sharma P. Burden of ocular motility disorders at a tertiary care institution: A case to enhance secondary level eye care. Indian J Community Med. 2016; 41(2):103–107. doi: 10.4103/0970-0218.177523

690. Schaal LF, Schellini SA, Pesci LT, Galindo A, Padovani CR, Corrente JE. The prevalence of strabismus and associated risk factors in a Southeastern region of Brazil. Semin Ophthalmol. 2018; 33(3):357–360. doi: 10.1080/08820538.2016.1247176

691. Scheiman M, Gallaway M, Coulter R, et al. Prevalence of vision and ocular disease conditions in a clinical pediatric population. J Am Optom Assoc. 1996; 67(4):193–202.

692. Schenk H, Haydn M. [Experiences with the Rodenstock vision test device for children, (serial examination in the Central Children’s Home of the City of Vienna)]. Klin Monbl Augenheilkd. 1969; 154(5):739–746. German

693. Schildwächter K. [Experience with screening the R5 device in Hessen.] Arbeitskreis Schielb. 1972; 4:102-112. German

694. Schimiti RB, Costa VP, Gregui MJF, Kara-José N, Temporini ER. Prevalence of refractive errors and ocular disorders in preschool and schoolchildren of Ibiporã-PR, Brazil (1989 to 1996). Arq Bras Oftalmol. 2001; 64(5):379-384. doi: 10.1590/S0004-27492001000500002

695. Schleich G. [Die Augen der Schüler und Schülerinnen der Tübinger Schulen.] Int Arch Schulhygiene. 1905; 1:19–27. German

696. Schlossman A, Priestley BS. Role of heredity in etiology and treatment of strabismus. AMA Arch Ophthalmol. 1952; 47(1):1–20. doi: 10.1001/archopht.1952.01700030004001

697. Schlossman A, Boruchoff SA. Correlation between physiologic and clinical aspects of exotropia. Am J Ophthalmol. 1955; 40(1):53–64. doi: 10.1016/0002-9394(55)92122-x

698. Schubert EW, Henriksson KM, McNeil TF. A prospective study of offspring of women with psychosis: visual dysfunction in early childhood predicts schizophrenia-spectrum disorders in adulthood. Acta Psychiatr Scand. 2005; 112(5):385–393. doi: 10.1111/j.1600-0447.2005.00584.x

699. Schulek W. [Symptomatologie und Aetiologie des Strabismus divergens.] Klin Monatsbl Augenheilkd. 1871; 9:407–421. German

700. Schuster AK, Elflein HM, Pokora R, Urschitz MS. Kindlicher Strabismus in Deutschland: Prävalenz und Risikogruppen: Ergebnisse der KiGGS-Studie [Childhood strabismus in Germany: Prevalence and risk groups: Results of the KiGGS survey]. Bundesgesundheitsblatt Gesundheitsforschung Gesundheitsschutz. 2017; 60(8):849–855. German. doi: 10.1007/s00103-017-2578-x

701. Schuster AK, Elflein HM, Pokora R, Schlaud M, Baumgarten F, Urschitz MS. Health-related quality of life and mental health in children and adolescents with strabismus - results of the representative population-based survey KiGGS. Health Qual Life Outcomes. 2019; 17(1):81. doi: 10.1186/s12955-019-1144-7

702. Schütte E, Groten H, Leymann J, Lizin F. [Ophthalmic and orthoptic investigations in the kindergarten (author’s transl)]. Klin Monbl Augenheilkd. 1976; 168(4):584–590. German

703. Schweigger C. [Klinische Untersuchungen über das Schielen: Eine Monographie.] Berlin: Verlag August Hirschwald, 1881, 152 pp.

704. Scobee RG. Esotropia. Incidence, etiology, and results of therapy. Am J Ophthalmol. 1951; 34(6):817–833.

705. See LC, Song HS, Ku WC, Lee JS, Liang YS, Shieh WB. Neglect of childhood strabismus: Keelung Ann-Lo Community ocular survey 1993-1995. Changgeng Yi Xue Za Zhi. 1996; 19(3):217–224.

706. Semanyenzil SE, Karimurio J, Nzayirambaho M. Prevalence and pattern of refractive errors in high schools in Nyarugenge district. Rwanda Med J. 2015; 72(3):8–13.

707. Sethee SK, FitzGerald DE, Krumholtz I. Exotropia in a pediatric population less than six years of age. J Behav Optometr. 2003; 14(6):149–157.

708. Sethi S, Khan MD. Pediatric ophthalmic disorders. J Postgrad Med Inst. 2001; 15(2):144–150.

709. Sethi S, Sethi MJ, Hussain I, Kundi NK. Causes of amblyopia in children coming to ophthalmology out patient department Khyber Teaching Hospital, Peshawar. J Pak Med Assoc. 2008; 58(3):125–128.

710. Sethi J, Shah A, Malik R, Rahil N. Gender differences & reasons of delay in presentation of childhood squint. Ophthalmol Update. 2012; 10(1):107–109.

711. Sethi S, Aggarwal R, Reddy VS, Dabas R. Pattern of ocular morbidity among children referred through a national screening program in a tertiary hospital in Northern India. Ann Int Med Dent Res. 2017; 3(1):3–8.

712. Shaffi M, Bejiga A. Common eye diseases in children of rural community in Goro district, Central Ethiopia. Ethiop J Health Dev. 2005; 19(2):148–152.

713. Shah MA, Khan S, Mohammad S. Presentation of childhood squint. J Postgrad Med Inst. 2002; 16:206–210.

714. Shah A, Sethi S, Ilyas O, Shah Z. The frequency of amblyopia & results of squint surgery in patients admitted in Khyber Teaching Hospital, Peshawar. Ophthalmology Update. 2013; 11:109–111.

715. Shahid E, Shaikh A, Aziz S, Rehman A. Frequency of Ocular Diseases in Infants at a Tertiary Care Hospital. Korean J Ophthalmol. 2019; 33(3):287–293. doi: 10.3341/kjo.2017.0142

716. Shaikh SP, Aziz TM. Pattern of eye diseases in children of 5-15 years at Bazzertaline area (South Karachi) Pakistan. J Coll Physicians Surg Pak. 2005; 15(5):291–294.

717. Shaikh TS, Shaikh SP, Shaikh WA. Pattern of horizontal squint presentation in Pediatric Eye Department at Civil Hospital Karachi. J Bahria Univ Med Dental Coll. 2014; 4(2):57–61.

718. Shaikh TS, Shaikh SP, Dareshani S, Shaikh WA. PATTERN OF SQUINT PRESENTATION IN PAEDIATRIC EYE DEPARTMENT AT CIVIL HOSPITAL KARACHI. Med Channel. 2015; 21(14):16–21.

719. Shankar J, Kaye S. Trends in squint surgical activity. Brit J Ophthalmol. 2003; 87(7):918. doi: 10.1136/bjo.87.7.918

720. Shapira Y, Machluf Y, Mimouni M, Chaiter Y, Mezer E. Amblyopia and strabismus: trends in prevalence and risk factors among young adults in Israel. Brit J Ophthalmol. 2018; 102(5):659–666. doi: 10.1136/bjophthalmol-2017-310364

721. Sharma RP, Gupta RK, Krishna G, Srivastava JP, Saxena SC, Srivastava VK, Sharma SN, Singh VP, Chandra S. A profile of ocular diseases among school children in slum areas of kanpur. Indian J Commun Health. 1995; 8(2, 3):28-30.

722. Sharma A, Maitreya A, Semwal J, Bahadur H. Ocular morbidity among school children in Uttarakhand: Himalayan state of India. Ann Trop Med Public Health. 2017; 10:149–153.

723. Sherief ST, Muhe LM, Mekasha A, Demtse A, Ali A. Prevalence and causes of ocular disorders and visual impairment among preterm children in Ethiopia. BMJ Paediatr Open. 2024; 8(1):e002317. doi: 10.1136/bmjpo-2023-002317

724. Sherpa D, Panta CR, Joshi N. Ocular morbidity among primary school children of Dhulikhel, Nepal. Nepal J Ophthalmol. 2011; 3(2):172–176. doi: 10.3126/nepjoph.v3i2.5272

725. Sherpa D. Ocular morbidity among primary school children. J Chitwan Med Coll. 2014; 4(2):32–34. doi: 10.3126/nepjoph.v3i2.5272

726. Shimauti AT, Pesci LT, Sousa RL, Padovani CR, Schellini SA. [Strabismus: Detection in a population-based sample and associated demographic factors.] Arq Bras Oftalmol. 2012; 75(2):92–96. Portuguese. doi: 10.1590/s0004-27492012000200004

727. Shrestha RK, Joshi MR, Ghising R, Pradhan P, Shakya S, Rizyal A. Ocular morbidity among children studying in private schools of Kathmandu valley: A prospective cross sectional study. Nepal Med Coll J. 2006; 8(1):43–46.

728. Shrestha RK, Joshi MR, Ghising R, Rizyal A. Ocular morbidity among children attending government and private schools of Kathmandu valley. J Nepal Med Assoc. (JNMA) 2011; 51(184):182–188.

729. Shrote VK, Thakre SS, Lanjewar AG, Brahmapurkar KP, Khakse GM. Ocular morbid conditions in the rural area of central India. Int J Collab Res Intern Med Public Health. 2012; 4(9):1692–1702.

730. Siddiqui SN, Asif M, Iqbal S, Shah MA, Habiba U, Rabia M. Frequency of Different Types of Manifest Strabismus in Children. Al-Shifa J Ophthalmol. 2017; 13(2)86-92.

731. Sigronde L, Blanc J, Aho S, Pallot C, Bron AM, Creuzot-Garcher C. Evaluation of the Spot Vision Screener in comparison with the orthoptic examination in visual screening in 3-5 year-old schoolchildren. J Fr Ophtalmol. 2020; 43(5):411–416. doi: 10.1016/j.jfo.2019.10.006

732. Simpson A, Kirkland C, Silva PA. Vision and eye problems in seven year olds: a report from the Dunedin Multidisciplinary Health and Development Research Unit. NZ Med J. 1984; 97:445–449.

733. Singh H. Pattern of ocular morbidity in school children in central India. Natl J Community Med. 2011; 2(3):429–431.

734. Singh V, Malik K, Malik VK, Jain K. Prevalence of ocular morbidity in school going children in West Uttar Pradesh. Indian J Ophthalmol. 2017; 65:500–508. doi: 10.4103/ijo.IJO_676_15

735. Singh A, Chawla O, Verma R, et al. Refractive errors and concomitant strabismus in children and adolescents: A hospital based observational study. Delhi J Ophthalmol. 2021; 32(2):24–29.

736. Skeller E. Anthropological and ophthalmological studies on the Angmagssalik Eskimos. Meddr Grønland. 1954; 107(4):1–231.

737. Skoglund P, Reich D. A genomic view of the peopling of the Americas. Curr Opin Genet Dev. 2016; 41:27–35. doi: 10.1016/j.gde.2016.06.016

738. Skoglund P, Posth C, Sirak K, et al. Genomic insights into the peopling of the Southwest Pacific. Nature. 2016; 538:510–513. doi: 10.1038/nature19844

739. Slater AM, Findlay JM. The measurement of fixation position in the newborn baby. J Exp Child Psychol. 1972; 14(3):349–364. doi: 10.1016/0022-0965(72)90056-2

740. Smith LK, Thompson JR, Woodruff G, Hiscox F. Factors affecting treatment compliance in amblyopia. J Pediatr Ophthalmol Strabismus. 1995; 32(2):98–101. doi: 10.3928/0191-3913-19950301-09

741. Sondhi N, Archer SM, Helveston EM. Development of normal ocular alignment. J Pediatr Ophthalmol Strabismus. 1988; 25(5):210–211. doi: 10.3928/0191-3913-19880901-03

742. Sorsby A, Sheridan M, Leary GA, Benjamin B. Vision, visual acuity, and ocular refraction of young men: findings in a sample of 1,033 subjects. Br Med J. 1960; 1(5183):1394-1398. doi: 10.1136/bmj.1.5183.1394

743. Speeg-Schatz C, Lobstein Y, Burget M, Berra O, Riehl C, Hoffmann C. A review of preschool vision screening for strabismus and amblyopia in France: 23 years experience in the Alsace region. Binocul Vis Strabismus Q. 2004; 19(3):151–158.

744. Speidel K. [Die Augen der Theologiestudierenden in Tübingen. Untersuchungen aus der Tübinger Universitätsaugenklinik. Int Arch Schulhygiene.] 1905; 1:28–52. German

745. Spencer R. Long-term visual outcomes in extremely low-birth-weight children (an American Ophthalmological Society thesis). Trans Am Ophthalmol Soc. 2006; 104:493-516.

746. Squint. Editorial. Brit Med J. 1974; 3:430-431.

747. Stayte M, Johnson A, Wortham C. Ocular and visual defects in a geographically defined population of 2-year-old children. Brit J Ophthalmol. 1990; 74(8):465–468. doi: 10.1136/bjo.74.8.465

748. Stayte M, Reeves B, Wortham C. Ocular and vision defects in preschool children. Brit J Ophthalmol. 1993; 77(4):228–232. doi: 10.1136/bjo.77.4.228

749. Sternberg A, Feher M. Five years experience with divergent squint. Ophthalmologica. 1958; 136(1):1–12. doi: 10.1159/000303386

750. Sterne JA, Egger M. Funnel plots for detecting bias in meta-analysis: guidelines on choice of axis. J Clin Epidemiol. 2001; 54(10):1046–1055. doi: 10.1016/s0895-4356(01)00377-8

751. Sterne JAC, Harbord RM. Funnel plots in meta-analysis. Stata J. 2004; 4(2):127–141.

752. Sterne JA, Sutton AJ, Ioannidis JP, et al. Recommendations for examining and interpreting funnel plot asymmetry in meta-analyses of randomized controlled trials. Brit Med J. 2011; 343:d4002. doi: 10.1136/bmj.d4002

753. Stidwill D. Epidemiology of strabismus. Ophthalmic Physiol Opt. 1997; 17:536–539.

754. Strazzi A. [La corrispondenza retinica nello strabismo funzioniale.] Boll ocul. 1950; 29:517–526. Italian

755. Streiner DL, Patten SB, Anthony JC, Cairney J. Has ‘lifetime prevalence’ reached the end of its life? An examination of the concept. Int J Methods Psychiatr Res. 2009; 18(4):221–228. doi: 10.1002/mpr.296

756. Stutterheim NA. The convergence of human binocular vision. Brit J Ophthalmol. 1932; 16(1):20–30. doi: 10.1136/bjo.16.1.20

757. Stutterheim NA. Eye Strain and Convergence. HK Lewis & Co, London, 1937, 89 pp.

758. Sweeting MJ, Sutton AJ, Lambert PC. What to add to nothing? Use and avoidance of continuity corrections in meta-analysis of sparse data. Stat Med. 2004; 23(9):1351–1375. doi: 10.1002/sim.1761

759. Tabansi IC, Anochie KE, Nkanginieme, Pedro-Egbe CN. Simple vision screening in lower primary school children in Port Harcourt city. Port Harcourt Med J. 2009; 3:239–246. doi: 10.4314/phmedj.v3i3.45268

760. Taha AO, Ibrahim SM. Prevalence of manifest horizontal strabismus among basic school children in Khartoum City, Sudan. Sudanese J Ophthalmol. 2015; 7:53–57. doi: 10.4103/1858-540X.169437

761. Taira Y, Matsuo T, Yamane T, Hasebe S, Ohtsuki H. Clinical features of comitant strabismus related to family history of strabismus or abnormalities in pregnancy and delivery. Jpn J Ophthalmol. 2003; 47(2):208–213. doi: 10.1016/s0021-5155(02)00685-8

762. Takemoto Y, Ota E, Yoneoka D, Mori R, Takeda S. Japanese secular trends in birthweight and the prevalence of low birthweight infants during the last three decades: A population-based study. Sci Rep. 2016; 6:31396. doi: 10.1038/srep31396

763. Tananuvat N, Manassakorn A, Worapong A, Kupat J, Chuwuttayakorn J, Wattananikorn S. Vision screening in schoolchildren: two years results. J Med Assoc Thai. 2004; 87(6):679–684.

764. Tang SM, Chan RY, Bin Lin S, Rong SS, Lau HH, Lau WW, Yip WW, Chen LJ, Ko ST, Yam JC. Refractive errors and concomitant strabismus: A systematic review and meta-analysis. Sci Rep. 2016; 6:35177. doi: 10.1038/srep35177

765. Tattersall I, Schwartz JH. Extinct Humans. Boulder, Colorado: Westview Press, 2000.

766. Taylor J. Letter: Retinitis pigmentosa and squint in Africans. Br Med J. 1974; 4:529. doi: 10.1136/bmj.4.5943.529

767. Taylor HR. Prevalence and causes of blindness in Australian aborigines. Med J Aust. 1980; 1(2):71–76. doi: 10.5694/j.1326-5377.1980.tb134630.x

768. Tegegne MM, Fekadu SA, Assem AS. Prevalence of Strabismus and Its Associated Factors Among School-Age Children Living in Bahir Dar City: A Community-Based Cross-Sectional Study. Clin Optom (Auckl). 2021; 13:103–112. doi: 10.2147/OPTO.S300124

769. Teoh GH, Yow CS. Prevalence of squints and visual defects in Malaysian primary one school children. Med J Malaysia. 1982; 37(4):336–337.

770. Thevi T, Basri M, Reddy SC. Prevalence of eye diseases and visual impairment among the rural population - a case study of Temerloh hospital. Malays Fam Physician. 2012; 7(1):6–10.

771. Thomson WE. Some considerations regarding the aetiology, the incidence and the course of concomitant convergent strabismus which have a bearing upon its treatment. Trans Ophthal Soc UK. 1924; 44:238–252.

772. Thorn F, Gwiazda J, Cruz AA, Bauer JA, Held R. The development of eye alignment, convergence, and sensory binocularity in young infants. Invest Ophthalmol Vis Sci. 1994; 35(2):544–553.

773. Tinley C, Grötte R. Comitant horizontal strabismus in South African black and mixed race children--a clinic-based study. Ophthalmic Epidemiol. 2012; 19(2):89–94. doi: 10.3109/09286586.2011.645107

774. Tishkoff SA, Reed FA, Friedlaender FR, et al. The genetic structure and history of Africans and African Americans. Science. 2009; 324(5930):1035-1044. doi: 10.1126/science.1172257

775. Toppel L. [Epidemiology of juvenile functional disorders of the eye]. Fortschr Med. 1978; 96(20):1087–1094. German

776. Torp-Pedersen T, Boyd HA, Poulsen G, et al. Perinatal risk factors for strabismus. Int J Epidemiol. 2010a; 39(5):1229–1239. doi: 10.1093/ije/dyq092

777. Torp-Pedersen T, Boyd HA, Poulsen G, et al. In-utero exposure to smoking, alcohol, coffee, and tea and risk of strabismus. Am J Epidemiol. 2010b; 171(8):868–875. doi: 10.1093/aje/kwq010

778. Torp-Pedersen T, Boyd HA, Skotte L, et al. Strabismus incidence in a Danish population-based cohort of children. JAMA Ophthalmol. 2017; 135(10):1047–1053. doi: 10.1001/jamaophthalmol.2017.3158

779. Toufeeq A, Oram AJ. School-entry vision screening in the United Kingdom: practical aspects and outcomes. Ophthalmic Epidemiol. 2014; 21(4):210–216. doi: 10.3109/09286586.2014.906627

780. Toyota T, Yoshitsugu K, Ebihara M, et al. Association between schizophrenia with ocular misalignment and polyalanine length variation in PMX2B. Hum Mol Genet. 2004; 13(5):551–561. doi: 10.1093/hmg/ddh047

781. Traboulsi EI, Cimino H, Mash C, Wilson R, Crowe S, Lewis H. Vision First, a program to detect and treat eye diseases in young children: the first four years. Trans Am Ophthalmol Soc. 2008; 106:179–185; discussion 185-186.

782. Tremblay M, Vézina H. New estimates of intergenerational time intervals for the calculation of age and origins of mutations. Am J Hum Genet. 2000; 66(2):651–658. doi: 10.1086/302770

783. Tsao WS, Hsieh HP, Chuang YT, Sheu MM. Ophthalmologic abnormalities among students with cognitive impairment in eastern Taiwan: The special group with undetected visual impairment. J Formos Med Assoc. 2017; 116(5):345–350. doi: 10.1016/j.jfma.2016.06.013

784. Tubing K, Patton T, Usharani L, et al. Study of concomitant strabismus amongst the ethnic population of Manipur. IOSR J Dent Med Sci. 2014; 13(1):23–28.

785. Tuppurainen K, Herrgård E, Martikainen A, Mäntyjärvi M. Ocular findings in prematurely born children at 5 years of age. Graefes Arch Clin Exp Ophthalmol. 1993; 231(5):261–266. doi: 10.1007/BF00919102

786. Turaçli ME, Aktan SG, Dürük K. Ophthalmic screening of school children in Ankara. Eur J Ophthalmol. 1995; 5(3):181–186. doi: 10.1177/112067219500500307

787. Tyser PA, Letchworth TW. A study in visual defects in young children. Br Med J. 1949; 2:1022–1023. doi: 10.1136/bmj.2.4635.1022

788. Uddin M, Omar R, Feizal V, Alam K. Ocular morbidity among preschool children in urban area of Chittagong in Bangladesh. Guoji Yanke Zazhi (Int Eye Sci.) 2017;17(1):16–20. doi: 10.3980/j.issn.1672-5123.2017.1.04

789. Urmil AC, Dutta PK, Ahmed KA, Ghosh AK. A PREVALENCE STUDY OF EYE DISEASES AMONG CHILDREN IN A SCHOOL IN PUNE. Indian J Commun Med. 1988; 13(3):134–141.

790. Urrets-Zavalía A, Solares-Zamora J, Olmos HR. Anthropological studies on the nature of cyclovertical squint. Br J Ophthalmol. 1961; 45(9):578–596. doi: 10.1136/bjo.45.9.578

791. Uzma N, Kumar BS, Khaja Mohinuddin Salar BM, Zafar MA, Reddy VD. A comparative clinical survey of the prevalence of refractive errors and eye diseases in urban and rural school children. Can J Ophthalmol. 2009; 44(3):328–333. doi: 10.3129/i09-030

792. van der Hoeve J. [Das Vorkommen von Strabismus an der Poliklinik für Augenheilkunde im “Ryks-Ziekenhuis zu Leiden” von Juli 1896 bis Dezember 1901.] Arch Augenh. 1902; 46:207–231. German

793. Varma R, Deneen J, Cotter S, et al. The multi-ethnic pediatric eye disease study: design and methods. Ophthalmic Epidemiol. 2006; 13(4):253–262. doi: 10.1080/09286580600719055

794. Varnakov SI, Utkin VF. [Concomitant strabismus and treatment of amblyopia according to data of the Office of Vision Protection for Children in Riazan]. Vestn Oftalmol. 1968; 81(2):38–41. Russian

795. Varshney AS, Solanki MG, Mahida MH, Patel C. The Prevalence and Association of Refractive Errors in Pediatric Strabismus: A Prospective Observational Study. SSR J Med Sci. 2024; 1(1):13–21. doi: 10.5281/zenodo.14005882

796. Vasantha N, Haleema A, Raheena KP, Parvathy PS, Haneef M. A study on the prevalence of ocular disorders among school going children in the age group of 5 to 12 years. Indian J Clin Exp Ophthalmol. 2021; 7(1):73–77. doi: 10.18231/j.ijceo.2021.015

797. Vats V, Arora D, Gupta P. Screening of the Squint among Residents Visiting a Tertiary Care Hospital in Uttarakhand. J Epidemiol Publ Health. 2024; 9(2):138–144. doi: 10.26911/jepublichealth.2024.09.02.01

798. Vattathil S, Akey JM. Small amounts of archaic admixture provide big insights into human history. Cell. 2015; 163(2):281–284. doi: 10.1016/j.cell.2015.09.042

799. Vázquez-Aguirre N, Arroyo-Yllanes ME, Fonte-Vázquez A. Dissociated deviation in sensorial strabismus. Rev Mex Oftalmol. 2018; 92(1):6–11. doi: 10.24875/RMO.M18000008

800. Vereecken E, Feron A, Evens L. [The importance of early detection of strabismus and amblyopia]. Bull Soc Belge Ophtalmol. 1966; 143:729–739. French

801. Verhoeven VJM, Morgan IG, Guggenheim JA. Myopia is predominantly genetic or predominantly environmental? Ophthalmic Physiol Opt. 2025; 45(4):911–917. doi: 10.1111/opo.13464

802. Verlee DL. Ophthalmic survey in the Solomon Islands. Am J Ophthalmol. 1968; 66(2):304–319. doi: 10.1016/0002-9394(68)92080-1

803. Vernot B, Tucci S, Kelso J, et al. Excavating Neandertal and Denisovan DNA from the genomes of Melanesian individuals. Science. 2016; 352(6282):235-239. doi: 10.1126/science.aad9416

804. Vinodhini K, Anuradha TR, Vaishnavi J, Nazeem FG. A study on prevalence of refractive errors, strabismus and amblyopia in pediatric age group in tertiary care hospital. J Evidence Based Med Healthcare. 2018; 5(8):709–713. doi: 10.18410/jebmh/2018/144

805. Vlachou C, Mellou K, Tsaras K, Sparos L. [Determinants of strabismus frequency]. Arch Hellenic Med. 2003; 20(3):276–280. Greek

806. Voirol AF. [Untersuchungen ueber Refraktion, Visus, Farbensinn und Muskelgleichgewicht an den Augen von 939 Schulkindern.] Zeitschr Augenheilk. 1912; 28:95–110. German

807. von Bartheld CS, Croes SA, Johnson LA. Strabismus. In: Levin LA & Albert DM (eds.) Ocular Disease: Mechanisms and Management. San Diego: Elsevier 454-460, 2010.

808. von Bartheld CS, Chand A, Wang L. Prevalence and Etiology of Strabismus in Down Syndrome: A Systematic Review and Meta-Analysis with a Focus on Ethnic Differences in the Esotropia/Exotropia Ratio. Ophthalmic Epidemiol. 2025; 32(6):633–651. doi: 10.1080/09286586.2025.2500018

809. von Csapody-Mocsy M. [Ein Beitrag zur Pathologie des Begleitschielens.] Klin Monatsbl Augenheilkd. 1934; 92:385–387. German

810. von Noorden GK, Campos EC. Binocular Vision and Ocular Motility. Sixth Edition, Mosby, St. Louis, 653 pages, 2002.

811. von Wecker L. [Ueber spontane Heilung des hypermetropischen Strabismus und die Dosierung der Tenotomie.] Klin Monatsbl Augenheilkd. 1871; 9:453–459. German

812. Voo I, Lee DA, Oelrich FO. Prevalences of ocular conditions among Hispanic, white, Asian, and black immigrant students examined by the UCLA Mobile Eye Clinic. J Am Optom Assoc. 1998; 69(4):255–261.

813. Vora U, Khandekar R, Natrajan S, Al-Hadrami K. Refractive error and visual functions in children with special needs compared with the first grade school students in Oman. Middle East Afr J Ophthalmol. 2010; 17(4):297–302. doi: 10.4103/0974-9233.71590

814. Vyas DB, Lee DA. Eye conditions among 5- to 7-year-old Asian-Pacific Islander schoolchildren in Southern California. Optometry. 2001; 72(7):426–434.

815. Waardenburg PJ. Squint and heredity. Doc Ophthalmol. 1954; 7-8:422-494. doi: 10.1007/BF00238145

816. Waheeda-Azwa H, Norihan I, Tai ELM, Kueh YC, Shatriah I. Visual outcome and factors influencing surgical outcome of horizontal strabismus surgery in a teaching hospital in Malaysia: A 5-year experience. Taiwan J Ophthalmol. 2020; 10(4):278–283. doi: 10.4103/tjo.tjo_71_19

817. Wallace DK, Christiansen SP, Sprunger DT, et al. Esotropia and Exotropia Preferred Practice Pattern®. Ophthalmology. 2018; 125(1):P143–P183. doi: 10.1016/j.ophtha.2017.10.007

818. Walline JJ, Johnson Carder ED. Vision problems of children with individualized education programs. J Behav Optom. 2012; 23(4):87-93.

819. Wan X, Wan L, Jiang M, Ding Y, Wang Y, Zhang J. A retrospective survey of strabismus surgery in a tertiary eye center in northern China, 2014-2019. BMC Ophthalmol. 2021; 21(1):40. doi: 10.1186/s12886-021-01805-w

820. Wang HW, Li YC, Sun F, et al. Revisiting the role of the Himalayas in peopling Nepal: insights from mitochondrial genomes. J Hum Genet. 2012; 57(4):228–234. doi: 10.1038/jhg.2012.8

821. Wang YC, Lee RF, Wang MW, Jeng ST, Lin SA. [Report of ocular examination of primary school children in south Taiwan.] Acta Soc Ophthalmol Sin. 1989; 28(1):29–33. Chinese

822. Wang Y, Zhao A, Zhang X, et al. Prevalence of strabismus among preschool children in eastern China and comparison at a 5-year interval: a population-based cross-sectional study. BMJ Open. 2021; 11(10):e055112. doi: 10.1136/bmjopen-2021-055112

823. Warkad VU, Panda L, Behera P, Das T, Mohanta BC, Khanna R. The Tribal Odisha Eye Disease Study (TOES) 1: prevalence and causes of visual impairment among tribal children in an urban school in Eastern India. J AAPOS. 2018; 22(2):145.e1-145.e6. doi: 10.1016/j.jaapos.2017.10.020

824. Weakley DR, Dabes EM, Birch E. Trends in surgical correction of strabismus: a 20-year experience, 1990-2009. J AAPOS. 2011; 15(3):219-223. doi: 10.1016/j.jaapos.2011.03.006

825. Wedner SH, Ross DA, Balira R, Kaji L, Foster A. Prevalence of eye diseases in primary school children in a rural area of Tanzania. Brit J Ophthalmol. 2000; 84(11):1291–1297. doi: 10.1136/bjo.84.11.1291

826. Weiss L. [Über das Schielen und seine Spontanheilung.] Bericht 23. Versamml. Ophthalmol Gesellsch. (Heidelberg). Stuttgart: Union, Dtsche Verlagsgesellsch. 1893, 23:122-140. German

827. Weiss L. Upon the relation between the internal and external recti as affected by increasing divergence of the orbits. Arch Ophthalmol. 1896; 25:341–348.

828. Wesson ME. Intermittent divergent strabismus of the divergence excess type. Brit Orthopt J. 1961; 18:114–116.

829. Wharton KR, Yolton RL. Visual characteristics of rural Central and South Americans. J Am Optom Assoc. 1986; 57(6):426–430.

830. Wick B, Crane S. A vision profile of American Indian children. Am J Optom Physiol Opt. 1976; 53(1):34–40. doi: 10.1097/00006324-197601000-00006

831. Wickelgren LW. Convergence in the human newborn. J Exp Child Psychol. 1967; 5(1):74–85.

832. Wiesinger H. Ocular findings in mentally retarded children. J Pediatr Ophthalmol. 1964; 1(3):37–41.

833. Williams C, Northstone K, Howard M, Harvey I, Harrad RA, Sparrow JM. Prevalence and risk factors for common vision problems in children: data from the ALSPAC study. Brit J Ophthalmol. 2008; 92(7):959–964. doi: 10.1136/bjo.2007.134700

834. Williams DR, Priest N, Anderson NB. Understanding associations among race, socioeconomic status, and health: Patterns and prospects. Health Psychol. 2016; 35(4):407–411. doi: 10.1037/hea0000242

835. Woodruff ME, Samek MJ. The refractive status of Belcher Island Eskimos. Can J Public Health. 1976; 67(4):314–320.

836. Woodruff ME. Prevalence of visual and ocular anomalies in 168 non-institutionalized mentally retarded children. Can J Public Health. 1977; 68(3):225–232.

837. Woodruff ME. Vision and refractive status among grade 1 children of the Province of New Brunswick. Am J Optom Physiol Opt. 1986; 63(7):545–552. doi: 10.1097/00006324-198607000-00008

838. Worku Y, Bayu S. Screening for ocular abnormalities and subnormal vision in school children of Butajira Town, southern Ethiopia. Ethiop J Health Dev. 2002; 16(2):165–171.

839. Worth CA. Squint: Its Causes, Pathology, And Treatment. 3^rd^ Edition. 234 pages, 1906.

840. Worth CA, Chavasse FB. Worth and Chavasse’s Squint: The Binocular Reflexes and the Treatment of Strabismus. Blakiston’s Son & Co, Philadelphia, 1939.

841. Wyatt HT, Boyd TA. Strabismus and strabismic amblyopia in Northern Canada. Can J Ophthalmol. 1973; 8(2):244–251.

842. Yang Y, Wang C, Gan Y, et al. Maternal smoking during pregnancy and the risk of strabismus in offspring: a meta-analysis. Acta Ophthalmol. 2019; 97(4):353–363. doi: 10.1111/aos.13953

843. Yassur Y, Yassur S, Zaifrani S, Sachs U, Ben-Sira I. Amblyopia among African pupils in Rwanda. Brit J Ophthalmol. 1972; 56(4):368–370. doi: 10.1136/bjo.56.4.368

844. Yazawa K. [Ocular examination on 10,000 school children.] Rinsho Ganka (Jpn J Clin Ophthalmol.) 1973; 27:557–569. Japanese

845. Ye XC, Pegado V, Patel MS, Wasserman WW. Strabismus genetics across a spectrum of eye misalignment disorders. Clin Genet. 2014; 86(2):103–111. doi: 10.1111/cge.12367

846. Yekta A, Fotouhi A, Hashemi H, et al. The prevalence of anisometropia, amblyopia and strabismus in schoolchildren of Shiraz, Iran. Strabismus. 2010; 18(3):104–110. doi: 10.3109/09273972.2010.502957

847. Yekta A, Hashemi H, Azizi E, et al. The prevalence of amblyopia and strabismus among schoolchildren in Northeastern Iran, 2011. Iran J Ophthalmol. 2012; 24:3-10.

848. Yekta A, Hashemi H, Ostadimoghaddam H, et al. Strabismus and near point of convergence and amblyopia in 4-6 year-old children. Strabismus. 2016; 24(3):113–119. doi: 10.1080/09273972.2016.1205103

849. Yekta A, Hashemi H, Norouzirad R, et al. The prevalence of amblyopia, strabismus, and ptosis in schoolchildren of Dezful. Eur J Ophthalmol. 2017; 27(1):109–112. doi: 10.5301/ejo.5000795

850. Yetkin AA, Turkman IH. Evaluation of clinical characteristics and risk factors of strabismus cases. North Clin Istanb. 2023; 10(2):157–162. doi: 10.14744/nci.2023.15579

851. Yetkin AA. Factors Affecting Surgical Success Rates in Pediatric Horizontal Strabismus Surgery. Cureus. 2024; 16(11):e74758. doi: 10.7759/cureus.74758

852. Yin L, Chen X. [The causative diseases, common comorbidities and surgical procedures of 948 cases of horizontal sensory strabismus]. Zhonghua Yan Ke Za Zhi. 2018; 54(4):283–287. Chinese. doi: 10.3760/cma.j.issn.0412-4081.2018.04.010

853. Ying GS, Maguire MG, Cyert LA, et al. Prevalence of vision disorders by racial and ethnic group among children participating in head start. Ophthalmology. 2014; 121(3):630–636. doi: 10.1016/j.ophtha.2013.09.036

854. Yoo EJ, Kim SH. Optimal surgical timing in infantile exotropia. Can J Ophthalmol. 2014; 49(4):358–362. doi: 10.1016/j.jcjo.2014.05.004

855. Yoon KC, Mun GH, Kim SD, et al. Prevalence of eye diseases in South Korea: data from the Korea National Health and Nutrition Examination Survey 2008-2009. Korean J Ophthalmol. 2011; 25(6):421–433. doi: 10.3341/kjo.2011.25.6.421

856. Yu YS, Kim SM, Kwon JY, et al. [Preschool vision screening in Korea: Preliminary study.] J Korean Ophthalmol Soc. 1991; 32(12):1092–1096. Korean

857. Yu CB, Fan DS, Wong VW, Wong CY, Lam DS. Changing patterns of strabismus: a decade of experience in Hong Kong. Brit J Ophthalmol. 2002; 86(8):854–856. doi: 10.1136/bjo.86.8.854

858. Yu X, Ji Z, Yu H, Xu M, Xu J. Exotropia is the main pattern of childhood strabismus surgery in the South of China: A six-year clinical review. J Ophthalmol. 2016; 2016:1489537. doi: 10.1155/2016/1489537

859. Zaki AAH, Keeney AH. The bony orbital walls in horizontal strabismus. AMA Arch Ophthalmol. 1957; 57(3):418–424. doi: 10.1001/archopht.1957.00930050430014

860. Zaki HA. A surgery on twenty six thousand and three hundred seventy eight patients with convergent squint (26378), thirteen thousand, one hundred and eighty one patients with divergent squint (13181), one thousand and twenty nine patients of vertical squint (1029), ten thousand and one hundred thirty nine patients of heterophoria (10139) and two thousand sixty one patients of paralytic squint (2061) seen during the past ten years at Giza Ophthalmic Hospital Squint Clinic from the year 1960 to the year 1970. Bull Ophthalmol Soc Egypt. 1972; 65(69):457–470.

861. Zehra Z, von Bartheld CS, Khan W, Azam M, Qamar R. Prevalence of strabismus in Pakistan: a systematic review and meta-analysis. Strabismus. 2025a; 33(1):44–53. doi: 10.1080/09273972.2024.2408416

862. Zehra Z, Hagen MM, Wang L, von Bartheld CS. The interpupillary distance differs between ethnicities and associates with horizontal strabismus patterns: Evidence from a systematic review and meta-analysis. medRxiv [Preprint] 2025b; posted December 30, 2025. doi: 10.64898/2025.12.30.25343217

863. Zeidan Z, Hashim K, Muhit MA, Gilbert C. Prevalence and causes of childhood blindness in camps for displaced persons in Khartoum: results of a household survey. East Mediterr Health J. 2007; 13(3):580–585.

864. Zhang XJ, Lau YH, Wang YM, et al. Prevalence of strabismus and its risk factors among school aged children: The Hong Kong Children Eye Study. Sci Rep. 2021; 11(1):13820. doi: 10.1038/s41598-021-93131-w

865. Zhao J, Pan X, Sui R, Munoz SR, Sperduto RD, Ellwein LB. Refractive error study in children: results from Shunyi District, China. Am J Ophthalmol. 2000; 129(4):427–435. doi: 10.1016/s0002-9394(99)00452-3

866. Zhu H, Yu JJ, Yu RB, et al. Association between childhood strabismus and refractive error in Chinese preschool children. PLoS One. 2015; 10(3):e0120720. doi: 10.1371/journal.pone.0120720

867. Zhu H, Pan C, Sun Q, et al. Prevalence of amblyopia and strabismus in Hani school children in rural southwest China: a cross-sectional study. BMJ Open. 2019; 9(2):e025441. doi: 10.1136/bmjopen-2018-025441

868. Ziakas NG, Woodruff G, Smith LK, Thompson JR. A study of heredity as a risk factor in strabismus. Eye (Lond). 2002; 16(5):519–521. doi: 10.1038/sj.eye.6700138

869. Zimmermann-Paiz MA, Ordonez-Rivas AM. [Frequency of various strabismus types in an ophthalmology facility of Guatemala City.] Rev Mex Oftalmol. 2013; 87(4):195–199. Spanish

## Other Cited Material

A. Sharbini SH. Prevalence of strabismus & associated risk factors: The Sydney Childhood Eye Studies. Dissertation, University of Sydney, Faculty of Health Sciences (Orthoptic), 2015, 290 pp.

B. Adinanto FC, French AN, Rose KA. The prevalence of strabismus: a systematic literature review. Invest Ophthalmol Vis Sci 2016; 57(12):2457.

C. Adinanto FC. Development of vision and strabismus in childhood: prevalence and risk factors. Orthoptic Thesis. University of Technology Sydney. 2020, 194 pages.

D. Roberts J, Rowland M. Refractive status and motility defects of persons 4-74 years, United States 1971-1972. Vital and health statistics: series 11, DHEW publication no. (PHS) 78-1654. Hyattsville, MD: National Center for Health Statistics, 1978.

E. PROSPERO (International prospective register of systematic reviews, CRD420251080354) https://www.crd.york.ac.uk/PROSPERO/view/CRD420251080354

F. Web of Science. https://www-webofscience-com.unr.idm.oclc.org/wos/woscc/cited-reference-search

G. Ward BA. Ophthalmic survey of Fiji with special reference to the problem of Trachoma, February-November 1955. In: Annual Report of the Medical Department for the year 1955 and File A48/201/1, pages 62–68. Suva: National Archives of Fiji.

H. United Nations Report: List of countries by population: https://en.wikipedia.org/wiki/List_of_countries_by_population_(United_Nations) https://en.wikipedia.org/wiki/List_of_countries_by_population_in_1989 (GeoHive - World Population 1950-2050). United Nations. World Population Prospects The 2017 Revision Key Findings and Advance Tables. Department of Economic and Social Affairs, Population Division. United Nations, New York, 2017 https://esa.un.org/unpd/wpp/Publications/Files/WPP2017_KeyFindings.pdf, United Nations, Department of Economic and Social Affairs, Population Division (2019). World Population Prospects 2019: Online Edition. https://population.un.org/wpp/ https://en.wikipedia.org/wiki/List_of_countries_by_population_(United_Nations)

I. Carpiuc KT. An appraisal of childhood vision problems: evaluating risk factors and the psychosocial impact of strabismus in children. Dissertation in Public Health, 2013, University of California, San Diego and San Diego State University. https://cloudfront.escholarship.org/dist/prd/content/qt9sw615jv/qt9sw615jv.pdf

J. Duelund N. Vision Screening of Children in Greenland (Doctoral dissertation, 2024, UNIVERSITY OF COPENHAGEN).

K. Polling JR, Tideman W, Jaddoe V, Hofman A, Klaver CCW. Refractive error determines type of strabismus: The generation R study. Invest Ophthalmol Vis Sci. 2015; 56:7 (5228).

L. Gunnufsen J. Strabismus in Oslo. Norwegian. Cited by Holst JC, Tjaland J (1962).

M. Dalz M, Gotz-Wieckowska A, Dalz M. [Schielenkrankheit bei den Kindern im Schulalter.] 107^th^ Kongress, Deutsche Ophthalmologische Gesellschaft (DOG), Leipzig, Germany, Abstract P 094, 2009 https://2009.archiv.dog.org/abstracts/P094.html

N. Quental H, Poças IM, Esteves C, Quintino W, Fortes CS. [Caracterização visual numa amostra infantil em idade pré-escolar e escolar - o estado da arte num rastreio.] XIV Congresso Nacional de Ortoptistas, 2013. https://pdfs.semanticscholar.org/00b8/c3287f0cbde1d04ec7514b0d0a1d707ba4dd.pdf

O. Roberts J. Eye examination findings among children, United States. Vital and health statistics: Series 11-No. 115, DHEW publication No. (HSM) 72-1057. Health Services and Mental Health Administration. Washington. U.S. Government Printing Office, June 1972.

P. Roberts J. Eye examination findings among youths aged 12-17 years, United States. Vital and health statistics: Series 11-No. 155, DHEW publication no. (HRA) 76-1637. Health Resources Administration. Washington. U.S. Government Printing Office, Nov. 1975.

Q. Vasquez SC, Lopez AMV. [Análisis de la situación de salud visual y ocular en los niños y niñas entre 3 y 18 años pertenecientes al proyecto de educacion en convivencia y ciudadanía pecc en la clínica de optometría de la universidad de la sale.] Thesis, Univ de la Salle, Bogota, 2014 https://ciencia.lasalle.edu.co/optometria/165/

R. Onsomu EM. Strabismus as seen in children aged 3 to 5 years attending Nairobi City Council day nursery schools in Nairobi province, Kenya. Dissertation, 2003, University of Nairobi, College of Health Sciences [2678]

S. Mutie DM. Ocular morbidity in nursery school children in Kilungu Division, Makueni District. Dissertation Master of Medicine, Ophthalmology, University of Nairobi, Kenya, 2008.

T. Atamah AO. Frequency and pattern of eye diseases among prisoners in Benin City. (A survey of Oko prisons, Benin City). Dissertation, Medical College of Nigeria, 2005.

U. Kwak J, Movahedan J, Tadjvidi P, Aref P, Maumenee IH. Mendelian inheritance of strabismus in a rural community of Northern Iran. Invest Ophthalmol Vis Sci. 2011; 52(14):6359

V. Hopkins S. A visual profile of Queensland indigenous and non-indigenous school children, and the association between vision and reading. Queensland University of Technology Thesis, 2014.

W. Pai ASI. Vision and refraction in Australian preschool children: Measures, prevalence, and associated factors. PhD Thesis, University of Sydney, 2013.

X. Mann I, Loschdörfer J. Ophthalmic survey of the Territories of Papua and New Guinea, 1955. Public Health Department publication, Port Moresby, 1956, 53 p.

Y. Phelan A. Child eye health in Nampula, Mozambique and Ireland. Masters dissertation, 2015. Dublin Institute of Technology. doi:10.21427/D7VK5Q.

Z. De Haas JH. [Geschiedkundig onderzoek omtrent de hypermetropie en hare Gevolgen.] Dissertation, Hoogeschool te Utrecht, Wed. Locke & Zoon, Rotterdam, 1862.

AA. Tasevska D. Prevalence and causes of visual impairment in children, Master thesis, Saints Cyril and Methodius Univ, Faculty of Philosophy, Skopje, 2019.

BB. Vladutiu C. Infant vision: First steps in Romania. Perception 1997; 26:785 (abstr). 6^th^ Meeting of the Child Vision Research Society (CVRS) Pisa, Italy, 6-8 June, 1997

CC. Contreras Roldán A. [Revisión y epidemiología de la cirugía del estrabismo en el área hospitalaria Virgen Macarena. Descripción de la casuística y resultados obtenidos.] Tesis Doctoral Inédita, Universidad de Sevilla, Sevilla, 2017.

DD. Belamkar AV, Haider KM. Prevalence and Clinical Characteristics of Adult Strabismus. Proceedings of IMPRS. 2022; 5(1) (Abstract). doi: 10.18060/27119

EE. Radenovich V, Balesh AM. Eye on the Border. From the Files of a Pediatric Ophthalmologist. [2006] http://childrenseyecenter.com/wp-content/uploads/2017/02/EyeontheBorder.pdf

FF. Melek NB. [La exotropia intermitente: observaciones clínicas y quirúrgicas.] Catálogo de la Biblioteca CAO, Buenos Aires, 1976. https://www.oftalmologos.org.ar/catalogo/items/show/5531

GG. Rodrigues EB. [Estrabismo na criança - triagem em um ambulatorio de pediatria.] Dissertation, Universidade Federal de Santa Catarina, Florianopolis, Brazil, 1998.

HH. Secin VKAV. [Construcao do Conceito Social de Intervencao Terapeutica Ortoptica: O ortoptista como agente informal da Educacao.] Dissertation, Universidade do Estado do Rio de Janeiro, Faculdade de Educacao, 2005, 193 pp.

II. Pereira CM. Estudo das características epidemiológicas da deficiência visual dos pacientes matriculados no setor de baixa visão infantil do Hospital São Geraldo-Hospital das Clínicas-Universidade Federal de Minas Gerais nos anos de 2012 a 2019. Dissertation, Universidade Federal de Minas Gerais, 2024.

JJ. Fazal AF. 2012. Review of outcome of horizontal childhood strabismus surgery at Kenyatta National Hospital and Kikuyu Eye Unit – a retrospective study. Dissertation, University of Nairobi, H58/78551.

KK. Oummad H. [Le strabisme de l’adulte.] Thesis de Medecine. Univ. Mohammed V, Faculte de Medecine et de Pharmacie, Rabat, Morocco, 2011.

LL. Mohammed K. [Chirurgie du Strabisme Horizontal (A propos de 55 cas)]. Thesis, Univ Sidi Mohammed Ben Abdellah, Fez City, Morocco, 2014.

MM. Mustafa MSE. Common eye disorders in children attending Khartoum Eye Teaching Hospital. Thesis, 2006, University of Khartoum.

NN. Fofana Z. Approche medicale du strabisme au chu de Cocody. Thesis Doctorate en Meecine, Universite de Cocody, Abidjan, Cote d”Ivoire. 2001, 139 pp.

OO. Akinsola FB. An analysis of eye disease in Nigerian children seen at Lagos University Teaching Hospital. Fellowship dissertation. National postgraduate medical college of Nigeria, Lagos 1990.

PP. Morgan RE. Pattern of eye diseases in children seen at Jos university teaching hospital. Fellowship dissertation. National postgraduate medical college of Nigeria, Lagos, 1995.

QQ. Onyekwe LO. Visual Impairment amongst school children in Jos plateau state, Nigeria. Fellowship dissertation. National postgraduate medical college of Nigeria, Lagos, 1995.

RR. Isawumi MA. Ocular disorders among school children in Ilesa- East, Osun state, Nigeria. *Fellowship dissertation. National postgraduate medical college of Nigeria, Lagos*, 2003.

SS. Ranjbar-Pazook M., Khorrami-Nejad M. The prevalence of refractive error and strabismus types in an academic hospital. 7th World Congress Controversies in Ophthalmology, Warsaw, Poland, B-16, 2016. http://www.comtecmed.com/cophy/2016/posters_B.aspx

TT. Sarıkaya İ. Çocuk ve ergenlerde şaşılık cerrahisinin yaşam kalitesine etkisi [EFFECT OF STRABISMUS SURGERY ON QUALITY OF LIFE IN CHILDREN AND ADOLESCENTS]. Thesis, PAMUKKALE UNIVERSITY, FACULTY OF MEDICINE, DEPARTMENT OF EYE DISEASES, 2024.

UU. Sharbini SH, Wang JJ, Burlutsky G, Rose KA, Mitchell P, Sydney Childhood Eye Study, Sydney Myopia Study. Parental factors associated with strabismus in a population-based sample of 12-year old Australian children. Invest Ophthalmol Vis Sci. 2008; 49:1807 (abstr).

VV. Annual Report for 1911 of the Chief Medical Officer of the [British] Board of Education. London, His Majesty’s Stationery Office, 1912, 1917, 1921 and 1922.

WW. Lee PP, Stein JD, Anthopolos R, Tootoo J, Andrews CA, Miranda ML. Assessing geographic variability in rates of strabismus among children on Medicaid in Michigan and North Carolina. Invest Ophthalmol Vis Sci. 2015; 56:7 (1394).

XX. Scott J. The variation of measurements: considering the morphology of late Pleistocene hominins. Thesis, Central Michigan University, 2016.

YY. von Bartheld CS, Agarwal A, Feng CY, Christensen A, Wen D, Cassinelli K, Kirkpatrick B, Johnson A. Molecular and developmental links between strabismus and schizophrenia. Invest Ophthalmol Vis Sci. 2016; 57(12):2460.

